# Comparative effectiveness of digital health interventions for low back pain: systematic review and network meta-analysis of randomised controlled trials

**DOI:** 10.64898/2026.09.23.26363780

**Authors:** Runfu Chen, Zhuotao Wu, Xiaojin Wei, Wenjie Yang, Caiyi Yan, Hai Chen, Xinyi Zeng, Jinpeng Zhang, Xianzheng Zeng, Chanyan Huang, Shu Zhang, Tao Xu, Hong Xiao, Jiangning Song, Hosam Alzahrani, Francis Fatoye, Daqing Ma, Chan Chen

**Affiliations:** Department of Anesthesiology, The Research Units of West China (2018RU012), Chinese Academy of Medical Sciences, West China Hospital, Sichuan University, Chengdu, Sichuan, China; Center of Biostatistics, Design, Measurement and Evaluation (CBDME), West China Hospital, Sichuan University, Chengdu, Sichuan, China; Department of Anesthesiology, Sichuan Public Health General Clinical Center, Jincheng Hospital, West China Hospital, Sichuan University, Chengdu, Sichuan, China; Department of Respiratory and Critical Care Medicine, Targeted Tracer Research and Development Laboratory, West China Hospital, Sichuan University, Chengdu, Sichuan, China; Department of Pain Management, West China Hospital, Sichuan University, Chengdu, Sichuan, China; Department of Anesthesiology, The First Affiliated Hospital, Sun Yat-sen University, Guangzhou, China; Department of Emergency Medicine, West China Hospital, Sichuan University, Chengdu, Sichuan, China; Department of Anesthesiology, Sixth People’s Hospital Affiliated to Shanghai Jiao Tong University School of Medicine, Shanghai, China; Key Laboratory of Anesthesiology, Ministry of Education, Shanghai Jiao Tong University, Shanghai, China; Biomedicine Discovery Institute and Department of Biochemistry and Molecular Biology, Monash University, Melbourne, Victoria, Australia; Department of Physical Therapy, College of Applied Medical Sciences, Taif University, Taif, Makkah Province, Saudi Arabia; Department of Health Professions, Manchester Metropolitan University, Manchester, UK; Department of Anesthesiology, Perioperative and Systems Medicine Laboratory, Children’s Hospital, Zhejiang University School of Medicine, National Clinical Research Center for Child and Adolescent’s Health and Disease, Hangzhou, China; Division of Anaesthetics, Pain Medicine and Intensive Care, Department of Surgery and Cancer, Faculty of Medicine, Imperial College London, Chelsea and Westminster Hospital, London, UK

## Abstract

**Background:** Digital health interventions are increasingly used for low back pain, but their comparative effectiveness across different delivery modalities, clinical outcomes, and follow-up periods remains uncertain. We aimed to compare the effects of major digital health modalities for adults with low back pain.

**Methods:** We conducted a systematic review and frequentist random-effects network meta-analysis of randomised controlled trials. PubMed, Embase, CENTRAL, CINAHL, APA PsycINFO, ClinicalTrials.gov, and WHO ICTRP were searched from inception to March 19, 2026, and updated to Aug 8, 2026. Eight treatment nodes were compared: telemedicine, web applications, mobile applications, virtual reality, exergaming, biofeedback devices, face-to-face care, and control. Primary outcomes were pain intensity and physical function; secondary outcomes were pain-related fear avoidance, health-related quality of life, self-efficacy, anxiety, and depression. The review was registered with PROSPERO (CRD420261339841).

**Findings:** We included 92 reports from 84 randomised controlled trials involving 11 803 participants. At post-intervention, virtual reality had the most favourable point estimate for pain intensity (SMD −1.43, 95% CI −1.87 to −0.98; moderate confidence). The estimate for physical function also favoured virtual reality (−0.86, −1.45 to −0.26), although confidence was very low. Virtual reality also probably reduced pain-related fear avoidance (−1.14, −1.68 to −0.61) and improved health-related quality of life (0.97, 0.57 to 1.36), both with moderate confidence. Other digital modalities showed favourable effects for selected outcomes, but confidence was generally low or very low. Networks became progressively sparse at later follow-up, and sustained effects were inconsistent. Global inconsistency was detected in the post-intervention pain-intensity network, with local inconsistency for selected exergaming comparisons.

**Interpretation:** Digital health interventions for low back pain showed different effects across outcomes and follow-up periods. Virtual reality had the most favourable post-intervention estimates for pain intensity, pain-related fear avoidance, and health-related quality of life compared with control, with moderate confidence in these estimates. Larger head-to-head trials with longer follow-up are needed to determine whether these effects are sustained and to clarify the comparative effectiveness of digital interventions.

**Funding:** National Key Research and Development Programme of China (2025YFC3607901).

**Research in context:** *Evidence before this study:* We searched PubMed, Embase, the Cochrane Central Register of Controlled Trials, CINAHL, APA PsycINFO, ClinicalTrials.gov, and the WHO International Clinical Trials Registry Platform from database inception to March 19, 2026, and updated the search on Aug 8, 2026. We used terms related to low back pain, digital health technologies, and randomised trials, and screened the reference lists of systematic reviews. Previous reviews examined individual modalities, such as mobile applications, telehealth, or virtual reality, and used pairwise meta-analysis. Existing network meta-analyses either included broader chronic pain populations or compared a narrower range of rehabilitation approaches for low back pain. Although these reviews suggested that some digital interventions might improve pain or physical function, their relative effects across clinical outcomes and follow-up periods remained uncertain. To our knowledge, no previous synthesis had compared digital delivery modalities for low back pain across pain, physical function, and psychosocial outcomes while assessing confidence in the network estimates.

*Added value of this study:* This systematic review and network meta-analysis included 92 reports from 84 randomised controlled trials involving 11 803 participants. We compared six digital modalities—telemedicine, web applications, mobile applications, virtual reality, exergaming, and biofeedback devices—with face-to-face care and control across seven outcomes and follow-up periods extending from treatment completion to 12 months or longer. Virtual reality had the broadest favourable estimates at post-intervention, including reductions in pain intensity and pain-related fear avoidance and improvement in health-related quality of life, all supported by moderate-confidence evidence. Other modalities showed favourable effects for selected outcomes, but no stable hierarchy was observed across outcomes and follow-up periods. Confidence in many estimates was low or very low, the post-intervention pain-intensity network showed inconsistency, and longer-term networks were sparse. These findings distinguish favourable post-intervention estimates from evidence of sustained comparative superiority.

*Implications of all the available evidence:* Digital health interventions might support the management of low back pain, but the available evidence does not establish that any single delivery modality should be preferred. Although virtual reality produced the broadest favourable post-intervention estimates, evidence of sustained superiority was not established, and the observed effects cannot be separated reliably from differences in therapeutic content, treatment intensity, and clinician support. Selection of a digital intervention should therefore consider the intended treatment outcome, therapeutic components, accessibility, digital literacy, costs, and patient preferences. Adequately powered head-to-head trials with longer follow-up are needed, together with component-based studies that distinguish the contribution of the technological platform from that of the treatment it delivers.

## Introduction

Low back pain is commonly defined as pain between the lower costal margin and the gluteal folds, with or without leg pain.^1^ In 2020, it affected approximately 619 million people worldwide and remains the leading cause of years lived with disability, imposing a substantial burden through pain, functional limitation, and psychological distress.^2,3^ Healthcare spending on low back and neck pain in the United States was estimated at US$134.5 billion in 2016.^4^ Management of low back pain includes both pharmacological and non-pharmacological treatments.^5^ However, pharmacological therapies generally provide only small, predominantly short-term benefits and may increase adverse events.^6^ Therefore, non-pharmacological strategies are central to guideline-based management, including education and supported self-management, exercise, psychological therapies, and multidisciplinary care.^5,7^

Digital health encompasses the use of digital technologies to support health and healthcare delivery.^8,9^ In low back pain, these technologies include telemedicine and telerehabilitation, web and mobile applications, virtual reality, exergaming, and biofeedback devices. Digital health interventions can deliver education, exercise, self-management support, monitoring, and interactive rehabilitation, and may complement conventional care.^10,11^ Previous systematic reviews only evaluated mobile applications, telerehabilitation, or virtual reality separately, using conventional pairwise meta-analyses.^12–15^ At the network level, a recent analysis of chronic non-specific low back pain compared AI-assisted telerehabilitation, telerehabilitation, in-person rehabilitation, and usual care, but did not incorporate the broader range of digital modalities available for low back pain.^16^ Consequently, the comparative effectiveness of these modalities for specific clinical outcomes and follow-up periods remains uncertain, limiting evidence-based treatment selection and implementation.

To address this evidence gap, we conducted a systematic review and network meta-analysis of randomised controlled trials evaluating digital health interventions for adults with low back pain. Unlike conventional pairwise meta-analysis, network meta-analysis combines direct and indirect evidence to enable simultaneous comparison of multiple interventions. We compared eight treatment nodes, including telemedicine, web applications, mobile applications, virtual reality, exergaming, biofeedback devices, face-to-face care, and control. Their effects on pain intensity, physical function, and other clinical outcomes were evaluated from post-intervention to 12 months or longer. This comprehensive comparative framework aims to provide clinically relevant evidence to inform treatment selection and future guideline recommendations for digital health in low back pain care.

## Methods

### Search strategy and selection criteria

This systematic review and network meta-analysis was reported in accordance with the PRISMA 2020 statement and the PRISMA extension for network meta-analyses.^17,18^ Completed reporting checklists are provided in the appendix (pp 6–12). The protocol was prospectively registered with PROSPERO before study selection and data extraction began (CRD420261339841), and deviations from the registered protocol are described in the appendix (p 13).

We searched PubMed, Embase, the Cochrane Central Register of Controlled Trials (CENTRAL), CINAHL, APA PsycINFO, ClinicalTrials.gov, and the World Health Organization International Clinical Trials Registry Platform (WHO ICTRP) from inception to 19 March 2026, with an updated search to 8 August 2026. Reference lists of relevant systematic reviews were also screened. Full search strategies are provided in appendix (pp 14–33). We included randomised controlled trials evaluating digital health interventions in adults aged 18 years or older with low back pain. Treatment nodes were defined before data synthesis according to the principal mode of digital delivery, informed by previous systematic reviews and network meta-analyses.^19–23^ Eight nodes were defined: control, face-to-face care, telemedicine, web applications (Web-App), mobile applications (Mobile-App), biofeedback devices, virtual reality (VR), and exergaming. Waiting-list, no-intervention, background usual care without study-specific active treatment, minimal advice or education, and sham or attention controls were classified as control; structured in-person physiotherapy or exercise programmes were classified as face-to-face care. Detailed operational definitions, boundary rules, and classifications of individual study arms are provided in appendix (pp 54–69).

Paired reviewers independently screened titles and abstracts, trial registry records where applicable, and potentially eligible full-text reports. Disagreements were resolved through discussion or consultation with another reviewer. Multiple reports or registry records from the same trial were linked and treated as a single study. The most complete published report was designated as the primary report, with companion reports and registry records used to supplement study characteristics, risk-of-bias assessments, and outcome data.

### Data analysis

Data were extracted independently by paired reviewers using a standardised form. Extracted information included study and participant characteristics, intervention characteristics, and outcomes; detailed study characteristics are provided in appendix (pp 70–78). Discrepancies were resolved by discussion. We assessed two primary outcomes, pain intensity and physical function, and five secondary outcomes, pain-related fear avoidance, health-related quality of life, self-efficacy, anxiety, and depression, all analysed as continuous outcomes.^24–27^ Outcome data were extracted at all available follow-up assessments and classified according to time since completion of the intervention as post-intervention (end of treatment or <2 months), short-term (≥2 to <6 months), mid-term (≥6 to <12 months), and long-term (≥12 months).^27^ Post-intervention was the primary time point for effectiveness analyses. When multiple assessments were available within the same interval, the assessment closest to the lower boundary of that interval was selected. When multiple instruments assessed the same outcome, one measure was selected according to the prespecified hierarchy described in appendix (pp 79–84).

Risk of bias was assessed independently by paired reviewers using the revised Cochrane risk-of-bias tool for randomised trials (RoB 2).^28,29^ Overall judgments were classified as low risk of bias, some concerns, or high risk of bias. Disagreements were resolved through discussion or consultation with a third reviewer. Detailed assessments are provided in appendix (pp 85–88).

Because outcomes were measured using different instruments, effects were expressed as standardised mean differences (SMDs; Hedges’ g) with 95% CIs. Scale directions were harmonised so that negative SMDs indicated improvement in pain intensity, physical function, pain-related fear avoidance, anxiety, and depression, and positive SMDs indicated improvement in health-related quality of life and self-efficacy. Endpoint scores were used at each follow-up. Missing or incomplete data were sought from investigators or derived or converted using established methods.^30,31^ Separate frequentist random-effects network meta-analyses were conducted for each outcome and follow-up interval with a connected network, using the graph-theoretical approach in netmeta.^32,33^ Between-study variance was estimated by restricted maximum likelihood and assumed common across treatment comparisons within each network. Multi-arm correlations were accounted for using netmeta’s multi-arm adjustment. Control was the reference treatment. An absolute SMD of 0.50 was used as a pragmatic threshold for potential clinical importance, consistent with a previous network meta-analysis of low back pain interventions.^27^ This threshold aided interpretation across outcomes but was not a validated outcome-specific minimal important difference. Based on median baseline standard deviations from eligible control groups, an SMD of 0.50 corresponded to approximately 0.85 points on a 0–10 numerical rating scale for pain intensity and 6.78 points on the 0–100 Oswestry Disability Index for physical function.^34^

The plausibility of the transitivity assumption was assessed by comparing potential effect modifiers across direct treatment comparisons, including intervention duration, mean age, proportion of female participants, and outcome measure (appendix pp 89–95). Between-study heterogeneity was quantified using τ^2^ and assessed with Cochran’s Q and *I*^2^ where estimable (appendix pp 127–131). Global inconsistency was assessed using the design-by-treatment interaction model, and local inconsistency using the Separate Indirect from Direct Evidence (SIDE) approach (appendix pp 132–133).^35–37^ Comparison-adjusted funnel plots were used to explore small-study effects in networks including at least 10 studies (appendix pp 134–140). Network plots, forest plots, and league tables are provided in appendix (pp 96–126). All analyses were conducted in R version 4.5.3, primarily using netmeta version 3.6.0.

Two sensitivity analyses were conducted for connected networks: excluding studies at overall high risk of bias and excluding studies with outcome data that were statistically estimated, converted, or otherwise derived rather than directly reported (appendix pp 141–165). Robustness was assessed by comparing the direction, magnitude, and 95% CIs of network estimates with those from the primary analyses. As a modification of the registered subgroup analysis by low back pain duration, we also conducted a restricted analysis including only trials enrolling participants with chronic low back pain, defined as symptoms lasting at least 12 weeks (appendix pp 166–178). Because this analysis did not formally compare treatment effects between chronic and non-chronic populations, it was interpreted as a restricted-population analysis rather than evidence of effect modification.

Confidence in all post-intervention network estimates and in short-term estimates for pain intensity and physical function was assessed using the CINeMA framework.^38,39^ Overall confidence was rated as high, moderate, low, or very low across six domains: within-study bias, reporting bias, indirectness, imprecision, heterogeneity, and incoherence. Most mid-term and long-term estimates were not formally assessed because the corresponding networks were sparse. Detailed CINeMA methods, domain-level judgments, and overall confidence ratings are provided in appendix (pp 179–188).

## Results

The searches identified 5579 records. After full-text assessment, 92 reports representing 84 unique randomised controlled trials involving 11 803 participants contributed to at least one network meta-analysis (Fig 1). Study and participant characteristics are presented in Table 1 and appendix pp 70–78. Five trials were judged to be at low risk of bias, 54 raised some concerns, and 25 were judged to be at high risk of bias (appendix pp 85–88). No clear imbalance was observed across the assessed effect modifiers (appendix pp 89–95). Network plots, forest plots, and league tables are provided in the appendix (pp 96–126). Network-level heterogeneity was substantial for pain intensity (*I*2=87.79%; τ^2^=0.57) and physical function (*I*2=87.41%; τ^2^=0.55) at post-intervention. Global inconsistency was detected for post-intervention pain intensity (Q=24.10, df=14; *P*<0.05), with local disagreement for selected exergaming comparisons (appendix pp 127, 132–133). Comparison-adjusted funnel plots showed no obvious asymmetry across evaluable networks (appendix pp 134–140). Figs 2 and 3 show the network geometry for pain intensity and physical function, respectively, and Fig 4 summarises post-intervention estimates versus control with CINeMA confidence ratings. Detailed CINeMA assessments and approximate back-transformed estimates for pain intensity and physical function are provided in the appendix (pp 189–191). Beyond post-intervention, networks became progressively sparser, and favourable estimates were less consistent across outcomes and follow-up periods. Confidence in most mid-term and long-term estimates was not formally assessed with CINeMA; complete later follow-up estimates are provided in the appendix (pp 102–126).

**Table 1:**
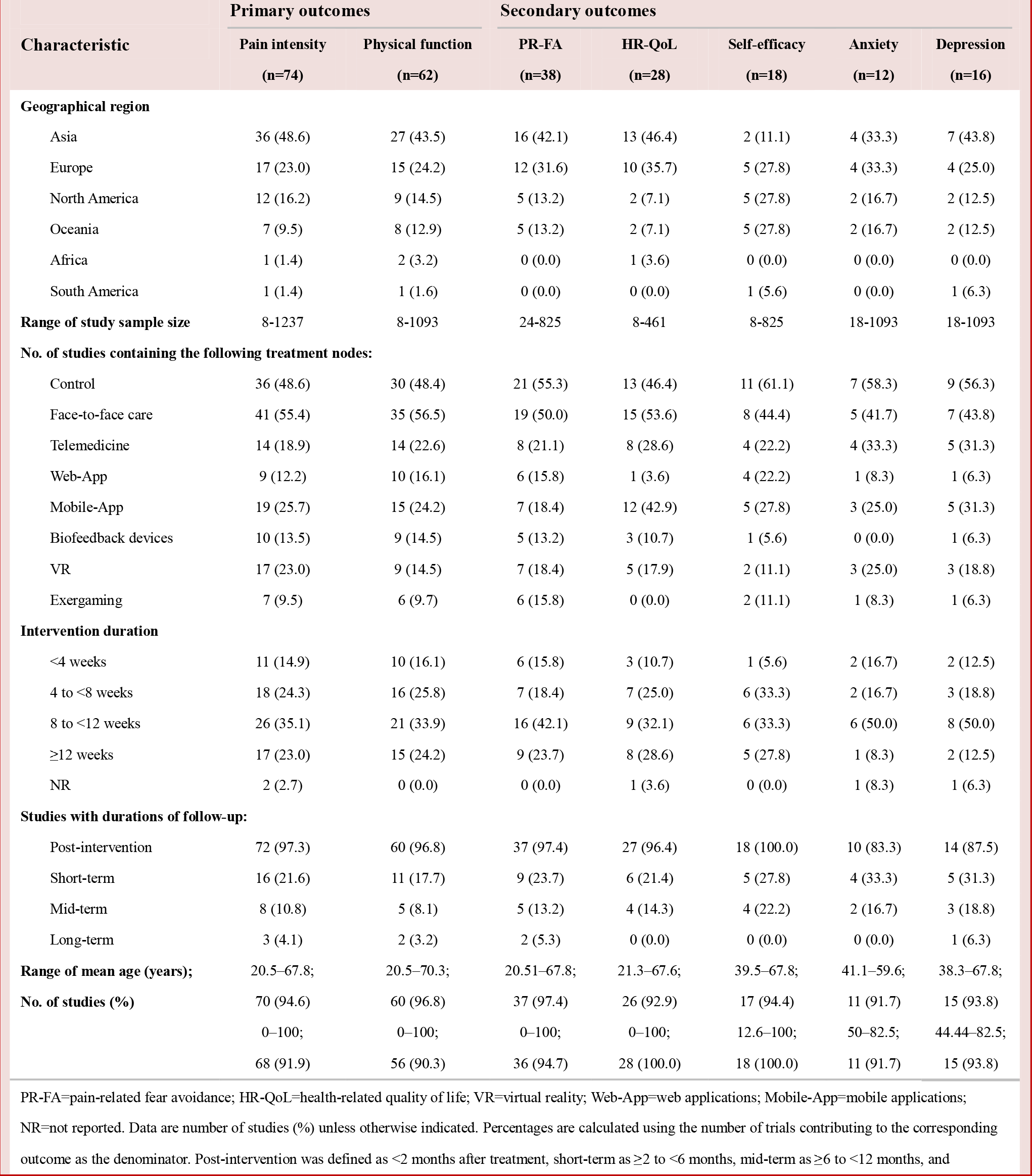
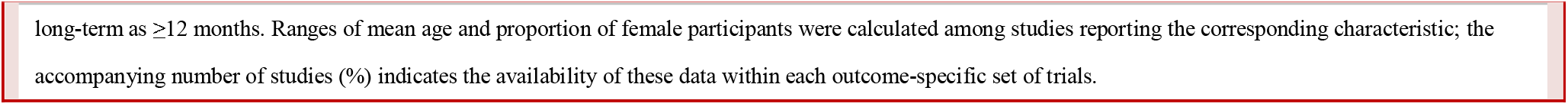
Summary characteristics of randomised controlled trials contributing to the network meta-analysis.

|  | Primary outcomes |  | Secondary outcomes |  |  |  |  |
|---|---|---|---|---|---|---|---|
| **Characteristic** | **Pain intensity<br>(n=74)** | **Physical function<br>(n=62)** | **PR-FA<br>(n=38)** | **HR-QoL<br>(n=28)** | **Self-efficacy<br>(n=18)** | **Anxiety<br>(n=12)** | **Depression<br>(n=16)** |
| **Geographical region** |  |  |  |  |  |  |  |
| Asia | 36 (48.6) | 27 (43.5) | 16 (42.1) | 13 (46.4) | 2 (11.1) | 4 (33.3) | 7 (43.8) |
| Europe | 17 (23.0) | 15 (24.2) | 12 (31.6) | 10 (35.7) | 5 (27.8) | 4 (33.3) | 4 (25.0) |
| North America | 12 (16.2) | 9 (14.5) | 5 (13.2) | 2 (7.1) | 5 (27.8) | 2 (16.7) | 2 (12.5) |
| Oceania | 7 (9.5) | 8 (12.9) | 5 (13.2) | 2 (7.1) | 5 (27.8) | 2 (16.7) | 2 (12.5) |
| Africa | 1 (1.4) | 2 (3.2) | 0 (0.0) | 1 (3.6) | 0 (0.0) | 0 (0.0) | 0 (0.0) |
| South America | 1 (1.4) | 1 (1.6) | 0 (0.0) | 0 (0.0) | 1 (5.6) | 0 (0.0) | 1 (6.3) |
| **Range of study sample size** | 8-1237 | 8-1093 | 24-825 | 8-461 | 8-825 | 18-1093 | 18-1093 |
| **No. of studies containing the following treatment nodes:** |  |  |  |  |  |  |  |
| Control | 36 (48.6) | 30 (48.4) | 21 (55.3) | 13 (46.4) | 11 (61.1) | 7 (58.3) | 9 (56.3) |
| Face-to-face care | 41 (55.4) | 35 (56.5) | 19 (50.0) | 15 (53.6) | 8 (44.4) | 5 (41.7) | 7 (43.8) |
| Telemedicine | 14 (18.9) | 14 (22.6) | 8 (21.1) | 8 (28.6) | 4 (22.2) | 4 (33.3) | 5 (31.3) |
| Web-App | 9 (12.2) | 10 (16.1) | 6 (15.8) | 1 (3.6) | 4 (22.2) | 1 (8.3) | 1 (6.3) |
| Mobile-App | 19 (25.7) | 15 (24.2) | 7 (18.4) | 12 (42.9) | 5 (27.8) | 3 (25.0) | 5 (31.3) |
| Biofeedback devices | 10 (13.5) | 9 (14.5) | 5 (13.2) | 3 (10.7) | 1 (5.6) | 0 (0.0) | 1 (6.3) |
| VR | 17 (23.0) | 9 (14.5) | 7 (18.4) | 5 (17.9) | 2 (11.1) | 3 (25.0) | 3 (18.8) |
| Exergaming | 7 (9.5) | 6 (9.7) | 6 (15.8) | 0 (0.0) | 2 (11.1) | 1 (8.3) | 1 (6.3) |
| **Intervention duration** |  |  |  |  |  |  |  |
| <4 weeks | 11 (14.9) | 10 (16.1) | 6 (15.8) | 3 (10.7) | 1 (5.6) | 2 (16.7) | 2 (12.5) |
| 4 to <8 weeks | 18 (24.3) | 16 (25.8) | 7 (18.4) | 7 (25.0) | 6 (33.3) | 2 (16.7) | 3 (18.8) |
| 8 to <12 weeks | 26 (35.1) | 21 (33.9) | 16 (42.1) | 9 (32.1) | 6 (33.3) | 6 (50.0) | 8 (50.0) |
| ≥12 weeks | 17 (23.0) | 15 (24.2) | 9 (23.7) | 8 (28.6) | 5 (27.8) | 1 (8.3) | 2 (12.5) |
| NR | 2 (2.7) | 0 (0.0) | 0 (0.0) | 1 (3.6) | 0 (0.0) | 1 (8.3) | 1 (6.3) |
| **Studies with durations of follow-up:** |  |  |  |  |  |  |  |
| Post-intervention | 72 (97.3) | 60 (96.8) | 37 (97.4) | 27 (96.4) | 18 (100.0) | 10 (83.3) | 14 (87.5) |
| Short-term | 16 (21.6) | 11 (17.7) | 9 (23.7) | 6 (21.4) | 5 (27.8) | 4 (33.3) | 5 (31.3) |
| Mid-term | 8 (10.8) | 5 (8.1) | 5 (13.2) | 4 (14.3) | 4 (22.2) | 2 (16.7) | 3 (18.8) |
| Long-term | 3 (4.1) | 2 (3.2) | 2 (5.3) | 0 (0.0) | 0 (0.0) | 0 (0.0) | 1 (6.3) |
| **Range of mean age (years);** | 20.5–67.8; | 20.5–70.3; | 20.51–67.8; | 21.3–67.6; | 39.5–67.8; | 41.1–59.6; | 38.3–67.8; |
| **No. of studies (%)** | 70 (94.6) | 60 (96.8) | 37 (97.4) | 26 (92.9) | 17 (94.4) | 11 (91.7) | 15 (93.8) |
|  | 0–100; | 0–100; | 0–100; | 0–100; | 12.6–100; | 50–82.5; | 44.44–82.5; |
|  | 68 (91.9) | 56 (90.3) | 36 (94.7) | 28 (100.0) | 18 (100.0) | 11 (91.7) | 15 (93.8) |

PR-FA=pain-related fear avoidance; HR-QoL=health-related quality of life; VR=virtual reality; Web-App=web applications; Mobile-App=mobile applications; NR=not reported. Data are number of studies (%) unless otherwise indicated. Percentages are calculated using the number of trials contributing to the corresponding outcome as the denominator. Post-intervention was defined as <2 months after treatment, short-term as ≥2 to <6 months, mid-term as ≥6 to <12 months, and long-term as ≥12 months. Ranges of mean age and proportion of female participants were calculated among studies reporting the corresponding characteristic; the accompanying number of studies (%) indicates the availability of these data within each outcome-specific set of trials.

**Figure 1:**
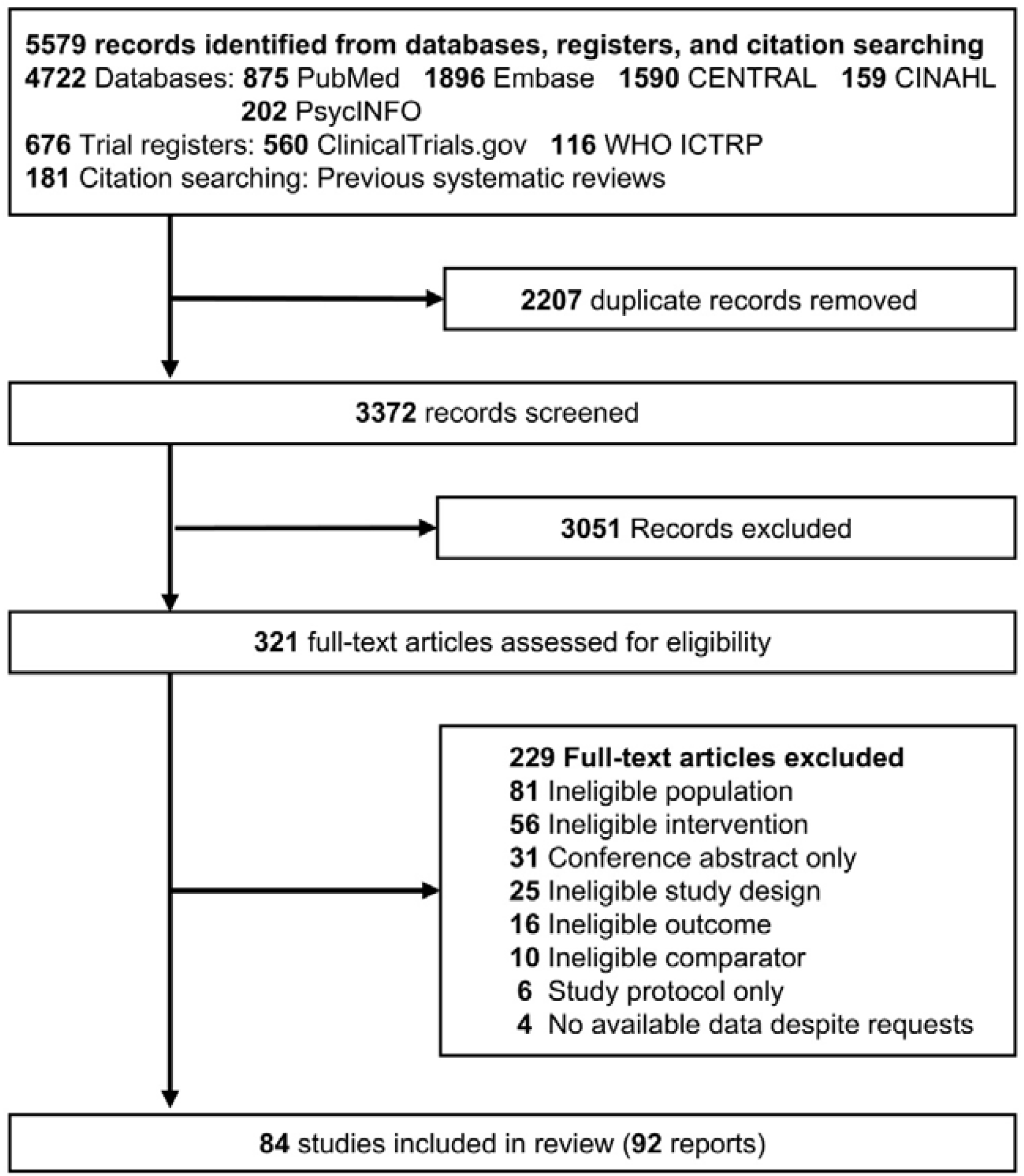
PRISMA flow diagram of study selection. Flow diagram showing the identification, screening, eligibility assessment, and inclusion of studies evaluating digital health interventions for low back pain. A total of 84 randomised controlled trials reported in 92 publications were included in the systematic review. PRISMA=Preferred Reporting Items for Systematic Reviews and Meta-Analyses; CENTRAL=Cochrane Central Register of Controlled Trials; CINAHL=Cumulative Index to Nursing and Allied Health Literature; ICTRP=International Clinical Trials Registry Platform.

**Figure 2:**
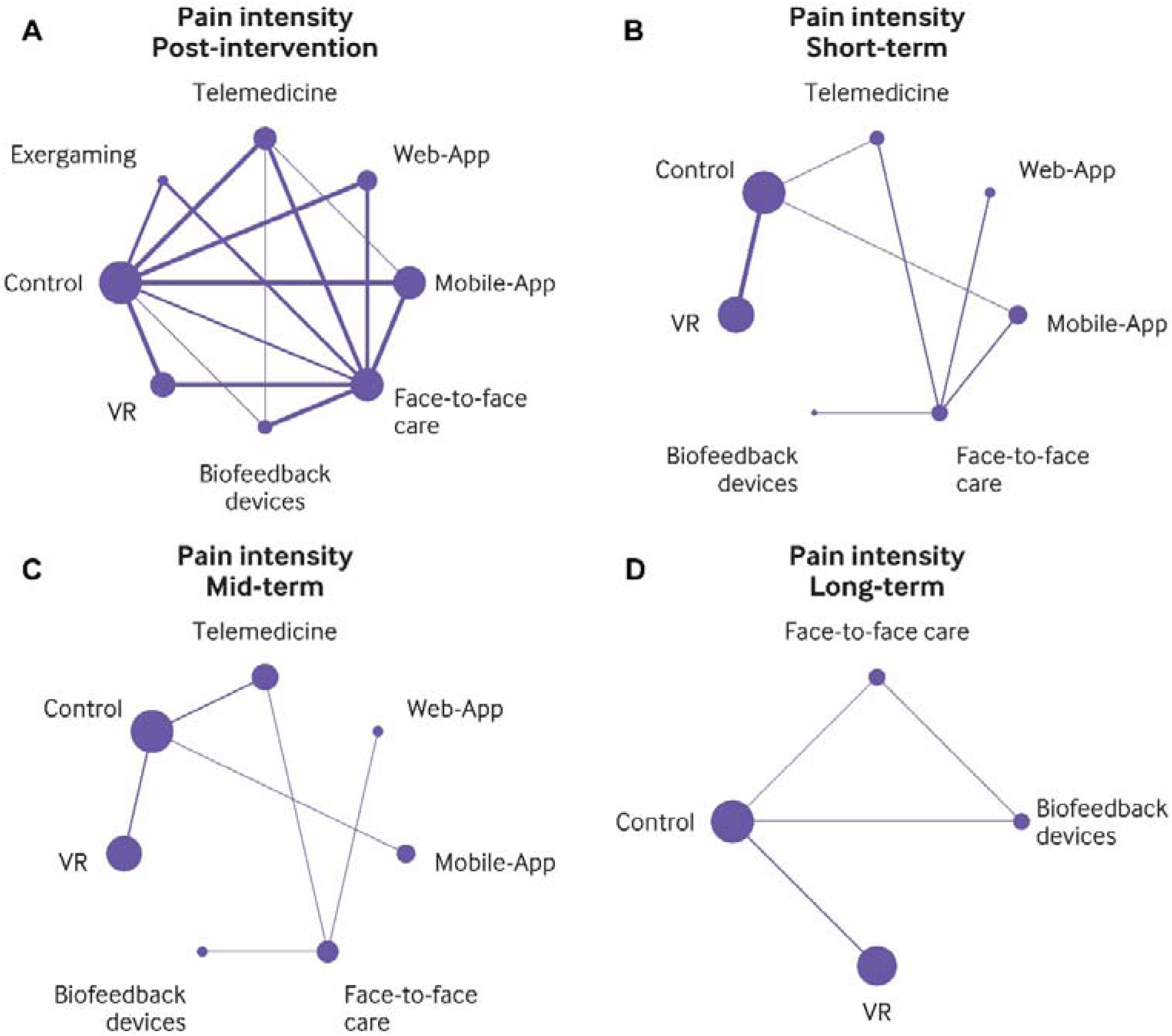
Network plots for pain intensity. Network plots are presented for post-intervention (A), short-term (B), mid-term (C), and long-term (D) follow-up. Lines connect interventions compared directly in at least one randomised trial, with line width proportional to the number of trials contributing to each direct comparison. Nodes represent intervention categories, with node size proportional to the number of participants. VR=virtual reality; Web-App=web applications; Mobile-App=mobile applications.

**Figure 3:**
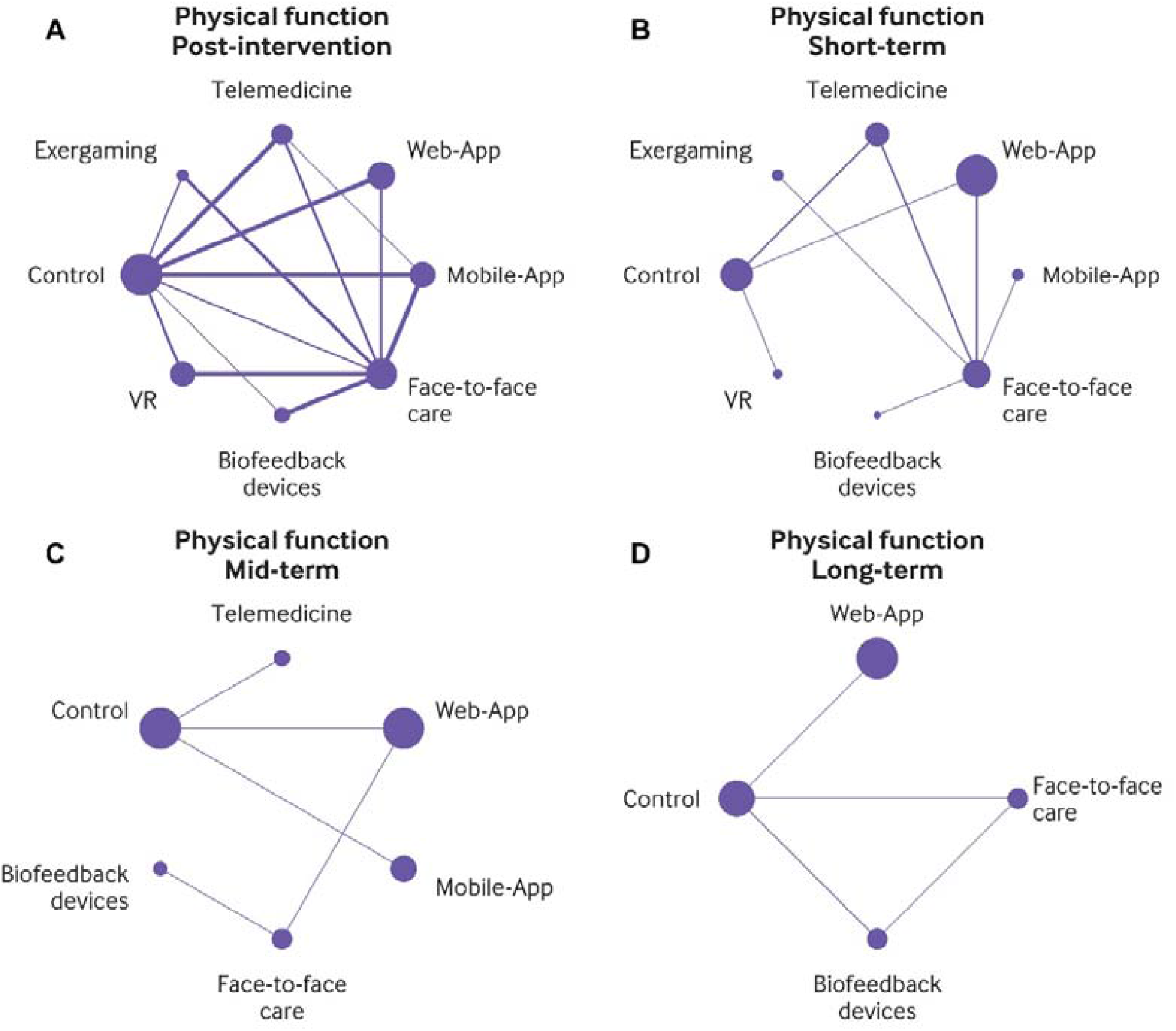
Network plots for physical function. Network plots are presented for post-intervention (A), short-term (B), mid-term (C), and long-term (D) follow-up. Lines connect interventions compared directly in at least one randomised trial, with line width proportional to the number of trials contributing to each direct comparison. Nodes represent intervention categories, with node size proportional to the number of participants. VR=virtual reality; Web-App=web applications; Mobile-App=mobile applications.

**Figure 4:**
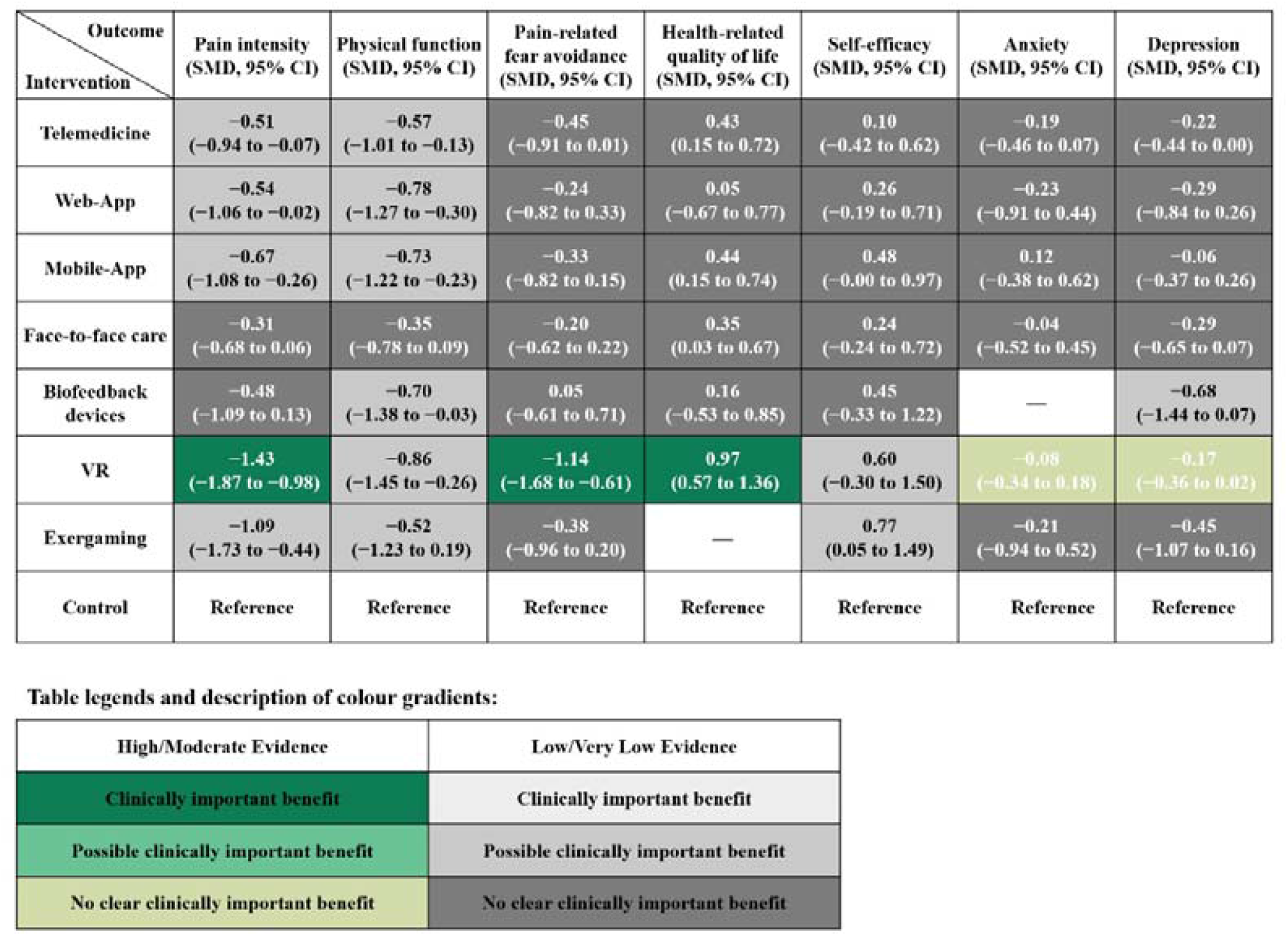
Comparative intervention effects and confidence in the evidence at post-intervention. Network meta-analysis estimates are presented as standardised mean differences (SMDs) with 95% confidence intervals (CIs), using control as the reference intervention. Negative SMDs favour the intervention for pain intensity, physical function, pain-related fear avoidance, anxiety, and depression, whereas positive SMDs favour the intervention for health-related quality of life and self-efficacy. Potential clinical importance was interpreted using an absolute SMD threshold of 0·50. Cell shading reflects potential clinical importance and confidence in the network evidence. Dashes indicate unavailable estimates. VR=virtual reality; Mobile-App=mobile applications; Web-App=web applications.

At post-intervention, VR probably reduced pain intensity compared with control (SMD −1.43, 95% CI −1.87 to −0.98; moderate confidence). Estimates also favoured lower pain intensity with exergaming (−1.09, −1.73 to −0.44), Mobile-App (−0.67, −1.08 to −0.26), Web-App (−0.54, −1.06 to −0.02), and telemedicine (−0.51, −0.94 to −0.07), but confidence in the evidence was very low. Estimates for the remaining interventions were uncertain. For post-intervention physical function, estimates favoured better physical function for VR (SMD −0.86, 95% CI −1.45 to −0.26), Web-App (−0.78, −1.27 to −0.30), Mobile-App (−0.73, −1.22 to −0.23), biofeedback devices (−0.70, −1.38 to −0.03), and telemedicine (−0.57, −1.01 to −0.13), but confidence in the evidence was very low. Estimates for the remaining interventions were uncertain, with low confidence in the evidence (Fig 4).

For post-intervention pain-related fear avoidance, VR probably reduced pain-related fear avoidance compared with control (SMD −1.14, 95% CI −1.68 to −0.61; moderate confidence). Estimates also favoured Mobile-App and face-to-face care, with low confidence; confidence for the remaining comparisons was very low. For health-related quality of life, VR probably improved health-related quality of life compared with control (SMD 0.97, 95% CI 0.57 to 1.36; moderate confidence). Estimates also favoured Mobile-App (0.44, 0.15 to 0.74), telemedicine (0.43, 0.15 to 0.72), and face-to-face care (0.35, 0.03 to 0.67), although confidence in the evidence was very low. For self-efficacy, Exergaming favoured better self-efficacy (SMD 0.77, 95% CI 0.05 to 1.49), whereas the estimate for Mobile-App (0.48, 0.00 to 0.97) included the null; confidence was very low for both comparisons. Estimates for the remaining interventions were imprecise, with CIs including the null (Fig 4). For anxiety and depression, connected networks were available only at post-intervention, with no convincing evidence of benefit across digital health interventions (Fig 4).

Sensitivity analyses excluding studies at high overall risk of bias and those with statistically estimated or converted outcome data were conducted across connected outcome networks. The direction of estimates was generally similar to that in the primary analyses, although exclusion of studies reduced network connectivity and precision for several sparse secondary-outcome networks (appendix pp 141–165). In restricted-population analyses including only participants with chronic low back pain, the direction of post-intervention treatment effects was generally consistent with the primary analyses, although some estimates were less precise (appendix pp 166–178).

## Discussion

In this systematic review and network meta-analysis of 84 randomised controlled trials involving 11 803 participants, the comparative effects of digital health interventions varied across outcomes and follow-up periods. At post-intervention, compared with control, VR showed the most favourable estimates across pain intensity, physical function, pain-related fear avoidance, and health-related quality of life, with moderate confidence for pain intensity, pain-related fear avoidance, and health-related quality of life, but very low confidence for physical function. Other digital modalities showed favourable estimates in a narrower range of outcomes: Mobile-App and telemedicine favoured lower pain intensity, better physical function, and better health-related quality of life; Web-App and biofeedback devices mainly favoured better physical function; and exergaming favoured lower pain intensity. Beyond post-intervention, favourable estimates were less consistent across outcomes and follow-up periods. Longer-term networks were increasingly sparse, and most mid-term and long-term estimates were not formally assessed with CINeMA, precluding conclusions about sustained comparative superiority.

Several features may help explain the modality-specific patterns observed in our analysis. VR combines immersive experience with therapeutic strategies such as distraction, pain education, behavioural skills training, and movement-based rehabilitation.^11,40,41^ These features may reduce attention to pain and threat and facilitate graded exposure, potentially contributing to the favourable post-intervention estimates for pain intensity and pain-related fear avoidance, although the independent contribution of immersion remains uncertain. Other digital modalities, including Mobile-App, Web-App, telemedicine, and biofeedback devices, share therapeutic components such as exercise, education, monitoring, feedback, and clinician support.^21,42,43^ The RESTORE trial found that adding movement-sensor biofeedback to cognitive functional therapy did not improve outcomes beyond the therapy itself,^44^ illustrating that adding technology does not necessarily provide incremental benefit. Together, these findings suggest that modality-level differences should be interpreted in the context of the therapeutic content delivered, rather than attributed to the technological platform alone.

Our findings are broadly consistent with previous evidence on digital interventions for low back pain. A meta-analysis of VR-based training reported improvements in pain, pain-related fear avoidance, and disability immediately after treatment, whereas effects at 3–6 months remained inconclusive.^15^ Reviews of mobile health, mobile apps, and telehealth have reported mixed findings for pain and disability, with benefits varying across interventions and assessment periods.^12–14^ Previous syntheses have evaluated digital interventions across broader populations with chronic or musculoskeletal pain,^20,21^ whereas a recent network meta-analysis focused on low back pain but compared only AI-assisted telerehabilitation, conventional telerehabilitation, in-person care, and usual care.^16^ Our study extends this evidence by comparing six digital modalities across seven clinical outcomes and multiple follow-up periods. Although VR showed favourable estimates across several outcomes at post-intervention, these findings do not establish its sustained superiority over other modalities. Limited direct comparisons, sparse longer-term networks, and predominantly low or very low confidence in the evidence constrain comparative treatment recommendations.

Current guidelines recommend education, exercise, psychological therapies, and supported self-management as core components of non-pharmacological care for low back pain,^45,46^ while WHO guidance emphasises a person-centred, multimodal approach.^47^ However, guidance on selecting digital modalities to deliver these interventions remains limited. Previous systematic reviews have emphasised the importance of considering intervention reach, uptake, feasibility, and implementation readiness alongside clinical effectiveness when evaluating digital health interventions.^48,49^ Our findings suggest that no single digital modality can currently be recommended as universally preferable. Clinical decisions should therefore consider the therapeutic content and certainty of comparative effects alongside accessibility, costs, digital literacy, and patient preferences, which may influence the uptake and sustained use of digital interventions.^50–52^

To our knowledge, this is the most comprehensive network meta-analysis comparing six digital health modalities for low back pain across seven outcomes and multiple follow-up periods. We used a modality-based classification and systematically assessed transitivity, heterogeneity, inconsistency, and confidence in network estimates using CINeMA. Several limitations arose from the primary trials. First, study populations were clinically heterogeneous in terms of pain duration (acute, subacute, chronic, or mixed) and clinical classification, which may have contributed to variation in treatment effects. Second, comparator conditions varied substantially, ranging from waiting-list or minimal-care controls to usual care and structured face-to-face rehabilitation, which may have influenced relative effect estimates. Third, many trials raised risk-of-bias concerns. Blinding was often infeasible, while missing data, selective reporting, and deviations from intended interventions further reduced confidence in the evidence. These limitations were partly mitigated by rigorous risk-of-bias assessment, sensitivity analyses, and evaluation of transitivity, inconsistency, and confidence in the evidence. Fourth, evidence on adverse events, acceptability, treatment discontinuation, and treatment burden was limited or not synthesised, preventing a complete assessment of the balance between benefit and burden.

Several limitations also arose from the network meta-analysis itself. First, substantial heterogeneity was present in several networks, and global inconsistency was detected in the post-intervention pain-intensity network, with local disagreement for selected exergaming comparisons; these findings warrant cautious interpretation. Second, direct comparisons between digital modalities were limited, and networks became increasingly sparse at longer follow-up, reducing the precision of long-term estimates. Third, analyses relied on aggregate study-level data, and interventions within each modality node varied in therapeutic content, intensity, duration, and clinician support, limiting our ability to explore treatment-effect modifiers and disentangle the effects of the technological platform from those of specific therapeutic components. Fourth, although SMDs allowed outcomes measured with different instruments to be combined, they are less intuitive clinically. Back-transformed estimates for pain intensity and physical function depended on the selected reference standard deviations and should not be interpreted as instrument-specific minimally important differences.

Future research on digital health for low back pain should focus on four priorities. First, adequately powered head-to-head trials with longer follow-up are needed. Second, factorial or component-based studies should distinguish the effects of therapeutic content, clinician support, treatment intensity, and technological features. Third, broader outcomes beyond pain and function, including anxiety, depression, sleep, and long-term improvement, should be evaluated in people with low back pain.^53^ These outcomes are clinically relevant because low back pain is often accompanied by psychological and behavioural comorbidities that may also be influenced by digital interventions. Finally, pragmatic studies should assess cost-effectiveness, accessibility, digital exclusion, and implementation in routine care.

Digital health interventions for low back pain showed effects that varied across outcomes and follow-up periods. Virtual reality had the most favourable post-intervention estimates, with moderate-confidence evidence for improvements in pain intensity, pain-related fear avoidance, and health-related quality of life. Although longer-term effects and the relative benefits of different modalities remain uncertain, these findings highlight the potential of digital interventions for low back pain management and the need for adequately powered head-to-head trials with longer follow-up.

## Data Availability

Data used for the analyses are available from the corresponding author on reasonable request. The R code for the network meta-analysis is publicly available at https://github.com/Zhuotao-Wu/digital-health-low-back-pain-nma

## Contributors

RC, ZW, XW, and WY contributed equally to this work and are joint first authors. DM and CC jointly supervised the study and are joint corresponding authors. RC, ZW, XW, WY, HX, JS, HA, FF, DM, and CC conceived and designed the study. RC, ZW, XW, and CY screened and selected studies. RC, ZW, XW, WY, CY, and HC extracted data. RC, ZW, XW, and XYZ assessed risk of bias. RC and ZW performed the statistical analyses. RC and WY directly accessed and verified the underlying data. RC, ZW, XW, and WY conducted the CINeMA assessments, which were reviewed by HX, JS, HA, FF, and CC. WY, CH, JS, HA, FF, DM, and CC provided methodological input. CC, TX, HX, XZZ, HA, FF, and SZ contributed to interpretation of the findings. RC, ZW, XW, WY, JZ, JS, and CC drafted the manuscript. HX, CH, TX, SZ, JS, HA, FF, DM, and CC critically revised the manuscript. All authors critically reviewed the manuscript for important intellectual content, approved the final version, and agreed to be accountable for all aspects of the work.

## Data sharing

The datasets generated from this review are available from the corresponding author upon request.

## Acknowledgments

We thank the librarians at West China Hospital, Sichuan University, for their assistance with literature retrieval. We also thank Yang Yang and Sarah L. Krein for providing additional unpublished data and clarifications on request. This work was supported by the National Key Research and Development Programme of China (grant 2025YFC3607901). During the preparation of this work, JS used ChatGPT by OpenAI to improve the readability and language of the manuscript. After using this tool, JS reviewed and edited the content as needed and takes full responsibility for the content of the publication.

