## Supplementary Materials for "Comparative effectiveness of digital health interventions for low back pain: systematic review and network meta-analysis of randomised controlled trials"

**Section 1.** PRISMA checklists

**1.1.** PRISMA 2020 checklist

| Section and Topic | Item # | Checklist item | Location where item is reported |
|---|---|---|---|
| **TITLE** | | | |
| Title | 1 | Identify the report as a systematic review. | Manuscript p 1 |
| **ABSTRACT** | | | |
| Abstract | 2 | See the PRISMA 2020 for Abstracts checklist. | Manuscript p 3; appendix pp 13–14 |
| **INTRODUCTION** | | | |
| Rationale | 3 | Describe the rationale for the review in the context of existing knowledge. | Manuscript pp 3–4 (Introduction) |
| Objectives | 4 | Provide an explicit statement of the objective(s) or question(s) the review addresses. | Manuscript p 4 (Introduction) |
| **METHODS** | | | |
| Eligibility criteria | 5 | Specify the inclusion and exclusion criteria for the review and how studies were grouped for the syntheses. | Manuscript pp 4–5; appendix pp 57–72 |
| Information sources | 6 | Specify all databases, registers, websites, organisations, reference lists and other sources searched or consulted to identify studies. Specify the date when each source was last searched or consulted. | Manuscript pp 3–4 (Summary; Methods) |
| Search strategy | 7 | Present the full search strategies for all databases, registers and websites, including any filters and limits used. | Appendix pp 17–35 (search strategies) |
| Selection process | 8 | Specify the methods used to decide whether a study met the inclusion criteria of the review, including how many reviewers screened each record and each report retrieved, whether they worked independently, and if applicable, details of automation tools used in the process. | Manuscript p 5 |

| Section and Topic | Item # | Checklist item | Location where item is reported |
|---|---|---|---|
| Data collection process | 9 | Specify the methods used to collect data from reports, including how many reviewers collected data from each report, whether they worked independently, any processes for obtaining or confirming data from study investigators, and if applicable, details of automation tools used in the process. | Manuscript p 5 |
| Data items | 10a | List and define all outcomes for which data were sought. Specify whether all results that were compatible with each outcome domain in each study were sought (e.g. for all measures, time points, analyses), and if not, the methods used to decide which results to collect. | Manuscript p 5; appendix pp 82–87 |
| Data items | 10b | List and define all other variables for which data were sought (e.g. participant and intervention characteristics, funding sources). Describe any assumptions made about any missing or unclear information. | Manuscript p 5; appendix pp 73–81 |
| Study risk of bias assessment | 11 | Specify the methods used to assess risk of bias in the included studies, including details of the tool(s) used, how many reviewers assessed each study and whether they worked independently, and if applicable, details of automation tools used in the process. | Manuscript p 5; appendix pp 88–91 |
| Effect measures | 12 | Specify for each outcome the effect measure(s) (e.g. risk ratio, mean difference) used in the synthesis or presentation of results. | Manuscript p 5 |
| Synthesis methods | 13a | Describe the processes used to decide which studies were eligible for each synthesis (e.g. tabulating the study intervention characteristics and comparing against the planned groups for each synthesis (item #5)). | Manuscript pp 4–6; appendix pp 57–72 |
| Synthesis methods | 13b | Describe any methods required to prepare the data for presentation or synthesis, such as handling of missing summary statistics, or data conversions. | Manuscript pp 5–6 |

| Section and Topic | Item # | Checklist item | Location where item is reported |
|---|---|---|---|
| Synthesis methods | 13c | Describe any methods used to tabulate or visually display results of individual studies and syntheses. | Manuscript pp 6–9; figs 2–4; appendix pp 99–129 |
| Synthesis methods | 13d | Describe any methods used to synthesize results and provide a rationale for the choice(s). If meta-analysis was performed, describe the model(s), method(s) to identify the presence and extent of statistical heterogeneity, and software package(s) used. | Manuscript pp 5–6; appendix pp 130–136 |
| Synthesis methods | 13e | Describe any methods used to explore possible causes of heterogeneity among study results (e.g. subgroup analysis, meta-regression). | Manuscript p 6; appendix pp 91–98, 130–136, 169–181 |
| Synthesis methods | 13f | Describe any sensitivity analyses conducted to assess robustness of the synthesized results. | Manuscript p 6; appendix pp 144–168 |
| Reporting bias assessment | 14 | Describe any methods used to assess risk of bias due to missing results in a synthesis (arising from reporting biases). | Manuscript p 6; appendix pp 137–143, 182–191 |
| Certainty assessment | 15 | Describe any methods used to assess certainty (or confidence) in the body of evidence for an outcome. | Manuscript p 6; appendix pp 182–191 |
| **RESULTS** | | | |
| Study selection | 16a | Describe the results of the search and selection process, from the number of records identified in the search to the number of studies included in the review, ideally using a flow diagram. | Manuscript p 7; fig 1 (p 16) |
| Study selection | 16b | Cite studies that might appear to meet the inclusion criteria, but which were excluded, and explain why they were excluded. | Appendix pp 39–56 (excluded reports; reasons require checking) |
| Study characteristics | 17 | Cite each included study and present its characteristics. | Manuscript pp 7–8; table 1 (p 8); appendix pp 73–81 |

| Section and Topic | Item # | Checklist item | Location where item is reported |
|---|---|---|---|
| Risk of bias in studies | 18 | Present assessments of risk of bias for each included study. | Manuscript p 7; appendix pp 88–91 |
| Results of individual studies | 19 | For all outcomes, present, for each study: (a) summary statistics for each group (where appropriate) and (b) an effect estimate and its precision (e.g. confidence/credible interval), ideally using structured tables or plots. | Not separately reported as study-level arm data; appendix provides network estimates |
| Results of syntheses | 20a | For each synthesis, briefly summarise the characteristics and risk of bias among contributing studies. | Manuscript pp 7–8; table 1; appendix pp 73–81, 88–91 |
| Results of syntheses | 20b | Present results of all statistical syntheses conducted. If meta-analysis was done, present for each the summary estimate and its precision (e.g. confidence/credible interval) and measures of statistical heterogeneity. If comparing groups, describe the direction of the effect. | Manuscript pp 7–9; fig 4; appendix pp 105–136 |
| Results of syntheses | 20c | Present results of all investigations of possible causes of heterogeneity among study results. | Manuscript pp 6–7, 9; appendix pp 91–98, 130–136, 169–181 |
| Results of syntheses | 20d | Present results of all sensitivity analyses conducted to assess the robustness of the synthesized results. | Manuscript p 9; appendix pp 144–168 |
| Reporting biases | 21 | Present assessments of risk of bias due to missing results (arising from reporting biases) for each synthesis assessed. | Manuscript p 7; appendix pp 137–143, 182–191 |
| Certainty of evidence | 22 | Present assessments of certainty (or confidence) in the body of evidence for each outcome assessed. | Manuscript pp 3, 7–9; fig 4; appendix pp 182–191 |
| **DISCUSSION** | | | |
| Discussion | 23a | Provide a general interpretation of the results in the context of other evidence. | Manuscript pp 9–10 |

| Section and Topic | Item # | Checklist item | Location where item is reported |
| --- | --- | --- | --- |
| Discussion | 23b | Discuss any limitations of the evidence included in the review. | Manuscript pp 10–11 |
| Discussion | 23c | Discuss any limitations of the review processes used. | Manuscript pp 10–11 (limitations; review-process limitations not explicit) |
| Discussion | 23d | Discuss implications of the results for practice, policy, and future research. | Manuscript pp 10–11 |
| **OTHER INFORMATION** | | | |
| Registration and protocol | 24a | Provide registration information for the review, including register name and registration number, or state that the review was not registered. | Manuscript pp 3–4 (PROSPERO CRD420261339841) |
| Registration and protocol | 24b | Indicate where the review protocol can be accessed, or state that a protocol was not prepared. | Manuscript p 4 (registration cited; protocol access not specified) |
| Registration and protocol | 24c | Describe and explain any amendments to information provided at registration or in the protocol. | Appendix p 16 |
| Support | 25 | Describe sources of financial or non-financial support for the review, and the role of the funders or sponsors in the review. | Manuscript pp 3, 6, 11 |
| Competing interests | 26 | Declare any competing interests of review authors. | Manuscript p 11 |
| Availability of data, code and other materials | 27 | Report which of the following are publicly available and where they can be found: template data collection forms; data extracted from included studies; data used for all analyses; analytic code; any other materials used in the review. | Manuscript p 11 (data on request; code/materials availability not specified) |

**1.2.** PRISMA 2020 for Abstracts checklist

| Section and Topic | Item # | Checklist item | Reported (Yes/No) |
|---|---|---|---|
| **TITLE** | | | |
| Title | 1 | Identify the report as a systematic review. | Yes |
| **BACKGROUND** | | | |
| Objectives | 2 | Provide an explicit statement of the main objective(s) or question(s) the review addresses. | Yes |
| **METHODS** | | | |
| Eligibility criteria | 3 | Specify the inclusion and exclusion criteria for the review. | Yes |
| Information sources | 4 | Specify the information sources (e.g. databases, registers) used to identify studies and the date when each was last searched. | Yes |
| Risk of bias | 5 | Specify the methods used to assess risk of bias in the included studies. | Yes |
| Synthesis of results | 6 | Specify the methods used to present and synthesise results. | Yes |
| **RESULTS** | | | |
| Included studies | 7 | Give the total number of included studies and participants and summarise relevant characteristics of studies. | Yes |
| Synthesis of results | 8 | Present results for main outcomes, preferably indicating the number of included studies and participants for each. If meta-analysis was done, report the summary estimate and confidence/credible interval. If comparing groups, indicate the direction of the effect (i.e. which group is favoured). | Yes |
| **DISCUSSION** | | | |
| Limitations of evidence | 9 | Provide a brief summary of the limitations of the evidence included in the review (e.g. study risk of bias, inconsistency and imprecision). | Yes |
| Interpretation | 10 | Provide a general interpretation of the results and important implications. | Yes |
| **OTHER** | | | |

| Section and Topic | Item # | Checklist item | Reported (Yes/No) |
|---|---|---|---|
| Funding | 11 | Specify the primary source of funding for the review. | Yes |
| Registration | 12 | Provide the register name and registration number. | Yes |

**Section 2.** Protocol deviations

(1) The PROSPERO registration specified broad categories of digital health interventions. For the network meta-analysis, these interventions were subsequently operationalised into eight treatment nodes according to the principal mode of digital delivery. This refinement was introduced to provide mutually interpretable treatment categories for quantitative synthesis and did not alter the underlying review question.

(2) The registered secondary outcomes included pain interference, psychological outcomes, self-efficacy, pain catastrophizing, and quality of life. In the final review, these domains were operationalised as pain-related fear avoidance, health-related quality of life, self-efficacy, anxiety, and depression according to the available validated measures and the outcome-selection rules described in Supplementary 6.

(3) The PROSPERO registration did not specify follow-up categories or a primary follow-up time point. For the final analysis, follow-up assessments were classified as post-intervention, short-term, mid-term, and long-term, with post-intervention designated as the primary time point, to harmonise heterogeneous assessment schedules across trials.

(4) The registered protocol indicated that subgroup analyses according to low back pain duration (acute, subacute, and chronic) would be conducted if sufficient data were available. Because the available networks did not support reliable comparative subgroup analyses across these categories, we instead conducted a restricted analysis including only trials of chronic low back pain. This analysis was interpreted as a restricted-population analysis rather than evidence of effect modification.

(5) The registered protocol proposed treatment rankings using ranking probabilities or SUCRA. These rankings were not reported in the final review because several networks were sparse and many network estimates had low or very low confidence; treatment rankings could therefore give an unwarranted impression of a stable treatment hierarchy. Interpretation instead focused on treatment effect estimates, their 95% confidence intervals, and CINeMA confidence ratings.

(6) Additional post-registration methodological specifications were introduced during development of the final analytic plan, including sensitivity analyses excluding studies at high risk of bias and studies requiring statistical estimation or conversion of outcome data, and explicit decision thresholds for CINeMA and interpretation of standardised effects. These additions were intended to improve the robustness and interpretability of the analyses and did not change the primary review question or outcomes.

**Section 3.** Search strategy and study selection

**Table S1.** Search strategies

| PubMed | | |
|---|---|---|
| **ID** | **Query** | **Records** |
| #1 | ((((((((((((((((((((((((((((((((((((((((((((((((low back pain[MeSH Terms]) OR (low back pain[Other Term])) OR (back pain, low[Other Term])) OR (back pains, low[Other Term])) OR (low back pains[Other Term])) OR (Pain, Low Back[Other Term])) OR (pains, low back[Other Term])) OR (low back ache[Other Term])) OR (ache, low back[Other Term])) OR (aches, low back[Other Term])) OR (back ache, low[Other Term])) OR (back aches, low[Other Term])) OR (low back aches[Other Term])) OR (low backache[Other Term])) OR (backache, low[Other Term])) OR (backaches, low[Other Term])) OR (low backaches[Other Term])) OR (lower back pain[Other Term])) OR (back pain, lower[Other Term])) OR (back pains, lower[Other Term])) OR (lower back pains[Other Term])) OR (pain, lower back[Other Term])) OR (pains, lower back[Other Term])) OR (lumbago[Other Term])) OR (low back pain, mechanical[Other Term])) OR (mechanical low back pain, posterior compartment[Other Term])) OR (low back pain, postural[Other Term])) OR (postural low back pain, recurrent[Other Term])) OR (recurrent low back pain[Other Term])) OR (acute low back pain[Other Term])) OR (chronic low back pain[Other Term]) ) OR (loin pain[Other Term])) OR (low backpain[Other Term])) OR (lowback pain[Other Term])) OR (lumbal pain[Other Term])) OR (lumbal syndrome[Other Term])) OR (lumbalgia[Other Term])) OR (lumbar pain[Other Term])) OR (lumbar spine syndrome[Other Term])) OR (lumbosacral pain[Other Term])) OR (lumbosacral root syndrome[Other Term])) OR (lumbosacroiliac strain[Other Term])) OR (pain, lumbosacral[Other Term])) OR (strain, lumbosacroiliac[Other Term])) OR (low back pain, posterior compartment[Other Term])) OR (low back pain, recurrent[Other Term])) OR (mechanical low back pain[Other Term])) OR (postural low back pain[Other Term])) OR ((((((((((((((((((((((((((((((((((((((((((((((((low back pain[MeSH Terms]) OR (low back pain[Title/Abstract])) OR (back pain, low[Title/Abstract])) OR (back pains, low[Title/Abstract])) OR (low back pains[Title/Abstract])) OR (Pain, Low Back[Title/Abstract])) OR (pains, low back[Title/Abstract])) OR (low back ache[Title/Abstract])) OR (ache, low back[Title/Abstract])) OR (aches, low back[Title/Abstract])) OR (back ache, low[Title/Abstract])) OR (back aches, low[Title/Abstract])) OR (low back aches[Title/Abstract])) OR (low backache[Title/Abstract])) OR (backache, low[Title/Abstract])) OR (backaches, low[Title/Abstract])) OR (low backaches[Title/Abstract])) OR (lower back pain[Title/Abstract])) OR (back pain, lower[Title/Abstract])) OR (back pains, lower[Title/Abstract])) OR (lower back pains[Title/Abstract])) OR (pain, lower back[Title/Abstract])) OR (pains, lower back[Title/Abstract])) OR (lumbago[Title/Abstract])) OR (low back pain, mechanical[Title/Abstract])) OR (mechanical low back pain, posterior compartment[Title/Abstract])) OR (low back pain, postural[Title/Abstract])) OR (postural low back pain, recurrent[Title/Abstract])) OR (recurrent | 64738 |

| | | |
|---|---|---|
| | low back pain[Title/Abstract])) OR (acute low back pain[Title/Abstract])) OR (chronic low back pain[Title/Abstract]) ) OR (loin pain[Title/Abstract])) OR (low backpain[Title/Abstract])) OR (lowback pain[Title/Abstract])) OR (lumbal pain[Title/Abstract])) OR (lumbal syndrome[Title/Abstract])) OR (lumbalgia[Title/Abstract])) OR (lumbar pain[Title/Abstract])) OR (lumbar spine syndrome[Title/Abstract])) OR (lumbosacral pain[Title/Abstract])) OR (lumbosacral root syndrome[Title/Abstract])) OR (lumbosacroiliac strain[Title/Abstract])) OR (pain, lumbosacral[Title/Abstract])) OR (strain, lumbosacroiliac[Title/Abstract])) OR (low back pain, posterior compartment[Title/Abstract])) OR (low back pain, recurrent[Title/Abstract])) OR (mechanical low back pain[Title/Abstract])) OR (postural low back pain[Title/Abstract])) | |
| #2 | ((((((((((((((((back pain[MeSH Terms]) OR (back pains[Other Term])) OR (pain, back[Other Term])) OR (pains, back[Other Term])) OR (back ache[Other Term])) OR (ache, back[Other Term])) OR (aches, back[Other Term])) OR (back aches[Other Term])) OR (backache[Other Term])) OR (backaches[Other Term])) OR (back pain with radiation[Other Term])) OR (back pain without radiation[Other Term])) OR (vertebrogenic pain syndrome[Other Term])) OR (pain syndromes, vertebrogenic[Other Term])) OR (pain syndrome, vertebrogenic[Other Term])) OR (syndromes, vertebrogenic pain[Other Term])) OR (syndrome, vertebrogenic pain[Other Term])) OR (vertebrogenic pain syndromes[Other Term])) OR ((((((((((((((((back pain[MeSH Terms]) OR (back pains[Title/Abstract])) OR (pain, back[Title/Abstract])) OR (pains, back[Title/Abstract])) OR (back ache[Title/Abstract])) OR (ache, back[Title/Abstract])) OR (aches, back[Title/Abstract])) OR (back aches[Title/Abstract])) OR (backache[Title/Abstract])) OR (backaches[Title/Abstract])) OR (back pain with radiation[Title/Abstract])) OR (back pain without radiation[Title/Abstract])) OR (vertebrogenic pain syndrome[Title/Abstract])) OR (pain syndromes, vertebrogenic[Title/Abstract])) OR (pain syndrome, vertebrogenic[Title/Abstract])) OR (syndromes, vertebrogenic pain[Title/Abstract])) OR (syndrome, vertebrogenic pain[Title/Abstract])) OR (vertebrogenic pain syndromes[Title/Abstract])) | 59542 |
| #3 | #1 OR #2 | 82109 |
| #4 | ((((((Digital Health[MeSH Terms]) OR (Health, Digital[Other Term])) OR (Digital Health Technology[Other Term])) OR (Digital Health Technologies[Other Term])) OR (Health Technologies, Digital[Other Term])) OR (Health Technology, Digital[Other Term])) OR ((((((Digital Health[MeSH Terms]) OR (Health, Digital[Title/Abstract])) OR (Digital Health Technology[Title/Abstract])) OR (Digital Health Technologies[Title/Abstract])) OR (Health Technologies, Digital[Title/Abstract])) OR (Health Technology, Digital[Title/Abstract])) | 13434 |
| #5 | ((((((((((((((((((((((((((((Mobile-Applications[MeSH Terms]) OR (Mobile-Applications[Other Term])) OR (application, mobile[Other Term])) OR (applications, mobile[Other Term])) OR (Mobile-Application[Other Term])) OR (Mobile-Apps[Other Term])) OR (app, mobile[Other Term])) OR (apps, mobile[Other Term])) OR (Mobile-App[Other Term])) OR (portable electronic apps[Other Term])) OR (app, portable electronic[Other Term])) OR (electronic app, portable[Other Term])) OR (portable electronic app[Other Term])) OR (portable electronic applications[Other | 34677 |

| | | |
|---|---|---|
| | Term])) OR (application, portable electronic[Other Term])) OR (electronic application, portable[Other Term])) OR (portable electronic application[Other Term])) OR (portable software apps[Other Term])) OR (app, portable software[Other Term])) OR (portable software app[Other Term])) OR (software app, portable[Other Term])) OR (portable software applications[Other Term])) OR (application, portable software[Other Term])) OR (portable software application[Other Term])) OR (software application, portable[Other Term])) OR (smartphone apps[Other Term])) OR (app, smartphone[Other Term])) OR (apps, smartphone[Other Term])) OR (smartphone app[Other Term])) OR (tablet application[Other Term])) OR ((((((((((((((((((((((((((((((Mobile-Applications[MeSH Terms]) OR (Mobile-Applications[Title/Abstract])) OR (application, mobile[Title/Abstract])) OR (applications, mobile[Title/Abstract])) OR (Mobile-Application[Title/Abstract])) OR (Mobile-Apps[Title/Abstract])) OR (app, mobile[Title/Abstract])) OR (apps, mobile[Title/Abstract])) OR (Mobile-App[Title/Abstract])) OR (portable electronic apps[Title/Abstract])) OR (app, portable electronic[Title/Abstract])) OR (electronic app, portable[Title/Abstract])) OR (portable electronic app[Title/Abstract])) OR (portable electronic applications[Title/Abstract])) OR (application, portable electronic[Title/Abstract])) OR (electronic application, portable[Title/Abstract])) OR (portable electronic application[Title/Abstract])) OR (portable software apps[Title/Abstract])) OR (app, portable software[Title/Abstract])) OR (portable software app[Title/Abstract])) OR (software app, portable[Title/Abstract])) OR (portable software applications[Title/Abstract])) OR (application, portable software[Title/Abstract])) OR (portable software application[Title/Abstract])) OR (software application, portable[Title/Abstract])) OR (smartphone apps[Title/Abstract])) OR (app, smartphone[Title/Abstract])) OR (apps, smartphone[Title/Abstract])) OR (smartphone app[Title/Abstract])) OR (tablet application[Title/Abstract])) | |
| #6 | ((((((((((((((((((telemedicine[MeSH Terms]) OR (telemedicine[Other Term])) OR (tele medicine[Other Term])) OR (virtual medicine[Other Term])) OR (medicine, virtual[Other Term])) OR (tele-referral[Other Term])) OR (tele referral[Other Term])) OR (mobile health[Other Term])) OR (health, mobile[Other Term])) OR (mhealth[Other Term])) OR (telehealth[Other Term])) OR (ehealth[Other Term])) OR (tele-intensive care[Other Term])) OR (tele intensive care[Other Term])) OR (tele-icu[Other Term])) OR (tele icu[Other Term])) OR (telecare[Other Term])) OR (tele-care[Other Term])) OR (tele care[Other Term])) OR ((((((((((((((((((telemedicine[MeSH Terms]) OR (telemedicine[Title/Abstract])) OR (tele medicine[Title/Abstract])) OR (virtual medicine[Title/Abstract])) OR (medicine, virtual[Title/Abstract])) OR (tele-referral[Title/Abstract])) OR (tele referral[Title/Abstract])) OR (mobile health[Title/Abstract])) OR (health, mobile[Title/Abstract])) OR (mhealth[Title/Abstract])) OR (telehealth[Title/Abstract])) OR (ehealth[Title/Abstract])) OR (tele-intensive care[Title/Abstract])) OR (tele intensive care[Title/Abstract])) OR (tele-icu[Title/Abstract])) OR (tele icu[Title/Abstract])) OR (telecare[Title/Abstract])) OR (tele-care[Title/Abstract])) OR (tele care[Title/Abstract])) | 114739 |
| #7 | (((((((((Remote Consultation[MeSH Terms]) OR (remote consultation[Other Term])) OR (consultation, remote[Other Term])) OR | 10855 |

| | | |
|---|---|---|
| | (teleconsultation[Other Term])) OR (teleconsultations[Other Term])) OR (long distance consultation[Other Term])) OR (tele−consultation[Other Term])) OR (telephone consultation[Other Term])) OR (telephone−based consultation[Other Term])) OR (((((((((Remote Consultation[MeSH Terms]) OR (remote consultation[Title/Abstract])) OR (consultation, remote[Title/Abstract])) OR (teleconsultation[Title/Abstract])) OR (teleconsultations[Title/Abstract])) OR (long distance consultation[Title/Abstract])) OR (tele−consultation[Title/Abstract])) OR (telephone consultation[Title/Abstract])) OR (telephone−based consultation[Title/Abstract])) | |
| #8 | (((((((((((((((((((((((((((((((((((cell phone[MeSH Terms]) OR (cell phone[Other Term])) OR (phone, cell[Other Term])) OR (phones, cell[Other Term])) OR (cellular phone[Other Term])) OR (cellular phones[Other Term])) OR (phone, cellular[Other Term])) OR (phones, cellular[Other Term])) OR (telephone, cellular[Other Term])) OR (cellular telephone[Other Term])) OR (cellular telephones[Other Term])) OR (telephones, cellular[Other Term])) OR (cell phones[Other Term])) OR (mobile phone[Other Term])) OR (mobile phones[Other Term])) OR (phone, mobile[Other Term])) OR (phones, mobile[Other Term])) OR (car phone[Other Term])) OR (car phones[Other Term])) OR (phone, car[Other Term])) OR (phones, car[Other Term])) OR (mobile telephone[Other Term])) OR (mobile telephones[Other Term])) OR (telephone, mobile[Other Term])) OR (telephones, mobile[Other Term])) OR (portable cellular phone[Other Term])) OR (cellular phone, portable[Other Term])) OR (cellular phones, portable[Other Term])) OR (portable cellular phones[Other Term])) OR (transportable cellular phone[Other Term])) OR (cellular phones, transportable[Other Term])) OR (cellular phone, transportable[Other Term])) OR (transportable cellular phones[Other Term])) OR (cellphone[Other Term])) OR (cellphones[Other Term])) OR (((((((((((((((((((((((((((((((((((cell phone[MeSH Terms]) OR (cell phone[Title/Abstract])) OR (phone, cell[Title/Abstract])) OR (phones, cell[Title/Abstract])) OR (cellular phone[Title/Abstract])) OR (cellular phones[Title/Abstract])) OR (phone, cellular[Title/Abstract])) OR (phones, cellular[Title/Abstract])) OR (telephone, cellular[Title/Abstract])) OR (cellular telephone[Title/Abstract])) OR (cellular telephones[Title/Abstract])) OR (telephones, cellular[Title/Abstract])) OR (cell phones[Title/Abstract])) OR (mobile phone[Title/Abstract])) OR (mobile phones[Title/Abstract])) OR (phone, mobile[Title/Abstract])) OR (phones, mobile[Title/Abstract])) OR (car phone[Title/Abstract])) OR (car phones[Title/Abstract])) OR (phone, car[Title/Abstract])) OR (phones, car[Title/Abstract])) OR (mobile telephone[Title/Abstract])) OR (mobile telephones[Title/Abstract])) OR (telephone, mobile[Title/Abstract])) OR (telephones, mobile[Title/Abstract])) OR (portable cellular phone[Title/Abstract])) OR (cellular phone, portable[Title/Abstract])) OR (cellular phones, portable[Title/Abstract])) OR (portable cellular phones[Title/Abstract])) OR (transportable cellular phone[Title/Abstract])) OR (cellular phones, transportable[Title/Abstract])) OR (cellular phone, transportable[Title/Abstract])) OR (transportable cellular phones[Title/Abstract])) OR (cellphone[Title/Abstract])) OR (cellphones[Title/Abstract])) | 44944 |
| #9 | ((((((smartphone[MeSH Terms]) OR (smartphone[Other Term])) OR (smartphones[Other Term])) OR (smart phone[Other Term])) OR (phones, | 40500 |

| | | |
|---|---|---|
| | smart[Other Term])) OR (smart phones[Other Term])) OR ((((((smartphone[MeSH Terms]) OR (smartphone[Title/Abstract])) OR (smartphones[Title/Abstract])) OR (smart phone[Title/Abstract])) OR (phones, smart[Title/Abstract])) OR (smart phones[Title/Abstract])) | |
| #10 | (((((((internet[MeSH Terms]) OR (world wide web[Other Term])) OR (web, world wide[Other Term])) OR (wide web, world[Other Term])) OR (cyberspace[Other Term])) OR (cyber space[Other Term])) OR (internet connection[Other Term])) OR (((((((internet[MeSH Terms]) OR (world wide web[Title/Abstract])) OR (web, world wide[Title/Abstract])) OR (wide web, world[Title/Abstract])) OR (cyberspace[Title/Abstract])) OR (cyber space[Title/Abstract])) OR (internet connection[Title/Abstract])) | 116564 |
| #11 | ((((((Videoconferencing[MeSH Terms]) OR (videoconferencing[Other Term])) OR (videoconference[Other Term])) OR (videoconferences[Other Term])) OR (video conference[Other Term])) OR (video conferences[Other Term])) OR ((((((Videoconferencing[MeSH Terms]) OR (videoconferencing[Title/Abstract])) OR (videoconference[Title/Abstract])) OR (videoconferences[Title/Abstract])) OR (video conference[Title/Abstract])) OR (video conferences[Title/Abstract])) | 8120 |
| #12 | (((((((((((telecommunications[MeSH Terms]) OR (telecommunications[Other Term])) OR (telecommunication[Other Term])) OR (teleconference[Other Term])) OR (teleconferences[Other Term])) OR (telegraphy[Other Term])) OR (broadcasting[Other Term])) OR (broadcasting industry[Other Term])) OR (radar[Other Term])) OR (radar wave[Other Term])) OR (satellite communications[Other Term])) OR (((((((((((telecommunications[MeSH Terms]) OR (telecommunications[Title/Abstract])) OR (telecommunication[Title/Abstract])) OR (teleconference[Title/Abstract])) OR (teleconferences[Title/Abstract])) OR (telegraphy[Title/Abstract])) OR (broadcasting[Title/Abstract])) OR (broadcasting industry[Title/Abstract])) OR (radar[Title/Abstract])) OR (radar wave[Title/Abstract])) OR (satellite communications[Title/Abstract])) | 167672 |
| #13 | ((((((((((((((((VR[MeSH Terms]) OR (VR[Other Term])) OR (reality, virtual[Other Term])) OR (VR, educational[Other Term])) OR (educational virtual realities[Other Term])) OR (educational reality, educational virtual[Other Term])) OR (virtual realities, educational[Other Term])) OR (VR, instructional[Other Term])) OR (instructional virtual realities[Other Term])) OR (instructional realities, instructional virtual[Other Term])) OR (reality, instructional virtual[Other Term])) OR (virtual realities, instructional[Other Term])) OR (educational VR[Other Term])) OR (instructional VR[Other Term])) OR (realities, instructional virtual[Other Term])) OR (reality, educational virtual[Other Term])) OR ((((((((((((((((VR[MeSH Terms]) OR (VR[Title/Abstract])) OR (reality, virtual[Title/Abstract])) OR (VR, educational[Title/Abstract])) OR (educational virtual realities[Title/Abstract])) OR (educational reality, educational virtual[Title/Abstract])) OR (virtual realities, educational[Title/Abstract])) OR (VR, instructional[Title/Abstract])) OR (instructional virtual realities[Title/Abstract])) OR (instructional realities, instructional virtual[Title/Abstract])) OR (reality, instructional virtual[Title/Abstract])) OR (virtual realities, instructional[Title/Abstract])) OR | 30615 |

| | | |
|---|---|---|
| | (educational VR[Title/Abstract])) OR (instructional VR[Title/Abstract])) OR (realities, instructional virtual[Title/Abstract])) OR (reality, educational virtual[Title/Abstract])) | |
| #14 | (((((((((((((((((((((((((Wearable Electronic Devices[MeSH Terms]) OR (wearable electronic devices[Other Term])) OR (device, wearable electronic[Other Term])) OR (electronic device, wearable[Other Term])) OR (wearable electronic device[Other Term])) OR (wearable devices[Other Term])) OR (device, wearable[Other Term])) OR (wearable device[Other Term])) OR (wearable technology[Other Term])) OR (technology, wearable[Other Term])) OR (wearable technologies[Other Term])) OR (electronic skin[Other Term])) OR (skin, electronic[Other Term])) OR (wearable computer[Other Term])) OR (computer, wearable[Other Term])) OR (wearable computers[Other Term])) OR (electronic wearable[Other Term])) OR (electronic wearables[Other Term])) OR (wearable electronic[Other Term])) OR (wearable electronical device[Other Term])) OR (wearable electronics[Other Term])) OR (devices, wearable[Other Term])) OR (devices, wearable electronic[Other Term])) OR (electronic devices, wearable[Other Term])) OR (technologies, wearable[Other Term])) OR (((((((((((((((((((((((((Wearable Electronic Devices[MeSH Terms]) OR (wearable electronic devices[Title/Abstract])) OR (device, wearable electronic[Title/Abstract])) OR (electronic device, wearable[Title/Abstract])) OR (wearable electronic device[Title/Abstract])) OR (wearable devices[Title/Abstract])) OR (device, wearable[Title/Abstract])) OR (wearable device[Title/Abstract])) OR (wearable technology[Title/Abstract])) OR (technology, wearable[Title/Abstract])) OR (wearable technologies[Title/Abstract])) OR (electronic skin[Title/Abstract])) OR (skin, electronic[Title/Abstract])) OR (wearable computer[Title/Abstract])) OR (computer, wearable[Title/Abstract])) OR (wearable computers[Title/Abstract])) OR (electronic wearable[Title/Abstract])) OR (electronic wearables[Title/Abstract])) OR (wearable electronic[Title/Abstract])) OR (wearable electronical device[Title/Abstract])) OR (wearable electronics[Title/Abstract])) OR (devices, wearable[Title/Abstract])) OR (devices, wearable electronic[Title/Abstract])) OR (electronic devices, wearable[Title/Abstract])) OR (technologies, wearable[Title/Abstract])) | 50127 |
| #15 | ((((((((((((((((((((((((((((((((((((Computers, Handheld[MeSH Terms]) OR (computers, handheld[Other Term])) OR (computer, handheld[Other Term])) OR (handheld computer[Other Term])) OR (palmtop computer[Other Term])) OR (palmtop computers[Other Term])) OR (computers, palm top[Other Term])) OR (palm-top computer[Other Term])) OR (palm-top computers[Other Term])) OR (personal digital assistant[Other Term])) OR (digital assistant, personal[Other Term])) OR (pda computer[Other Term])) OR (computer, pda[Other Term])) OR (computers, pda[Other Term])) OR (pda computers[Other Term])) OR (palm pilot[Other Term])) OR (palm pilots[Other Term])) OR (pilot, palm[Other Term])) OR (pilots, palm[Other Term])) OR (pocket pc[Other Term])) OR (pc, pocket[Other Term])) OR (pcs, pocket[Other Term])) OR (pocket pcs[Other Term])) OR (tablet computers[Other Term])) OR (computers, tablet[Other Term])) OR (computer, tablet[Other Term])) OR (tablet computer[Other Term])) OR (mobile devices[Other Term])) OR (device, mobile[Other Term])) OR (devices, mobile[Other Term])) OR (mobile | 32944 |

| | | |
|---|---|---|
| | device[Other Term])) OR (hand held computer[Other Term])) OR (handheld computers[Other Term])) OR (palm PC[Other Term])) OR (personal data assistant[Other Term])) OR (pocket computer[Other Term])) OR (pocket−sized computer[Other Term])) OR (((((((((((((((((((((((((((((((((((((Computers, Handheld[MeSH Terms]) OR (computers, handheld[Title/Abstract])) OR (computer, handheld[Title/Abstract])) OR (handheld computer[Title/Abstract])) OR (palmtop computer[Title/Abstract])) OR (palmtop computers[Title/Abstract])) OR (computers, palm top[Title/Abstract])) OR (palm-top computer[Title/Abstract])) OR (palm-top computers[Title/Abstract])) OR (personal digital assistant[Title/Abstract])) OR (digital assistant, personal[Title/Abstract])) OR (pda computer[Title/Abstract])) OR (computer, pda[Title/Abstract])) OR (computers, pda[Title/Abstract])) OR (pda computers[Title/Abstract])) OR (palm pilot[Title/Abstract])) OR (palm pilots[Title/Abstract])) OR (pilot, palm[Title/Abstract])) OR (pilots, palm[Title/Abstract])) OR (pocket pc[Title/Abstract])) OR (pc, pocket[Title/Abstract])) OR (pcs, pocket[Title/Abstract])) OR (pocket pcs[Title/Abstract])) OR (tablet computers[Title/Abstract])) OR (computers, tablet[Title/Abstract])) OR (computer, tablet[Title/Abstract])) OR (tablet computer[Title/Abstract])) OR (mobile devices[Title/Abstract])) OR (device, mobile[Title/Abstract])) OR (devices, mobile[Title/Abstract])) OR (mobile device[Title/Abstract])) OR (hand held computer[Title/Abstract])) OR (handheld computers[Title/Abstract])) OR (palm PC[Title/Abstract])) OR (personal data assistant[Title/Abstract])) OR (pocket computer[Title/Abstract])) OR (pocket−sized computer[Title/Abstract])) | |
| #16 | (((((((((((((((((((((Artificial Intelligence[MeSH Terms]) OR (artificial intelligence[Other Term])) OR (intelligence, artificial[Other Term])) OR (computer reasoning[Other Term])) OR (reasoning, computer[Other Term])) OR (ai (artificial intelligence[Other Term]))) OR (machine intelligence[Other Term])) OR (intelligence, machine[Other Term])) OR (computational intelligence[Other Term])) OR (intelligence, computational[Other Term])) OR (computer vision systems[Other Term])) OR (computer vision system[Other Term])) OR (system, computer vision[Other Term])) OR (systems, computer vision[Other Term])) OR (vision system, computer[Other Term])) OR (vision systems, computer[Other Term])) OR (knowledge acquisition (computer[Other Term]))) OR (acquisition, knowledge (computer[Other Term]))) OR (knowledge representation (computer[Other Term]))) OR (knowledge representations (computer[Other Term]))) OR (representation, knowledge (computer[Other Term]))) OR (((((((((((((((((((((Artificial Intelligence[MeSH Terms]) OR (artificial intelligence[Title/Abstract])) OR (intelligence, artificial[Title/Abstract])) OR (computer reasoning[Title/Abstract])) OR (reasoning, computer[Title/Abstract])) OR (ai (artificial intelligence[Title/Abstract]))) OR (machine intelligence[Title/Abstract])) OR (intelligence, machine[Title/Abstract])) OR (computational intelligence[Title/Abstract])) OR (intelligence, computational[Title/Abstract])) OR (computer vision systems[Title/Abstract])) OR (computer vision system[Title/Abstract])) OR (system, computer vision[Title/Abstract])) OR (systems, computer vision[Title/Abstract])) OR (vision system, computer[Title/Abstract])) OR (vision systems, computer[Title/Abstract])) OR (knowledge acquisition (computer[Title/Abstract]))) OR | 391482 |

| | | |
|---|---|---|
| | (acquisition, knowledge (computer[Title/Abstract]))) OR (knowledge representation (computer[Title/Abstract]))) OR (knowledge representations (computer[Title/Abstract]))) OR (representation, knowledge (computer[Title/Abstract]))) | |
| #17 | ((((((Telephone[MeSH Terms]) OR (telephone[Other Term])) OR (switch board service[Other Term])) OR (Service, switch board[Other Term])) OR (Services, switch board[Other Term])) OR (switch board services[Other Term])) OR ((((((Telephone[MeSH Terms]) OR (telephone[Title/Abstract])) OR (switch board service[Title/Abstract])) OR (Service, switch board[Title/Abstract])) OR (Services, switch board[Title/Abstract])) OR (switch board services[Title/Abstract])) | 111258 |
| #18 | ((((((((Text Messaging[MeSH Terms]) OR (Messaging, Text[Other Term])) OR (Texting[Other Term])) OR (Short Message Service[Other Term])) OR (Text Messages[Other Term])) OR (Messages, Text[Other Term])) OR (Message, Text[Other Term])) OR (Text Message[Other Term])) OR ((((((((Text Messaging[MeSH Terms]) OR (Messaging, Text[Title/Abstract])) OR (Texting[Title/Abstract])) OR (Short Message Service[Title/Abstract])) OR (Text Messages[Title/Abstract])) OR (Messages, Text[Title/Abstract])) OR (Message, Text[Title/Abstract])) OR (Text Message[Title/Abstract])) | 10917 |
| #19 | ((((((((Augmented Reality[MeSH Terms]) OR (Augmented Realities[Other Term])) OR (Realities, Augmented[Other Term])) OR (Reality, Augmented[Other Term])) OR (Mixed Reality[Other Term])) OR (Mixed Realities[Other Term])) OR (Realities, Mixed[Other Term])) OR (Reality, Mixed[Other Term])) OR ((((((((Augmented Reality[MeSH Terms]) OR (Augmented Realities[Title/Abstract])) OR (Realities, Augmented[Title/Abstract])) OR (Reality, Augmented[Title/Abstract])) OR (Mixed Reality[Title/Abstract])) OR (Mixed Realities[Title/Abstract])) OR (Realities, Mixed[Title/Abstract])) OR (Reality, Mixed[Title/Abstract])) | 8648 |
| #20 | (((((((((((exergaming[MeSH Terms]) OR (VR Exercise[Other Term])) OR (Exercises, VR[Other Term])) OR (Exercise, VR[Other Term])) OR (VR Exercises[Other Term])) OR (Active-Video Gaming[Other Term])) OR (Active Video Gaming[Other Term])) OR (Gaming, Active-Video[Other Term])) OR (Gamings, Active-Video[Other Term])) OR (Exergames[Other Term])) OR (Exergame[Other Term])) OR (((((((((((exergaming[MeSH Terms]) OR (VR Exercise[Title/Abstract])) OR (Exercises, VR[Title/Abstract])) OR (Exercise, VR[Title/Abstract])) OR (VR Exercises[Title/Abstract])) OR (Active-Video Gaming[Title/Abstract])) OR (Active Video Gaming[Title/Abstract])) OR (Gaming, Active-Video[Title/Abstract])) OR (Gamings, Active-Video[Title/Abstract])) OR (Exergames[Title/Abstract])) OR (Exergame[Title/Abstract])) | 3292 |
| #21 | (((Remote Patient Monitoring[MeSH Terms]) OR (Monitoring, Remote Patient[Other Term])) OR (Patient Monitoring, Remote[Other Term])) OR (((Remote Patient Monitoring[MeSH Terms]) OR (Monitoring, Remote Patient[Title/Abstract])) OR (Patient Monitoring, Remote[Title/Abstract])) | 4252 |
| #22 | ((((((((((((((((((((((((Biofeedback, Psychology[MeSH Terms]) OR (Biofeedbacks, Psychology[Other Term])) OR (Psychology Biofeedback[Other | 19979 |

| | | |
|---|---|---|
| | Term])) OR (Psychology Biofeedbacks[Other Term])) OR (Biofeedback[Other Term])) OR (Biofeedbacks[Other Term])) OR (Biofeedback (Psychology[Other Term]))) OR (Biofeedbacks (Psychology[Other Term]))) OR (Feedback, Psychophysiologic[Other Term])) OR (Feedback, Psychophysiological[Other Term])) OR (Psychophysiologic Feedback[Other Term])) OR (False Physiological Feedback[Other Term])) OR (False Physiological Feedbacks[Other Term])) OR (Feedback, False Physiological[Other Term])) OR (Feedbacks, False Physiological[Other Term])) OR (Physiological Feedback, False[Other Term])) OR (Physiological Feedbacks, False[Other Term])) OR (Bogus Physiological Feedback[Other Term])) OR (Bogus Physiological Feedbacks[Other Term])) OR (Feedback, Bogus Physiological[Other Term])) OR (Feedbacks, Bogus Physiological[Other Term])) OR (Physiological Feedback, Bogus[Other Term])) OR (Physiological Feedbacks, Bogus[Other Term])) OR (Myofeedback[Other Term])) OR ((((((((((((((((((((((Biofeedback, Psychology[MeSH Terms]) OR (Biofeedbacks, Psychology[Title/Abstract])) OR (Psychology Biofeedback[Title/Abstract])) OR (Psychology Biofeedbacks[Title/Abstract])) OR (Biofeedback[Title/Abstract])) OR (Biofeedbacks[Title/Abstract])) OR (Biofeedback (Psychology[Title/Abstract]))) OR (Biofeedbacks (Psychology[Title/Abstract]))) OR (Feedback, Psychophysiologic[Title/Abstract])) OR (Feedback, Psychophysiological[Title/Abstract])) OR (Psychophysiologic Feedback[Title/Abstract])) OR (False Physiological Feedback[Title/Abstract])) OR (False Physiological Feedbacks[Title/Abstract])) OR (Feedback, False Physiological[Title/Abstract])) OR (Feedbacks, False Physiological[Title/Abstract])) OR (Physiological Feedback, False[Title/Abstract])) OR (Physiological Feedbacks, False[Title/Abstract])) OR (Bogus Physiological Feedback[Title/Abstract])) OR (Bogus Physiological Feedbacks[Title/Abstract])) OR (Feedback, Bogus Physiological[Title/Abstract])) OR (Feedbacks, Bogus Physiological[Title/Abstract])) OR (Physiological Feedback, Bogus[Title/Abstract])) OR (Physiological Feedbacks, Bogus[Title/Abstract])) OR (Myofeedback[Title/Abstract])) | |
| #23 | #4 OR #5 OR #6 OR #7 OR #8 OR #9 OR #10 OR #11 OR #12 OR #13 OR #14 OR #15 OR #16 OR #17 OR #18 OR #19 OR #20 OR #21 OR #22 | 896718 |
| #24 | #3 AND #23 | 2476 |
| #25 | (randomized controlled trial[MeSH Terms]) OR ((((((randomized controlled trial[Publication Type]) OR (controlled clinical trial[Publication Type])) OR (random*[Title/Abstract])) OR (placebo[Title/Abstract])) OR (randomly[Title/Abstract])) OR (trial[Title/Abstract])) | 2522684 |
| #26 | #24 AND #25 | 875 |
| **Embase** | | |
| **ID** | **Query** | **Records** |
| #1 | 'backache'/exp OR 'backache' OR 'back ache' OR 'back pain' OR 'back pain syndrome' OR 'backpain' OR 'dorsalgia' OR 'pain, back' | 190416 |
| #2 | 'low back pain'/exp OR 'low back pain' OR 'acute low back pain' OR 'back pain, low' OR 'chronic low back pain' OR 'loin pain' OR 'low backache' | 106550 |

| | | |
|---|---|---|
| | OR 'low backpain' OR 'lowback pain' OR 'lower back pain' OR 'lumbago' OR 'lumbal pain' OR 'lumbal syndrome' OR 'lumbalgesia' OR 'lumbalgia' OR 'lumbar pain' OR 'lumbar spine syndrome' OR 'lumbodynia' OR 'lumbosacral pain' OR 'lumbosacral root syndrome' OR 'lumbosacroiliac strain' OR 'pain, low back' OR 'pain, lumbosacral' OR 'strain, lumbosacroiliac' | |
| #3 | #1 OR #2 | 192338 |
| #4 | 'digital health'/exp OR 'digital health' | 40890 |
| #5 | 'Mobile-Application'/exp OR 'Mobile-Application' OR 'Mobile-App' OR 'Mobile-Applications' OR 'Mobile-Apps' OR 'portable software app' OR 'portable software application' OR 'portable software applications' OR 'portable software apps' OR 'tablet application' | 46731 |
| #6 | 'telemedicine'/exp OR 'telemedicine' OR 'tele medicine' OR 'virtual medicine' | 115091 |
| #7 | 'teleconsultation'/exp OR 'teleconsultation' OR 'long distance consultation' OR 'remote consultation' OR 'tele-consultation' OR 'telephone consultation' OR 'telephone-based consultation' | 21917 |
| #8 | 'mobile phone'/exp OR 'mobile phone' OR 'cell phone' OR 'cell phones' OR 'cellphone' OR 'cellphones' OR 'cellular phone' OR 'cellular telephone' OR 'mobile telephone' | 84044 |
| #9 | 'smartphone'/exp OR 'smartphone' OR 'smart phone' OR 'smartphones' | 63203 |
| #10 | 'internet'/exp OR 'internet' OR 'internet connection' OR 'world wide web' | 221796 |
| #11 | 'videoconferencing'/exp OR 'videoconferencing' OR 'video conference' OR 'video conferencing' OR 'videoconference' | 16957 |
| #12 | 'telecommunications'/exp OR 'telecommunications' OR 'broadcasting' OR 'broadcasting industry' OR 'radar' OR 'radar wave' OR 'satellite communications' | 203529 |
| #13 | 'VR'/exp OR 'VR' | 54317 |
| #14 | 'wearable electronic device'/exp OR 'wearable electronic device' OR 'electronic wearable' OR 'electronic wearables' OR 'wearable electronic' OR 'wearable electronic devices' OR 'wearable electronical device' OR 'wearable electronics' | 35833 |
| #15 | 'personal digital assistant'/exp OR 'personal digital assistant' OR 'computers, handheld' OR 'hand held computer' OR 'handheld computer' OR 'handheld computers' OR 'palm pc' OR 'palmtop' OR 'personal data assistant' OR 'pocket computer' OR 'pocket-sized computer' | 3422 |
| #16 | 'artificial intelligence'/exp OR 'artificial intelligence' OR 'machine intelligence' | 298034 |
| #17 | 'telephone'/exp OR 'telephone' OR 'dataphone' OR 'telephone line' | 173926 |
| #18 | 'e-mail'/exp OR 'e-mail' OR 'electronic mail' OR 'electronic messaging' OR 'email' OR 'mail, electronic' OR 'mailing, electronic' | 218146 |
| #19 | 'text messaging'/exp OR 'text messaging' OR 'texting' | 14777 |

| | | |
|---|---|---|
| #20 | 'augmented reality'/exp OR 'augmented reality' | 10550 |
| #21 | 'exergaming'/exp OR 'exergaming' OR 'active video gaming' OR 'active videogaming' OR 'exer-gaming' OR 'VR (vr) exergaming' OR 'VR (vr) -based exercise' OR 'VR exercise' OR 'VR-based exercise' OR 'vr exercise' OR 'vr exergaming' OR 'vr-based exercise' | 2015 |
| #22 | 'telemonitoring'/exp OR 'telemonitoring' OR 'distant monitoring (patient)' OR 'distant patient monitoring' OR 'remote distance patient monitoring' OR 'remote monitoring (patient)' OR 'remote patient monitoring' OR 'remote patient surveillance' OR 'tele monitoring' OR 'tele surveillance' OR 'telesurveillance' | 11747 |
| #23 | 'biofeedback'/exp OR 'biofeedback' OR 'bio feed back' OR 'bio-feedback' OR 'biofeed-back' OR 'biofeedback (psychology)' OR 'biofeedback system' OR 'biofeedback, psychology' OR 'myobiofeedback' OR 'myofeedback' OR 'psychophysiological feedback' | 24249 |
| #24 | #4 OR #5 OR #6 OR #7 OR #8 OR #9 OR #10 OR #11 OR #12 OR #13 OR #14 OR #15 OR #16 OR #17 OR #18 OR #19 OR #20 OR #21 OR #22 OR #23 | 1252596 |
| #25 | #3 AND #24 | 6316 |
| #26 | 'randomized controlled trial'/exp OR 'randomized controlled trial' | 1573917 |
| #27 | (randomized:ti,ab,kw AND controlled:ti,ab,kw AND trial:ti,ab,kw OR controlled:ti,ab,kw) AND clinical:ti,ab,kw AND trial:ti,ab,kw OR random*:ti,ab,kw OR randomly:ti,ab,kw OR randomization:ti,ab,kw OR randomisation:ti,ab,kw | 2747859 |
| #28 | #26 OR #27 | 3053397 |
| #29 | #25 AND #28 | 1896 |
| **CENTRAL** | | |
| **ID** | **Search** | **Records** |
| #1 | MeSH descriptor: [Low Back Pain] explode all trees | 6714 |
| #2 | (ache, low back OR aches, low back OR back ache, low OR back aches, low OR back pain, low OR back pain, lower OR back pains, low OR back pains, lower OR backache, low OR backaches, low OR low back ache OR low back aches OR low back pain OR low back pain, mechanical OR low back pain, posterior compartment OR low back pain, postural OR low back pain, recurrent OR low back pains OR low backache OR low backaches OR lower back pain OR lower back pains OR lumbago OR mechanical low back pain OR pain, low back OR pain, lower back OR pains, low back OR pains, lower back OR postural low back pain OR recurrent low back pain):ti,ab,kw | 21239 |
| #3 | MeSH descriptor: [Back Pain] explode all trees | 8219 |
| #4 | (Back Pain without Radiation OR Back Pain with Radiation OR Backaches OR Pain, Back OR Back Pains OR Back Aches OR Ache, Back OR | 29864 |

| | | |
|---|---|---|
| | Backache OR Aches, Back OR Pains, Back OR Back Ache OR Vertebrogenic Pain Syndrome OR Pain Syndrome, Vertebrogenic OR Pain Syndromes, Vertebrogenic OR Vertebrogenic Pain Syndromes OR Syndrome, Vertebrogenic Pain OR Syndromes, Vertebrogenic Pain):ti,ab,kw | |
| #5 | #1 OR #2 OR #3 OR #4 | 30018 |
| #6 | MeSH descriptor: [Digital Technology] explode all trees | 36 |
| #7 | (Technology, Digital OR Technologies, Digital OR Digital Technologies OR Electronics, Digital OR Digital Electronics):ti,ab,kw | 3632 |
| #8 | #6 OR #7 | 3632 |
| #9 | MeSH descriptor: [Mobile-Applications] explode all trees | 3129 |
| #10 | (Mobile-Application OR Apps, Mobile OR Mobile-Apps OR App, Mobile OR Application, Mobile OR Applications, Mobile OR Mobile-App OR Smartphone Apps OR Apps, Smartphone OR App, Smartphone OR Smartphone App OR Portable Electronic Application OR Portable Electronic Apps OR Electronic App, Portable OR App, Portable Electronic OR Portable Electronic Applications OR Application, Portable Electronic OR Electronic Application, Portable OR Portable Electronic App OR Software Application, Portable OR Application, Portable Software OR Portable Software Applications OR Software App, Portable OR Portable Software Apps OR App, Portable Software OR Portable Software App OR Portable Software Application):ti,ab,kw | 15003 |
| #11 | #9 OR #10 | 15003 |
| #12 | MeSH descriptor: [Telemedicine] explode all trees | 6490 |
| #13 | (health, mobile OR mobile health OR telehealth OR telemedicine OR ehealth OR mhealth OR Medicine, Virtual OR Tele-Referrals OR Tele Referral OR Virtual Medicine OR Tele-Referral OR Tele-Intensive Care OR Tele ICU OR Tele Intensive Care OR Tele-ICU OR Telecare OR Tele-Care OR Tele Care):ti,ab,kw | 27577 |
| #14 | #12 OR #13 | 28411 |
| #15 | MeSH descriptor: [Remote Consultation] explode all trees | 498 |
| #16 | (Consultation, Remote OR Teleconsultations OR Teleconsultation OR remote consultation):ti,ab,kw | 1814 |
| #17 | #15 OR #16 | 1840 |
| #18 | MeSH descriptor: [Cell Phone] explode all trees | 4129 |
| #19 | (car phone OR car phones OR cell phone OR cell phones OR cellular phone OR cellular phone, portable OR cellular phone, transportable OR cellular phones OR cellular phones, portable OR cellular phones, transportable OR cellular telephone OR cellular telephones OR mobile phone OR mobile phones OR mobile telephone OR mobile telephones OR phone, car OR phone, cell OR phone, cellular OR phone, mobile OR phones, | 9296 |

| | | |
|---|---|---|
| | car OR phones, cell OR phones, cellular OR phones, mobile OR portable cellular phone OR portable cellular phones OR telephone, cellular OR telephone, mobile OR telephones, cellular OR telephones, mobile OR transportable cellular phone OR transportable cellular phones):ti,ab,kw | |
| #20 | #18 OR #19 | 11906 |
| #21 | MeSH descriptor: [Smartphone] explode all trees | 1482 |
| #22 | (phones, smart OR smart phone OR smart phones OR smartphone OR smartphones):ti,ab,kw | 13365 |
| #23 | #21 OR #22 | 13365 |
| #24 | MeSH descriptor: [Internet] explode all trees | 7354 |
| #25 | (Wide Web, World OR Web, World Wide OR World Wide Web OR Cyberspace OR Cyber Space OR internet):ti,ab,kw | 19528 |
| #26 | #24 OR #25 | 20147 |
| #27 | MeSH descriptor: [Videoconferencing] explode all trees | 419 |
| #28 | (Videoconferencings OR Videoconferences OR Video Conferences OR Videoconference OR Video Conference OR videoconferencing):ti,ab,kw | 5655 |
| #29 | #27 OR #28 | 5697 |
| #30 | MeSH descriptor: [Telecommunications] explode all trees | 13914 |
| #31 | (Teleconference OR Teleconferences OR Telecommunication OR Telegraphy OR Telegraphies OR telecommunications):ti,ab,kw | 1073 |
| #32 | #30 OR #31 | 14733 |
| #33 | MeSH descriptor: [VR] explode all trees | 1972 |
| #34 | (Reality, Educational Virtual OR Virtual Realities, Educational OR Instructional VR OR Reality, Instructional Virtual OR VR, Instructional OR Virtual Realities, Instructional OR Educational Virtual Realities OR Realities, Instructional Virtual OR VR, Educational OR Instructional Virtual Realities OR Educational VR OR Reality, Virtual OR VR):ti,ab,kw | 10777 |
| #35 | #33 OR #34 | 10802 |
| #36 | MeSH descriptor: [Wearable Electronic Devices] explode all trees | 1307 |
| #37 | (Wearable Electronic Device OR Wearable Devices OR Technology, Wearable OR Wearable Technologies OR Wearable Technology OR Wearable Device OR Device, Wearable OR Device, Wearable Electronic OR Electronic Device, Wearable OR Wearable Computers OR Wearable Computer OR Computer, Wearable OR Electronic Skin OR Skin, Electronic OR Electronic Devices, Wearable OR Devices, Wearable Electronic):ti,ab,kw | 4073 |

| | | |
|---|---|---|
| #38 | #36 OR #37 | 4862 |
| #39 | MeSH descriptor: [Computers, Handheld] explode all trees | 1887 |
| #40 | (PCs, Pocket OR Pocket PCs OR Pocket PC OR PC, Pocket OR Computer, Tablet OR Tablet Computer OR Computers, Tablet OR Tablet Computers OR Handheld Computer OR Computer, Handheld OR Handheld Computers OR Palm-Top Computer OR Computers, Palm-Top OR Computers, Palmtop OR Palm-Top Computers OR Palmtop Computers OR Computer, Palmtop OR Palmtop Computer OR Computers, Palm Top OR Computer, Palm-Top OR Mobile Devices OR Devices, Mobile OR Device, Mobile OR Mobile Device OR Pilot, Palm OR Pilots, Palm OR Palm Pilot OR Palm Pilots OR Digital Assistant, Personal OR PDA Computers OR Computers, PDA OR PDA Computer OR Personal Digital Assistant OR Computer, PDA):ti,ab,kw | 6602 |
| #41 | #39 OR #40 | 7941 |
| #42 | MeSH descriptor: [Artificial Intelligence] explode all trees | 4318 |
| #43 | (Vision System, Computer OR Systems, Computer Vision OR Computer Vision System OR Computer Vision Systems OR System, Computer Vision OR Vision Systems, Computer OR AI (Artificial Intelligence) OR Intelligence, Artificial OR Computational Intelligence OR Intelligence, Machine OR Machine Intelligence OR Computer Reasoning OR Intelligence, Computational OR Reasoning, Computer OR Knowledge Acquisition (Computer) OR Acquisition, Knowledge (Computer) OR Knowledge Representation (Computer) OR Representation, Knowledge (Computer) OR Knowledge Representations (Computer) OR artificial intelligence OR computational intelligence OR computer reasoning OR computer vision system OR computer vision systems OR knowledge acquisition (computer) OR knowledge representation (computer) OR knowledge representations (computer) OR machine intelligence):ti,ab,kw | 5599 |
| #44 | #42 OR #43 | 8624 |
| #45 | MeSH descriptor: [Telephone] explode all trees | 6934 |
| #46 | (Telephones OR Switchboard Services OR Services, Switchboard OR Service, Switchboard OR Switchboard Service OR service, switchboard OR services, switchboard OR switchboard service OR switchboard services OR telephone OR telephones):ti,ab,kw | 28023 |
| #47 | #45 OR #46 | 31805 |
| #48 | MeSH descriptor: [Electronic Mail] explode all trees | 450 |
| #49 | (Emails OR Mail, Electronic OR E-Mail OR E-Mails OR Email OR E Mail OR e mail OR e-mail OR e-mails OR electronic mail OR email OR emails OR mail, electronic):ti,ab,kw | 8753 |
| #50 | #48 OR #49 | 8753 |

| | | |
|---|---|---|
| #51 | MeSH descriptor: [Text Messaging] explode all trees | 1937 |
| #52 | (Short Message Service OR Messages, Text OR Text Message OR Message, Text OR Text Messages OR Messaging, Text OR Texting OR Textings OR text messaging):ti,ab,kw | 8747 |
| #53 | #51 OR #52 | 8747 |
| #54 | MeSH descriptor: [Augmented Reality] explode all trees | 166 |
| #55 | (Realities, Augmented OR Realities, Mixed OR Mixed Realities OR Mixed Reality OR Reality, Augmented OR Reality, Mixed OR Augmented Realities OR augmented reality OR mixed reality):ti,ab,kw | 1812 |
| #56 | #54 OR #55 | 1812 |
| #57 | MeSH descriptor: [Exergaming] explode all trees | 102 |
| #58 | VR Exercises OR Exercises, VR OR Exercise, VR OR Active-Video Gamings OR Active Video Gaming OR VR Exercise OR Exergamings OR Active-Video Gaming OR Gamings, Active-Video OR Gaming, Active-Video OR Exergame OR Exergames OR exergaming | 3528 |
| #59 | #57 OR #58 | 3528 |
| #60 | MeSH descriptor: [Biofeedback, Psychology] explode all trees | 2461 |
| #61 | (Myofeedbacks OR Myofeedback OR Feedbacks, Bogus Physiological OR Physiological Feedback, Bogus OR Feedbacks, False Physiological OR False Physiological Feedback OR Feedback, False Physiological OR Physiological Feedbacks, Bogus OR Physiological Feedbacks, False OR Bogus Physiological Feedbacks OR Physiological Feedback, False OR Feedback, Bogus Physiological OR Bogus Physiological Feedback OR False Physiological Feedbacks OR Feedback, Psychophysiologic OR Psychology Biofeedbacks OR Biofeedback OR Biofeedbacks OR Biofeedbacks, Psychology OR Psychology Biofeedback OR Biofeedback (Psychology) OR Feedback, Psychophysiological OR Psychophysiologic Feedback OR Biofeedbacks (Psychology)):ti,ab,kw | 5594 |
| #62 | #60 OR #61 | 6397 |
| #63 | #8 OR #11 OR #14 OR #17 OR #20 OR #23 OR #26 OR #29 OR #32 OR #35 OR #38 OR #41 OR #44 OR #47 OR #50 OR #53 OR #56 OR #59 OR #62 | 126192 |
| #64 | #5 AND #63 | 2153 |
| #65 | MeSH descriptor: [Randomized Controlled Trials as Topic] explode all trees | 66000 |
| #66 | (randomized controlled trial OR controlled clinical trial OR random* OR randomly OR randomization OR randomisation):ti,ab,kw | 1576443 |
| #67 | #65 OR #66 | 1576532 |

| | | |
|---|---|---|
| #68 | #64 AND #67 | 1590 |
| **CINAHL** | | |
| **ID** | **Query** | **Records** |
| #1 | MH "Back Pain" OR ("Back Pains" OR "Backache" OR "Pain, Back" OR "Spinal Pain") | 13885 |
| #2 | MH "Low Back Pain" OR ("Low Back Pains" OR "Lumbago" OR "Pain, Low Back") | 25148 |
| #3 | #1 OR #2 | 37585 |
| #4 | MH "Digital Health" OR Health, Digital | 8642 |
| #5 | MH "Mobile-Applications" OR ("App, Portable Electronic" OR "App, Software" OR "Apps, Portable Electronic" OR "Apps, Software" OR "Mobile-App" OR "Software App" OR "Software Apps" OR "Software Apps, Portable") | 19039 |
| #6 | MH "Telemedicine" | 20361 |
| #7 | MH "Remote Consultation" OR ("Consultation, Remote" OR "Consultations Remote" OR "Remote Consultations" OR "Teleconsultation" OR "Teleconsultations") | 4667 |
| #8 | MH "Cellular Phone" OR ("Car Phone" OR "Car Phones" OR "Cell Phones" OR "Cellular Phones" OR "Cellular Telephone" OR "Cellular Telephones" OR "Mobile Phone" OR "Mobile Phones" OR "Phone, Car" OR "Phone, Cellular" OR "Phone, Mobile" OR "Phones, Car" OR "Phones, Cellular" OR "Phones, Mobile" OR "Telephone, Cellular" OR "Telephones, Cellular") | 7296 |
| #9 | MH "Smartphone" OR ("Android Phone" OR "Smartphones" OR "Windows Phone" OR "iPhone") | 10642 |
| #10 | MH "Internet" | 55857 |
| #11 | MH "Videoconferencing" OR ("Video Conferencing" OR "Videoconference" OR "Videoconferences") | 5951 |
| #12 | MH "Telecommunications" OR ("Broadband Communication Systems" OR "Mobile Communication Systems" OR "Telecommunication" OR "Telecommunication Systems") | 3377 |
| #13 | MH "VR" OR Virtual Realities | 12944 |
| #14 | MH "Electrical Equipment and Supplies" OR ("Electrical Equipment" OR "Electronic Equipment" OR "Electronic Equipment and Supplies" OR "Equipment and Supplies, Electrical") | 820 |
| #15 | MH "Computers, Hand-Held" OR ("Computer, Hand-Held" OR "Computers, Handheld" OR "Computers, Palmtop" OR "Hand-Held Computers" OR "Handheld Computer" OR "Handheld Computers" OR "PDA" OR "Palmtop" OR "Palmtop Computers" OR "Personal Digital Assistant" OR "Personal Digital Assistants") | 7605 |

| | | |
|---|---|---|
| #16 | MH "Artificial Intelligence" OR ("Computer Reasoning" OR "Intelligence, Artificial") | 26220 |
| #17 | MH "Telephone" OR ("Phone" OR "Phones" OR "Telephone Use" OR "Telephones") | 37301 |
| #18 | MH "Text Messaging" OR ("Messaging, Text" OR "Short Message Service" OR "Text Message" OR "Text Messages" OR "Textings") | 6997 |
| #19 | MH "Augmented Reality" | 1232 |
| #20 | MH "Exergames" OR ("Exergame" OR "Exergaming") | 877 |
| #21 | MH "Remote Patient Monitoring" | 519 |
| #22 | MH "Biofeedback" OR ("Feedback, Psychophysiologic" OR "Neurofeedback") | 5104 |
| #23 | #4 OR #5 OR #6 OR #7 OR #8 OR #9 OR #10 OR #11 OR #12 OR #13 OR #14 OR #15 OR #16 OR #17 OR #18 OR #19 OR #20 OR #21 OR #22 | 198023 |
| #24 | #3 AND #23 | 763 |
| #25 | MH "Randomized Controlled Trials" OR ("Clinical Trial, Randomized" OR "Clinical Trials, Controlled, Randomized" OR "Clinical Trials, Randomized" OR "Controlled Clinical Trials, Randomized" OR "Controlled Trials, Randomized" OR "Randomised Controlled Trial" OR "Randomised Controlled Trials" OR "Randomized Clinical Trial" OR "Randomized Clinical Trials" OR "Randomized Controlled Clinical Trials" OR "Randomized Controlled Trial") | 241120 |
| #26 | #24 AND #25 | 159 |
| **PsycINFO** | | |
| **ID** | **Search** | **Records** |
| #1 | exp Back Pain/ | 5332 |
| #2 | (backache* or back ache* or backpain* or back pain* or dorsalgia* or low back pain* or low backache* or low backpain* or lowback pain* or lower back pain* or lumbago* or lumbal pain* or lumbalgia* or lumbar pain* or lumbodynia* or lumbosacral pain* or loin pain*).tw. | 7776 |
| #3 | 1 or 2 | 8048 |
| #4 | exp Telemedicine/ | 19157 |
| #5 | exp Internet/ | 35977 |
| #6 | exp Telephone Systems/ | 12445 |
| #7 | exp Text Messaging/ | 2048 |

| | | |
|---|---|---|
| #8 | exp Computer Mediated Communication/ | 58487 |
| #9 | exp Mobile Devices/ | 14120 |
| #10 | exp Biofeedback/ | 31078 |
| #11 | (digital health or digital healthcare or digital intervention* or digital therap* or digital treatment* or ehealth or e-health or electronic health or mhealth or m-health or mobile health or internet-based intervention* or internet based intervention* or technology-based intervention* or technology based intervention*).tw. | 17831 |
| #12 | (Mobile-App* or Mobile-Application* or smartphone* or smart phone* or mobile phone* or mobile telephone* or cell phone* or cellphone* or cellular phone* or tablet app* or tablet application*).tw. | 23337 |
| #13 | (telemedicine or tele-medicine or telehealth or tele-health or telecare or tele-care or teleconsult* or tele-consult* or remote consult* or distant consult* or virtual consult* or virtual care or telephone consult* or telephone-based consult*).tw. | 11619 |
| #14 | (videoconferenc* or video conferenc* or video consult* or video-based consult* or video visit* or virtual visit*).tw. | 4752 |
| #15 | (internet or online or web-based or web based or web-delivered or web delivered or internet-based or internet based or online intervention* or web intervention* or Web-Application* or Web-App* or website* or web portal*).tw. | 237940 |
| #16 | (VR or virtual realities or virtual environment* or immersive VR or immersive technolog* or VR-based or VR based or VR-based or VR based).tw. | 17631 |
| #17 | (augmented reality or mixed reality or extended reality or AR-based or AR based or XR-based or XR based).tw. | 3030 |
| #18 | (exergam* or exer-gam* or active video gam* or active videogam* or exercise gam* or exercise-based gam* or virtual exercise* or VR exercise* or VR exercise* or VR exergam* or VR exergam*).tw. | 1071 |
| #19 | (wearable* or wearable device* or wearable electronic* or wearable technolog* or wearable sensor* or activity tracker* or fitness tracker* or smart watch* or smartwatch*).tw. | 4503 |
| #20 | (artificial intelligence or machine intelligence or machine learning or AI-based or AI based or intelligent system* or chatbot* or conversational agent*).tw. | 37670 |
| #21 | (telephone* or phone-based or phone based or telephone-based or telephone based).tw. | 30351 |
| #22 | (email* or e-mail* or electronic mail* or electronic messag*).tw. | 14205 |
| #23 | (text messag* or texting or SMS or short message service*).tw. | 6589 |
| #24 | (telemonitor* or tele-monitor* or remote monitor* or remote patient monitor* or distant patient monitor* or remote patient surveillance or | 819 |

| | | |
|---|---|---|
| | telesurveillance).tw. | |
| #25 | (biofeedback or bio-feedback or bio feedback or biofeedback system* or psychophysiological feedback or myobiofeedback or myofeedback).tw. | 6550 |
| #26 | or/4-25 | 720786 |
| #27 | 3 and 26 | 688 |
| #28 | exp Randomized Controlled Trials/ | 1970 |
| #29 | (randomized controlled trial).ti,ab. OR ((((randomized.ti,ab. AND controlled.ti,ab. AND trial.ti,ab.) OR controlled.ti,ab.) AND clinical.ti,ab. AND trial.ti,ab.) OR random*.ti,ab. OR randomly.ti,ab. OR randomization.ti,ab. OR randomisation.ti,ab.) | 285685 |
| #30 | 28 OR 29 | 285775 |
| #31 | 27 AND 30 | 202 |
| **Clinicaltrials.gov** | | |
| **ID** | **Query** | **Records** |
| #1 | ( "low back pain" OR "lower back pain" OR lumbago OR lumbalgia OR "lumbar pain" OR "lumbosacral pain" OR "mechanical low back pain" OR "chronic low back pain" OR "acute low back pain" OR "recurrent low back pain" OR "back pain" OR backache OR "back ache" OR "back pain with radiation" OR "back pain without radiation" OR "vertebrogenic pain syndrome" ) | 3963 |
| #2 | ("digital health" OR "digital health technology" OR "digital health technologies" OR "digital intervention" OR "digital interventions" OR "digital medicine" OR "digital therapeutic" OR "digital therapeutics" OR "connected health" OR "Mobile-App" OR "Mobile-Apps" OR "Mobile-Application" OR "Mobile-Applications" OR "smartphone app" OR "smartphone apps" OR "tablet application" OR app OR apps OR telemedicine OR "tele medicine" OR telehealth OR ehealth OR mhealth OR "mobile health" OR telecare OR "tele care" OR "tele-ICU" OR "tele ICU" OR "virtual care" OR "virtual visit" OR "remote consultation" OR teleconsultation OR teleconsultations OR "telephone consultation" OR "telephone-based consultation" OR "virtual consultation" OR "cell phone" OR "cell phones" OR cellphone OR cellphones OR "mobile phone" OR "mobile phones" OR "cellular phone" OR "cellular phones" OR smartphone OR smartphones OR "smart phone" OR "smart phones" OR "smartphone-based" OR web-based OR "web based" OR webbased OR internet-based OR "internet based" OR online OR "online intervention" OR "online program" OR "internet intervention" OR "web intervention" OR "web-based intervention" OR email OR e-mail OR emails OR "electronic mail" OR videoconferencing OR videoconference OR videoconferences OR "video conference" OR "video conferences" OR "video visit" OR teleconference OR teleconferences OR "VR" OR VR OR "immersive VR" OR "augmented reality" OR AR OR "mixed reality" OR "wearable device" OR "wearable devices" OR "wearable technology" OR "wearable technologies" OR "wearable electronic device" OR "wearable electronic | 82092 |

| | | |
|---|---|---|
| | devices" OR wearables OR "fitness tracker" OR smartwatch OR smartwatches OR "handheld computer" OR "handheld computers" OR "tablet computer" OR "tablet computers" OR "mobile device" OR "mobile devices" OR "personal digital assistant" OR PDA OR tablet OR tablets OR "artificial intelligence" OR AI OR "machine learning" OR "deep learning" OR "computer vision" OR "clinical decision support" OR "decision support system" OR telephone OR phone OR "phone call" OR "telephone call" OR "telephone-based" OR "phone-based" OR "text messaging" OR texting OR "text message" OR "text messages" OR SMS OR "short message service" OR "mobile messaging" OR exergaming OR exergame OR exergames OR "active video gaming" OR "active video game" OR "VR exercise" OR "remote patient monitoring" OR "remote monitoring" OR "home monitoring" OR "patient monitoring" OR biofeedback OR myofeedback OR "physiologic feedback" OR "physiological feedback" OR neurofeedback) | |
| #3 | #1 and #2 | 560 |
| WHO ICTRP | | |
| **ID** | **Query** | **Records** |
| #1 | ("low back pain" OR "lower back pain" OR lumbago OR lumbalgia OR "lumbar pain" OR "lumbosacral pain" OR "mechanical low back pain" OR "chronic low back pain" OR "acute low back pain" OR "recurrent low back pain" OR "back pain" OR backache OR "back ache" OR "back pain with radiation" OR "back pain without radiation" OR "vertebrogenic pain syndrome") | 1335 |
| #2 | ("digital health" OR "digital health technology" OR "digital health technologies" OR "digital intervention" OR "digital interventions" OR "digital medicine" OR "digital therapeutic" OR "digital therapeutics" OR "connected health" OR "Mobile-App" OR "Mobile-Apps" OR "Mobile-Application" OR "Mobile-Applications" OR "smartphone app" OR "smartphone apps" OR "tablet application" OR app OR apps OR telemedicine OR "tele medicine" OR telehealth OR ehealth OR mhealth OR "mobile health" OR telecare OR "tele care" OR "tele-ICU" OR "tele ICU" OR "virtual care" OR "virtual visit" OR "remote consultation" OR teleconsultation OR teleconsultations OR "telephone consultation" OR "telephone-based consultation" OR "virtual consultation") | 2140 |
| #3 | ("cell phone" OR "cell phones" OR cellphone OR cellphones OR"mobile phone" OR "mobile phones" OR "cellular phone" OR "cellular phones" OR smartphone OR smartphones OR "smart phone" OR "smart phones" OR "smartphone-based" OR "web-based" OR "web based" OR webbased OR "internet-based" OR "internet based" OR online OR "online intervention" OR "online program" OR "internet intervention" OR "web intervention" OR "web-based intervention" OR email OR "e-mail" OR emails OR "electronic mail" OR videoconferencing OR videoconference | 5374 |

| | | |
|---|---|---|
| | OR videoconferences OR "video conference" OR "video conferences" OR "video visit" OR teleconference OR teleconferences OR telephone OR phone OR "phone call" OR "telephone call" OR "telephone-based" OR "phone-based" OR "text messaging" OR texting OR "text message" OR "text messages" OR SMS OR "short message service" OR "mobile messaging") | |
| #4 | ("VR" OR VR OR "immersive VR" OR "augmented reality" OR AR OR "mixed reality" OR "wearable device" OR "wearable devices" OR "wearable technology" OR "wearable technologies" OR "wearable electronic device" OR "wearable electronic devices" OR wearables OR "fitness tracker" OR smartwatch OR smartwatches OR "handheld computer" OR "handheld computers" OR "tablet computer" OR "tablet computers" OR "mobile device" OR "mobile devices" OR "personal digital assistant" OR PDA OR tablet OR tablets OR exergaming OR exergame OR exergames OR "active video gaming" OR "active video game" OR "VR exercise") | 15847 |
| #5 | ("artificial intelligence" OR AI OR "machine learning" OR "deep learning" OR "computer vision" OR "clinical decision support" OR "decision support system" OR "remote patient monitoring" OR "remote monitoring" OR "home monitoring" OR "patient monitoring" OR biofeedback OR myofeedback OR "physiologic feedback" OR "physiological feedback" OR neurofeedback) | 1751 |
| #6 | #1 AND #2 | 17 |
| #7 | #1 AND #3 | 36 |
| #8 | #1 AND #4 | 43 |
| #9 | #1 AND #5 | 20 |
| #10 | #6 OR #7 OR #8 OR #9 | 116 |

**Table S2.** Previous systematic reviews and network meta-analyses

| Author–year ID | Title | Population and intervention scope | Number of included RCTs |
|---|---|---|---|
| Dario 2017[1] | Effectiveness of telehealth-based interventions in the management of non-specific low back pain: a systematic review with meta-analysis | People with non-specific low back pain; telehealth interventions delivered alone or as an adjunct to usual care, including telephone, text messaging, applications, websites, and email | 11 RCTs |
| Slattery 2019[2] | An Evaluation of the Effectiveness of the Modalities | Adults with chronic pain; eHealth interventions delivered through | 30 RCTs |

| | | | |
|---|---|---|---|
| | Used to Deliver Electronic Health Interventions for Chronic Pain: Systematic Review With Network Meta-Analysis | modalities including web-based programmes, Mobile-Applications, VR, and other electronic platforms | |
| Du 2020[3] | The efficacy of e-health in the self-management of chronic low back pain: a meta analysis | People with chronic low back pain; eHealth-based self-management programmes, including mobile- and web-based interventions | 8 RCTs |
| Chen 2021[4] | Efficacy of Mobile Health in Patients With Low Back Pain: Systematic Review and Meta-analysis of Randomized Controlled Trials | People with low back pain; mobile health interventions delivered through mobile phones or Mobile-Applications, generally compared with usual care | 9 RCTs |
| Brea-Gómez 2021[5] | VR in the Treatment of Adults with Chronic Low Back Pain: A Systematic Review and Meta-Analysis of Randomized Clinical Trials | Adults with chronic low back pain; VR–based exercise, training, rehabilitation, or behavioural interventions | 14 RCTs |
| Lara-Palomo 2022[6] | Efficacy of e-Health Interventions in Patients with Chronic Low-Back Pain: A Systematic Review with Meta-Analysis | People with chronic low back pain; eHealth interventions compared with physiotherapy, home-based treatment, or minimal intervention | 8 RCTs |
| Valentijn 2022[7] | Digital Health Interventions for Musculoskeletal Pain Conditions: Systematic Review and Meta-analysis of Randomized Controlled Trials | Adults with musculoskeletal pain conditions, including low back, neck, shoulder, knee, elbow, and ankle pain and whiplash; digital health interventions with at least one month of follow-up | 56 RCTs |
| Wu 2026[8] | Efficacy of Various VR Exposure Therapies for Chronic Low Back Pain: Systematic Review and Network Meta-Analysis. | Adults with chronic low back pain lasting >3 months; any VR game or VR-based therapeutic intervention. | 25 RCTs |
| Gu 2026[9] | Comparative Effectiveness of AI-Assisted Telerehabilitation, Telerehabilitation, In-Person Care, and Usual Care for Chronic Nonspecific Low Back Pain: Bayesian Network Meta-Analysis. | Adults with chronic nonspecific low back pain lasting ≥12 weeks; AI-assisted telerehabilitation, conventional telerehabilitation, in-person rehabilitation, and usual care. | 20 RCTs |

**3.1.** Studies excluded after full-text assessment

1. Nouwen A. EMG biofeedback used to reduce standing levels of paraspinal muscle tension in chronic low back pain. Pain. 1983;17(4):353-360. doi:10.1016/0304-3959(83)90166-5
2. Stuckey SJ, Jacobs A, Goldfarb J. EMG biofeedback training, relaxation training, and placebo for the relief of chronic back pain. Percept Mot Skills. 1986;63(3):1023-1036. doi:10.2466/pms.1986.63.3.1023
3. Donaldson S, Romney D, Donaldson M, Skubick D. Randomized study of the application of single motor unit biofeedback training to chronic low back pain. J Occup Rehabil. 1994;4(1):23-37. doi:10.1007/BF02109994
4. Newton-John TR, Spence SH, Schotte D. Cognitive-behavioural therapy versus EMG biofeedback in the treatment of chronic low back pain. Behav Res Ther. 1995;33(6):691-697. doi:10.1016/0005-7967(95)00008-l
5. Moore JE, Von Korff M, Cherkin D, Saunders K, Lorig K. A randomized trial of a cognitive-behavioral program for enhancing back pain self care in a primary care setting. Pain. 2000;88(2):145-153. doi:10.1016/S0304-3959(00)00314-6
6. Ström L, Pettersson R, Andersson G. A controlled trial of self-help treatment of recurrent headache conducted via the Internet. J Consult Clin Psychol. 2000;68(4):722-727.
7. Cherkin DC, Eisenberg D, Sherman KJ, et al. Randomized trial comparing traditional Chinese medical acupuncture, therapeutic massage, and self-care education for chronic low back pain. Arch Intern Med. 2001;161(8):1081-1088. doi:10.1001/archinte.161.8.1081
8. Lorig KR, Laurent DD, Deyo RA, Marnell ME, Minor MA, Ritter PL. Can a Back Pain E-mail Discussion Group improve health status and lower health care costs?: A randomized study. Arch Intern Med. 2002;162(7):792-796. doi:10.1001/archinte.162.7.792
9. Taylor S, Ellis I, Gallagher M Patient Satisfaction with a New Physiotherapy Telephone Service for Back Pain Patients Physiotherapy, 88, 645-657
10. Blixen CE, Bramstedt KA, Hammel JP, Tilley BC. A pilot study of health education via a nurse-run telephone self-management programme for elderly people with osteoarthritis. J Telemed Telecare. 2004;10(1):44-49. doi:10.1258/135763304322764194
11. Buhrman M, Fältenhag S, Ström L, Andersson G. Controlled trial of Internet-based treatment with telephone support for chronic back pain. Pain. 2004;111(3):368-377. doi:10.1016/j.pain.2004.07.021
12. Bruce B, Lorig K, Laurent D, Ritter P. The impact of a moderated e-mail discussion group on use of complementary and alternative therapies in subjects with recurrent back pain. Patient Educ Couns. 2005;58(3):305-311. doi:10.1016/j.pec.2004.08.012
13. Devineni T, Blanchard EB. A randomized controlled trial of an internet-based treatment for chronic headache. Behav Res Ther. 2005;43(3):277-292. doi:10.1016/j.brat.2004.01.008
14. Desmoulin, G., Yasin, N., and Chen, D. (2007). Initial Results Using Khan Kinetic Treatment™ as a Low Back Pain Treatment Option. Journal of Musculoskeletal Pain - J MUSCULOSKELET PAIN 15, 91–102. 10.1300/J094v15n03_12.
15. Oleske DM, Lavender SA, Andersson GB, Kwasny MM. Are back supports plus education more effective than education alone in promoting recovery from low back pain?: Results from a

randomized clinical trial. Spine (Phila Pa 1976). 2007;32(19):2050-2057. doi:10.1097/BRS.0b013e3181453fcc
16. Brattberg G. Self-administered EFT (Emotional Freedom Techniques) in Individuals With Fibromyalgia: A Randomized Trial. Integr Med (IMCJ). 2008;7(4):30-35
17. Hartwich-Tersek J, Glombiewski JA, Rief W. Positive Effekte von Informationsvermittlung bei Patienten mit chronischen Rückenschmerzen. Verhaltenstherapie. 2008;18(3):153-161. doi:10.1159/000151379
18. Lorig KR, Ritter PL, Laurent DD, Plant K. The internet-based arthritis self-management program: a one-year randomized trial for patients with arthritis or fibromyalgia. Arthritis Rheum. 2008;59(7):1009-1017. doi:10.1002/art.23817
19. Naylor MR, Keefe FJ, Brigidi B, Naud S, Helzer JE. Therapeutic Interactive Voice Response for chronic pain reduction and relapse prevention. Pain. 2008;134(3):335-345. doi:10.1016/j.pain.2007.11.001
20. Berman RLH, Iris MA, Bode R, Drengenberg C. The effectiveness of an online mind-body intervention for older adults with chronic pain. J Pain. 2009;10(1):68-79. doi:10.1016/j.jpain.2008.07.006
21. Glombiewski JA, Hartwich-Tersek J, Rief W. Two psychological interventions are effective in severely disabled, chronic back pain patients: a randomised controlled trial. Int J Behav Med. 2010;17(2):97-107. doi:10.1007/s12529-009-9070-4
22. Leveille SG, Huang A, Tsai SB, Allen M, Weingart SN, Iezzoni LI. Health coaching via an internet portal for primary care patients with chronic conditions: a randomized controlled trial. Med Care. 2009;47(1):41-47. doi:10.1097/MLR.0b013e3181844dd0
23. Vermani, E., Mittal, R. and Weeks, A. (2010), Pelvic Girdle Pain and Low Back Pain in Pregnancy: A Review. Pain Practice,10:60-71.DOI: 10.1111/j.1533-2500.2009.00267.x
24. Cherkin DC, Sherman KJ, Kahn J, et al. A comparison of the effects of 2 types of massage and usual care on chronic low back pain: a randomized, controlled trial. Ann Intern Med. 2010;153(9):570-579. doi:10.7326/0003-4819-153-9-201011020-00006
25. Ang DC, Chakr R, Mazzuca S, France CR, Steiner J, Stump T. Cognitive-behavioral therapy attenuates nociceptive responding in patients with fibromyalgia: a pilot study. Arthritis Care Res (Hoboken). 2010;62(5):618-623. doi:10.1002/acr.20119
26. Chiauzzi E, Pujol LA, Wood M, et al. painACTION-back pain: a self-management website for people with chronic back pain. Pain Med. 2010;11(7):1044-1058. doi:10.1111/j.1526-4637.2010.00879.x
27. Hørdam B, Sabroe S, Pedersen PU, Mejdahl S, Søballe K. Nursing intervention by telephone interviews of patients aged over 65 years after total hip replacement improves health status: a randomised clinical trial. Scand J Caring Sci. 2010;24(1):94-100. doi:10.1111/j.1471-6712.2009.00691.x
28. Johnston M, Foster M, Shennan J, Starkey NJ, Johnson A. The effectiveness of an Acceptance and Commitment Therapy self-help intervention for chronic pain. Clin J Pain. 2010;26(5):393-402. doi:10.1097/AJP.0b013e3181cf59ce
29. Kosterink SM, Huis in 't Veld RM, Cagnie B, Hasenbring M, Vollenbroek-Hutten MM. The clinical effectiveness of a myofeedback-based teletreatment service in patients with non-

specific neck and shoulder pain: a randomized controlled trial. J Telemed Telecare. 2010;16(6):316-321. doi:10.1258/jtt.2010.006005

42. Bromberg J, Wood ME, Black RA, Surette DA, Zacharoff KL, Chiauzzi EJ. A randomized trial of a web-based intervention to improve migraine self-management and coping. Headache.

2012;52(2):244-261. doi:10.1111/j.1526-4610.2011.02031.x
43. Cerrada CJ, Weinberg J, Sherman KJ, Saper RB. Inter-method reliability of paper surveys and computer assisted telephone interviews in a randomized controlled trial of yoga for low back pain. BMC Res Notes. 2014;7:227. Published 2014 Apr 9. doi:10.1186/1756-0500-7-227
44. Cerrada C, Weinberg J, Dresner D, et al. Comparison of paper surveys and computer-assisted telephone interviews in a randomized controlled trial of yoga for low back pain [conference abstract]. BMC Complement Altern Med. 2012;12(Suppl 1):P370. doi:10.1186/1472-6882-12-S1-P370.
45. McBeth J, Prescott G, Scotland G, et al. Cognitive Behavior Therapy, Exercise, or Both for Treating Chronic Widespread Pain. Archives of Internal Medicine. 2012;172(1):48. doi:10.1001/archinternmed.2011.555
46. Moessner M, Schiltenwolf M, Neubauer E. Internet-based aftercare for patients with back pain-a pilot study. Telemed J E Health. 2012;18(6):413-419. doi:10.1089/tmj.2011.0221
47. Ruehlman LS, Karoly P, Enders C. A randomized controlled evaluation of an online chronic pain self management program. Pain. 2012;153(2):319-330. doi:10.1016/j.pain.2011.10.025
48. Simon D, Kriston L, von Wolff A, et al. Effectiveness of a web-based, individually tailored decision aid for depression or acute low back pain: A randomized controlled trial. Patient Education and Counseling. 2012;87(3):360-368. doi:10.1016/j.pec.2011.10.009
49. Slater M, Atkinson J, Weickgenant A, et al. Six-month follow-up of a telehealth intervention for chronic back pain. The Journal of Pain. 2012;13(4):S97. doi:10.1016/j.jpain.2012.01.401
50. Slater M, Chircop-Rollick T, Patel S, et al. Telephone-delivered cognitive behavioral therapy for chronic back pain [conference abstract]. Psychosom Med. 2012;74(3):A58. doi:10.1097/PSY.0b013e3182583b27.
51. Oh HW, Lee MG, Jang JY, et al. Time-effects of horse simulator exercise on psychophysiological responses in men with chronic low back pain. Isokinet Exerc Sci. 2014;22(2):153-163. doi:10.3233/IES-140533.
52. Yoo JH, Kim SE, Lee MG, et al. The effect of horse simulator riding on visual analogue scale, body composition and trunk strength in the patients with chronic low back pain. Int J Clin Pract. 2014;68(8):941-949. doi:10.1111/ijcp.12414.
53. Kim TY, Lee JH, Oh SJ, et al. Effectiveness of simulated horseback riding for patients with chronic low back pain: a randomized controlled trial. J Sport Rehabil. 2020;29(2):179-185. doi:10.1123/jsr.2018-0252.
54. Park S, Park S, Min S, et al. A randomized controlled trial investigating the effects of equine simulator riding on low back pain, morphological changes, and trunk musculature in elderly women. Medicina (Kaunas). 2020;56(11):610. doi:10.3390/medicina56110610.
55. Yasir A, Quddus N, Grover S. The efficacy of motor control exercise on Physical function of spine in chronic low back pain: pre-test post-test experimental group design. Indian J Physiother Occup Ther. 2012;6(4):145-148
56. Shahzad Ali, Syed Murtaza Ali, Khalida Naz Memon. Effectiveness of core stabilization exercises versus Mckenzie's exercises in chronic lower back pain. Med Forum Mon. 2013;24(12):82-85

70. Li, Lun-Lan & Gan, Yu-Yun & Zhang, Li-Na & Wang, Ya-Bing & Zhang, Fan & Qi, Jin-Mei. (2014). The effect of post-discharge telephone intervention on rehabilitation following total hip replacement surgery. International Journal of Nursing Sciences. 1. 10.1016/j.ijnss.2014.05.005.

82. Trost Z, Nowlin L, Guck A, Madi D, Davis M. (491) Exploring the role of pain-related fear and catastrophizing in response to a VR gaming intervention for chronic low back pain. The

Journal of Pain. 2015;16(4):S98. doi:10.1016/j.jpain.2015.01.411
83. Valenzuela-Pascual F, Molina F, Corbi F, Blanco-Blanco J, Gil RM, Soler-Gonzalez J. The influence of a biopsychosocial educational internet-based intervention on pain, dysfunction, quality of life, and pain cognition in chronic low back pain patients in primary care: a mixed methods approach. BMC Med Inform Decis Mak. 2015;15:97. Published 2015 Nov 23. doi:10.1186/s12911-015-0220-0
84. Geraghty AWA, Stanford R, Little P, et al. Using an internet intervention to support self-management of low back pain in primary care: protocol for a randomised controlled feasibility trial (SupportBack).BMJ Open 2015;5:e009524. doi: 10.1136/bmjopen-2015-009524
85. Chen SY, Kim SK, Kim KH, Lee IS, HwangBo G. Effects of Horse Riding Simulator on Pain, Oswestry Physical function Index and Balance in Adults with Nonspecific Chronic Low Back Pain. J Korean Soc Phys Med. 2016;11(4):79-84. doi:10.13066/kspm.2016.11.4.79
86. Müller R, Gertz KJ, Molton IR, et al. Effects of a Tailored Positive Psychology Intervention on Well-Being and Pain in Individuals With Chronic Pain and a Physical Physical function: A Feasibility Trial. Clin J Pain. 2016;32(1):32-44. doi:10.1097/AJP.0000000000000225
87. Thomas JS, France CR, Applegate ME, Leitkam ST, Walkowski S. Feasibility and Safety of a VR Dodgeball Intervention for Chronic Low Back Pain: A Randomized Clinical Trial. J Pain. 2016;17(12):1302-1317. doi:10.1016/j.jpain.2016.08.011
88. Williams AJ, Wiggers J, O'Brien KM, Wolfenden L, Yoong S, Williams CM. A telephone-based lifestyle behavioural intervention for overweight or obese patients with low back pain. Obes Rev. 2016;17(Suppl 2):156. doi:10.1111/obr.12403.
89. Bennell KL, Nelligan R, Dobson F, et al. Effectiveness of an Internet-Delivered Exercise and Pain-Coping Skills Training Intervention for Persons With Chronic Knee Pain: A Randomized Trial. Ann Intern Med. 2017;166(7):453-462. doi:10.7326/M16-1714
90. Bini SA, Mahajan J. Clinical outcomes of remote asynchronous telerehabilitation are equivalent to traditional therapy following total knee arthroplasty: A randomized control study. J Telemed Telecare. 2017;23(2):239-247. doi:10.1177/1357633X16634518
91. Calner T, Nordin C, Eriksson MK, Nyberg L, Gard G, Michaelson P. Effects of a self-guided, web-based activity programme for patients with persistent musculoskeletal pain in primary healthcare: A randomized controlled trial. Eur J Pain. 2017;21(6):1110-1120. doi:10.1002/ejp.1012
92. Dear BF, Gandy M, Karin E, et al. The pain course: a randomised controlled trial comparing a remote-delivered chronic pain management program when provided in online and workbook formats. Pain. 2017;158(7):1289-1301. doi:10.1097/j.pain.0000000000000916
93. Heapy AA, Higgins DM, Goulet JL, et al. Interactive Voice Response-Based Self-management for Chronic Back Pain: The COPES Noninferiority Randomized Trial. JAMA Intern Med. 2017;177(6):765-773. doi:10.1001/jamainternmed.2017.0223
94. Herbert MS, Afari N, Liu L, et al. Telehealth Versus In-Person Acceptance and Commitment Therapy for Chronic Pain: A Randomized Noninferiority Trial. J Pain. 2017;18(2):200-211. doi:10.1016/j.jpain.2016.10.014

doi:10.1016/j.cct.2018.05.001
109. Matheve T, Brumagne S, Demoulin C, Timmermans A. Sensor-based postural feedback is more effective than conventional feedback to improve lumbopelvic movement control in patients with chronic low back pain: a randomised controlled trial. J Neuroeng Rehabil. 2018;15(1):85. Published 2018 Sep 26. doi:10.1186/s12984-018-0423-6
110. Mecklenburg G, Smittenaar P, Erhart-Hledik JC, Perez DA, Hunter S. Effects of a 12-Week Digital Care Program for Chronic Knee Pain on Pain, Mobility, and Surgery Risk: Randomized Controlled Trial. J Med Internet Res. 2018;20(4):e156. Published 2018 Apr 25. doi:10.2196/jmir.9667
111. Rutledge T, Atkinson JH, Chircop-Rollick T, et al. Randomized Controlled Trial of Telephone-delivered Cognitive Behavioral Therapy Versus Supportive Care for Chronic Back Pain. Clin J Pain. 2018;34(4):322-327. doi:10.1097/AJP.0000000000000555
112. Rutledge T, Atkinson JH, Holloway R, et al. Randomized Controlled Trial of Nurse-Delivered Cognitive-Behavioral Therapy Versus Supportive Psychotherapy Telehealth Interventions for Chronic Back Pain. J Pain. 2018;19(9):1033-1039. doi:10.1016/j.jpain.2018.03.017
113. Sobie, T. J. (2016). Body schema acuity training and Feldenkrais movements compared to core stabilization biofeedback and motor control exercises: Comparative effects on chronic non-specific low back pain in an outpatient clinical setting: A randomized controlled comparative efficacy study (Publication No. 10251703) [Doctoral dissertation, Saybrook University]. ProQuest Dissertations & Theses Global.
114. Bishop F, Greville-Harris M, Bostock J, Din A, Informing Adults With Back Pain About Placebo Effects: Randomized Controlled Evaluation of a New Website With Potential to Improve Informed Consent in Clinical Research J Med Internet Res 2019;21(1):e9955. DOI: 10.2196/jmir.9955
115. Coli, T., & Aspell, J. . Investigating the long-term efficacy of an ‘out of body’ illusion in the management of chronic pain [Conference abstract]. Brain and Neuroscience Advances,2019;3, 391. https://doi.org/10.1177/2398212819855490
116. Critchley D, McCracken L, Wileman V, Galea Holmes M, Norton S, Godfrey E. Physiotherapy informed by Acceptance and Commitment Therapy (PACT) for people with chronic low back pain: a randomised controlled trial. Physiotherapy. 2019;105:e34-e35. doi:10.1016/j.physio.2018.11.279
117. Day MA, Ward LC, Ehde DM, et al. A Pilot Randomized Controlled Trial Comparing Mindfulness Meditation, Cognitive Therapy, and Mindfulness-Based Cognitive Therapy for Chronic Low Back Pain. Pain Med. 2019;20(11):2134-2148. doi:10.1093/pm/pny273
118. Gannon, Jamie & Atkinson, Joseph & Chircop-Rollick, Telehealth Therapy Effects of Nurses and Mental Health Professionals From 2 Randomized Controlled Trials for Chronic Back Pain. The Clinical journal of pain. 2019;35. 295-303. 10.1097/AJP.0000000000000678.
119. Jamison RN, Wan L, Edwards RR, Mei A, Ross EL. Outcome of a High-Frequency Transcutaneous Electrical Nerve Stimulator (hfTENS) Device for Low Back Pain: A Randomized Controlled Trial. Pain Pract. 2019;19(5):466-475. doi:10.1111/papr.12764
120. McCurry SM, Von Korff M, Morin CM, et al. Telephone interventions for co-morbid insomnia and osteoarthritis pain: The OsteoArthritis and Therapy for Sleep (OATS) randomized trial design. Contemp Clin Trials. 2019;87:105851. doi:10.1016/j.cct.2019.105851

148. Rhon DI, Mayhew RJ, Greenlee TA, Fritz JM. The influence of a MOBile-based video Instruction for Low back pain (MOBIL) on initial care decisions made by primary care providers: a randomized controlled trial. BMC Fam Pract. 2021;22(1):200. Published 2021 Oct 9. doi:10.1186/s12875-021-01549-y
149. Solar C, Halat AM, MacLean RR, et al. Predictors of engagement in an internet-based cognitive behavioral therapy program for veterans with chronic low back pain. Transl Behav Med. 2021;11(6):1274-1282. doi:10.1093/tbm/ibaa098
150. Adhia DB, Mani R, Turner PR, Vanneste S, De Ridder D. Infraslow Neurofeedback Training Alters Effective Connectivity in Individuals with Chronic Low Back Pain: A Secondary Analysis of a Pilot Randomized Placebo-Controlled Study. Brain Sci. 2022;12(11):1514. Published 2022 Nov 8. doi:10.3390/brainsci12111514
151. Ashar YK, Gordon A, Schubiner H, et al. Effect of Pain Reprocessing Therapy vs Placebo and Usual Care for Patients With Chronic Back Pain: A Randomized Clinical Trial. JAMA Psychiatry. 2022;79(1):13-23. doi:10.1001/jamapsychiatry.2021.2669
152. Blanchard A, Nguyen SM, Devanne M, Simonnet M, Le Goff-Pronost M, Rémy-Néris O. Technical Feasibility of Supervision of Stretching Exercises by a Humanoid Robot Coach for Chronic Low Back Pain: The R-COOL Randomized Trial. Biomed Res Int. 2022;2022:5667223. Published 2022 Mar 9. doi:10.1155/2022/5667223
153. Büyükşireci DE, Demirsoy N, Mit S, Geçioğlu E, Onurlu İ, Günendi Z. Comparison of the Effects of Myofascial Meridian Stretching Exercises and Acupuncture in Patients with Low Back Pain. J Acupunct Meridian Stud. 2022;15(6):347-355. doi:10.51507/j.jams.2022.15.6.347
154. Ho KKN, Simic M, Pinheiro MB, et al. Efficacy of a digital cognitive behavioral therapy for insomnia in people with low back pain: a feasibility randomized co-twin and singleton-controlled trial. Pilot Feasibility Stud. 2022;8(1):125. Published 2022 Jun 14. doi:10.1186/s40814-022-01087-z
155. Sergeenko O, Savin D, Ryabykh S. A101: Spinal deformity and split cord malformations type I: One-stage or two-stage treatment? Global Spine Journal. 2022;12(3):595-608. doi:10.1177/21925682221096074
156. Marugán-Rubio D, Chicharro JL, Becerro-de-Bengoa-Vallejo R, et al. Effectiveness of Ultrasonography Visual Biofeedback of the Diaphragm in Conjunction with Inspiratory Muscle Training on Muscle Thickness, Respiratory Pressures, Pain, Physical function, Quality of Life and Pulmonary Function in Athletes with Non-Specific Low Back Pain: A Randomized Clinical Trial. J Clin Med. 2022;11(15):4318. Published 2022 Jul 25. doi:10.3390/jcm11154318
157. Meng XY, Bu L, Chen JY, et al. Comparative effectiveness of electroacupuncture VS neuromuscular electrical stimulation in the treatment of chronic low back pain in active-duty personals: A single-center, randomized control study. Front Neurol. 2022;13:945210. Published 2022 Sep 13. doi:10.3389/fneur.2022.945210
158. Mueller J, Niederer D, Tenberg S, Oberheim L, Moesner A, Mueller S. Acute effects of game-based biofeedback training on trunk motion in chronic low back pain: a randomized cross-over pilot trial. BMC Sports Sci Med Rehabil. 2022;14(1):192. Published 2022 Nov 13. doi:10.1186/s13102-022-00586-z
159. Paganini S, Terhorst Y, Sander LB, et al. Internet- and mobile-based intervention for depression in adults with chronic back pain: A health economic evaluation. J Affect Disord. 2022;308:607-615. doi:10.1016/j.jad.2022.04.004
160. Papanikolaou F, Petsiti A, Liampas I, Chalkias A, Gouva M, Arnaoutoglou E. Massage-electroacupuncture versus epidural analgesia for patients with chronic low back pain: a randomized

controlled trial [abstract]. Reg Anesth Pain Med. 2022;47(suppl 1). doi:10.1136/rapm-2022-ESRA.452. Presented at: 39th Annual ESRA Congress; June 22-25, 2022

173. O'Hagan ET, Cashin AG, Rizzo RRN, et al. Development of a booster intervention for graded sensorimotor retraining (RESOLVE) in people with persistent low back pain: A nested,

randomised, feasibility trial. Musculoskeletal Care. 2023;21(2):444-452. doi:10.1002/msc.1715
174. Okhotin D, Maddox T, Sparks C, et al. ID: 207542 Durable Chronic Low Back Pain Relief by RelieVRx, an at-Home VR Program, 18 months Post-Treatment . Neuromodulation. 2023;26(4)(suppl):S33. doi:10.1016/j.neurom.2023.04.057
175. Rughani G, Nilsen TIL, Wood K, et al. The selfBACK artificial intelligence-based smartphone app can improve low back pain outcome even in patients with high levels of depression or stress. Eur J Pain. 2023;27(5):568-579. doi:10.1002/ejp.2080
176. Talbot LA, Webb L, Ramirez VJ, et al. Non-pharmacological Home Therapies for Subacute Low Back Pain in Active Duty Military Personnel: A Randomized Controlled Trial. Mil Med. 2023;188(1-2):12-19. doi:10.1093/milmed/usab382
177. Albers R, Lemke S, Fauser D, Knapp S, Krischak G, Bethge M. Non-inferiority of hybrid outpatient telerehabilitation for patients with back pain: 3-month follow-up of a randomized controlled trial. Eur J Phys Rehabil Med. 2024;60(6):1009-1018. doi:10.23736/S1973-9087.24.08458-2
178. Čeko M, Baeuerle T, Webster L, Wager TD, Lumley MA. The effects of VR neuroscience-based therapy on clinical and neuroimaging outcomes in patients with chronic back pain: a randomized clinical trial. Pain. 2024;165(8):1860-1874. doi:10.1097/j.pain.0000000000003198
179. Desgagnés A, Côté-Picard C, Gaumond A, et al. Efficacy of a Psychologically-Informed Physiotherapy Intervention in Patients with Chronic Low Back Pain at High Risk of Poor Prognosis: A Pilot and Feasibility Randomized Controlled Trial. Physiother Can. 2024;76(2):163-174. Published 2024 May 8. doi:10.3138/ptc-2023-0038
180. Fritsch CG, Ferreira ML, Halliday MH, et al. Health coaching intervention with or without the support of an exercise buddy to increase physical activity of people with chronic low back pain compared to usual care: a feasibility and pilot randomised controlled trial. Musculoskelet Sci Pract. 2024;71:102941. doi:10.1016/j.msksp.2024.102941
181. Fritz JM, Ford I, George SZ, et al. Telehealth delivery of physical therapist-led interventions for persons with chronic low back pain in underserved communities: lessons from pragmatic clinical trials. Front Pain Res (Lausanne). 2024;5:1324096. Published 2024 Apr 19. doi:10.3389/fpain.2024.1324096
182. George SZ, France C, Coffman CJ, et al. Cohort Profile: Baseline Characteristics of Veterans from Improving Veteran Access to Integrated Management of Back Pain (AIM-Back) - an Embedded Pragmatic, Cluster Randomized Trial in the United States. Preprint. medRxiv. 2024;2024.11.23.24317833. Published 2024 Nov 26. doi:10.1101/2024.11.23.24317833
183. Hancock M, Smith A, O'Sullivan P, et al. Patients with worse Physical function respond best to cognitive functional therapy for chronic low back pain: a pre-planned secondary analysis of a randomised trial. J Physiother. 2024;70(4):294-301. doi:10.1016/j.jphys.2024.08.005
184. Harvie DS, Kelly J, Kluver J, Deen M, Spitzer E, Coppieters MW. A randomized controlled pilot study examining immediate effects of embodying a VR superhero in people with chronic low back pain. Disabil Rehabil Assist Technol. 2024;19(3):851-858. doi:10.1080/17483107.2022.2129846
185. Knoop J, Slatman S, Staal B. Response to In-Home VR Program for Chronic Lower Back Pain: A Randomized Sham-Controlled Effectiveness Trial in a Clinically Severe and Diverse Sample. Mayo Clin Proc Digit Health. 2023;2(1):38-40. Published 2023 Dec 26. doi:10.1016/j.mcpdig.2023.11.010
186. López-Marcos JJ, Díaz-Arribas MJ, Valera-Calero JA, et al. The Added Value of Face-to-Face Supervision to a Therapeutic Exercise-Based App in the Management of Patients with

Chronic Low Back Pain: A Randomized Clinical Trial. Sensors (Basel). 2024;24(2):567. Published 2024 Jan 16. doi:10.3390/s24020567
187. O'Hagan ET, Traeger AC, Schabrun SM, et al. It's OK to Move! Effect of a Brief Video on Community Confidence in Activity Despite Back Pain: A Randomized Trial. J Orthop Sports Phys Ther. 2024;54(6):400-407. doi:10.2519/jospt.2024.12412
188. Özden F, Güçlü B, Tümtürk İ, Doğrukök ÖN, İmerci A, Tuğay BU. The effect of visual feedback-based clinical monitoring application in patients with chronic low back pain: a randomized controlled trial. Eur Spine J. 2024;33(2):505-516. doi:10.1007/s00586-023-08098-1
189. Poolman EY, Vorstermans L, Donker MH, et al. How people with persistent pain experience in-person physiotherapy blended with biopsychosocial digital health - A qualitative study on participants' experiences with Back2Action. Internet Interv. 2024;36:100731. Published 2024 Feb 28. doi:10.1016/j.invent.2024.100731
190. Rasmussen CDN, Sandal LF, Holtermann A, Stochkendahl MJ, Mork PJ, Søgaard K. Effect of a smartphone self-management digital support system for low-back pain (selfBACK) among workers with high physical work demands - secondary analysis of a randomized controlled trial. Scand J Work Environ Health. 2024;50(8):613-621. doi:10.5271/sjweh.4186
191. Sobie T. RCT Study: Augmented Reality and Feldenkrais® vs. Traditional Core Stabilization: Comparative Effects on Chronic LBP [abstract]. Arch Phys Med Rehabil. 2024;105(4):e162-e163. doi:10.1016/j.apmr.2024.02.573
192. Son S, Yoo BR, Jeong YM. Digital therapeutics-based lumbar core exercise for patients with low back pain: A prospective exploratory pilot study. Digit Health. 2024;10:20552076231218154. Published 2024 Jan 9. doi:10.1177/20552076231218154
193. Yalfani A, Abedi M, Raeisi Z, Asgarpour A. The effects of VR training on postural sway and physical function performance on older women with chronic low back pain: A double-blind randomized clinical trial. J Back Musculoskelet Rehabil. 2024;37(3):761-770. doi:10.3233/BMR-230260
194. Abdala DW, Castro T, Costa WDSD, et al. Effect of thigh muscle strength training through telerehabilitation in volunteers with low back pain. A controlled and randomized clinical trial. J Bodyw Mov Ther. 2025;43:28-34. doi:10.1016/j.jbmt.2025.04.016
195. Ashar YK, Low EL, Knight K, et al. Pain Reprocessing Therapy vs Placebo and Usual Care for Patients With Chronic Back Pain: 5-Year Follow-Up of a Randomized Clinical Trial. JAMA Psychiatry. 2025;82(10):1049-1051. doi:10.1001/jamapsychiatry.2025.1844
196. Dosbaba F, Senkyr V, Vlazna D, et al. Comparison of hybrid guided home-based and outpatient rehabilitation in patients with chronic low back pain: A randomized controlled trial. J Bodyw Mov Ther. 2025;45:110-118. doi:10.1016/j.jbmt.2025.08.009
197. Espin A, Irazusta J, Aiestaran M, et al. Videoconference-Supervised Group Exercise Reduces Low Back Pain in Eldercare Workers: Results from the ReViEEW Randomised Controlled Trial. J Occup Rehabil. 2025;35(1):66-77. doi:10.1007/s10926-024-10182-2
198. Espin A, Irazusta J, Urquiza M, et al. Videoconference-Supervised Exercise for Low Back Pain in Eldercare Workers: One-Year Follow-up of the ReViEEW Randomized Controlled Trial. J Occup Environ Med. 2025;67(10):e720-e728. doi:10.1097/JOM.0000000000003474
199. Fourré A, Michielsen J, Ris L, et al. Comparing the impact of interactive versus traditional e-learning on physiotherapists' knowledge, attitudes, and clinical decision-making in low back

pain management: a randomized controlled trial. J Man Manip Ther. 2025;33(6):505-518. doi:10.1080/10669817.2025.2476670
200. Geraghty AWA, Hughes S, Roberts L, et al. Understanding internet-supported self-management for low back pain in primary care: a qualitative process evaluation of the SupportBack 2 randomised controlled trial. BMJ Open. 2025;15(10):e103428. Published 2025 Oct 20. doi:10.1136/bmjopen-2025-103428
201. Hüsing P, Busmann M, Löwe B, Engelmann P. Effectiveness and acceptance of a personalized online intervention to enhance pain-related self-efficacy in back pain: A Two-Arm Randomized Controlled Trial (IDRIS). J Psychosom Res. 2025;196:112246. doi:10.1016/j.jpsychores.2025.112246
202. Hüsing P, Busmann M, Löwe B, Engelmann P. A personalized online intervention to enhance back pain-related self-efficacy: A two-arm randomized controlled trial (IDRIS). Internet Interv. 2025;42:100892. Published 2025 Nov 25. doi:10.1016/j.invent.2025.100892
203. Jones Berkeley S, Wedin S, Patidar SM, et al. Design and implementation of online acceptance and commitment therapy with enhanced therapist support for chronic low back pain (ACT for PAIN). Pain Med. 2025;26(8):451-458. doi:10.1093/pm/pnaf026
204. Kaczorowski S, Trompeter K, Kramer C, et al. Usability and Acceptability of an App-Based Approach to Treat Low Back Pain: Preplanned Secondary Analysis of a Randomized Controlled Trial. JMIR Form Res. 2025;9:e59866. Published 2025 Aug 25. doi:10.2196/59866
205. Kałużna A, Hagner-Derengowska M, Kałużny K, Pyskir M, Grigorief M, Budzyński J. Kinesiotherapy supported by visual biofeedback on a stabilometric platform is more effective than kinesiotherapy alone for patients with non-specific low back pain. A randomized, open-label study with a 6-month follow-up. Med Res J. 2025;10(2):145-153. doi:10.5603/mrj.106722.
206. Li YH, Li N, Liu ZX, et al. The effectiveness of artificial intelligence health education accurately linking system on self-management in non-specific lower back pain patients. Front Public Health. 2025;13:1630329. Published 2025 Sep 11. doi:10.3389/fpubh.2025.1630329
207. Lindner N, Kornder N, Heisig J, Becker A, der Wardt VV, Viniol A. Digitally supported physical activity counselling for people with chronic back pain: a randomised controlled parallel feasibility study. BMC Prim Care. 2025;26(1):58. Published 2025 Feb 27. doi:10.1186/s12875-025-02742-z
208. Abstracts from the PAINWeek 2025 Annual Conference. Postgrad Med. 2025;137(suppl 2):2-117. doi:10.1080/00325481.2025.2597706
209. Marins EF, Primo TT, Vasconcelos BB, et al. Effectiveness of an app-delivered, self-management exercise program in public safety workers with chronic low back pain: a randomized controlled trial. Braz J Phys Ther. 2025;29(5):101232. doi:10.1016/j.bjpt.2025.101232
210. Moshe I, Haslbeck J, Terhorst Y, et al. Network intervention analysis of a digital intervention for the prevention of depression in patients with chronic back pain. Behav Res Ther. 2025;192:104784. doi:10.1016/j.brat.2025.104784
211. Moudjari I, Pautard C, Jouanneau C, Servel AG, Le Cavorzin P, Margo B, et al. Evaluation of the Performance of Blueback Physio® Medical Device in the Management of Patients Suffering from Chronic Low Back Pain: Randomized Clinical Trial. IRBM. 2025;46(1):100875. doi:10.1016/j.irbm.2024.100875
212. Neason C, Samanna CL, Tagliaferri SD, et al. Fears and beliefs about running in adults with chronic low back pain: a mixed methods study from the ASTEROID randomised controlled trial. Sports Med Open. 2025;11(1):66. Published 2025 Jun 8. doi:10.1186/s40798-025-00861-6

213. Perotti L, Stamm O, Strohm H, et al. Learning Transversus Abdominis Activation in Older Adults with Chronic Low Back Pain Using an Ultrasound-Based Wearable: A Randomized Controlled Pilot Study. J Funct Morphol Kinesiol. 2025;10(1):14. Published 2025 Jan 1. doi:10.3390/jfmk10010014
214. Saki F, Ziya M. Effects of integrating Feldenkrais method with dynamic neuromuscular stabilization exercises on clinical outcomes in older women with nonspecific chronic low back pain: A randomized controlled trial. BMC Geriatr. 2025;25(1):573. Published 2025 Jul 31. doi:10.1186/s12877-025-06219-7
215. Sarafadeen R, Ganiyu SO, Ibrahim AA, et al. Lumbar stabilization exercise with and without real-time ultrasound imaging biofeedback in chronic low back pain patients: a randomized controlled trial. Sci Rep. 2025;15(1):36975. Published 2025 Oct 22. doi:10.1038/s41598-025-20942-6
216. Schlett C, van der Keylen P, Schöpf-Lazzarino AC, et al. The Effectiveness of a Physician-Led Web Portal on Back Pain: A Cluster Randomized Controlled Trial. Dtsch Arztebl Int. 2025;122(8):203-209. doi:10.3238/arztebl.m2025.0015
217. Zheng X, Zhang M, Huang Y, Shan S. Improvement of lower limb walking function in patients with chronic non-specific low back pain by biofeedback assisted electrical stimulation. Chin J Tissue Eng Res. 2025;29(3):547-553. doi:10.12307/2024.690
218. Xiao C, Zhao Y, Li G, et al. Clinical Efficacy of Multimodal Exercise Telerehabilitation Based on AI for Chronic Nonspecific Low Back Pain: Randomized Controlled Trial. JMIR Mhealth Uhealth. 2025;13:e56176. Published 2025 May 22. doi:10.2196/56176
219. Yalfani A, Asgarpoor A. Comparison of cognitive functional therapy and neurofeedback training on kinetic gait in patients with chronic non-specific low back pain: a randomised controlled trial. Disabil Rehabil. 2025;47(17):4443-4452. doi:10.1080/09638288.2025.2451219
220. Espin A, Rodriguez-Larrad A, Ruiz-Fernández A, et al. Predictors of response to physical exercise for low back pain: a secondary analysis of the ReViEEW trial. Musculoskelet Sci Pract. 2026;81:103465. doi:10.1016/j.msksp.2025.103465
221. Lukkahatai N, Benjasirisan C, Huang X, et al. Smartphone-Based Ecological Momentary Assessment of Pain in Older Adults Undergoing Auricular Point Acupressure for Chronic Low Back Pain: Secondary Analysis of a Randomized Controlled Trial. JMIR Form Res. 2026;10:e79612. Published 2026 Mar 4. doi:10.2196/79612
222. van der Veen SM, Stamenkovic A, France CR, et al. The Effects of Virtual Immersive Gaming to Optimize Recovery (VIGOR) in Low Back Pain: A Phase II Randomized Controlled Trial. Healthcare (Basel). 2026;14(2):142. Published 2026 Jan 6. doi:10.3390/healthcare14020142
223. Wang IL, Lu FF, Chiu CH, Su Y, Tao H, Lai CY. Impact of Nintendo Ring Fit exergaming on lower-limb asymmetry and joint angles in women with chronic nonspecific low back pain during obstacle crossing. BMC Sports Sci Med Rehabil. 2026;18(1):117. Published 2026 Feb 6. doi:10.1186/s13102-025-01516-5
224. Tuck N, Pollard C, Good C, et al. Active Virtual Reality for Chronic Primary Pain: Mixed Methods Randomized Pilot Study. JMIR Form Res. 2022;6(7):e38366. Published 2022 Jul 13. doi:10.2196/38366
225. Iles R, A randomized controlled trial of telephone coaching for return to usual activity in low back pain. Spine Journal 2011;Issue 10 Pages 62S. doi: 10.1016/j.spinee.2011.08.158.
226. Kent P, Laird R, Haines T. The effect of changing movement and posture using motion-sensor biofeedback, versus guidelines-based care, on the clinical outcomes of people with sub-acute

or chronic low back pain-a multicentre, cluster-randomised, placebo-controlled, pilot trial. BMC Musculoskelet Disord. 2015;16:131. Published 2015 May 29. doi:10.1186/s12891-015-0591-5

227. Fatoye F, Gebrye T, Fatoye C, et al. The Clinical and Cost-Effectiveness of Telerehabilitation for People With Nonspecific Chronic Low Back Pain: Randomized Controlled Trial. JMIR Mhealth Uhealth. 2020;8(6):e15375. Published 2020 Jun 24. doi:10.2196/15375

228. Villatoro-Luque FJ, Rodríguez-Almagro D, Aibar-Almazán A, et al. In non-specific low back pain, is an exercise program carried out through telerehabilitation as effective as one carried out in a physiotherapy center? A controlled randomized trial. Musculoskelet Sci Pract. 2023;65:102765. doi:10.1016/j.msksp.2023.102765

229. Feldwieser F, Kiselev J, Hardy S, Garcia-Agundez A, Eicher C, Steinhagen-Thiessen E, Göbel S. Evaluation of biofeedback based bridging exercises on older adults with low back pain: a randomized controlled trial. Physiother Pract Res. 2018;39(1):15–25. doi:10.3233/PPR-170109

**Section 4.** Classification of intervention nodes

We assigned each eligible intervention and comparator arm to one of eight treatment nodes, primarily according to its principal mode of delivery. For multicomponent interventions, therapeutic content, clinician involvement, and treatment dose were considered only to resolve ambiguous boundary cases and were not used as independent node-defining criteria. The eight nodes comprised control, face-to-face care, telemedicine, web applications (Web-App), mobile applications (Mobile-App), biofeedback devices, virtual reality (VR), and exergaming. Detailed operational definitions and study-level assignments are provided in Supplementary Tables 4.1 and 4.2, respectively.

**Table S3.** Treatment nodes included in the network meta-analysis

| Node | Specific definition |
|---|---|
| Control | Comparator condition without a study-specific active therapeutic intervention beyond background usual care, waiting-list management, no intervention, minimal advice or education, or sham/attention control. |
| Face-to-face care | Active treatment whose principal therapeutic dose is delivered through real-time, in-person contact between patient and clinician at the same location. |
| Telemedicine | Active clinical care whose principal therapeutic dose is delivered through scheduled, synchronous, real-time remote encounters, such as telephone calls or videoconferencing. |
| Web-App | Asynchronous digital intervention delivered primarily through a browser-based website, portal, or computer-accessed online platform. |
| Mobile-App | Asynchronous digital intervention delivered primarily through a native smartphone or tablet application, or through a mobile mini-program. |
| Biofeedback devices | Intervention in which a real-time physiological or movement-feedback loop generated by a device is the core active therapeutic mechanism. |
| VR | Intervention in which the principal treatment is delivered within an immersive virtual environment, such that the virtual environment itself is the therapeutic container. |
| Exergaming | Digitally mediated exercise intervention in which gameplay mechanics are inseparably coupled to bodily movement and constitute the main driver of treatment. |

**Note:** Interventions were classified according to their principal mode of delivery. Multicomponent interventions were assigned to the node representing the principal mode through which treatment was delivered. Control included usual care, waiting list, no intervention, minimal advice or education, and sham or attention control. VR=virtual reality; Web-App=web-based application; Mobile-App=mobile applications.

**Table S4.** Treatment node assignments and rationale

| Study ID | Description | Treatment node assignment |
|---|---|---|
| Iles 2011 | Usual care delivered according to routine clinical practice, with modality selection, frequency, and duration determined at the discretion of the treating therapist. | Control |
| | Telephone-delivered health coaching provided by a physiotherapist with three years of clinical experience, applied once per week for four weeks plus one additional session three weeks later, using individualized motivational interviewing and cognitive-behavioural strategies, in addition to usual physiotherapy care including exercise therapy and manual techniques. | Telemedicine |
| Carpenter 2012 | Waitlist control without access to the web-based intervention during the initial 3-week period while continuing usual activities and care | Control |
| | Internet-based CBT self-help intervention provided via an automated web platform, delivered over 3 weeks with approximately two chapters per week, with minimal supervision supported by email reminders, in addition to participants' ongoing usual treatments including medication and prior care. | Web-App |
| Del Pozo-Cruz 2012 | Usual care including standard preventive medicine services with annual clinical visits and access to general self-care information, without access to the structured web-based exercise and postural education programme | Control |
| | Internet-based web program for occupational postural and physical exercise delivered via daily online videos, provided by a clinical exercise physiologist and preventive medicine clinician, performed Monday through Friday for nine months, including postural reminders and exercises targeting strength, flexibility, mobility, and stretching of abdominal, lumbar, hip, and thigh muscles, in addition to standard preventive care with patient visits and self-care web information.Because we aimed to mimic real-life implementation as much as possible, only 1 e-mail, which always contained the same information | Web-App |
| Krein 2013 | Enhanced usual care pedometer program with monthly email reminders to upload data, without individualized goals, feedback, or access to educational content or e-community, in addition to standard monitoring of adverse events. | Control |
| | Internet-based walking program delivered via website and pedometer, provided with automated goal setting, individualized weekly step count targets, motivational messages, educational materials, and moderated e-community, with weekly email reminders to upload data. | Web-App |
| McDonough 2013 | Face-to-face education/advice program provided by a physiotherapist in a single one-hour session, including brief physical examination and standardized guidance to remain active using "The Back Book," without pedometer use or ongoing telephone feedback. | Face-to-face care |
| | Telephone-delivered pedometer-driven walking program plus education provided by a physiotherapist, initially including a one-hour face-to-face session with physical examination and education, followed by weekly individualized step goals and feedback via phone for six weeks, tailored using participant step diaries and self-efficacy walks, in addition to standard education/advice to remain active using "The Back Book." | Telemedicine |

| | | |
|---|---|---|
| Park 2013 | Physical agent modalities consisting of a hot pack for 30 minutes, interferential current therapy for 15 minutes, and deep heat with ultrasound for 5 minutes, without an additional exercise programme. | Control |
| | Physiotherapist-prescribed lumbar stabilization exercise comprising seven bridge, quadruped, and side-bridge positions, performed for 15 seconds for three sets, three times weekly for eight weeks, in addition to the physical modalities provided to all groups. | Face-to-face care |
| | Nintendo Wii Sports exercise using wakeboard, Frisbee dog, jet ski, and canoe games, in which participants controlled virtual characters through swinging, rowing, and tilting motion-sensitive controllers, three times weekly for eight weeks, in addition to common physical modalities. | Exergaming |
| Kim 2014 | Conventional trunk-stabilization exercise delivered for 30 minutes per session, three times per week for 4 weeks, including exercises targeting the transversus abdominis, multifidus, and other trunk muscles, together with conventional physical therapy. | Face-to-face care |
| | Wii Fit yoga–based exergaming delivered for 30 minutes per session, three times per week for 4 weeks, using a Wii Balance Board and on-screen visual feedback during seven yoga-based therapeutic exercises. | Exergaming |
| Irvine 2015 | Usual care consisting of email contacts only to complete study assessments, without access to FitBack or educational websites. | Control |
| | Internet-based education program delivered via eight emails containing links to six websites with information about low back pain, without interactive self-monitoring or tailored behavioral interventions. | |
| | Internet-based self-tailored cognitive-behavioral NLBP management program delivered via FitBack app and website, provided with expert-developed content in consultation with orthopedic, physical therapy, and pain psychology professionals, including weekly emails, interactive self-monitoring of pain and activity, gain-framed messages, journaling, and tracking of daily self-management activities over eight weeks | Mobile-App |
| Geraghty 2018 | Usual care for low back pain continued over the trial period, unrestricted and varying across patients, including no further care beyond the initial consultation or access to treatments such as physiotherapy or pain clinics. | Control |
| | Internet-based tailored self-management program for low back pain delivered over six weekly sessions with automated weekly email reminders, using goal setting, self-monitoring, and tailored feedback for physical activity, in addition to unrestricted usual care. | Web-App |
| | Internet-based tailored self-management program with physiotherapist telephone support delivered over six weekly sessions plus three calls totaling up to one hour by senior musculoskeletal physiotherapists, with individualized encouragement and reassurance, in addition to unrestricted usual care. | |
| Tan 2015 | Face-to-face self-hypnosis training delivered by a therapist in eight weekly sessions, with individualized hypnotic suggestions selected after the first two sessions, without home-practice recommendations. | Face-to-face care |
| | Face-to-face self-hypnosis training delivered by a therapist in eight weekly sessions, with individualized hypnotic suggestions plus audio recordings and recommendations for self-hypnosis home practice. | |

| | | |
|---|---|---|
| | Face-to-face plus telephone-delivered self-hypnosis training delivered by a therapist in two face-to-face sessions plus six brief weekly calls, with individualized hypnotic suggestions, audio recordings, and recommended home practice | Telemedicine |
| | Face-to-face sEMG biofeedback-assisted relaxation training delivered in eight sessions, with forehead electrodes and visual and auditory feedback to reduce muscle tension. | Biofeedback devices |
| Schaller 2016 | Face-to-face educational intervention delivering two general physical activity presentations during rehabilitation with additional downloadable materials available online without interactive or tailored support | Face-to-face care |
| | The intervention (Movement Coaching) was designed as a multicomponent approach and comprised of three different components: face-to-face contact (small group intervention, twice during inpatient rehabilitation), a tailored telephone aftercare (8 weeks and 12 weeks after rehabilitation) and an Internet-based aftercare (web 2.0 platform; available up to six months after rehabilitation) | Web-App |
| Yilmaz Yelvar 2017 | Face-to-face traditional physiotherapy delivered by physiotherapists five times per week for 2 weeks, including physical modalities and supervised therapeutic exercises with additional home exercise practice | Face-to-face care |
| | Virtual reality–based physiotherapy delivering virtual walking–integrated rehabilitation by physiotherapists, applied five times per week for 2 weeks, with supervised exercise sessions plus passive viewing of virtual walking videos during treatment sessions | VR |
| Chhabra 2018 | Face-to-face physician-prescribed physical activity, home exercise, and medication program delivered as a written prescription, including recommended activity level and prescribed medicines with dosages. | Face-to-face care |
| | App-based physical activity and home exercise program delivered via Snapcare, with individualized daily goals and automated monitoring, reminders, and reinforcement adjusted to health status, activities of daily living, and progress, in addition to a written physician prescription and usual medicines. | Mobile-App |
| Goode 2018 | Wait-list control maintaining usual lifestyle without intervention during the 12-week period | Control |
| | Telemedicine-delivered home-based physical activity program supported by structured telephone follow-up from physical therapist and exercise counselor over 12 weeks | Telemedicine |
| | Telemedicine-delivered home-based physical activity combined with cognitive behavioral therapy for pain via structured telephone sessions over 12 weeks | |
| Williams 2018 | Usual care consisting of remaining on the orthopaedic waiting list with unrestricted access to healthcare services without structured lifestyle intervention | Control |
| | Telephone-delivered healthy lifestyle coaching provided by qualified health professionals over 10 individually tailored calls across 6 months, following brief telephone advice and a face-to-face physiotherapist consultation using behavior change techniques, in addition to usual care | Telemedicine |
| Amorim 2019 | Usual care physical activity advice provided by a study investigator with a booklet and brief guidance to stay active, including one post-randomization telephone contact encouraging independent goal progression | Control |

| | | |
|---|---|---|
| | Face-to-face plus telephone-delivered health coaching delivering a tailored physical activity intervention by trained coaches over 6 months with one initial session and 12 fortnightly calls, with individualized goal setting plus support from a mobile web applications and activity tracker and provision of an information booklet | Telemedicine |
| Mbada 2019 | Face-to-face McKenzie therapy delivered in the clinic using repeated extension exercises, plus back care education instructions for home. | Face-to-face care |
| | App-based McKenzie therapy delivered at home via a mobile phone application, using personalized guided self-therapy with tele-monitoring by phone calls and SMS, plus back care education | Mobile-App |
| Shebib 2019 | Control group receiving three digital education articles only with continued access to treatment-as-usual without structured digital care program | Control |
| | App-based digital care program for low back pain delivered remotely over 12 weeks, with a personal coach and peer support team, requiring three weekly sensor-guided exercise sessions, one to two education articles, symptom logging, intermittent CBT, and three aerobic activities, in addition to treatment as usual. | Mobile-App |
| Petrozzi 2019 | Face-to-face standard physical treatment provided by a registered chiropractor or physiotherapist in up to 12 sessions, with treatment frequency determined by clinical judgment, including manual therapy, advice, education, and exercise. | Face-to-face care |
| | Internet-based CBT self-management program delivered via MoodGYM with weekly telephone adherence support, completed as one module per week in addition to 12 sessions of practitioner-directed physical treatment provided by a chiropractor or physiotherapist, including manual therapy, advice, education, and exercise. | Web-App |
| Toelle 2019 | Face-to-face standard physiotherapy provided by a certified physiotherapist in six individual weekly sessions, with exercises tailored to symptoms and fitness level plus manual therapy, in addition to weekly emailed educational website links and motivating messages. | Face-to-face care |
| | App-based multidisciplinary self-management program delivered via the Kaia app, used at least four times weekly for three months, with individually tailored education, physiotherapy exercises, and mindfulness or relaxation content adapted to user progress. | Mobile-App |
| Yang 2019 | Face-to-face physiotherapy provided as prescribed by the physiotherapist, including manual therapy, electrophysical therapy, or traction, without the app-supported self-management program. | Face-to-face care |
| | App-based self-management exercise program delivered with physiotherapist-prescribed individualized exercises, performed four times daily for four weeks with exercise and pain-diary reminders via the Pain Care app, in addition to physiotherapy including manual therapy, electrophysical therapy, or traction. | Mobile-App |
| Zadro 2019 | Control group instructed to maintain usual activities and care-seeking behaviors without structured exercise intervention | Control |
| | Home-based video-game exercise delivered using Nintendo Wii Fit U, introduced by a physical therapist during one initial 1–2 hour home visit, performed unsupervised for 60 minutes three times weekly over 8 weeks, with fortnightly telephone follow-up for progression and adverse-event monitoring. | Exergaming |
| Almhdawi 2020 | App-based placebo intervention delivering general nutrition information via a smartphone application over 6 weeks, with automated daily notifications unrelated to low back pain management and no therapeutic exercise content | Control |

| | | |
|---|---|---|
| | Full version of the “Relieve my back” smartphone application used for 6 weeks, providing evidence-based low back pain self-management advice, office-based stretching exercises, and home-based strengthening exercises for the lower back and abdominal muscles, together with four daily notifications reminding participants to take walking breaks, maintain correct posture, perform stretching exercises, and complete home exercises; participants were allowed to continue their usual medical care. | Mobile-App |
| Matheve 2020 | Face-to-face exercise intervention delivering pelvic tilt training guided by auditory cues in a supervised setting, performed in two 2-minute sessions with standardized tempo and no feedback | Face-to-face care |
| | Virtual reality–based exercise intervention delivering pelvic tilt training via interactive games controlled by motion sensors, performed in a supervised setting in two 2-minute sessions with standardized instructions and no real-time feedback. | VR |
| Nambi 2020 | Conventional non-VR trunk strengthening using isokinetic flexion-extension exercise, performed at 60°/s, 90°/s, and 120°/s for 15 repetitions × 3 sets, 5 days/week for 4 weeks, with warm-up and stretching. | Control |
| | Conventional non-VR core and balance exercise training, including isotonic and isometric exercises for the abdominal and back muscles, performed 10–15 repetitions/day, 5 days/week for 4 weeks, with stretching exercises. | |
| | Face-to-face virtual reality balance training delivered by an experienced physiotherapist, performed in sitting for 30 minutes 5 days per week for 4 weeks with graded difficulty progression, plus prescribed home exercise, hot pack, and ultrasound. | VR |
| Tomruk 2020 | Computer-based postural balance training delivered using the Biodex Balance System under physiotherapist supervision for approximately 30 minutes per session, twice weekly for 12 weeks. Training included postural stability, limits of stability, weight shifting, and maze-control tasks, with immediate visual feedback from the device and progressive platform instability. | Biofeedback devices |
| | Traditional postural control training delivered face-to-face under physiotherapist supervision for the same frequency and duration, including limits-of-stability exercises, static postural stability exercises, weight shifting, and visual, vestibular, and proprioceptive stimulation, without the computerized feedback system. | Face-to-face care |
| Alzahrani 2021 | Face-to-face usual physiotherapy care delivered by physiotherapists over 8 weeks, including strengthening, stabilization, manual therapy, home exercises, and education, without wearable support or structured walking program | Face-to-face care |
| | Wearable device–based walking intervention delivering a progressive step-count program via Fitbit combined with access to a web platform over 8 weeks, added to usual physiotherapy care including exercise, manual therapy, and education, with individualized weekly step targets and self-monitoring feedback | Biofeedback devices |
| Dadarkhah 2021 | Face to face exercise instruction on the pain and disability of people with nonspecific low back pain | Face-to-face care |
| | Home-based core stability exercise program delivered with physical therapist education and a written illustrated guide, performed twice daily for four weeks, with telephone follow-up by a physical therapist three times weekly to address barriers, self-efficacy, and adverse events. | Telemedicine |

| | | |
|---|---|---|
| Garcia 2021 | Sham virtual reality program delivered via VR headset for one daily session over 56 days, using noninteractive 2D nature videos with neutral music, with online setup materials and study staff technical support. | Control |
| | Home-based immersive virtual reality pain self-management program delivered via VR headset for one daily session over 56 days, incorporating CBT, mindfulness, pain education, breathing biofeedback, relaxation, and interactive games, with online setup materials and study staff technical support | VR |
| Kazemi 2021 | Usual care no educational intervention during the study period | Control |
| | Face-to-face educational program delivered in two 60-minute sessions including lectures, role-playing, and group discussions covering ergonomics, spine health, and exercise, with weekly SMS reminders to reinforce adherence | Face-to-face care |
| | Social media–based educational program delivered via website and mobile-access platform providing structured content on ergonomics and exercise, with weekly interactive messaging and reminders to support engagement | Web-App |
| Kim 2021 | Usual care consisting of provision of an educational booklet only without ongoing counseling or structured follow-up during the 8-week study period | Control |
| | Face-to-face plus telephone-delivered individualized educational program providing an educational booklet with personalized counseling through biweekly face-to-face and telephone sessions over 8 weeks to support self-management of low back pain | Telemedicine |
| Lang 2021 | Standardized education and advice on self-management and physical activity delivered prior to allocation without ongoing structured walking program or follow-up support | Control |
| | Face-to-face plus telephone-delivered physiotherapist-guided pedometer-based walking program with individualized weekly step goals over 12 weeks, in addition to standardized education and advice on self-management and staying active | Telemedicine |
| Li 2021 | Face-to-face physical therapy delivered as thermal magnetic therapy for 20 minutes per day over two weeks without additional exercise or behavioral intervention. | Face-to-face care |
| | Face-to-face motor control exercise delivered by a physiotherapist for approximately 30 minutes per day over two weeks with real-time guidance and supervised training, in addition to daily thermal magnetic therapy. | |
| | Face-to-face virtual reality–based exercise delivered using a Kinect system for approximately 30 minutes per day over two weeks, with repeated sessions and instructed movement control, in addition to daily thermal magnetic therapy. | VR |
| Nambi 2021a | Conventional non-VR core muscle training delivered by a trained physiotherapist, performed five days per week for four weeks with active exercises and stretching, together with prescribed home exercise, hot pack, and ultrasound. | Control |
| | Conventional non-VR isokinetic trunk training delivered by a trained physiotherapist, performed five days per week for four weeks with supervised progressive sets and repetitions, together with prescribed home exercise, hot pack, and ultrasound. | |

| | | |
|---|---|---|
| | Virtual reality–based balance training delivered by a trained physiotherapist in a sitting position for 30 minutes, five days per week for four weeks, with graded progression, together with prescribed home exercise, hot pack, and ultrasound. | VR |
| Nambi 2021b | Conventional non-VR balance and core muscle training delivered by an experienced physiotherapist, performed for 10–15 repetitions five days per week for four weeks with stretching, together with prescribed home exercise, hot pack, and ultrasound. | Control |
| | Conventional non-VR Swiss-ball core balance training delivered by an experienced physiotherapist, performed as 15 repetitions for three sets, five days per week for four weeks, with 10-second holds and 3-second rests, together with prescribed home exercise, hot pack, and ultrasound. | |
| | Virtual reality–based balance training delivered by an experienced physiotherapist in a sitting position for 30 minutes, five days per week for four weeks, with graded progression in task difficulty, together with prescribed home exercise, hot pack, and ultrasound. | VR |
| Nambi 2021c | Conventional non-VR active balance exercise for the abdominal and back muscles, performed five times per week for four weeks, together with lower-limb self-stretching, heat therapy, and therapeutic ultrasound. | Control |
| | Conventional non-VR core stability exercise using a therapeutic ball, performed as 15 repetitions for 3 sets with 15-second holds, five times per week for four weeks, in addition to heat therapy and therapeutic ultrasound. | |
| | Virtual reality–based core muscle training using the Pro-Kin system for 30 minutes daily, five times per week for four weeks, with graded progression according to performance, in addition to heat therapy and therapeutic ultrasound. | VR |
| Sandal 2021 | Usual care low back pain management delivered according to the advice or treatment offered by the participant's clinician. | Control |
| | App-based self-management support delivered via the AI-based SELFBACK app with a connected step-detecting wristband, introduced by a research assistant in one face-to-face session, providing weekly individualized recommendations for physical activity, strength and flexibility exercises, and educational messages, in addition to usual care | Mobile-App |
| Sato 2021 | Usual care medication management with continued prior drug therapy and stepwise addition or dose escalation of oral analgesic treatment every 2 weeks, without exergaming or exercise therapy. | Control |
| | Face-to-face exergaming delivered using Ring Fit Adventure for about 40 minutes once weekly for 8 weeks, with game-adjusted exercise intensity based on patient response, in addition to continued previously prescribed medication. | Exergaming |
| Afzal 2022 | Face-to-face routine physiotherapy with moist hot pack heat therapy, hamstring stretching, and back strengthening exercises, provided three sessions weekly on alternate days for 12 sessions. | Face-to-face care |
| | Face-to-face virtual reality exercise delivered using non-immersive Kinect exergames for 10 minutes in addition to routine physiotherapy with heat therapy, hamstring stretching, and back strengthening exercises, provided three sessions weekly on alternate days for 12 sessions. | VR |

| | | |
|---|---|---|
| Eccleston 2022 | No structured intervention or additional instruction with continuation of usual care over 6–8 weeks | Control |
| | Home-based sham virtual reality program delivered over 6 to 8 weeks using the same headset and immersive seashore environment, without therapeutic cognitive-behavioral content and with instructions only to relax and enjoy the environment. | |
| | Home-based immersive virtual reality cognitive-behavioral pain self-management program delivered in 30 modules over 6 to 8 weeks, scheduled for five sessions weekly lasting 15 to 60 minutes, using a virtual mentor, homework, and gamified movement activities. | VR |
| Itoh 2022 | Usual care routine medical care provided at each clinic, without app-based patient education or daily exercise therapy. | Control |
| | App-based patient education and exercise therapy delivered via the Secaide mobile messaging app for 60 days, with daily Artificial Intelligence chatbot messages providing cognitive-behavioral education and simple exercise guidance, in addition to routine medical care. | Mobile-App |
| Koppenaal 2022 | Face-to-face physiotherapy delivered according to low back pain guideline recommendations, with information, exercises, and physical activity advice tailored by physiotherapists, without web-based applications. | Face-to-face care |
| | Blended stratified physiotherapy delivered through a smartphone app in addition to face-to-face physiotherapy, tailored by physiotherapists to individual risk profile, needs, and progress, including self-management information, exercises, and goal-oriented physical activity. | Mobile-App |
| Lara-Palomo 2022 | Conventional home-based rehabilitation combining self-administered lumbar TENS with individually prescribed McKenzie exercises, performed three times weekly for 8 weeks following initial therapist instruction. | Control |
| | Internet-based telerehabilitation delivering individualized McKenzie exercise and transcutaneous electrical nerve stimulation over 8 weeks with three weekly sessions, supported by video-guided instructions and continuous digital platform access | Web-App |
| Meinke 2022 | No intervention with continuation of usual activity without structured exercise or feedback over the study period | Control |
| | Wearable sensor–based exergame delivering trunk movement and postural feedback via inertial measurement units and tablet-guided visual interface, performed as unsupervised home exercises for 20 minutes per session across 9 sessions over 3 weeks with adaptive difficulty and self-directed continuation thereafter | Exergaming |
| Nambi 2022 | Face-to-face conventional lower trunk exercise delivered by a trained physical therapist, for 30 minutes per session, 5 days per week for 4 weeks, with active exercises and lower-limb stretching, plus hydrocollator packs and continuous ultrasound. | Face-to-face care |
| | Face-to-face isokinetic trunk strengthening delivered by a trained physical therapist, for 30 minutes per session, 5 days per week for 4 weeks, with supervised progressive flexion-extension training, in addition to hydrocollator packs and continuous ultrasound. | |
| | Face-to-face virtual reality trunk strengthening delivered by a trained physical therapist, for 30 minutes per session, 5 days per week for 4 weeks, with graded progression in pain-free trunk movements, in addition to hydrocollator packs and continuous ultrasound. | VR |
| Yalfani 2022 | Usual activity without structured exercise or therapeutic intervention during the study period | Control |

| | | |
|---|---|---|
| | Virtual reality–based exercise program delivered via immersive HTC Vive system by supervised sessions, performed three times weekly for 30 minutes over 8 weeks with progressive game-based training involving whole-body movements and real-time audiovisual feedback | VR |
| Fatoye 2022 | Virtual Reality Game (BE-VRG) delivered using a Microsoft Kinect–based interactive virtual game, three sessions per week for 8 weeks, with participants performing standing trunk-extension movements and accompanying left/right side-gliding to interact with virtual balls, with progressively increasing task difficulty and real-time visual and textual feedback, in addition to warm-up/cool-down stretching and back-care education. | Exergaming |
| | Clinic-based McKenzie therapy (CBMT) consisting of supervised McKenzie extension-in-standing exercises, three sessions per week for 8 weeks, with participants repeatedly extending the trunk backwards in standing while keeping the knees straight, up to 10 repetitions per exercise bout, in addition to warm-up/cool-down stretching and back-care education. | Face-to-face care |
| Cui 2023 | Face-to-face physiotherapy delivering exercise, education, manual therapy, and physical modalities by physiotherapists over 8 weeks, with two weekly sessions, individualized adjustment, and additional home exercise practice | Face-to-face care |
| | App-based telerehabilitation delivering exercise, education, and cognitive behavioral therapy via a digital platform supervised by physiotherapists over 8 weeks, with three weekly sessions, individualized programs, real-time biofeedback, asynchronous monitoring, and additional communication through app-based chat or calls | Mobile-App |
| Garreta-Catala 2023 | Usual care follow-up after conservative treatment failure, arranged at the discretion of healthcare providers according to patient preference, including referral to a physiatrist or physiotherapist, pain clinic specialist, general practitioner, and/or spine surgeon. | Control |
| | Internet-based biopsychosocial rehabilitation delivered as eight weekly 2-hour group videoconferencing sessions of physical rehabilitation or physiotherapy and psychosocial intervention, in addition to usual care follow-up after conservative treatment failure. | Telemedicine |
| Groenveld 2023 | Wait-list follow-up for advanced pain treatment, with no additional treatment or virtual reality intervention during the 4-week study period. | Control |
| | Home-based virtual reality pain self-management delivered via Oculus Go, used independently for at least 10 minutes daily for 4 weeks after one supervised home session, while patients remained on the waiting list for advanced pain treatment. | VR |
| Kałużna 2023 | Face-to-face kinesiotherapy provided by the same physiotherapist in 10 daily 60-minute sessions over 2 weeks, including warm-up, general exercises, relaxation, and balance training on a rehabilitation mat. | Face-to-face care |
| | Face-to-face kinesiotherapy provided by the same physiotherapist in 10 daily 60-minute sessions over 2 weeks, including warm-up, general exercises, relaxation, and balance training on a stabilometric platform with visual feedback. | Biofeedback devices |
| Lazaridou 2023 | Usual care pain management information delivered as a brochure with psychoeducational materials, with no active treatment during the 12-week study period. | Control |
| | Internet-based EMG biofeedback and relaxation training delivered by a trained biofeedback instructor in weekly 40–45-minute Zoom sessions, with home practice of learned exercises for at least 30 minutes three times weekly, including posture training. | Telemedicine |

| | | |
|---|---|---|
| Maddox 2023 | Home-based sham virtual reality program delivered via the same head-mounted device, completed as one daily session for 56 days, with 2–16-minute sessions consisting of non-immersive 2D nature videos and music without pain management content. | Control |
| | Home-based immersive virtual reality pain self-management program delivered via RelieVRx using a head-mounted device, completed as one daily session for 56 days, with 2–16-minute sessions incorporating breathing biofeedback, mindfulness, and pain education. | VR |
| Park 2023 | Face-to-face conventional physical therapy provided by two licensed experienced physical therapists for 30 minutes three times weekly over four weeks, including heat, ultrasound, TENS, mobilization, manipulation, and therapeutic exercises according to standardized protocols. | Face-to-face care |
| | App-based AI-guided exercise program introduced in one initial face-to-face session and performed for 30 minutes three times weekly over four weeks, with individualized diagnosis and exercise prescription by the Dr AI platform confirmed by the investigator. | Mobile-App |
| Porwal 2023 | Face-to-face trunk-stabilization exercise without EMG biofeedback, performed under investigator supervision for 30–40 minutes per session, three times weekly for four weeks, plus warm-up, static bicycling, and stretching before exercise. | Face-to-face care |
| | Face-to-face trunk-stabilization exercise with EMG biofeedback, performed under investigator supervision for 30–40 minutes per session, three times weekly for four weeks, plus warm-up, static bicycling, and stretching before exercise. | Biofeedback devices |
| Sanabria-Mazo 2023 | Internet-based group acceptance and commitment therapy delivered by trained therapists in eight weekly 1.5-hour synchronous videoconference sessions, with homework documents and weekly reminders, in addition to treatment-as-usual. | Telemedicine |
| | Internet-based group behavioral activation therapy for depression delivered by trained therapists in eight weekly 1.5-hour synchronous videoconference sessions, with homework documents and weekly reminders, in addition to treatment-as-usual. | |
| | Usual care chronic pain management delivered by general practitioners in regular consultations of approximately 10 minutes, including medication prescription and recommendations for aerobic exercise, without additional active treatment during the study period. | Control |
| Fanuscu 2024 | Face-to-face supervised motor control exercise delivered by a physiotherapist three times per week for 8 weeks in a clinical setting, with real-time feedback, question handling, and attendance monitoring during sessions. | Face-to-face care |
| | Home-based motor control exercise delivered via instructional videos three times per week for 8 weeks, with weekly written consultations, exercise diary monitoring, optional text message support, and video-based feedback when needed. | Web-App |
| Holden 2024 | Face-to-face physiotherapy delivered by physical therapists once weekly for 6 weeks, including individualized treatment modalities such as exercise, manual therapy, advice, and education tailored to patient needs and progress. | Face-to-face care |
| | Face-to-face physiotherapy delivered by trained physical therapists once weekly for 6 weeks, incorporating motivational interviewing within sessions plus a self-directed smartphone app used every 1–3 days, with content tailored to patient readiness. | Mobile-App |

| | | |
|---|---|---|
| Kang 2024 | Face-to-face and home-based exercise therapy delivered by physicians over 8 weeks, including up to four in-person sessions with controlled intensity plus self-directed exercises based on educational materials. | Face-to-face care |
| | Smartphone app based digital therapeutics delivering CBT and exercise therapy over 8 weeks via a smartphone application with weekly CBT sessions and daily exercises, using AI-based tailoring and patient-reported feedback to individualize treatment plus education. | Mobile-App |
| McConnell 2024 | Face-to-face physiotherapy delivered by physical therapists with flexible frequency and duration according to routine clinical practice without additional digital or educational augmentation. | Face-to-face care |
| | Face-to-face physiotherapy delivered by physical therapists with flexible frequency and duration, augmented by VR-based pain neuroscience education during clinic visits using a head-mounted device with standardized sessions and relaxation and mindfulness components. | VR |
| Öztürk 2024 | Face-to-face physiotherapy delivering conservative therapy plus supervised stabilization exercises with pressure biofeedback over 4 weeks, 5 days per week, including progressive training with device-assisted feedback and home-based practice. | Biofeedback devices |
| | Face-to-face physiotherapy delivering conservative therapy plus supervised lumbar dynamic strengthening exercises over 4 weeks, 5 days per week, with therapist-guided feedback and home-based practice. | Face-to-face care |
| Priebe 2024 | Usual care delivered by general practitioners according to national guidelines, including clinical assessment, follow-up visits and standard management without access to the digital intervention. | Control |
| | App-based self-management intervention using the Kaia back pain app with ad libitum use, providing digital exercise, education and behavioral support, plus GP-led care and optional teleconsultation with pain specialists based on risk stratification. | Mobile-App |
| Santos 2024 | Participants received an evidence-based printed educational booklet during an individual consultation with a physical therapist lasting up to 1 hour. The booklet addressed pain self-management, relaxation exercises, walking, active coping, and remaining physically active, without specific or individually tailored exercise recommendations. | Control |
| | Participants attended weekly 90-minute, physiotherapist-led group sessions for 8 weeks, comprising 20 minutes of pain education and 60 minutes of supervised multimodal exercise based on cognitive-behavioral principles, supplemented by home exercise and motivational mobile-phone messages. | Telemedicine |
| Shi 2024 | Face-to-face physiotherapy delivering supervised strengthening and stretching exercises in outpatient settings 3 times weekly for 8 weeks, with in-person guidance, monitoring, and assessment by physiotherapists. | Face-to-face care |
| | App-based telerehabilitation delivering strengthening and stretching exercises via a smartphone platform with integrated sensors for monitoring performance, conducted 3 times weekly for 8 weeks with automated reminders, remote physiotherapist support, and digital assessment tracking. | Mobile-App |
| Yang 2024 | Waitlist control with usual activities and care without structured intervention during the study period | Control |

| | | |
|---|---|---|
| | Telemedicine-delivered tai chi, qigong, and meditation program provided via live online group classes by an instructor twice weekly for 12 weeks with guided supervision and optional home practice plus usual care | Telemedicine |
| Yeldan 2024 | Face-to-face physiotherapy delivering the same core stabilization and activity training three times weekly for 4 weeks, with therapist-provided feedback to guide posture and movement during exercises. | Face-to-face care |
| | Face-to-face physiotherapy delivering core stabilization and activity training three times weekly for 4 weeks, with exercises supported by wearable sensor–based auditory and tactile biofeedback via a mobile applications to guide movement. | Biofeedback devices |
| Choi 2025 | Face-to-face physiotherapy delivering core stability exercise sessions three times weekly for 6 weeks plus instructed abdominal contraction during daily activities without real-time biofeedback. | Face-to-face care |
| | Face-to-face physiotherapy delivering core stability exercise sessions three times weekly for 6 weeks plus additional training using a wearable waist sensor providing real-time vibratory biofeedback to maintain abdominal contraction during daily activities. | Biofeedback devices |
| Feng 2025 | Home-based paper-manual exercise therapy delivered 3 times weekly for 8 weeks, plus weekly app-based patient education, with initial physiotherapist instruction but no routine ongoing supervision unless help was requested. | Control |
| | App-based individualized exercise therapy delivered 3 times weekly for 8 weeks, plus weekly app-based patient education and weekly WeChat video–based health coaching, with physiotherapist supervision and progression adjustment during video sessions. | Mobile-App |
| Geraghty 2025 | Usual care for low back pain allowing unrestricted access to primary and secondary care services, including assessment, advice for self-management, pharmacotherapy, and referrals to physiotherapy, pain clinics, or psychological interventions. | Control |
| | Internet-based self-management intervention providing automated behavioural support and advice through six weekly sessions over 6 weeks with goal setting, self-monitoring, and tailored feedback, in addition to unrestricted usual care. | Web-App |
| | Internet-based self-management intervention plus physiotherapist telephone support delivered over 6 weeks, including up to three calls providing encouragement, reassurance, and goal-setting support, in addition to unrestricted usual care. | |
| Hsieh 2025 | Face-to-face traditional exercise delivered under therapist guidance for six sessions over 2 weeks, with 15-minute therapeutic exercise following 20 minutes of hot pack therapy plus 20 minutes of TENS, and weekly difficulty adjustment. | Face-to-face care |
| | Face-to-face exergaming delivered under investigator supervision for six sessions over 2 weeks, with 15-minute interactive video game training following 20 minutes of hot pack therapy plus 20 minutes of TENS, and weekly difficulty adjustment. | Exergaming |
| Massah 2025 | Face-to-face core stability exercise delivered on a yoga mat under therapist supervision in a single session matched in duration and intensity monitoring to the intervention group. | Face-to-face care |

| | | |
|---|---|---|
| | Virtual reality–based exergaming exercise delivered using an Xbox Kinect system under therapist supervision in a single 30–45 minute session including warm-up and cool-down with brief rest intervals between activities. | Exergaming |
| Kent 2023 | Usual care allowing participants to receive any treatment recommended by their health-care providers or chosen independently in the community without structured intervention from the study. | Control |
| | Face-to-face physiotherapy delivering individualized cognitive functional therapy over 12 weeks with up to 7 sessions plus a booster session, incorporating movement retraining, behavioral change, and lifestyle coaching based on a biopsychosocial approach. | Face-to-face care |
| | Face-to-face physiotherapy delivering individualized cognitive functional therapy over 12 weeks with up to 7 sessions plus a booster session, augmented by wearable movement sensors providing biofeedback for assessment and movement retraining. | Biofeedback devices |
| Villatoro-Luque 2025 | Face-to-face physiotherapy delivered by a physiotherapist over 8 weeks with two 30-minute sessions per week, using supervised therapeutic exercise with pain-guided individualization plus pain neurophysiology education. | Face-to-face care |
| | Telephone-delivered telerehabilitation exercise delivered by a physiotherapist over 8 weeks with two 30-minute sessions per week, using instructional videos plus weekly videoconference follow-up, pain-guided individualization, and pain neurophysiology education. | Telemedicine |
| Mudd 2025 | Face-to-face guideline-informed physiotherapy over 12 weeks, including three consultations providing back pain education, advice, and exercise, without lifestyle counseling beyond exercise recommendations. | Face-to-face care |
| | Face-to-face plus telephone-delivered physiotherapy and lifestyle care over 12 weeks, including up to four physiotherapist sessions and one dietitian session, plus educational resources and tailored health coaching for weight, physical activity, and diet. | Telemedicine |
| Tawfek 2025 | Home-based exercise delivered as an unsupervised home program over 12 weeks, following the same warm-up, core stabilization, stretching, and cool-down protocol, plus patient education. | Control |
| | Internet-based exercise delivered by a certified physiotherapist in real-time Zoom sessions twice weekly for 24 sessions over 12 weeks, following a warm-up, core stabilization, stretching, and cool-down program, plus patient education | Telemedicine |
| | Internet-based exercise delivered as prerecorded videos over 12 weeks, with weekly follow-up messages by email, WhatsApp, or Telegram according to patient preference, following the same exercise program, plus patient education. | Mobile-App |
| Menekseoglu 2026 | Face-to-face physiotherapy delivered over 3 weeks with 15 supervised sessions, including 20-minute TENS, 20-minute hot pack application, and 30-minute dynamic lumbar stabilization exercises without virtual reality hypnotherapy. | Face-to-face care |
| | Face-to-face physiotherapy plus virtual reality hypnotherapy delivered over 3 weeks with 15 sessions, including supervised TENS, hot pack, and dynamic lumbar stabilization exercises in addition to 20-minute VR sessions using a headset-based seaside relaxation program. | VR |

<table>
<tr><td rowspan="2">Moreno-Ligero 2026</td><td>Face-to-face booklet-based self-management exercise introduced by a physiotherapist with recommended home exercise three times per week, plus written pain information, exercise descriptions and photos, and self-management strategies to control exercise intensity.</td><td>Face-to-face care</td></tr>
<tr><td>App-based self-management exercise delivered through a mobile applications with recommended home exercise three times per week, following an initial face-to-face physiotherapist instruction session, plus a booklet with pain information and self-management strategies.</td><td>Mobile-App</td></tr>
<tr><td rowspan="3">Spiegel 2026</td><td>Virtual reality–based sham control delivered remotely as a prescribed daily 56-session program over 56 days, using 2D nature footage with neutral music and no therapeutic or interactive components.</td><td>Control</td></tr>
<tr><td>Virtual reality–based skills training delivered remotely as a prescribed daily 56-session program over 56 days, including mindfulness, relaxation, biofeedback, pain education, breathing training, and executive-function games, plus on-demand access to additional modules.</td><td rowspan="2">VR</td></tr>
<tr><td>Virtual reality–based distraction therapy delivered remotely as a prescribed daily 56-session program over 56 days, using immersive 360-degree videos matched to the skills-based program in hardware, schedule, duration, and interface.</td></tr>
<tr><td rowspan="2">Ahmed 2026</td><td>Conventional pelvic rocking exercise delivered three sessions per week for 8 weeks, involving posterior pelvic tilting performed without game-based digital feedback, together with the study-prescribed conventional physical therapy components.</td><td>Face-to-face care</td></tr>
<tr><td>TBed-based game-mediated pelvic rocking exercise delivered three sessions per week for 8 weeks, in which trunk and pelvic pressure generated during therapeutic movement controlled an on-screen game and provided real-time interactive feedback, together with the same background physical therapy provided to the conventional group.</td><td>Exergaming</td></tr>
<tr><td rowspan="2">Alahmri 2026</td><td>Conventional in-person rehabilitation delivered over 6 weeks according to routine clinical practice, with treatment individualized by the physiotherapist and including supervised exercise, hands-on techniques, and other therapeutic modalities as clinically appropriate; a fixed session frequency or session duration was not reported.</td><td>Face-to-face care</td></tr>
<tr><td>Telerehabilitation delivered remotely over 6 weeks using Zoom videoconferencing supplemented by telephone calls and WhatsApp messaging, providing individualized aerobic, stretching, and core-strengthening exercise, pain education, self-management support, and treatment progression according to patient tolerance; a fixed session frequency or duration was not reported.</td><td>Telemedicine</td></tr>
<tr><td rowspan="2">Jeong 2026</td><td>Face-to-face deep muscle strengthening exercise with the abdominal drawing-in manoeuvre, delivered for approximately 30 minutes per session, three times per week for 4 weeks, including leg raises, bilateral leg tilts, and curl-up exercises, with warm-up and cool-down stretching and no biofeedback.</td><td>Face-to-face care</td></tr>
<tr><td>Biofeedback-assisted deep muscle strengthening exercise delivered for approximately 30 minutes per session, three times per week for 4 weeks, using the same abdominal drawing-in and strengthening program plus Stabilizer pressure biofeedback to provide real-time lumbar pressure feedback during exercise.</td><td>Biofeedback devices</td></tr>
<tr><td>Lechauve 2026</td><td>Face-to-face multidisciplinary rehabilitation and self-management education delivered 5 days per week for 3 weeks, including daily physiotherapy, occupational therapy, adapted physical activity, hydrotherapy, and self-management education.</td><td>Face-to-face care</td></tr>
</table>

| | | |
|---|---|---|
| | Smartphone app–supported multidisciplinary rehabilitation delivered alongside the same 5-day-per-week, 3-week face-to-face program, plus access to the Mon Coach Dos app and three 1-hour app-training sessions delivered once weekly, providing exercise videos, pain information, physical-activity guidance, and support for a personalized home exercise program. | Mobile-App |
| Slatman 2026 | Guideline-based usual-care physiotherapy delivered over a 12-week treatment period, individualized according to Dutch clinical physiotherapy guidelines for chronic low back pain; no uniform visit frequency was imposed because of the pragmatic design. | Face-to-face care |
| | Guideline-based physiotherapy integrated with home-based immersive VR over 12 weeks, comprising the same usual-care physiotherapy plus unsupervised VR headset use recommended five times per week for 10–30 minutes per session, with pain education, relaxation/distraction, exercise, and graded-exposure modules selected according to patient needs. | VR |
| Valenzuela-Pascual 2026 | Conventional primary-care management during the 15-day intervention period, without access to the study-specific web intervention; participants were instructed not to seek additional treatment other than consultation with their family physician if required. | Control |
| | Gamified web-based pain neuroscience education provided for 15 days, with password-protected access available at any time throughout the intervention period and no fixed number of sessions prescribed, including educational videos, nonlinear personalized navigation, interactive pain-belief tasks, and gamified learning activities. | Web-App |

Note: Treatment node assignment was based on the classification of each intervention arm according to its principal mode of delivery. AI, artificial intelligence; CBT, cognitive behavioural therapy; EMG, electromyography; GP, general practitioner; SMS, short message service; VR, virtual reality; Mobile-App, mobile applications; Web-App, web applications.

**Section 5.** Study characteristics

**Table S5.** Characteristics of the 84 randomised controlled trials included in the systematic review

| Study ID | Country | Intervention duration | Total sample size | Overall mean age | Female (%) | Pain type | Study design | Treatment nodes | Funding source |
|---|---|---|---|---|---|---|---|---|---|
| Iles 2011[10] | Australia | 7 weeks | 30 | 39.5 | 40 | A/S-NSLBP | RCT | Control<br>Telemedicine | Angliss Hospital |
| Carpenter 2012[11] | USA | 6 weeks | 141 | 42.5 | 83 | CLBP | RCT | Control<br>Web-App | National Institute of Arthritis and Musculoskeletal and Skin Diseases. |
| Del Pozo-Cruz 2012[12-14] | Spain | 9 months | 100 | 46.2 | 86.7 | S-NSLBP | RCT | Control<br>Web-App | University of Extremadura,Junta de Extremadura and European Union Regional Development Funds |
| Krein 2013[15] | USA | 12 months | 229 | 51.6 | 12.6 | CNSLBP | RCT | Control<br>Web-App | US Department of Veterans Affairs (HSR&D); additional institutional and investigator support. |
| McDonough 2013[16] | Ireland | 8 weeks | 57 | 48.9 | 55.4 | CNSLBP | RCT | Face-to-face care<br>Telemedicine | Economic and Social Research Council(ESRC) |
| Park 2013[17] | South Korea | 8 weeks | 24 | 44.3 | NR | CLBP | RCT | Control<br>Face-to-face care<br>Exergaming | Soonchunhyang University |
| Kim 2014[18] | Korea | 4 weeks | 30 | 44.33 | 100.00 | CLBP | RCT | Face-to-face care<br>Exergaming | NR |
| Irvine 2015[19] | USA | 8 weeks | 597 | NR | 59.97 | NSLBP | RCT | Control<br>Mobile-App | US National Institutes of Health, National Institute of Arthritis and Musculoskeletal and Skin Diseases |

| | | | | | | | | | |
|---|---|---|---|---|---|---|---|---|---|
| Geraghty2018[20] | UK | 6 weeks | 87 | 57.95 | 58.62 | LBP | RCT | Control<br>Web-App | National Institute for Health Research (NIHR) Research for Patient Benefit programme |
| Tan 2015[21] | USA | 8 weeks | 159 | 55 | NR | CLBP | RCT | Face-to-face care<br>Telemedicine<br>Biofeedback devices | Veterans Health Administration Rehabilitation Research and Development Service. |
| Schaller 2016[22] | Germany | 6 months | 412 | 50.42 | 69.42 | CLBP | RCT | Face-to-face care<br>Web-App | German Statutory Pension Insurance Rhineland |
| Yilmaz Yelvar 2017[23] | Turkey | 2 weeks | 46 | 49.55 | 63.64 | S/C-NSLBP | RCT | Face-to-face care<br>VR | NR |
| Chhabra 2018[24] | India | 12 weeks | 93 | 41.19 | NR | CLBP | RCT | Face-to-face care<br>Mobile-App | Snapcare Technologies Pvt. Ltd |
| Goode 2018[25] | USA | 12 weeks | 60 | 70.3 | 6.67 | CLBP | RCT | Control<br>Telemedicine | Department of Veterans Affairs Rehabilitation and Research Development and the Center of Innovation for Health Services Research in Primary Care at the Durham VA Health Care System. |
| Williams 2018[26] | Australia | 6 months | 160 | 56.7 | 59.1 | CLBP | RCT | Control<br>Telemedicine | Hunter New England Local Health District, the University of Newcastle and the Hunter Medical Research Institute |

| | | | | | | | | | |
|---|---|---|---|---|---|---|---|---|---|
| Amorim 2019[27] | Australia | 6 months | 68 | 58.3 | 50 | CLBP | Pilot RCT | Control<br>Telemedicine | Medibank Health Research Fund |
| Mbada 2019[28] | Nigeria | 8 weeks | 47 | 48.8 | 72.34 | CNSLBP | RCT | Face-to-face care<br>Mobile-App | African Doctoral Dissertation Research Fellowship (ADDRF) and the African Population and Health Research Center (APHRC) |
| Petrozzi 2019[29] | Australia | 8 weeks | 108 | 50.4 | 50 | CNSLBP | RCT | Face-to-face care<br>Web-App | NR |
| Shebib 2019[30] | USA | 12 weeks | 177 | 43 | 41 | CNSLBP | RCT | Control<br>Mobile-App | Hinge Health, Inc |
| Toelle 2019[31] | Germany | 12 weeks | 101 | 41.98 | 70.21 | S/C-NSLBP | RCT | Face-to-face care<br>Mobile-App | Kaia (Kaia Health Software GmbH, Munich, Germany) |
| Yang 2019[32] | China | 4 weeks | 8 | 40.75 | 50 | CNSLBP | RCT | Face-to-face care<br>Mobile-App | NR |
| Zadro 2019[33] | Australia | 8 weeks | 60 | 67.8 | 51.7 | CLBP | RCT | Control<br>Exergaming | NR |
| Almhdawi 2020[34] | Jordan | 6 weeks | 41 | 41.08 | 53.66 | CNSLBP | RCT | Control<br>Mobile-App | Jordan University and European Union |
| Matheve 2020[35] | Belgium | 1 day | 84 | 43.15 | 64 | CLBP | RCT | Face-to-face care<br>VR | NR |
| Nambi 2020[36] | Saudi Arabia | 4 weeks | 45 | 20.75 | 0 | CLBP | RCT | Control<br>VR | Princess Nourah Bint Abdulrahman University |
| Tomruk 2020[37] | Turkey | 12 weeks | 42 | NR | NR | CLBP | RCT | Face-to-face care<br>Biofeedback devices | NR |

| | | | | | | | | | |
|---|---|---|---|---|---|---|---|---|---|
| Alzahrani 2021[38] | Australia | 8 weeks | 26 | 43.6 | 42.3 | NSLBP | RCT | Face-to-face care<br>Biofeedback devices | Taif University |
| Dadarkhah 2021[39] | Iran | 4 weeks | 56 | 49.5 | 57.14 | CNSLBP | RCT | Face-to-face care<br>Telemedicine | NR |
| Garcia 2021[40-43] | USA | 8 weeks | 188 | 51.45 | 76.54 | CLBP | RCT | Control<br>VR | AppliedVR, Inc |
| Kazemi 2021[44] | Iran | 1 day | 180 | 36.66 | 100 | LBP | RCT | Control<br>Face-to-face care<br>Web-App | Tarbiat Modares University |
| Kim 2021[45] | Korea | 8 weeks | 43 | 57.82 | 25.58 | CLBP | RCT | Control<br>Telemedicine | NR |
| Lang 2021[46] | Canada | 12 weeks | 174 | 46 | 60.1 | CLBP | RCT | Control<br>Telemedicine | Saskatchewan Health Research Foundation (SHRF) |
| Li 2021[47] | China | 2 weeks | 34 | 23.68 | 73.53 | CNSLBP | RCT | Face-to-face care<br>VR | the National Natural Science Foundation of China;the Guangdong Province Medical Science Technology Research Grant;the Guangdong Basic and Applied Basic Research Foundation; the Guangzhou Science;Technology Program key projects and the NonProfit Central Research Institute Fund of Chinese Academy of Medical Sciences |
| Nambi 2021a[48] | Saudi Arabia | 4 weeks | 60 | 23.1 | 0 | CLBP | RCT | Control<br>VR | - |

| | | | | | | | | | |
|---|---|---|---|---|---|---|---|---|---|
| Nambi 2021b[49] | Saudi Arabia | 4 weeks | 54 | 21.87 | 0 | CLBP | RCT | Control<br>VR | Prince Sattam Bin Abdulaziz University |
| Nambi 2021c[50] | Saudi Arabia | 4 weeks | 60 | 21.27 | 0 | CLBP | RCT | Control<br>VR | Princess Nourah bint Abdulrahman University |
| Sandal 2021[51] | Denmark | 12 weeks | 461 | 47.5 | 55.31 | NSLBP | RCT | Control<br>Mobile-App | European Union |
| Sato 2021[52] | Japan | 8 weeks | 40 | 52.46 | 47.5 | CLBP | RCT | Control<br>Exergaming | NR |
| Afzal 2022[53] | Pakistan | 4 weeks | 84 | 37.85 | 66.66 | CLBP | RCT | Face-to-face care<br>VR | NR |
| Eccleston 2022[54] | UK | 8 weeks | 42 | 54.69 | 88.1 | CLBP | RCT | Control<br>VR | Orion Corporation, Business Finland and the Finnish government |
| Itoh 2022[55] | Japan | 12 weeks | 101 | 47.38 | 44.44 | CLBP | RCT | Control<br>Mobile-App | Shionogi & Co, Ltd, Osaka, Japan |
| Koppenaal 2022[56] | Netherlands | 3/12 weeks | 208 | 47.68 | 49.04 | NSLBP | Cluster RCT | Face-to-face care<br>Mobile-App | NR |
| Lara-Palomo 2022[57] | Spain | 8 weeks | 80 | 47.91 | 58.1 | CLBP | RCT | Control<br>Web-App | Andalusian Health Service, Junta de Andalucia,Carlos III Health Institute and European Social Fund |
| Meinke 2022[58] | Switzerland | 3 weeks | 27 | 40.48 | 62.96 | NSLBP | RCT | Control<br>Exergaming | Swiss National Science Foundation |
| Nambi 2022[59] | Saudi Arabia | 4 weeks | 60 | 22.97 | 0 | CNSLBP | RCT | Face-to-face care<br>VR | Prince Sattam bin Abdulaziz University |
| Yalfani 2022[60] | Iran | 8 weeks | 25 | 67.56 | 100 | CLBP | RCT | Control<br>VR | NR |

| | | | | | | | | | |
|---|---|---|---|---|---|---|---|---|---|
| Fatoye 2022[61] | Nigeria | 8 weeks | 57 | 48.24 | NR | CNSLBP | RCT | Face-to-face care<br>Exergaming | African Population and Health Research Centre (APHRC) and International Development Research Centre (IDRC) |
| Cui 2023[62] | China | 8 weeks | 140 | 52.5 | 67.86 | CLBP | RCT | Face-to-face care<br>Mobile-App | NR |
| Garreta-Catala 2023[63] | Spain | 8 weeks | 18 | 53.15 | 72.22 | CLBP | RCT | Control<br>Telemedicine | Justimplant and the Catalan Institute of Health |
| Groenveld 2023[64] | Netherlands | 4 weeks | 40 | 51.5 | 82.5 | CLBP | RCT | Control<br>VR | European Regional Development Fund (ERDF) |
| Kałużna 2023[65] | Poland | 2 weeks | 100 | 38.08 | 76 | CNSLBP | RCT | Face-to-face care<br>Biofeedback devices | NR |
| Kent 2023[66 67] | Australia | 12 weeks | 492 | 47.3 | 59.35 | CLBP | RCT | Control<br>Biofeedback devices<br>Face-to-face care | Australian National Health and Medical Research Council (NHMRC; grant 1145271) and Curtin University. |
| Lazaridou 2023[68] | USA | 8 weeks | 81 | 45.8 | NR | CLBP | RCT | Control<br>Telemedicine | the JOGO Health Inc |
| Maddox 2023[69 70] | USA | 8 weeks | 1093 | 50.75 | 72.35 | CLBP | RCT | Control<br>VR | AppliedVR, Inc |
| Park 2023[71] | Korea | 4 weeks | 100 | 35.5 | 40 | CLBP | RCT | Face-to-face care<br>Mobile-App | Brain Korea and National Research Foundation of Korea (NRF) |
| Porwal 2023[72] | Saudi Arabia | 4 weeks | 40 | 30.62 | NR | CNSLBP | RCT | Face-to-face care<br>Biofeedback devices | King Khalid University, Kingdom of Saudi Arabia |
| Sanabria-Mazo 2023[73] | Spain | 8 weeks | 234 | 54.53 | 67.52 | CLBP | RCT | Control<br>Telemedicine | Institute of Health Carlos III and European Union ERDF |

| | | | | | | | | | |
|---|---|---|---|---|---|---|---|---|---|
| Fanuscu 2024[74 75] | Turkey | 8 weeks | 42 | 39.6 | 64.3 | CLBP | RCT | Face-to-face care<br>Web-App | NR |
| Holden 2024[76] | Australia | 6 weeks | 46 | 43.7 | 52.2 | SLBP | Cluster RCT | Face-to-face care<br>Mobile-App | La Trobe University |
| Kang 2024[77] | Korea | 8 weeks | 46 | 38.3 | 79.1 | CLBP | Pilot RCT | Face-to-face care<br>Mobile-App | Daegu-Gyeongbuk Medical Innovation Foundation |
| McConnell 2024[78] | USA | 6 weeks | 52 | 46.2 | 62.5 | CLBP | RCT | Face-to-face care<br>VR | NR |
| Öztürk 2024[79] | Turkey | 4 weeks | 40 | 34 | 50 | CLBP | RCT | Biofeedback devices<br>Face-to-face care | NR |
| Priebe 2024[80] | Germany | NR | 1237 | 40.8 | 64.8 | A/S-NSLBP | Cluster RCT | Control<br>Mobile-App | German Innovationsfonds (G-BA) |
| Santos 2024[81] | Brazil | 8 weeks | 40 | 67.8 | 82.5 | CNSLBP | RCT | Control<br>Telemedicine | Conselho Nacional Desenvolvimento Científico e Tecnológico(CNPq), Ministry of Science, Technology and Innovations, Brazil |
| Shi 2024[82] | China | 8 weeks | 54 | 38.7 | 59.3 | CNSLBP | RCT | Face-to-face care<br>Mobile-App | National High-Level Hospital Clinical Research Funding |
| Yang 2024[83] | USA | 12 weeks | 350 | 58.8 | 79.4 | LBP | RCT | Control<br>Telemedicine | Qi Balance LLC |
| Yeldan 2024[84] | Turkey | 4 weeks | 40 | 46.5 | 63.2 | CNSLBP | RCT | Face-to-face care<br>Biofeedback devices | NR |
| Choi 2025[85] | Korea | 6 weeks | 32 | 54.7 | 55 | CLBP | RCT | Face-to-face care<br>Biofeedback devices | NR |

| | | | | | | | | | |
|---|---|---|---|---|---|---|---|---|---|
| Feng 2025[86] | China | 8 weeks | 78 | NR | 50 | CLBP | RCT | Control<br>Mobile-App | Natural Science Foundation of Sichuan Province;National Natural Science Foundation of China;Sichuan University;West China Hospital, Sichuan University and Deyang City Science and Technology Bureau |
| Geraghty 2025[87] | UK | 6 weeks | 825 | 54.2 | 58.3 | LBP | RCT | Control<br>Web-App | National Institute for Health and Care Research (NIHR) and Health Technology Assessment |
| Hsieh 2025[88] | China<br>Taiwan | 2 weeks | 70 | 59.6 | 62.85 | CNSLBP | RCT | Face-to-face care<br>Exergaming | Shin Kong Wu HoSu Memorial Hospital and the Ministry of Science and Technology, Taiwan |
| Massah 2025[89] | Iran | 1 day | 40 | 31 | 40 | CNSLBP | RCT | Face-to-face care<br>Exergaming | Tarbiat Modares University |
| Villatoro-Luque 2025[90] | Spain | 8 weeks | 70 | 43.07 | 50 | CNSLBP | RCT | Face-to-face care<br>Telemedicine | NR |
| Mudd 2025[91] | Australia | 26 weeks | 346 | 50.2 | 31.98 | CNSLBP | RCT | Face-to-face care<br>Telemedicine | NR |
| Tawfek 2025[92] | Turkey | 12 weeks | 72 | 41.26 | 43.06 | CNSLBP | RCT | Control<br>Telemedicine<br>Mobile-App | NR |
| Menekseoglu 2026[93] | Turkey | 3 weeks | 62 | 47.62 | 66.7 | CNSLBP | RCT | Face-to-face care<br>VR | NR |
| Moreno-Ligero 2026[94] | Spain | NR | 99 | NR | 67.7 | CLBP | RCT | Face-to-face care<br>Mobile-App | NR |

| | | | | | | | | | |
|---|---|---|---|---|---|---|---|---|---|
| Spiegel 2026[95] | USA | 8 weeks | 385 | 54.1 | 62.6 | CLBP | RCT | Control<br>VR | NIH/NIAMS, AppliedVR and the Marc and Sheri Rapaport Fund for Digital Health Science and Precision Health at Cedars-Sinai and NIH National Center |
| Ahmed 2026[96] | Saudi Arabia | 8 weeks | 100 | 40.48 | 63.00 | NSLBP | RCT | Face-to-face care<br>Exergaming | Scientific Research Deanship, University of Ha'il, Saudi Arabia (project number RG-20202) |
| Alahmri 2026[97] | Saudi Arabia | 2 weeks | 46 | 49.55 | 63.60 | NSLBP | RCT | Face-to-face care<br>Telemedicine | NR |
| Jeong 2026[98] | Korea | 12 weeks | 32 | 49.93 | 75.00 | CNSLBP | RCT | Face-to-face care<br>Biofeedback devices | NR |
| Lechauve 2026[99] | France | 3 weeks | 110 | 20.55 | 0.00 | CLBP | RCT | Face-to-face care<br>Mobile-App | University Hospital of Clermont-Ferrand (AOI funding DGA/AB/MFC 2019-8) |
| Slatman 2026[100] | Netherlands | 2 weeks | 48 | 47.4 | 100.00 | LBP | Cluster RCT | Face-to-face care<br>VR | ZonMw, case 10270032021502 |
| Valenzuela-Pascual 2026[101] | Spain | 8 weeks | 37 | 20.51 | 0.00 | CLBP | RCT | Control<br>Web-App | Catedra ASISA-UdL, University of Lleida, Spain |

**Note:** A/S-NSLBP, acute and subacute non-specific low back pain; CLBP, chronic low back pain; CNSLBP, chronic non-specific low back pain; LBP, low back pain; Mobile-App, mobile applications; NR, not reported; NSLBP, non-specific low back pain; RCT, randomised controlled trial; S/C-NSLBP, subacute and chronic non-specific low back pain; SLBP, subacute low back pain; S-NSLBP, subacute non-specific low back pain; VR, virtual reality; Web-App, web applications.

**Section 6.** Outcome measures and selection

**Table S6.** Outcome measures reported in included studies

| Study ID | Intervention arms | Pain intensity | Physical function | Pain-related fear avoidance | Health-related quality of life | Self-efficacy | Depression | Anxiety |
|---|---|---|---|---|---|---|---|---|
| Iles 2011 | Control; Telemedicine | NR | ODI | NR | NR | PSEQ | NR | NR |
| Carpenter 2012 | Control; Web-App | PAQ | RMDQ | NR | NR | PSES | NR | NR |
| Del Pozo-Cruz 2012 | Control; Web-App | NR | RMDQ | NR | NR | NR | NR | NR |
| Krein 2013 | Control; Web-App | NRS | RMDQ | FABQ-PA | NR | ERS | NR | NR |
| McDonough 2013 | Face-to-face care; Telemedicine | NRS | ODI | FABQ-PA | EQ-5D | PASES | NR | NR |
| Park 2013 | Control; Face-to-face care; Exergaming | VAS | ODI | FABQ | NR | NR | NR | NR |
| Kim 2014 | Face-to-face care; Exergaming | VAS | NR | NR | NR | NR | NR | NR |
| Irvine 2015 | Control; Mobile-App | NRS | NR | TSK | NR | Other scale | NR | NR |
| Geraghty 2018 | Control; Web-App | NRS | RMDQ | NR | NR | NR | NR | NR |
| Schaller 2016 | Face-to-face care; Telemedicine; Biofeedback devices | BPI intensity | NR | NR | NR | NR | NR | NR |
| Tan 2015 | Face-to-face care; Web-App | Other scale | NR | NR | NR | NR | NR | NR |
| Yilmaz Yelvar 2017 | Face-to-face care; VR | VAS | ODI | TSK | NR | NR | NR | NR |
| Chhabra 2018 | Face-to-face care; Mobile-App | NRS | mODI | NR | NR | NR | NR | NR |
| Goode 2018 | Control; Telemedicine | NR | RMDQ | NR | NR | NR | NR | NR |
| Williams 2018 | Control; Telemedicine | NRS | RMDQ | FABQ-PA | SF-12 PCS | NR | DASS-D | DASS-A |
| Amorim 2019 | Control; Telemedicine | NRS | RMDQ | NR | NR | NR | NR | NR |
| Mbada 2019 | Face-to-face care; Mobile-App | QVAS | RMDQ | NR | SF-12 PCS | NR | NR | NR |
| Petrozzi 2019 | Face-to-face care; Web-App | NRS | RMDQ | PCS | NR | PSEQ | DASS-D | DASS-A |
| Shebib 2019 | Control; Mobile-App | VAS | ODI | NR | NR | NR | NR | NR |

| | | | | | | | | |
|---|---|---|---|---|---|---|---|---|
| Toelle 2019 | Face-to-face care; Mobile-App | NRS | NR | NR | VR-12 PCS | NR | NR | NR |
| Yang 2019 | Face-to-face care; Mobile-App | VAS | RMDQ | NR | SF-36 | PSEQ | NR | NR |
| Zadro 2019 | Control; Exergaming | NRS | RMDQ | TSK | NR | PSEQ | NR | NR |
| Almhdawi 2020 | Control; Mobile-App | VAS | ODI | NR | SF-12 PCS | NR | DASS-D | DASS-A |
| Matheve 2020 | Face-to-face care; VR | NRS | NR | NR | NR | NR | NR | NR |
| Nambi 2020 | Control; VR | VAS | NR | NR | NR | NR | NR | NR |
| Tomruk 2020 | Biofeedback devices; Face-to-face care | NRS | ODI | NR | NR | NR | NR | NR |
| Alzahrani 2021 | Face-to-face care; Biofeedback devices | VAS | ODI | TSK | NR | NR | NR | NR |
| Dadarkhah 2021 | Face-to-face care; Telemedicine | VAS | ODI | NR | NR | NR | NR | NR |
| Garcia 2021 | Control; VR | DVPRS | NR | NR | NR | NR | NR | NR |
| Kazemi 2021 | Control; Face-to-face care; Web-App | VAS | QBPDS | NR | NR | NR | NR | NR |
| Kim 2021 | Control; Telemedicine | BPI intensity | ODI | NR | NR | NR | NR | NR |
| Lang 2021 | Control; Telemedicine | ODI Pain rating | ODI | FABQ-PA | EQ-5D-5L | PASES | NR | NR |
| Li 2021 | Face-to-face care; VR | VAS | ODI | NR | NR | NR | NR | NR |
| Nambi 2021a | Control; VR | VAS | NR | TSK | NR | NR | NR | NR |
| Nambi 2021b | Control; VR | VAS | NR | TSK | NR | NR | NR | NR |
| Nambi 2021c | Control; VR | NRS | NR | NR | PCS fitness index | NR | NR | NR |
| Sandal 2021 | Control; Mobile-App | NRS | RMDQ | FABQ-PA | EQ-5D-5L | PSEQ | NR | NR |
| Sato 2021 | Control; Exergaming | VAS | NR | TSK | NR | PSEQ | NR | NR |
| Afzal 2022 | Face-to-face care; VR | NRS | mODI | NR | NR | NR | NR | NR |
| Eccleston 2022 | Control; VR | NRS | ODI | TSK | EQ-5DHealth state | NR | NR | NR |
| Itoh 2022 | Control; Mobile-App | NRS | NR | TSK | NR | NR | Other scale | NR |

| | | | | | | | | |
|---|---|---|---|---|---|---|---|---|
| Koppenaal 2022 | Face-to-face care; Mobile-App | NRS | ODI | FABQ | EQ-5DHealth state | GSES | NR | NR |
| Lara-Palomo 2022 | Control; Web-App | VAS | ODI | TSK | SF-36 | NR | NR | NR |
| Meinke 2022 | Control; Exergaming | NRS | RMDQ | TSK | NR | NR | NR | NR |
| Nambi 2022 | Face-to-face care; VR | VAS | NR | NR | NR | NR | NR | NR |
| Yalfani 2022 | Control; VR | VAS | NR | NR | SF-36 | NR | NR | NR |
| Fatoye 2022 | Face-to-face care; Exergaming | NR | ODI | NR | NR | NR | NR | NR |
| Cui 2023 | Face-to-face care; Mobile-App | VAS | ODI | NR | NR | NR | PHQ-9 | GAD-7 |
| Garreta-Catala 2023 | Control; Telemedicine | NR | ODI | NR | SF-12 PCS | NR | HADS-D | HADS-A |
| Groenveld 2023 | Control; VR | NR | mODI | PCS | SF-12 PCS | NR | HADS-D | HADS-A |
| Kałużna 2023 | Face-to-face care; Biofeedback devices | NRS | ODI | NR | SF-36 PCS | NR | NR | NR |
| Kent 2023 | Control; Face-to-face care; Biofeedback devices | NRS | RMDQ | FABQ-PA | NR | PSEQ | NR | NR |
| Lazaridou 2023 | Control; Telemedicine | BPI intensity | ODI | PCS | NR | NR | HADS-D | HADS-A |
| Maddox 2023 | Control; VR | BPI intensity | ODI | NR | NR | NR | PROMIS-Depression | PROMIS-Anxiety |
| Park 2023 | Face-to-face care; Mobile-App | NRS | ODI | NR | SF-12 PCS | NR | NR | NR |
| Porwal 2023 | Face-to-face care; Biofeedback devices | VAS | ODI | NR | NR | NR | NR | NR |
| Sanabria-Mazo 2023 | Telemedicine; Control | NRS | NR | PCS | NR | NR | DASS-D | DASS-A |
| Fanuscu 2024 | Face-to-face care; Web-App | VAS | ODI | PCS | NR | NR | NR | NR |
| Holden 2024 | Face-to-face care; Mobile-App | NR | ODI | NR | NR | PSEQ | NR | NR |
| Kang 2024 | Face-to-face care; Mobile-App | NRS | ODI | FABQ | EQ-5D-3L | NR | PHQ-9 | NR |
| McConnell 2024 | Face-to-face care; VR | NRS | ODI | FABQ-PA | NR | PSEQ | NR | NR |
| Öztürk 2024 | Biofeedback devices; Face-to-face care | VAS | ODI | FABQ-PA | SF-12 PCS | NR | NR | NR |
| Priebe 2024 | Control; Mobile-App | NRS | NR | NR | NR | NR | NR | NR |
| Santos 2024 | Control; Telemedicine | NRS | RMDQ | NR | NR | CPSS | CES-D | NR |

| | | | | | | | | |
|---|---|---|---|---|---|---|---|---|
| Shi 2024 | Face-to-face care; Mobile-App | NRS | ODI | FABQ | SF-36 | NR | NR | NR |
| Yang 2024 | Control; Telemedicine | VAS | ODI | NR | SF-36 | NR | NR | NR |
| Yeldan 2024 | Face-to-face care; Biofeedback devices | VAS | RMDQ | FABQ-PA | SF-36 | NR | BDI | NR |
| Choi 2025 | Face-to-face care; Biofeedback devices | VAS | ODI | NR | NR | NR | NR | NR |
| Feng 2025 | Control; Mobile-App | NRS | RMDQ | NR | SF-12 PCS | NR | NR | NR |
| Geraghty 2025 | Control; Web-App | NR | RMDQ | PCS | NR | PSEQ | NR | NR |
| Hsieh 2025 | Face-to-face care; Exergaming | NR | ODI | FABQ-PA | NR | NR | HADS-D | HADS-A |
| Massah 2025 | Face-to-face care; Exergaming | VAS | NR | TSK | NR | NR | NR | NR |
| Villatoro-Luque 2025 | Face-to-face care; Telemedicine | NR | NR | TSK | NR | NR | NR | NR |
| Mudd 2025 | Face-to-face care; Telemedicine | NRS | NR | NR | SF-12 PCS | NR | NR | NR |
| Tawfek 2025 | Control; Telemedicine; Mobile-App | VAS | ODI | TSK | SF-12 PCS | NR | NR | NR |
| Menekseoglu 2026 | Face-to-face care; VR | VAS | RMDQ | NR | EQ-5D-5L | NR | HADS-D | HADS-A |
| Moreno-Ligero 2026 | Face-to-face care; Mobile-App | NRS | NR | NR | SF-12 PCS | NR | HADS-D | HADS-A |
| Spiegel 2026 | Control; VR | NRS | NR | NR | NR | NR | NR | NR |
| Ahmed 2026 | Face-to-face care; Exergaming | NRS | ODI | NR | NR | NR | NR | NR |
| Alahmri 2026 | Face-to-face care; Telemedicine | NRS | ODI | PCS | SF-12 PCS | NR | NR | NR |
| Jeong 2026 | Face-to-face care; Biofeedback devices | NRS | K-ODI | PCS | NR | NR | NR | NR |
| Lechauve 2026 | Face-to-face care; Mobile-App | NRS | ODI | NR | NR | NR | NR | NR |
| Slatman 2026 | Face-to-face care; VR | VAS | ODI | FABQ | NR | PSEQ | NR | NR |
| Valenzuela-Pascual 2026 | Control; Web-App | NRS | RMQ | FABQ | NR | NR | NR | NR |

**Note:** BDI, Beck Depression Inventory; BPI, Brief Pain Inventory; CES-D, Center for Epidemiologic Studies Depression Scale; CPSS, Chronic Pain Self-Efficacy Scale; DASS-A, Depression Anxiety Stress Scales–Anxiety subscale; DASS-D, Depression Anxiety Stress Scales–Depression subscale; DVPRS, Defense and Veterans Pain Rating Scale; EQ-5D, EuroQol 5-Dimension questionnaire; EQ-5D-3L, EuroQol 5-Dimension 3-Level questionnaire; EQ-5D-5L, EuroQol 5-Dimension 5-Level questionnaire; ERS, Exercise Regularly Scale; FABQ, Fear-Avoidance Beliefs Questionnaire; FABQ-PA, physical activity subscale of the Fear-Avoidance Beliefs Questionnaire; GAD-7, Generalized Anxiety Disorder 7-item scale; GSES, General Self-Efficacy Scale; HADS-A, Hospital Anxiety and Depression Scale–Anxiety subscale; HADS-D, Hospital Anxiety and Depression Scale–Depression subscale; K-ODI, Korean version of the Oswestry

Disability Index; mODI, modified Oswestry Disability Index; NPRS, Numeric Pain Rating Scale; NR, not reported; NRS, Numeric Rating Scale; ODI, Oswestry Disability Index; PAQ, Pain Assessment Questionnaire; PASES, Physical Activity Self-Efficacy Scale; PCS, Pain Catastrophizing Scale; PHQ-9, Patient Health Questionnaire-9; PROMIS, Patient-Reported Outcomes Measurement Information System; PSEQ, Pain Self-Efficacy Questionnaire; PSES, Pain Self-Efficacy Scale; QBPDS, Quebec Back Pain Disability Scale; QVAS, Quadruple Visual Analogue Scale; RMDQ, Roland–Morris Disability Questionnaire; RMQ, Roland–Morris Questionnaire; SF-12 PCS, Physical Component Summary of the 12-Item Short Form Health Survey; SF-36, 36-Item Short Form Health Survey; SF-36 PCS, Physical Component Summary of the 36-Item Short Form Health Survey; TSK, Tampa Scale for Kinesiophobia; VAS, Visual Analogue Scale; VR-12 PCS, Physical Component Summary of the Veterans RAND 12-Item Health Survey.

**Table S7.** Outcome-measure selection hierarchy

| Outcome domain | Priority | Abbreviation | Full name | Additional rules |
|---|---|---|---|---|
| **Pain intensity** | 1 | NRS | Numerical Rating Scale | Average or usual pain was preferred over worst pain or other pain summaries when multiple ratings were reported |
| | 2 | VAS/QVAS | Visual Analogue Scale | Conventional VAS was preferred; equivalent VAS-based variants were accepted |
| | 3 | BPI | Brief Pain Inventory—Pain Severity/Intensity | The average pain item was preferred when individual BPI pain-intensity items were available |
| | 4 | DVPRS | Defense and Veterans Pain Rating Scale | |
| | 5 | Other | Other validated pain-intensity measures | Included PAQ, ODI pain rating, or other study-specific validated pain-intensity measures when none of the above were available |
| **Physical function** | 1 | ODI/mODI | Oswestry Disability Index/Modified Oswestry Disability Index | ODI and mODI were considered equivalent for the purpose of the selection hierarchy |
| | 2 | RMDQ | Roland–Morris Disability Questionnaire | |
| | 3 | QBPDS | Quebec Back Pain Disability Scale | |
| | 4 | Other | Other validated measures of low-back-pain-related disability | |
| **Pain-related fear avoidance** | 1 | FABQ/FABQ-PA | Fear-Avoidance Beliefs Questionnaire/Fear-Avoidance Beliefs Questionnaire—Physical Activity subscale | The physical activity subscale was preferred over the total score or other subscales when more than one FABQ score was reported |

| | | | | |
|---|---|---|---|---|
| | 2 | TSK | Tampa Scale for Kinesiophobia | Used as a measure of pain-related fear of movement when FABQ was unavailable |
| | 3 | PCS | Pain Catastrophizing Scale | Included only when no fear-specific measure was available; catastrophizing was considered a related but conceptually distinct construct |
| | 4 | Other | Other validated measures of pain-related fear or avoidance | |
| **Health-related quality of life** | 1 | EQ-5D | EuroQol Five-Dimension Questionnaire | EQ-5D-3L and EQ-5D-5L were included; the utility index was preferred over the EQ VAS/health-state score |
| | 2 | SF-12 PCS/SF-36 PCS | 12-Item/36-Item Short Form Health Survey—Physical Component Summary | PCS was preferred over MCS or individual SF-12/SF-36 domain scores |
| | 3 | VR-12 PCS | Veterans RAND 12-Item Health Survey—Physical Component Summary | |
| | 4 | SF-36 | 36-Item Short Form Health Survey | A study-reported summary score was used when a PCS score was not separately available |
| | 5 | EQ VAS | EuroQol Visual Analogue Scale | Used only when an EQ-5D utility index was unavailable |
| | 6 | Other | Other validated measures of health-related quality of life | |

**Section 7.** Risk of bias

Risk of bias was assessed using the Cochrane Risk of Bias 2 (RoB 2) tool, targeting the effect of assignment to intervention. Individually randomised trials were assessed across five domains: bias arising from the randomisation process, bias due to deviations from intended interventions, bias due to missing outcome data, bias in measurement of the outcome, and bias in selection of the reported result. Cluster-randomised trials were assessed using the cluster-specific RoB 2 tool, which includes an additional domain addressing bias arising from the timing of identification or recruitment of participants in a cluster-randomised trial. Overall risk of bias was classified as low risk, some concerns, or high risk according to the RoB 2 algorithm. Assessments were completed using the RoB 2 Excel tools.[102 103]

**Table S8.** Risk-of-bias judgments

| Study ID | Randomization process | Deviations from intended interventions | Missing outcome data | Measurement of the outcome | Selection of the reported result | Overall |
|---|---|---|---|---|---|---|
| Iles 2011 | Low | Low | Some concerns | Some concerns | Some concerns | Some concerns |
| Carpenter 2012 | Some concerns | High | High | Some concerns | High | High |
| Del Pozo-Cruz 2012 | Low | Low | High | Some concerns | Some concerns | High |
| Krein 2013 | Low | Low | Low | Some concerns | Low | Some concerns |
| McDonough 2013 | Low | Low | Some concerns | Some concerns | Low | Some concerns |
| Park 2013 | Some concerns | Low | Low | Some concerns | Some concerns | Some concerns |
| Kim 2014 | Low | Low | Low | Some concerns | Some concerns | Some concerns |
| Irvine 2015 | Some concerns | Low | Low | Some concerns | Low | Some concerns |
| Geraghty 2018 | Low | Low | Low | Some concerns | Low | Some concerns |
| Schaller 2016 | Low | Some concerns | Some concerns | Some concerns | Low | Some concerns |
| Tan 2015 | Some concerns | High | High | Some concerns | Some concerns | High |
| Yilmaz Yelvar 2017 | Some concerns | Some concerns | Low | Some concerns | Some concerns | Some concerns |
| Chhabra 2018 | Some concerns | Low | Low | Some concerns | Some concerns | Some concerns |
| Goode 2018 | Low | Low | Some concerns | Some concerns | Low | Some concerns |
| Williams 2018 | Low | Low | Low | Some concerns | Low | Some concerns |
| Amorim 2019 | Low | Low | Some concerns | Some concerns | Low | Some concerns |

| | | | | | | |
|---|---|---|---|---|---|---|
| Mbada 2019 | Low | High | Some concerns | Some concerns | Some concerns | High |
| Petrozzi 2019 | Low | Low | Low | Some concerns | Low | Some concerns |
| Shebib 2019 | Low | High | High | Some concerns | Low | High |
| Toelle 2019 | Low | Some concerns | Some concerns | Some concerns | Low | Some concerns |
| Yang 2019 | Some concerns | High | Low | Some concerns | High | Some concerns |
| Zadro 2019 | Low | Some concerns | Low | Some concerns | High | High |
| Almhdawi 2020 | Some concerns | Some concerns | Low | Some concerns | High | High |
| Matheve 2020 | Low | Low | Low | Some concerns | Some concerns | Some concerns |
| Nambi 2020 | Low | Low | Low | Some concerns | Some concerns | Some concerns |
| Tomruk 2020 | Low | Low | Low | Some concerns | Some concerns | Some concerns |
| Alzahrani 2021 | Low | High | High | Some concerns | Low | High |
| Dadarkhah 2021 | Low | Low | Low | Some concerns | Some concerns | Some concerns |
| Garcia 2021 | Low | Low | Low | Some concerns | Low | Some concerns |
| Kazemi 2021 | Low | Some concerns | Some concerns | Some concerns | Low | Some concerns |
| Kim 2021 | Low | High | High | Some concerns | Some concerns | High |
| Lang 2021 | Low | High | High | Some concerns | Low | High |
| Li 2021 | Some concerns | Low | Low | Some concerns | Some concerns | Some concerns |
| Nambi 2021a | Low | Low | Low | Some concerns | Some concerns | Some concerns |
| Nambi 2021b | Low | Low | Low | Some concerns | Some concerns | Some concerns |
| Nambi 2021c | Low | Low | Low | Some concerns | Some concerns | Some concerns |
| Sandal 2021 | Low | Low | Some concerns | Some concerns | Low | Some concerns |
| Sato 2021 | Some concerns | Low | Low | Some concerns | Some concerns | Some concerns |
| Afzal 2022 | Some concerns | High | High | Some concerns | Some concerns | High |
| Eccleston 2022 | Low | Low | Low | Low | High | High |
| Itoh 2022 | Some concerns | Low | Some concerns | Some concerns | Some concerns | Some concerns |
| Lara-Palomo 2022 | Low | Low | Low | Some concerns | Some concerns | Some concerns |

| | | | | | | |
|---|---|---|---|---|---|---|
| Meinke 2022 | Low | Low | Low | Some concerns | Low | Some concerns |
| Nambi 2022 | Low | Low | Low | Some concerns | Some concerns | Some concerns |
| Yalfani 2022 | Low | High | Some concerns | Some concerns | Some concerns | High |
| Fatoye 2022 | Low | Some concerns | High | Some concerns | Some concerns | High |
| Cui 2023 | Low | Low | Some concerns | Some concerns | Some concerns | Some concerns |
| Garreta-Catala 2023 | Low | High | Some concerns | Some concerns | Low | High |
| Groenveld 2023 | Low | Low | Some concerns | Some concerns | Some concerns | Some concerns |
| Kałużna 2023 | Some concerns | Some concerns | Low | Some concerns | High | High |
| Kent 2023 | Low | Low | Low | Some concerns | Low | Some concerns |
| Lazaridou 2023 | Some concerns | High | Some concerns | Some concerns | Some concerns | High |
| Maddox 2023 | Low | Low | Low | Low | Low | Low |
| Park 2023 | Some concerns | Low | Low | Some concerns | High | High |
| Porwal 2023 | Low | High | Some concerns | Some concerns | Some concerns | High |
| Sanabria-Mazo 2023 | Low | Some concerns | High | Some concerns | Low | High |
| Fanuscu 2024 | Low | Low | Some concerns | Some concerns | Low | Some concerns |
| Kang 2024 | Low | High | Some concerns | Some concerns | Some concerns | High |
| McConnell 2024 | Low | Low | High | Some concerns | Some concerns | High |
| Öztürk 2024 | Low | High | High | Some concerns | Some concerns | High |
| Santos 2024 | Low | Some concerns | Low | Some concerns | Low | Some concerns |
| Shi 2024 | Low | Low | Some concerns | Some concerns | Low | Some concerns |
| Yang 2024 | Low | Low | High | Some concerns | Low | High |
| Yeldan 2024 | Low | Some concerns | Low | Some concerns | Low | Some concerns |
| Choi 2025 | Some concerns | Some concerns | Low | Some concerns | Low | Some concerns |
| Feng 2025 | Low | Low | Some concerns | Some concerns | Low | Some concerns |
| Geraghty 2025 | Low | Low | Low | Some concerns | Low | Some concerns |
| Hsieh 2025 | Low | Low | Some concerns | Some concerns | Low | Some concerns |

| | | | | | | |
|---|---|---|---|---|---|---|
| Massah 2025 | Low | Low | Low | Some concerns | Some concerns | Some concerns |
| Villatoro-Luque 2025 | Low | Some concerns | Low | Some concerns | Some concerns | Some concerns |
| Mudd 2025 | Low | Low | Low | Some concerns | Low | Some concerns |
| Tawfek 2025 | Low | Low | Low | Some concerns | Some concerns | Some concerns |
| Menekseoglu 2026 | Low | Some concerns | Low | Some concerns | Low | Some concerns |
| Moreno-Ligero 2026 | Low | Low | High | Some concerns | Some concerns | High |
| Spiegel 2026 | Low | Low | Low | Low | Low | Low |
| Ahmed 2026 | Low | Low | Low | Some concerns | Low | Low |
| Alahmri 2026 | Low | Low | Low | Some concerns | Low | Low |
| Jeong 2026 | Low | Some concerns | Low | Some concerns | Low | Some concerns |
| Lechauve 2026 | Low | Low | Some concerns | Some concerns | Low | Some concerns |
| Valenzuela-Pascual 2026 | Low | Low | Low | Some concerns | Low | Low |

**Table S9.** Risk-of-bias judgments for cluster-randomised trials

| Study ID | Randomization process | Timing of identification or recruitment of participants | Deviations from intended interventions | Missing outcome data | Measurement of the outcome | Selection of the reported result | Overall |
|---|---|---|---|---|---|---|---|
| Koppenaal 2022 | Low | Some concerns | Low | Low | Some concerns | Some concerns | Some concerns |
| Priebe 2024 | Low | Some concerns | Some concerns | Some concerns | Some concerns | Some concerns | Some concerns |
| Holden 2024 | Low | Some concerns | Some concerns | Some concerns | Some concerns | Some concerns | Some concerns |
| Slatman 2026 | Some concerns | Some concerns | Low | Low | Some concerns | Low | Some concerns |

**Section 8.** Assessment of transitivity across outcome networks

Transitivity was assessed at the outcome level across all available follow-up intervals. For each outcome, a direct comparison was included if at least one trial contributed usable data to that comparison at any available follow-up interval. Each trial was counted once within a given direct comparison, irrespective of the number of follow-up intervals to which it contributed.

**Table S10.** Pain intensity

| Comparisons | n studies | Intervention duration (mode) | Mean age (years) | Female (%) | Outcome measures |
|---|---|---|---|---|---|
| Control: Face-to-face care | 3 | 1 day / 8 weeks / 12 weeks | <50 | ≥50% | VAS; NRS |
| Control: Telemedicine | 9 | 8 weeks | <50 / ≥50 | <50% / ≥50% | NRS; BPI intensity; ODI Pain rating; VAS |
| Control: Web-App | 6 | 6 weeks / 8 weeks | <50 / ≥50 | <50% / ≥50% | PAQ; NRS; VAS; NPRS |
| Control: Mobile-App | 7 | 12 weeks | <50 | <50% / ≥50% | NRS; VAS |
| Control: VR | 9 | 8 weeks | <50 / ≥50 | <50% / ≥50% | VAS; DVPRS; NRS; BPI intensity |
| Control: Exergaming | 4 | 8 weeks | <50 / ≥50 | <50% / ≥50% | VAS; NRS |
| Control: Biofeedback devices | 1 | 12 weeks | <50 | ≥50% | NRS |
| Face-to-face care: Telemedicine | 5 | 8 weeks | <50 / ≥50 | <50% / ≥50% | NRS; BPI intensity; VAS |
| Face-to-face care: Web-App | 4 | 8 weeks | <50 / ≥50 | ≥50% | Other scale; NRS; VAS |
| Face-to-face care: Mobile-App | 10 | 8 weeks | <50 | <50% / ≥50% | NRS; QVAS; VAS |
| Face-to-face care: VR | 7 | 4 weeks | <50 | <50% / ≥50% | VAS; NRS |
| Face-to-face care: Exergaming | 4 | 1 day / 3 weeks / 4 weeks / 8 weeks | <50 | <50% / ≥50% | VAS; NRS |
| Face-to-face care: Biofeedback devices | 10 | 4 weeks / 12 weeks | <50 / ≥50 | <50% / ≥50% | BPI intensity; NRS; VAS |
| Telemedicine: Mobile-App | 1 | 12 weeks | <50 | <50% | VAS |
| Telemedicine: Biofeedback devices | 1 | 8 weeks | ≥50 | NR | BPI intensity |

**Note:** Intervention duration represents the most frequently reported duration within each comparison; where multiple durations occurred with equal frequency, all are shown. Mean age was categorised according to whether the study-level mean was <50 or ≥50 years, and female proportion according to whether <50% or ≥50% of participants were female. BPI, Brief Pain Inventory; DVPRS, Defense and Veterans Pain Rating Scale; NPRS, Numeric Pain Rating Scale; NR, not reported; NRS, Numeric Rating Scale; ODI, Oswestry Disability Index; PAQ, Pain Assessment Questionnaire; QVAS, Quadruple Visual Analogue Scale; VAS, Visual Analogue Scale; VR, virtual reality; Mobile-App, Mobile-Applications; Web-App, Web-Applications.

**Table S11.** Physical function

| Comparisons | n studies | Intervention duration (mode) | Mean age (years) | Female (%) | Outcome measures |
|---|---|---|---|---|---|
| Control: Face-to-face care | 2 | 1 day / 12 weeks | <50 | ≥50% | QBPDS; RMDQ |
| Control: Telemedicine | 10 | 8 weeks | <50 / ≥50 | <50% / ≥50% | ODI; RMDQ |
| Control: Web-App | 8 | 6 weeks | <50 / ≥50 | <50% / ≥50% | RMDQ; QBPDS; ODI; RMQ |
| Control: Mobile-App | 5 | 12 weeks | <50 | <50% / ≥50% | ODI; RMDQ |
| Control: VR | 3 | 8 weeks | ≥50 | ≥50% | ODI; mODI |
| Control: Exergaming | 2 | 3 weeks / 8 weeks | <50 / ≥50 | ≥50% | RMDQ |
| Control: Biofeedback devices | 1 | 12 weeks | <50 | ≥50% | RMDQ |
| Face-to-face care: Telemedicine | 3 | 2 weeks / 4 weeks / 8 weeks | <50 | ≥50% | ODI |
| Face-to-face care: Web-App | 3 | 8 weeks | <50 / ≥50 | ≥50% | RMDQ; QBPDS; ODI |
| Face-to-face care: Mobile-App | 9 | 8 weeks | <50 | <50% / ≥50% | mODI; RMDQ; ODI |
| Face-to-face care: VR | 5 | 2 weeks / 4 weeks | <50 | ≥50% | ODI; mODI; RMDQ |
| Face-to-face care: Exergaming | 4 | 2 weeks / 3 weeks / 4 weeks / 8 weeks | <50 / ≥50 | ≥50% | ODI |
| Face-to-face care: Biofeedback devices | 9 | 4 weeks / 12 weeks | <50 / ≥50 | <50% / ≥50% | ODI; RMDQ; K-ODI |
| Telemedicine: Mobile-App | 1 | 12 weeks | <50 | <50% | ODI |

**Note:** Intervention duration represents the most frequently reported duration within each comparison; where multiple durations occurred with equal frequency, all are shown. Mean age was categorised according to whether the study-level mean was <50 or ≥50 years, and female proportion according to whether <50% or ≥50% of participants were female. K-ODI, Korean version of the Oswestry Disability Index; mODI, modified Oswestry Disability Index; ODI, Oswestry Disability Index; QBPDS, Quebec Back Pain Disability Scale; RMDQ, Roland–Morris Disability Questionnaire; RMQ, Roland–Morris Questionnaire; VR, virtual reality; Mobile-App, Mobile-Applications; Web-App, Web-Applications.

**Table S12.** Pain-related fear avoidance

| Comparisons | n studies | Intervention duration (mode) | Mean age (years) | Female (%) | Outcome measures |
|---|---|---|---|---|---|
| Control: Face-to-face care | 1 | 12 weeks | <50 | ≥50% | FABQ-PA |
| Control: Telemedicine | 5 | 8 weeks / 12 weeks | <50 / ≥50 | <50% / ≥50% | FABQ-PA; PCS; TSK |
| Control: Web-App | 4 | 8 weeks | <50 / ≥50 | <50% / ≥50% | FABQ-PA; TSK-17; PCS; FABQ |
| Control: Mobile-App | 3 | 12 weeks | <50 | <50% / ≥50% | TSK; FABQ-PA |
| Control: VR | 4 | 4 weeks | <50 / ≥50 | <50% / ≥50% | TSK; PCS |
| Control: Exergaming | 3 | 8 weeks | <50 / ≥50 | <50% / ≥50% | TSK |
| Control: Biofeedback devices | 1 | 12 weeks | <50 | ≥50% | FABQ-PA |
| Face-to-face care: Telemedicine | 3 | 8 weeks | <50 | ≥50% | FABQ-PA; TSK; PCS |
| Face-to-face care: Web-App | 2 | 8 weeks | <50 / ≥50 | ≥50% | PCS |
| Face-to-face care: Mobile-App | 3 | 8 weeks | <50 | <50% / ≥50% | FABQ |
| Face-to-face care: VR | 2 | 2 weeks / 4 weeks | <50 | ≥50% | TSK; FABQ |
| Face-to-face care: Exergaming | 3 | 1 day / 2 weeks / 4 weeks | <50 / ≥50 | <50% / ≥50% | FABQ-PA; TSK; FABQ |
| Face-to-face care: Biofeedback devices | 5 | 4 weeks / 12 weeks | <50 | <50% / ≥50% | TSK; FABQ-PA; PCS-13 |
| Telemedicine: Mobile-App | 1 | 12 weeks | <50 | <50% | TSK |

**Note:** Intervention duration is presented as the mode; where multiple durations occurred with equal frequency, all modes are shown. Mean age was categorised according to whether the study-level mean was <50 or ≥50 years, and female proportion according to whether <50% or ≥50% of participants were female. FABQ, Fear-Avoidance Beliefs Questionnaire; FABQ-PA, physical activity subscale of the Fear-Avoidance Beliefs Questionnaire; PCS, Pain Catastrophizing Scale; PCS-13, 13-item Pain Catastrophizing Scale; TSK, Tampa Scale for Kinesiophobia; VR, virtual reality; Mobile-App, Mobile-Applications; Web-App, Web-Applications.

**Table S13.** Health-related quality of life

| Comparisons | n studies | Intervention duration (mode) | Mean age (years) | Female (%) | Outcome measures |
|---|---|---|---|---|---|
| Control: Telemedicine | 5 | 12 weeks | <50 / ≥50 | <50% / ≥50% | SF-12 PCS; EQ-5D-5L; SF-36 |
| Control: Web-App | 1 | 8 weeks | <50 | ≥50% | SF-36 |
| Control: Mobile-App | 4 | 12 weeks | <50 | <50% / ≥50% | SF-12 PCS; EQ-5D-5L |
| Control: VR | 4 | 4 weeks / 8 weeks | <50 / ≥50 | <50% / ≥50% | PCS fitness index; EQ-5DHealth state; SF-36; SF-12 PCS |
| Face-to-face care: Telemedicine | 3 | 2 weeks / 8 weeks / 26 weeks | <50 / ≥50 | <50% / ≥50% | EQ-5D; SF-12 PCS |
| Face-to-face care: Mobile-App | 8 | 8 weeks | <50 | <50% / ≥50% | SF-12 PCS; VR-12 PCS; SF-36; EQ-5DHealth state; EQ-5D-3L |
| Face-to-face care: VR | 1 | 3 weeks | <50 | ≥50% | EQ-5D-5L |
| Face-to-face care: Biofeedback devices | 3 | 4 weeks | <50 | ≥50% | SF-36 PCS; SF-12 PCS; SF-36 |
| Telemedicine: Mobile-App | 1 | 12 weeks | <50 | <50% | SF-12 PCS |

**Note:** Intervention duration is presented as the mode; where multiple durations occurred with equal frequency, all modes are shown. Mean age was categorised according to whether the study-level mean was <50 or ≥50 years, and female proportion according to whether <50% or ≥50% of participants were female. EQ-5D, EuroQol 5-Dimension questionnaire; EQ-5D-3L, EuroQol 5-Dimension 3-Level questionnaire; EQ-5D-5L, EuroQol 5-Dimension 5-Level questionnaire; PCS, Physical Component Summary; SF-12 PCS, Physical Component Summary of the 12-Item Short Form Health Survey; SF-36, 36-Item Short Form Health Survey; SF-36 PCS, Physical Component Summary of the 36-Item Short Form Health Survey; VR-12 PCS, Physical Component Summary of the Veterans RAND 12-Item Health Survey; VR, virtual reality; Mobile-App, Mobile-Applications; Web-App, Web-Applications.

**Table S14.** Self-efficacy

| Comparisons | n studies | Intervention duration (mode) | Mean age (years) | Female (%) | Outcome measures |
|---|---|---|---|---|---|
| Control: Face-to-face care | 1 | 12 weeks | <50 | ≥50% | PSEQ |
| Control: Telemedicine | 3 | 7 weeks / 8 weeks / 12 weeks | <50 / ≥50 | <50% / ≥50% | PSEQ; PASES; CPSS |
| Control: Web-App | 3 | 6 weeks | <50 / ≥50 | <50% / ≥50% | PSES; ERS; PSEQ |
| Control: Mobile-App | 2 | 8 weeks / 12 weeks | <50 | ≥50% | Other scale; PSEQ |
| Control: Exergaming | 2 | 8 weeks | ≥50 | <50% / ≥50% | PSEQ |
| Control: Biofeedback devices | 1 | 12 weeks | <50 | ≥50% | PSEQ |
| Face-to-face care: Telemedicine | 1 | 8 weeks | <50 | ≥50% | PASES |
| Face-to-face care: Web-App | 1 | 8 weeks | ≥50 | ≥50% | PSEQ |
| Face-to-face care: Mobile-App | 3 | 4 weeks / 6 weeks / 3 or 12 weeks | <50 | <50% / ≥50% | PSEQ; GSES |
| Face-to-face care: VR | 1 | 4 weeks | <50 | ≥50% | PSEQ |
| Face-to-face care: Biofeedback devices | 1 | 12 weeks | <50 | ≥50% | PSEQ |

**Note:** Intervention duration is presented as the mode; where multiple durations occurred with equal frequency, all modes are shown. Mean age was categorised according to whether the study-level mean was <50 or ≥50 years, and female proportion according to whether <50% or ≥50% of participants were female. CPSS, Chronic Pain Self-Efficacy Scale; ERS, Exercise Regularly Scale; GSES, General Self-Efficacy Scale; PASES, Physical Activity Self-Efficacy Scale; PSEQ, Pain Self-Efficacy Questionnaire; PSES, Pain Self-Efficacy Scale; VR, virtual reality; Mobile-App, Mobile-Applications; Web-App, Web-Applications.

**Table S15.** Anxiety

| Comparisons | n studies | Intervention duration (mode) | Mean age (years) | Female (%) | Outcome measures |
|---|---|---|---|---|---|
| Control: Telemedicine | 4 | 8 weeks | <50 / ≥50 | ≥50% | DASS-A; HADS-A |
| Control: Mobile-App | 1 | 6 weeks | <50 | ≥50% | DASS-A |
| Control: VR | 2 | 4 weeks / 8 weeks | ≥50 | ≥50% | HADS-A; PROMIS-Anxiety |
| Face-to-face care: Web-App | 1 | 8 weeks | ≥50 | ≥50% | DASS-A |
| Face-to-face care: VR | 1 | 3 weeks | <50 | ≥50% | HADS-A |
| Face-to-face care: Exergaming | 1 | 2 weeks | ≥50 | ≥50% | HADS-A |

**Note:** Intervention duration is presented as the mode; where multiple durations occurred with equal frequency, all modes are shown. Mean age was categorised according to whether the study-level mean was <50 or ≥50 years, and female proportion according to whether <50% or ≥50% of participants were female. DASS-A, Depression Anxiety Stress Scales–Anxiety subscale; GAD-7, Generalized Anxiety Disorder 7-item scale; HADS-A, Hospital Anxiety and Depression Scale–Anxiety subscale; PROMIS-Anxiety, Patient-Reported Outcomes Measurement Information System Anxiety; VR, virtual reality; Mobile-App, Mobile-Applications; Web-App, Web-Applications.

**Table S16.** Depression

| Comparisons | n studies | Intervention duration (mode) | Mean age (years) | Female (%) | Outcome measures |
|---|---|---|---|---|---|
| Control: Telemedicine | 5 | 8 weeks | <50 / ≥50 | ≥50% | DASS-D; HADS-D; CES-D |
| Control: Mobile-App | 1 | 6 weeks | <50 | ≥50% | DASS-D |
| Control: VR | 3 | 8 weeks | ≥50 | ≥50% | HADS-D; PROMIS-Depression; PROMIS Depression |
| Face-to-face care: Web-App | 1 | 8 weeks | ≥50 | ≥50% | DASS-D |
| Face-to-face care: Mobile-App | 2 | 8 weeks / NR | <50 | ≥50% | PHQ-9; HADS-D |
| Face-to-face care: VR | 1 | 3 weeks | <50 | ≥50% | HADS-D |
| Face-to-face care: Exergaming | 1 | 2 weeks | ≥50 | ≥50% | HADS-D |
| Face-to-face care: Biofeedback devices | 1 | 4 weeks | <50 | ≥50% | BDI |

**Note:** Intervention duration is presented as the mode; where multiple durations occurred with equal frequency, all modes are shown. Mean age was categorised according to whether the study-level mean was <50 or ≥50 years, and female proportion according to whether <50% or ≥50% of participants were female. BDI, Beck Depression Inventory; CES-D, Center for Epidemiologic Studies Depression Scale; DASS-D, Depression Anxiety Stress Scales–Depression subscale; HADS-D, Hospital Anxiety and Depression Scale–Depression subscale; PHQ-9, Patient Health Questionnaire-9; PROMIS-Depression, Patient-Reported Outcomes Measurement Information System Depression; VR, virtual reality; Mobile-App, Mobile-Applications; Web-App, Web-Applications.

**Section 9.** Network geometry

Each node represents an intervention category, and node size is proportional to the total number of participants assigned to that intervention. Each connecting line represents a direct comparison between two interventions, with line thickness proportional to the number of randomised controlled trials contributing direct evidence. The absence of a connecting line indicates that no direct comparison was available. VR, virtual reality; Mobile-App, mobile application-based intervention; Web-App, web-based intervention.

**Figure S1.** Network plots for pain-related fear avoidance

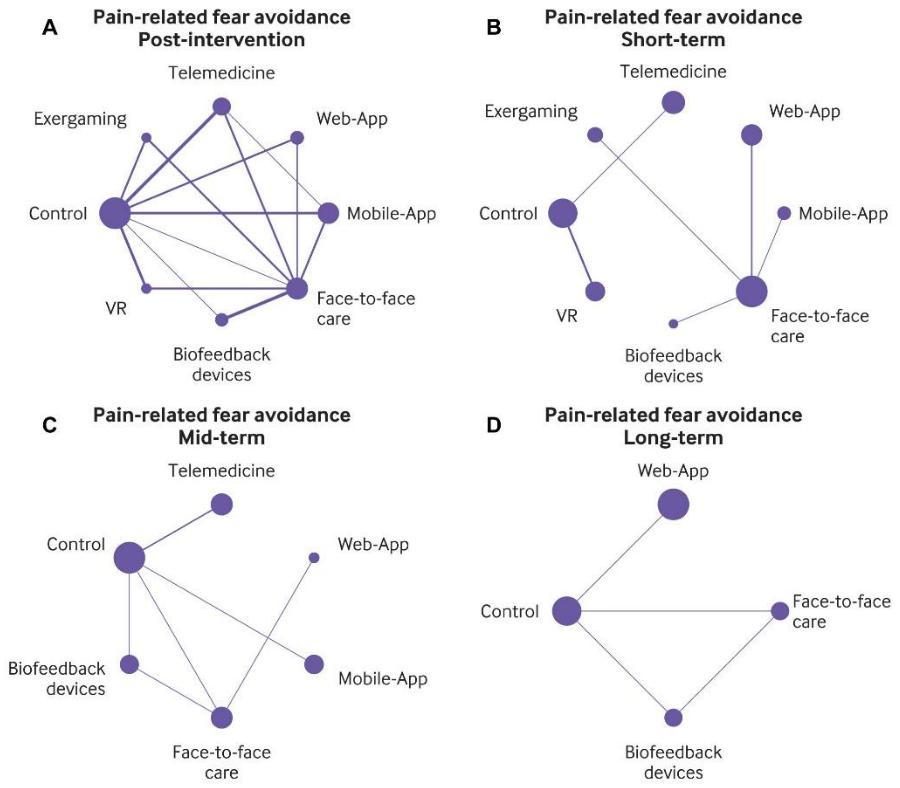

**Figure S2.** Network plots for health-related quality of life

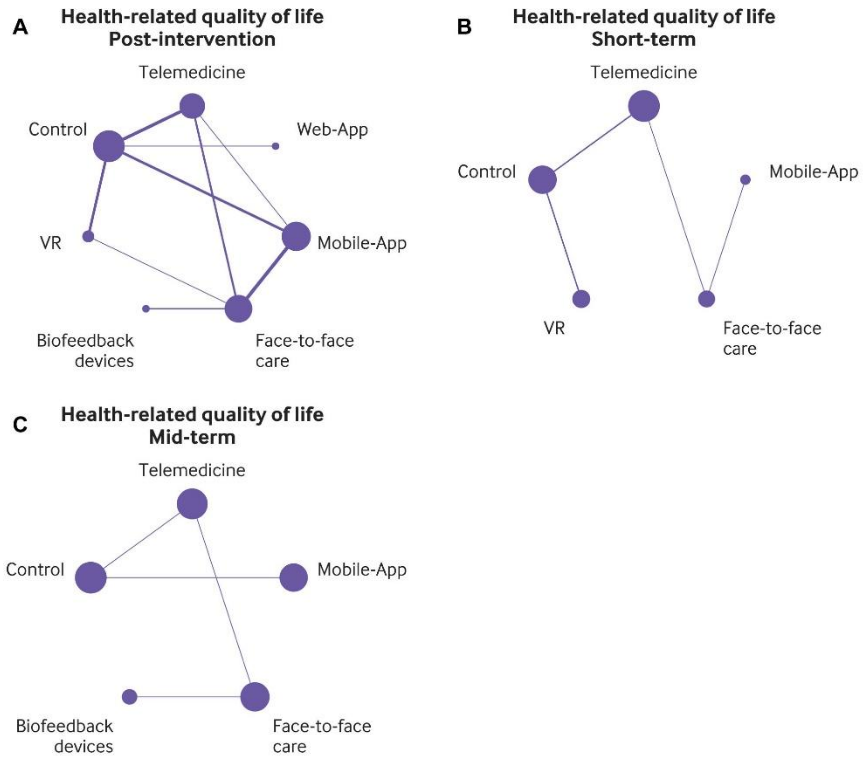

**Figure S3.** Network plots for self-efficacy

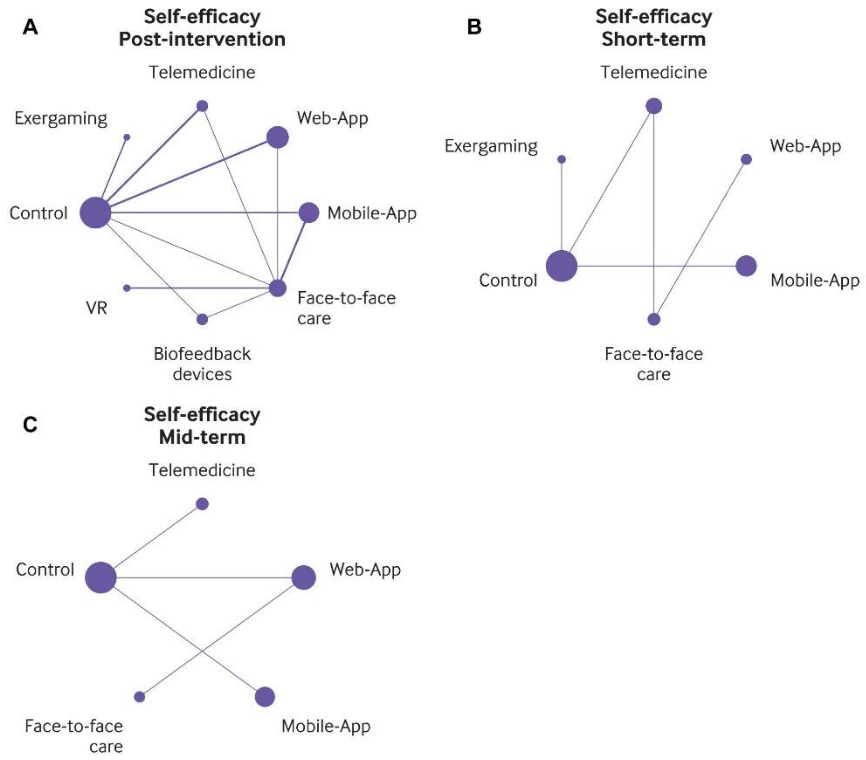

**Figure S4.** Network plots for anxiety

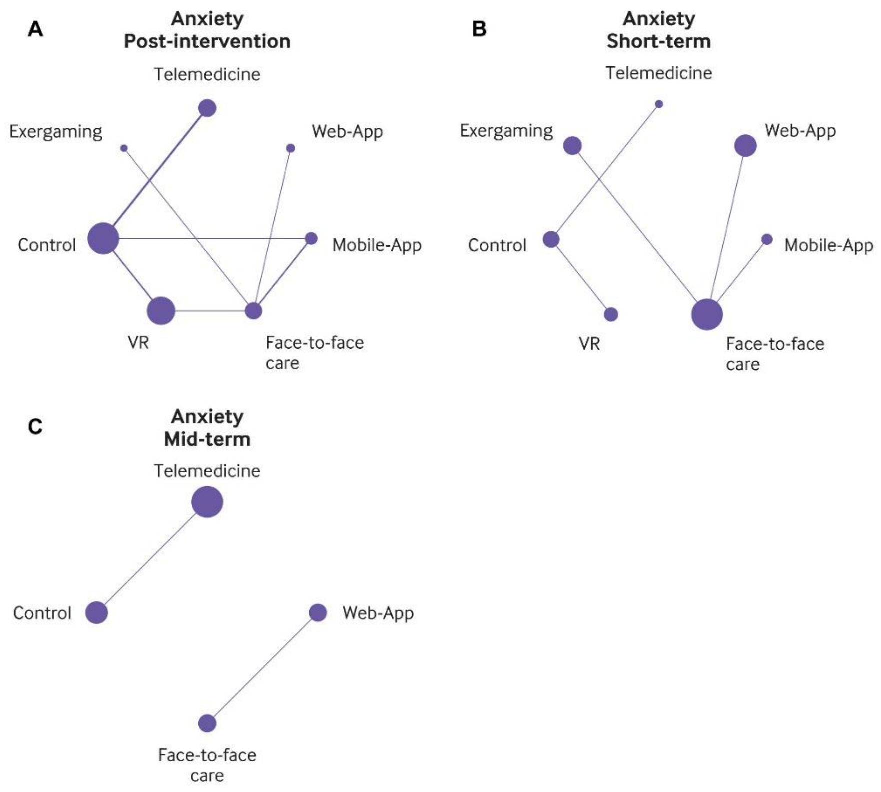

**Figure S5.** Network plots for depression

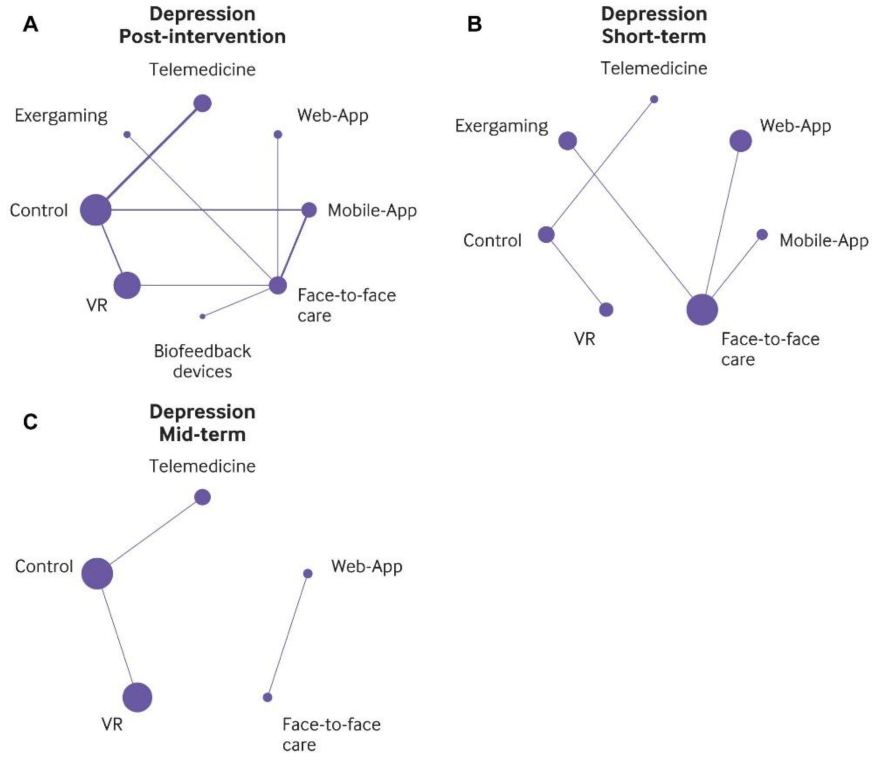

**Section 10.** Forest plots of network meta-analysis estimates

Network meta-analysis estimates are presented as standardised mean differences (SMDs) with 95% confidence intervals (CIs) using random-effects models. For pain intensity, physical function, pain-related fear avoidance, anxiety, and depression, negative SMD values indicate improvement; for health-related quality of life and self-efficacy, positive SMD values indicate improvement. Estimates with 95% CIs excluding zero indicate statistical significance. VR, virtual reality; Mobile-App, mobile application-based intervention; Web-App, web-based intervention.

**Figure S6.** Forest plot for pain intensity at post-intervention

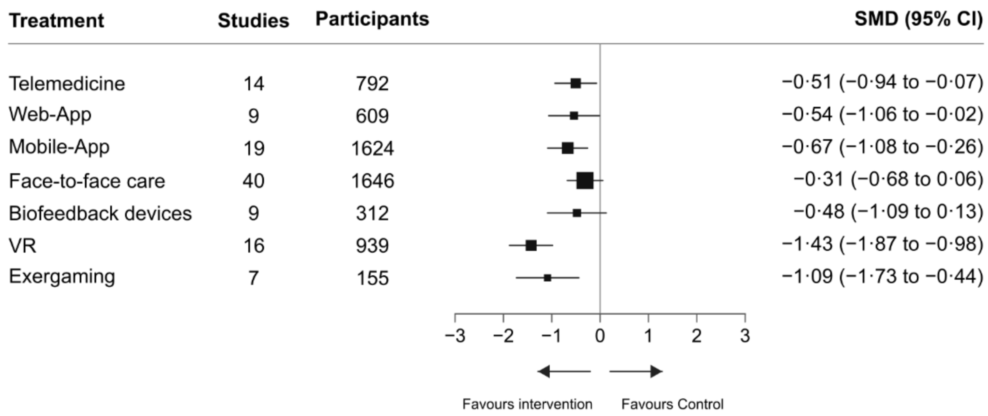

**Figure S7.** Forest plot for pain intensity at short-term

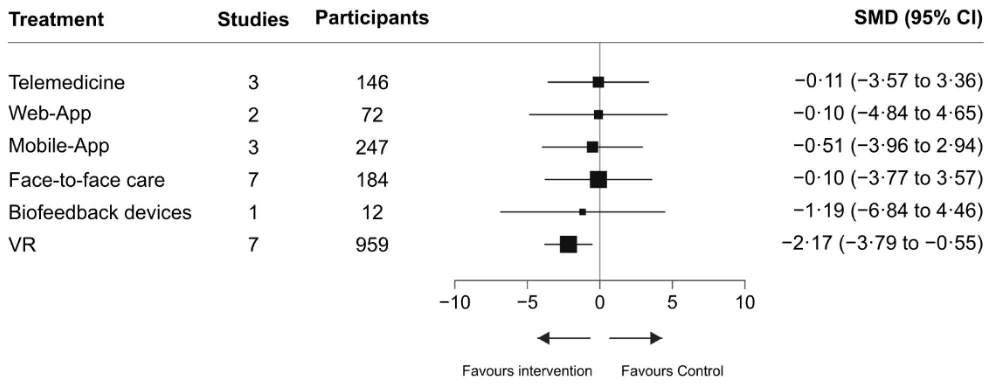

**Figure S8.** Forest plot for pain intensity at mid-term

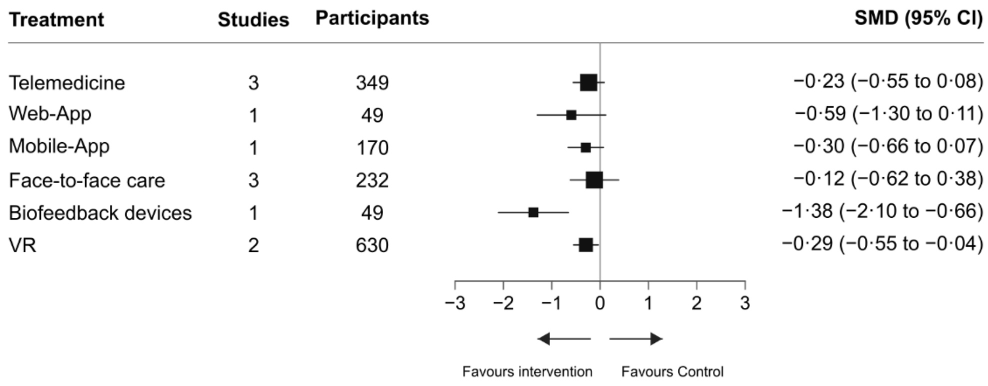

**Figure S9.** Forest plot for pain intensity at long-term

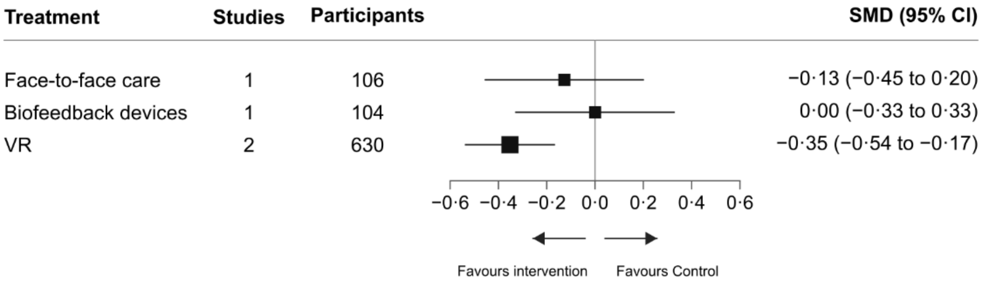

**Figure S10.** Forest plot for physical function at post-intervention

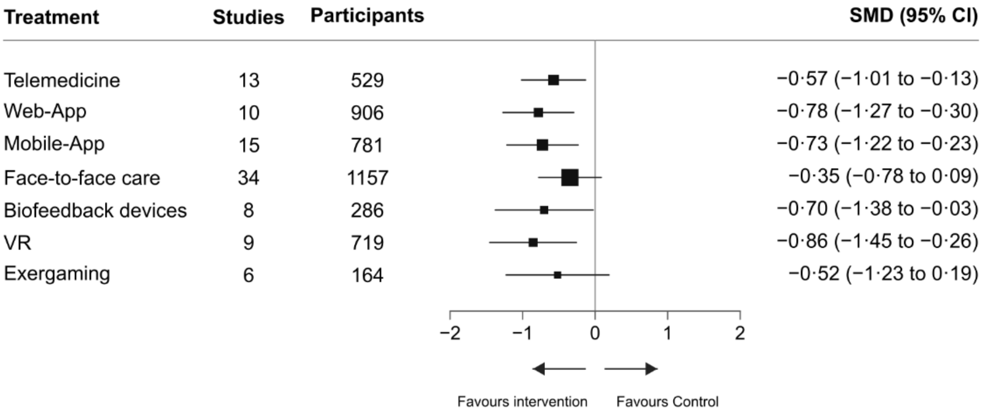

**Figure S11.** Forest plot for physical function at short-term

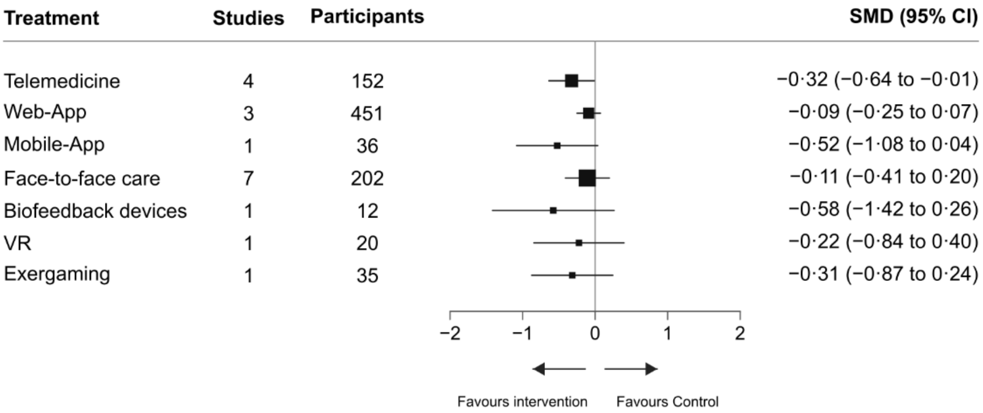

**Figure S12.** Forest plot for physical function at mid-term

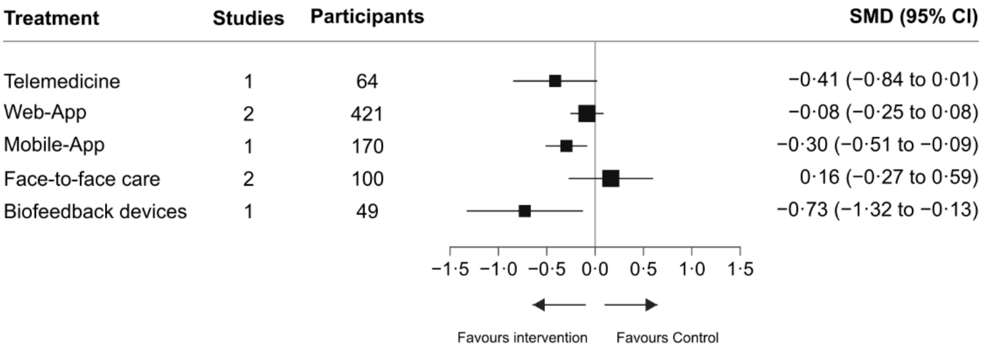

**Figure S13.** Forest plot for physical function at long-term

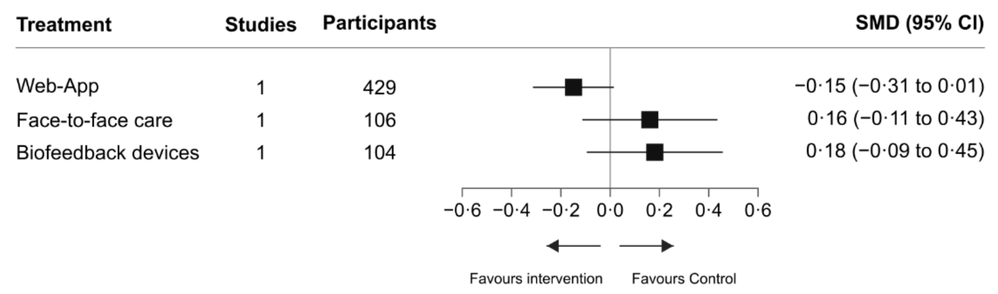

**Figure S14.** Forest plot for pain-related fear avoidance at post-intervention

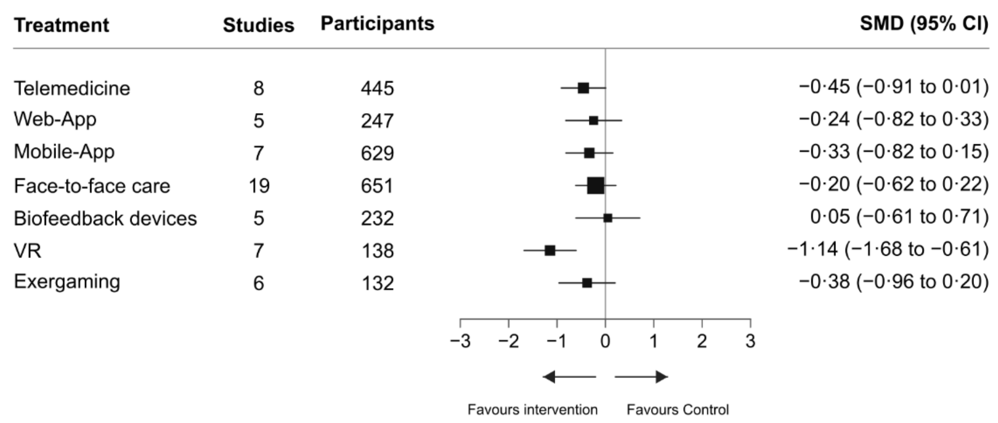

**Figure S15.** Forest plot for pain-related fear avoidance at mid-term

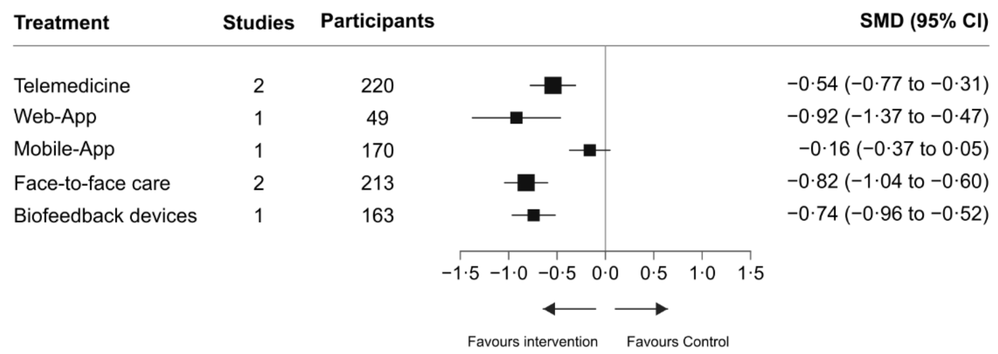

**Figure S16.** Forest plot for pain-related fear avoidance at long-term

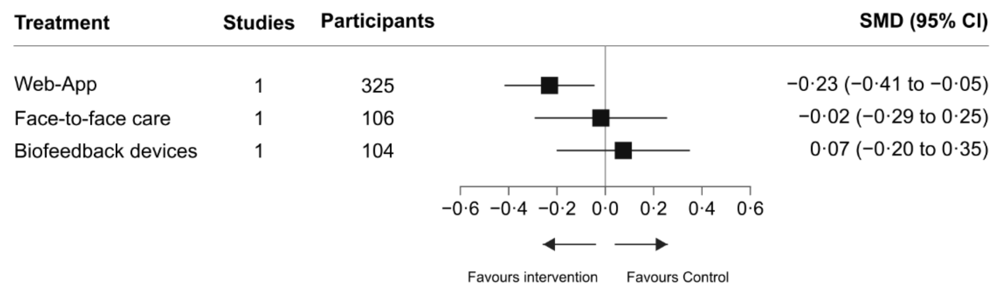

**Figure S17.** Forest plot for health-related quality of life at post-intervention

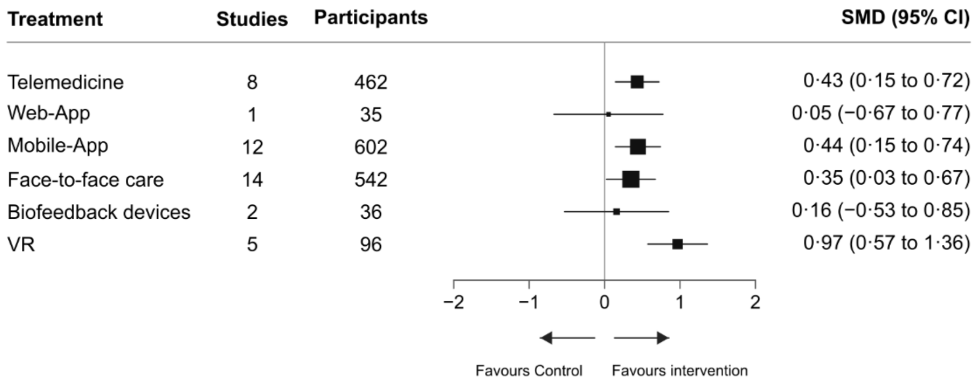

**Figure S18.** Forest plot for health-related quality of life at short-term

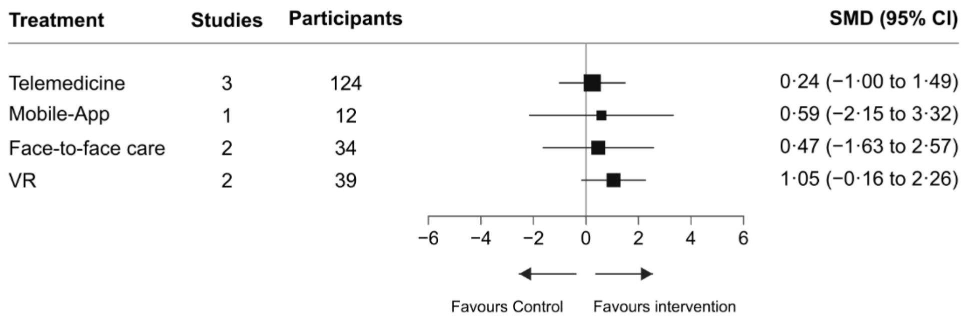

**Figure S19.** Forest plot for health-related quality of life at mid-term

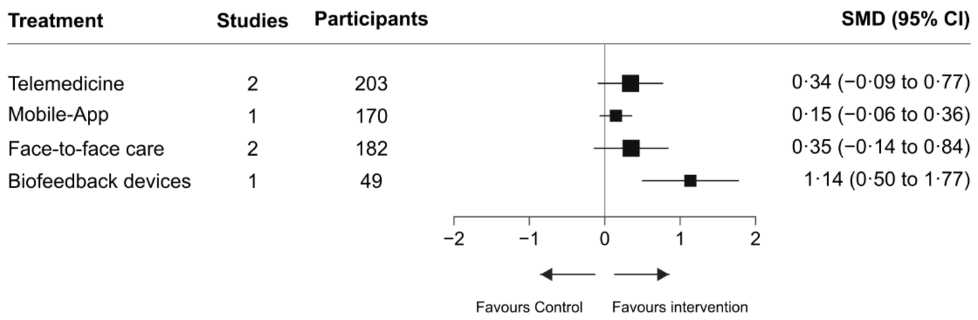

**Figure S20.** Forest plot for self-efficacy at post-intervention

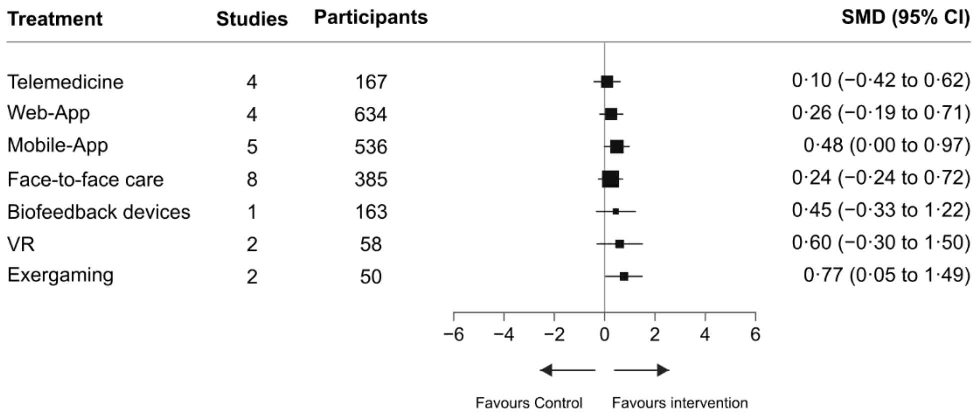

**Figure S21.** Forest plot for self-efficacy at short-term

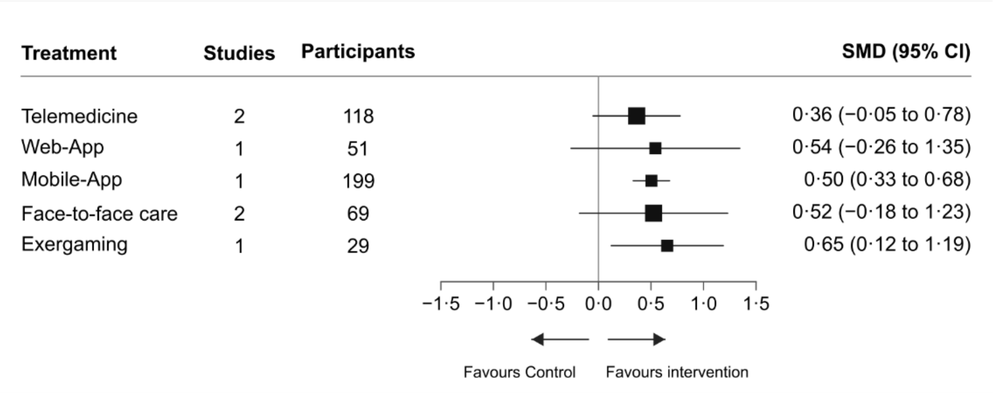

**Figure S22.** Forest plot for self-efficacy at mid-term

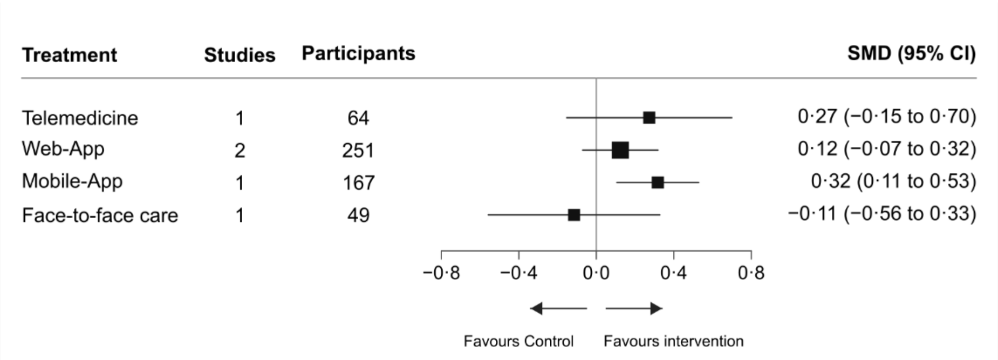

**Figure S23.** Forest plot for anxiety at post-intervention

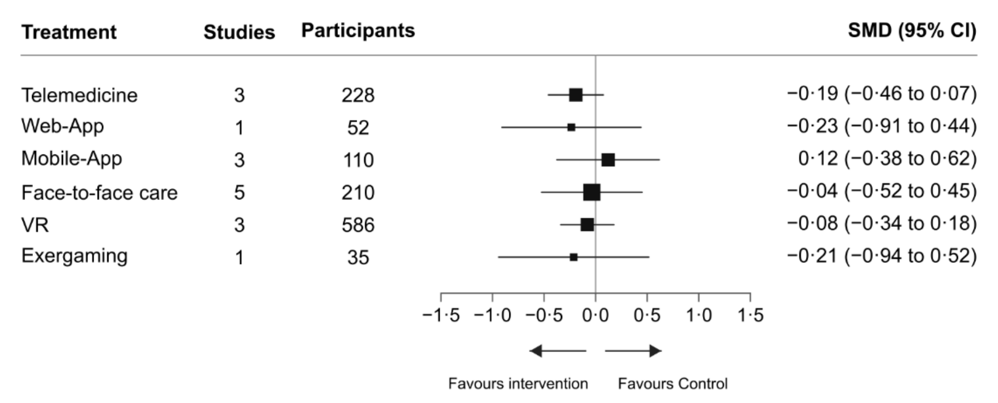

**Figure S24.** Forest plot for depression at post-intervention

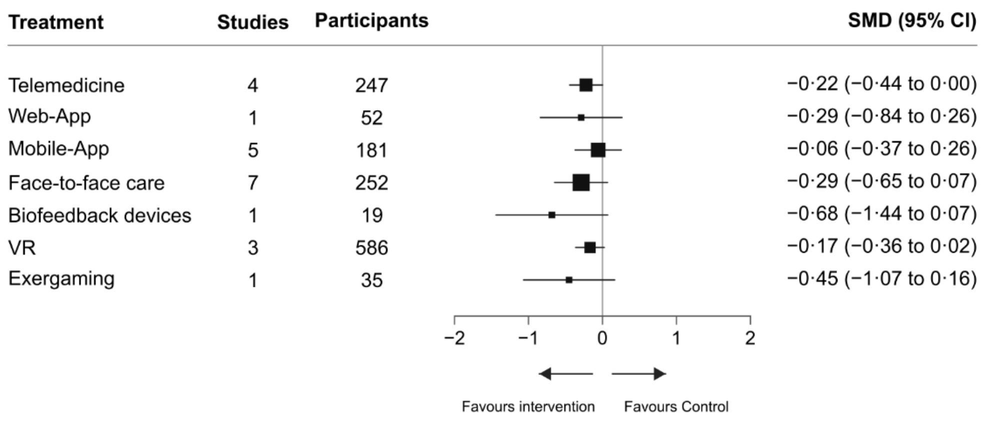

**Section 11.** League tables of network meta-analysis estimates

Results are presented as standardised mean differences (SMDs) with 95% confidence intervals (CIs). Network meta-analysis estimates are shown in the lower-left triangle and direct pairwise estimates in the upper-right triangle. For pain intensity, physical function, pain-related fear avoidance, anxiety, and depression, negative network estimates favour the column-defining intervention and negative direct estimates favour the row-defining intervention; the direction is reversed for health-related quality of life and self-efficacy. Estimates with 95% CIs excluding zero are shown in bold; a dot indicates no available direct estimate. VR, virtual reality; Mobile-App, mobile application-based intervention; Web-App, web-based intervention.

**Table S17.** Pain intensity at post-intervention

| | | | | | | | |
|---|---|---|---|---|---|---|---|
| **Telemedicine** | . | 0.22 (−1.36 to 1.80) | −0.16 (−0.86 to 0.54) | −0.84 (−2.43 to 0.74) | . | −0.48 (−1.00 to 0.04) | . |
| 0.03 (−0.61 to 0.68) | **Web-App** | . | −0.26 (−1.02 to 0.51) | . | . | −0.59 (−1.22 to 0.03) | . |
| 0.17 (−0.37 to 0.70) | 0.13 (−0.49 to 0.75) | **Mobile-App** | −0.34 (−0.81 to 0.14) | . | . | **−0.71 (−1.25 to −0.16)** | . |
| −0.19 (−0.66 to 0.27) | −0.23 (−0.78 to 0.32) | −0.36 (−0.75 to 0.03) | **Face-to-face care** | 0.19 (−0.35 to 0.72) | **0.89 ( 0.32 to 1.46)** | −0.67 (−1.59 to 0.24) | **1.38 ( 0.52 to 2.24)** |
| −0.02 (−0.69 to 0.64) | −0.06 (−0.80 to 0.68) | −0.19 (−0.83 to 0.45) | 0.17 (−0.35 to 0.69) | **Biofeedback devices** | . | −0.55 (−2.04 to 0.95) | . |
| **0.92 ( 0.35 to 1.50)** | **0.89 ( 0.24 to 1.54)** | **0.76 ( 0.22 to 1.30)** | **1.12 ( 0.67 to 1.56)** | **0.95 ( 0.28 to 1.62)** | **VR** | **−1.66 (−2.24 to −1.08)** | . |
| **−0.51 (−0.94 to −0.07)** | **−0.54 (−1.06 to −0.02)** | **−0.67 (−1.08 to −0.26)** | −0.31 (−0.68 to 0.06) | −0.48 (−1.09 to 0.13) | **−1.43 (−1.87 to −0.98)** | **Control** | 0.31 (−0.52 to 1.14) |
| 0.58 (−0.16 to 1.33) | 0.55 (−0.25 to 1.35) | 0.42 (−0.30 to 1.13) | **0.78 ( 0.13 to 1.43)** | 0.61 (−0.21 to 1.43) | −0.34 (−1.08 to 0.40) | **1.09 ( 0.44 to 1.73)** | **Exergaming** |

**Table S18.** Pain intensity at short-term

| | | | | | | |
|---|---|---|---|---|---|---|
| **Telemedicine** | . | . | 0.13 (−2.88 to 3.14) | . | . | −0.38 (−4.62 to 3.86) |
| −0.01 (−4.08 to 4.06) | **Web-App** | . | −0.00 (−3.00 to 3.00) | . | . | . |
| 0.41 (−3.06 to 3.87) | 0.41 (−3.66 to 4.48) | **Mobile-App** | −0.55 (−3.57 to 2.46) | . | . | −0.24 (−4.46 to 3.98) |
| −0.01 (−2.75 to 2.74) | 0.00 (−3.00 to 3.00) | −0.41 (−3.16 to 2.34) | **Face-to-face care** | 1.09 (−3.21 to 5.39) | . | . |
| 1.08 (−4.01 to 6.18) | 1.09 (−4.15 to 6.33) | 0.68 (−4.42 to 5.78) | 1.09 (−3.21 to 5.39) | **Biofeedback devices** | . | . |
| 2.07 (−1.76 to 5.89) | 2.07 (−2.94 to 7.09) | 1.66 (−2.16 to 5.47) | 2.07 (−1.94 to 6.09) | 0.98 (−4.90 to 6.86) | **VR** | **−2.17 (−3.79 to −0.55)** |
| −0.11 (−3.57 to 3.36) | −0.10 (−4.84 to 4.65) | −0.51 (−3.96 to 2.94) | −0.10 (−3.77 to 3.57) | −1.19 (−6.84 to 4.46) | **−2.17 (−3.79 to −0.55)** | **Control** |

**Table S19.** Pain intensity at mid-term

| | | | | | |
|---|---|---|---|---|---|
| **Telemedicine** | . | . | −0.12<br>(−0.50 to 0.27) | . | . |
| 0.36<br>(−0.27 to 0.99) | **Web-App** | . | −0.48<br>(−0.98 to 0.02) | . | . |
| 0.06<br>(−0.42 to 0.55) | −0.30<br>(−1.09 to 0.50) | **Mobile-App** | . | . | . |
| −0.12<br>(−0.50 to 0.27) | −0.48<br>(−0.98 to 0.02) | −0.18<br>(−0.80 to 0.44) | **Face-to-face care** | **1.26**<br>**( 0.74 to 1.79)** | . |
| **1.14**<br>**( 0.50 to 1.79)** | **0.79**<br>**( 0.06 to 1.51)** | **1.08**<br>**( 0.27 to 1.89)** | **1.26**<br>**( 0.74 to 1.79)** | **Biofeedback devices** | . |
| 0.06<br>(−0.35 to 0.47) | −0.30<br>(−1.05 to 0.45) | −0.00<br>(−0.45 to 0.44) | 0.18<br>(−0.38 to 0.74) | **−1.09**<br>**(−1.85 to −0.32)** | **VR** |

**Table S20.** Pain intensity at long-term

| | | | |
|---|---|---|---|
| **Face-to-face care** | −0.13 (−0.45 to 0.20) | . | −0.13 (−0.45 to 0.20) |
| −0.13 (−0.45 to 0.20) | **Biofeedback devices** | . | 0.00 (−0.33 to 0.33) |
| 0.23 (−0.15 to 0.60) | 0.35 (−0.02 to 0.73) | **VR** | **−0.35 (−0.54 to −0.17)** |
| −0.13 (−0.45 to 0.20) | −0.00 (−0.33 to 0.33) | **−0.35 (−0.54 to −0.17)** | **Control** |

**Table S21.** Physical function at post-intervention

| | | | | | | | |
|---|---|---|---|---|---|---|---|
| **Telemedicine** | . | 0.20 (-1.36 to 1.76) | -0.63 (-1.54 to 0.27) | . | . | -0.45 (-0.94 to 0.04) | . |
| 0.21 (-0.43 to 0.85) | **Web-App** | . | -0.08 (-0.97 to 0.80) | . | . | **−0.91 (−1.45 to −0.37)** | . |
| 0.15 (-0.45 to 0.75) | -0.06 (-0.70 to 0.59) | **Mobile-App** | -0.18 (-0.68 to 0.31) | . | . | **−1.09 (−1.78 to −0.39)** | . |
| -0.23 (-0.76 to 0.31) | -0.44 (-1.01 to 0.14) | -0.38 (-0.81 to 0.05) | **Face-to-face care** | 0.33 (-0.23 to 0.89) | **0.79 ( 0.14 to 1.45)** | -0.68 (-1.73 to 0.37) | 0.34 (-0.45 to 1.12) |
| 0.13 (-0.62 to 0.88) | -0.08 (-0.86 to 0.70) | -0.02 (-0.71 to 0.67) | 0.36 (-0.19 to 0.91) | **Biofeedback devices** | . | -0.77 (-2.24 to 0.71) | . |
| 0.28 (-0.42 to 0.98) | 0.07 (-0.66 to 0.80) | 0.13 (-0.53 to 0.79) | 0.51 (-0.04 to 1.06) | 0.15 (-0.62 to 0.92) | **VR** | -0.33 (-1.22 to 0.56) | . |
| **−0.57 (−1.01 to −0.13)** | **−0.78 (−1.27 to −0.30)** | **−0.73 (−1.22 to −0.23)** | -0.35 (-0.78 to 0.09) | **−0.70 (−1.38 to −0.03)** | **−0.86 (−1.45 to −0.26)** | **Control** | 0.16 (-0.98 to 1.30) |
| -0.06 (-0.85 to 0.74) | -0.27 (-1.09 to 0.56) | -0.21 (-0.97 to 0.55) | 0.17 (-0.49 to 0.83) | -0.19 (-1.04 to 0.67) | -0.34 (-1.17 to 0.50) | 0.52 (-0.19 to 1.23) | **Exergaming** |

**Table S22.** Physical function at short-term

| | | | | | | | |
|---|---|---|---|---|---|---|---|
| **Telemedicine** | . | . | −0.20 (−0.59 to 0.18) | . | . | −0.34 (−0.72 to 0.05) | . |
| −0.25 (−0.57 to 0.07) | **Web-App** | . | 0.04 (−0.24 to 0.33) | . | . | −0.09 (−0.25 to 0.08) | . |
| 0.21 (−0.35 to 0.77) | 0.46 (−0.08 to 1.00) | **Mobile-App** | −0.42 (−0.89 to 0.05) | . | . | . | . |
| −0.21 (−0.51 to 0.10) | 0.04 (−0.21 to 0.30) | −0.42 (−0.89 to 0.05) | **Face-to-face care** | 0.47 (−0.31 to 1.25) | . | . | 0.21 (−0.26 to 0.68) |
| 0.26 (−0.58 to 1.10) | 0.51 (−0.31 to 1.34) | 0.05 (−0.86 to 0.97) | 0.47 (−0.31 to 1.25) | **Biofeedback devices** | . | . | . |
| −0.11 (−0.81 to 0.58) | 0.14 (−0.51 to 0.78) | −0.32 (−1.15 to 0.51) | 0.09 (−0.59 to 0.78) | −0.38 (−1.41 to 0.66) | **VR** | −0.22 (−0.84 to 0.40) | . |
| **−0.34 (−0.64 to −0.03)** | −0.09 (−0.25 to 0.07) | −0.55 (−1.10 to 0.00) | −0.13 (−0.41 to 0.15) | −0.60 (−1.43 to 0.23) | −0.22 (−0.84 to 0.40) | **Control** | . |
| 0.00 (−0.56 to 0.56) | 0.25 (−0.28 to 0.79) | −0.21 (−0.87 to 0.46) | 0.21 (−0.26 to 0.68) | −0.26 (−1.17 to 0.65) | 0.11 (−0.71 to 0.94) | 0.34 (−0.21 to 0.89) | **Exergaming** |

**Table S23.** Physical function at mid-term

| | | | | |
|---|---|---|---|---|
| **Telemedicine** | . | . | . | . |
| −0.33 (−0.79 to 0.13) | **Web-App** | . | −0.25 (−0.64 to 0.15) | . |
| −0.12 (−0.59 to 0.36) | 0.21 (−0.06 to 0.48) | **Mobile-App** | . | . |
| −0.58 (−1.18 to 0.03) | −0.25 (−0.64 to 0.15) | −0.46 (−0.94 to 0.02) | **Face-to-face care** | **0.89 ( 0.48 to 1.30)** |
| 0.31 (−0.42 to 1.05) | **0.64 ( 0.07 to 1.21)** | 0.43 (−0.20 to 1.06) | **0.89 ( 0.48 to 1.30)** | **Biofeedback devices** |

**Table S24.** Physical function at long-term

| | | | |
|---|---|---|---|
| **Web−App** | . | . | −0.15 (−0.31 to 0.01) |
| −0.31 (−0.63 to 0.01) | **Face−to−face care** | −0.02 (−0.29 to 0.25) | 0.16 (−0.11 to 0.43) |
| **−0.33 (−0.65 to −0.01)** | −0.02 (−0.29 to 0.25) | **Biofeedback devices** | 0.18 (−0.09 to 0.45) |
| −0.15 (−0.31 to 0.01) | 0.16 (−0.11 to 0.43) | 0.18 (−0.09 to 0.45) | **Control** |

**Table S25.** Pain-related fear avoidance at post-intervention

| | | | | | | | |
|---|---|---|---|---|---|---|---|
| **Telemedicine** | . | 0.16 (−1.11 to 1.44) | −0.43 (−1.16 to 0.30) | . | . | −0.40 (−0.94 to 0.15) | . |
| −0.21 (−0.91 to 0.49) | **Web-App** | . | 0.09 (−0.79 to 0.97) | . | . | −0.33 (−1.04 to 0.38) | . |
| −0.12 (−0.72 to 0.48) | 0.09 (−0.62 to 0.80) | **Mobile-App** | −0.18 (−0.90 to 0.53) | . | . | −0.29 (−0.89 to 0.31) | . |
| −0.25 (−0.76 to 0.25) | −0.04 (−0.65 to 0.56) | −0.13 (−0.64 to 0.38) | **Face-to-face care** | −0.37 (−0.95 to 0.21) | 0.69 (−0.11 to 1.48) | −0.74 (−1.91 to 0.42) | 0.68 (−0.08 to 1.44) |
| −0.50 (−1.23 to 0.23) | −0.29 (−1.10 to 0.51) | −0.38 (−1.12 to 0.36) | −0.25 (−0.81 to 0.31) | **Biofeedback devices** | . | −0.86 (−2.02 to 0.31) | . |
| **0.69 ( 0.02 to 1.36)** | **0.90 ( 0.15 to 1.65)** | **0.81 ( 0.13 to 1.49)** | **0.95 ( 0.38 to 1.51)** | **1.19 ( 0.42 to 1.97)** | **VR** | **−1.33 (−1.99 to −0.66)** | . |
| −0.45 (−0.91 to 0.01) | −0.24 (−0.82 to 0.33) | −0.33 (−0.82 to 0.15) | −0.20 (−0.62 to 0.22) | 0.05 (−0.61 to 0.71) | **−1.14 (−1.68 to −0.61)** | **Control** | −0.14 (−0.90 to 0.63) |
| −0.07 (−0.77 to 0.62) | 0.14 (−0.64 to 0.91) | 0.05 (−0.66 to 0.75) | 0.18 (−0.40 to 0.76) | 0.43 (−0.36 to 1.22) | **−0.77 (−1.51 to −0.02)** | 0.38 (−0.20 to 0.96) | **Exergaming** |

**Table S26.** Pain-related fear avoidance at mid-term

| | | | | |
|---|---|---|---|---|
| **Telemedicine** | . | . | . | . |
| 0.38 (−0.13 to 0.89) | **Web-App** | . | −0.10 (−0.50 to 0.30) | . |
| **−0.38 (−0.69 to −0.07)** | **−0.76 (−1.26 to −0.26)** | **Mobile-App** | . | . |
| 0.28 (−0.04 to 0.60) | −0.10 (−0.50 to 0.30) | **0.66 ( 0.35 to 0.96)** | **Face-to-face care** | −0.08 (−0.29 to 0.14) |
| 0.20 (−0.12 to 0.52) | −0.18 (−0.63 to 0.27) | **0.58 ( 0.28 to 0.88)** | −0.08 (−0.29 to 0.14) | **Biofeedback devices** |

**Table S27.** Pain-related fear avoidance at long-term

| | | | |
|---|---|---|---|
| **Web-App** | . | . | **−0.23 (−0.41 to −0.05)** |
| −0.21 (−0.54 to 0.12) | **Face-to-face care** | −0.09 (−0.36 to 0.18) | −0.02 (−0.29 to 0.25) |
| −0.31 (−0.63 to 0.02) | −0.09 (−0.36 to 0.18) | **Biofeedback devices** | 0.07 (−0.20 to 0.35) |
| **−0.23 (−0.41 to −0.05)** | −0.02 (−0.29 to 0.25) | 0.07 (−0.20 to 0.35) | **Control** |

**Table S28.** Health-related quality of life at post-intervention

| | | | | | | |
|---|---|---|---|---|---|---|
| **Telemedicine** | . | −0.29 (−1.09 to 0.50) | −0.03 (−0.44 to 0.38) | . | . | **0.56 ( 0.23 to 0.89)** |
| 0.38 (−0.39 to 1.16) | **Web-App** | . | . | . | . | 0.05 (−0.67 to 0.77) |
| −0.01 (−0.34 to 0.32) | −0.39 (−1.17 to 0.39) | **Mobile-App** | 0.16 (−0.11 to 0.43) | . | . | 0.33 (−0.02 to 0.69) |
| 0.08 (−0.23 to 0.40) | −0.30 (−1.09 to 0.49) | 0.09 (−0.15 to 0.34) | **Face-to-face care** | 0.19 (−0.42 to 0.80) | −0.50 (−1.26 to 0.26) | . |
| 0.27 (−0.41 to 0.96) | −0.11 (−1.10 to 0.89) | 0.28 (−0.37 to 0.94) | 0.19 (−0.42 to 0.80) | **Biofeedback devices** | . | . |
| **−0.53 (−1.00 to −0.07)** | **−0.92 (−1.74 to −0.09)** | **−0.53 (−0.98 to −0.07)** | **−0.62 (−1.07 to −0.17)** | **−0.81 (−1.57 to −0.05)** | **VR** | **1.01 ( 0.56 to 1.46)** |
| **0.43 ( 0.15 to 0.72)** | 0.05 (−0.67 to 0.77) | **0.44 ( 0.15 to 0.74)** | **0.35 ( 0.03 to 0.67)** | 0.16 (−0.53 to 0.85) | **0.97 ( 0.57 to 1.36)** | **Control** |

**Table S29.** Health-related quality of life at short-term

| | | | | |
|---|---|---|---|---|
| **Telemedicine** | . | −0.23 (−1.92 to 1.46) | . | 0.24 (−1.00 to 1.49) |
| −0.35 (−2.78 to 2.09) | **Mobile-App** | 0.12 (−1.64 to 1.87) | . | . |
| −0.23 (−1.92 to 1.46) | 0.12 (−1.64 to 1.87) | **Face-to-face care** | . | . |
| −0.81 (−2.55 to 0.93) | −0.46 (−3.45 to 2.53) | −0.58 (−3.00 to 1.84) | **VR** | 1.05 (−0.16 to 2.26) |
| 0.24 (−1.00 to 1.49) | 0.59 (−2.15 to 3.32) | 0.47 (−1.63 to 2.57) | 1.05 (−0.16 to 2.26) | **Control** |

**Table S30.** Health-related quality of life at mid-term

| | | | |
|---|---|---|---|
| **Telemedicine** | . | −0.01 (−0.25 to 0.23) | . |
| 0.19 (−0.28 to 0.67) | **Mobile-App** | . | . |
| −0.01 (−0.25 to 0.23) | −0.20 (−0.73 to 0.33) | **Face-to-face care** | **−0.79 (−1.19 to −0.38)** |
| **−0.79 (−1.27 to −0.32)** | **−0.99 (−1.66 to −0.32)** | **−0.79 (−1.19 to −0.38)** | **Biofeedback devices** |

**Table S31.** Self-efficacy at post-intervention

| | | | | | | | |
|---|---|---|---|---|---|---|---|
| **Telemedicine** | . | . | −0.22 (−1.22 to 0.78) | . | . | 0.13 (−0.46 to 0.71) | . |
| −0.16 (−0.83 to 0.51) | **Web-App** | . | 0.07 (−0.84 to 0.98) | . | . | 0.25 (−0.25 to 0.75) | . |
| −0.39 (−1.06 to 0.29) | −0.22 (−0.85 to 0.41) | **Mobile-App** | 0.48 (−0.15 to 1.12) | . | . | 0.28 (−0.32 to 0.87) | . |
| −0.14 (−0.76 to 0.48) | 0.02 (−0.55 to 0.60) | 0.25 (−0.26 to 0.75) | **Face-to-face care** | −0.01 (−0.86 to 0.84) | −0.36 (−1.12 to 0.40) | 0.64 (−0.21 to 1.49) | . |
| −0.35 (−1.25 to 0.55) | −0.18 (−1.05 to 0.68) | 0.04 (−0.82 to 0.90) | −0.21 (−0.98 to 0.57) | **Biofeedback devices** | . | 0.65 (−0.20 to 1.50) | . |
| −0.50 (−1.48 to 0.48) | −0.33 (−1.29 to 0.62) | −0.11 (−1.02 to 0.80) | −0.36 (−1.12 to 0.40) | −0.15 (−1.24 to 0.94) | **VR** | . | . |
| 0.10 (−0.42 to 0.62) | 0.26 (−0.19 to 0.71) | 0.48 ( 0.00 to 0.97) | 0.24 (−0.24 to 0.72) | 0.45 (−0.33 to 1.22) | 0.60 (−0.30 to 1.50) | **Control** | **−0.77 (−1.49 to −0.05)** |
| −0.67 (−1.56 to 0.22) | −0.51 (−1.36 to 0.34) | −0.28 (−1.15 to 0.59) | −0.53 (−1.40 to 0.33) | −0.32 (−1.38 to 0.74) | −0.17 (−1.32 to 0.98) | **−0.77 (−1.49 to −0.05)** | **Exergaming** |

**Table S32.** Self-efficacy at short-term

| | | | | | |
|---|---|---|---|---|---|
| **Telemedicine** | . | . | −0.16 (−0.73 to 0.41) | 0.36 (−0.05 to 0.78) | . |
| −0.18 (−0.87 to 0.51) | **Web-App** | . | 0.02 (−0.37 to 0.40) | . | . |
| −0.14 (−0.59 to 0.31) | 0.04 (−0.78 to 0.86) | **Mobile-App** | . | **0.50 ( 0.33 to 0.68)** | . |
| −0.16 (−0.73 to 0.41) | 0.02 (−0.37 to 0.40) | −0.02 (−0.75 to 0.70) | **Face-to-face care** | . | . |
| 0.36 (−0.05 to 0.78) | 0.54 (−0.26 to 1.35) | **0.50 ( 0.33 to 0.68)** | 0.52 (−0.18 to 1.23) | **Control** | **−0.65 (−1.19 to −0.12)** |
| −0.29 (−0.97 to 0.38) | −0.11 (−1.08 to 0.85) | −0.15 (−0.71 to 0.41) | −0.13 (−1.01 to 0.75) | **−0.65 (−1.19 to −0.12)** | **Exergaming** |

**Table S33.** Self-efficacy at mid-term

| | | | | |
|---|---|---|---|---|
| **Telemedicine** | . | . | . | 0.27<br>(−0.15 to 0.70) |
| 0.15<br>(−0.32 to 0.62) | **Web-App** | . | 0.24<br>(−0.16 to 0.64) | 0.12<br>(−0.07 to 0.32) |
| −0.04<br>(−0.52 to 0.43) | −0.19<br>(−0.48 to 0.09) | **Mobile-App** | . | **0.32**<br>**( 0.11 to 0.53)** |
| 0.39<br>(−0.23 to 1.00) | 0.24<br>(−0.16 to 0.64) | 0.43<br>(−0.06 to 0.92) | **Face-to-face care** | . |
| 0.27<br>(−0.15 to 0.70) | 0.12<br>(−0.07 to 0.32) | **0.32**<br>**( 0.11 to 0.53)** | −0.11<br>(−0.56 to 0.33) | **Control** |

**Table S34.** Anxiety at post-intervention

| | | | | | | |
|---|---|---|---|---|---|---|
| **Telemedicine** | . | . | . | . | −0.19 (−0.46 to 0.07) | . |
| 0.04 (−0.68 to 0.77) | **Web-App** | . | −0.20 (−0.66 to 0.27) | . | . | . |
| −0.31 (−0.88 to 0.25) | −0.36 (−0.93 to 0.22) | **Mobile-App** | 0.18 (−0.17 to 0.54) | . | 0.02 (−0.66 to 0.70) | . |
| −0.16 (−0.71 to 0.40) | −0.20 (−0.66 to 0.27) | 0.16 (−0.18 to 0.49) | **Face-to-face care** | 0.11 (−0.46 to 0.69) | . | 0.18 (−0.36 to 0.72) |
| −0.11 (−0.48 to 0.26) | −0.15 (−0.81 to 0.51) | 0.20 (−0.29 to 0.70) | 0.04 (−0.42 to 0.51) | **VR** | −0.07 (−0.33 to 0.20) | . |
| −0.19 (−0.46 to 0.07) | −0.23 (−0.91 to 0.44) | 0.12 (−0.38 to 0.62) | −0.04 (−0.52 to 0.45) | −0.08 (−0.34 to 0.18) | **Control** | . |
| 0.02 (−0.75 to 0.80) | −0.02 (−0.73 to 0.69) | 0.33 (−0.30 to 0.97) | 0.18 (−0.36 to 0.72) | 0.13 (−0.58 to 0.85) | 0.21 (−0.52 to 0.94) | **Exergaming** |

**Table S35.** Depression at post-intervention

| | | | | | | | |
|---|---|---|---|---|---|---|---|
| **Telemedicine** | . | . | . | . | . | −0.22 (−0.44 to 0.00) | . |
| 0.07 (−0.53 to 0.66) | **Web-App** | . | −0.00 (−0.42 to 0.42) | . | . | . | . |
| −0.16 (−0.55 to 0.22) | −0.23 (−0.73 to 0.26) | **Mobile-App** | 0.22 (−0.07 to 0.50) | . | . | −0.04 (−0.40 to 0.32) | . |
| 0.07 (−0.35 to 0.49) | 0.00 (−0.42 to 0.42) | 0.23 (−0.03 to 0.49) | **Face-to-face care** | 0.40 (−0.27 to 1.06) | −0.16 (−0.70 to 0.37) | . | 0.16 (−0.34 to 0.66) |
| 0.46 (−0.33 to 1.25) | 0.40 (−0.39 to 1.18) | 0.63 (−0.09 to 1.34) | 0.40 (−0.27 to 1.06) | **Biofeedback devices** | . | . | . |
| −0.05 (−0.35 to 0.24) | −0.12 (−0.68 to 0.44) | 0.11 (−0.23 to 0.45) | −0.12 (−0.49 to 0.25) | −0.51 (−1.27 to 0.24) | **VR** | −0.17 (−0.38 to 0.03) | . |
| −0.22 (−0.44 to 0.00) | −0.29 (−0.84 to 0.26) | −0.06 (−0.37 to 0.26) | −0.29 (−0.65 to 0.07) | −0.68 (−1.44 to 0.07) | −0.17 (−0.36 to 0.02) | **Control** | . |
| 0.23 (−0.42 to 0.88) | 0.16 (−0.49 to 0.81) | 0.39 (−0.17 to 0.96) | 0.16 (−0.34 to 0.66) | −0.23 (−1.06 to 0.60) | 0.28 (−0.34 to 0.90) | 0.45 (−0.16 to 1.07) | **Exergaming** |

**Section 12.** Heterogeneity

Heterogeneity was assessed using the between-study variance ($\tau^2$) estimated from random-effects models. $I^2$ values represent the proportion of total variability attributable to between-study heterogeneity rather than sampling error. Higher values indicate greater variability across included studies. VR, virtual reality; Mobile-App, mobile application-based intervention; Web-App, web-based intervention.

**Table S36.** Overall heterogeneity across outcomes and time points

| Outcome | Time point | $\tau^2$ | $I^2$ (%) |
|---|---|---|---|
| Pain intensity | Post-intervention | 0.57 | **87.79** |
| | Short-term | 4.63 | **94.75** |
| | Mid-term | 0.02 | 55.45 |
| | Long-term | 0.01 | 40.20 |
| Physical function | Post-intervention | 0.55 | **87.41** |
| | Short-term | 0.00 | 0.00 |
| | Mid-term | 0.00 | 0.00 |
| | Long-term | 0.00 | 0.00 |
| Pain-related fear avoidance | Post-intervention | 0.34 | **80.80** |
| | Mid-term | 0.00 | 0.00 |
| | Long-term | 0.00 | 0.00 |
| Health-related quality of life | Post-intervention | 0.08 | 61.35 |
| | Short-term | 0.66 | **85.60** |
| | Mid-term | 0.00 | 0.00 |
| Self-efficacy | Post-intervention | 0.18 | **79.97** |
| | Short-term | 0.00 | 0.00 |
| | Mid-term | 0.00 | 0.00 |
| Anxiety | Post-intervention | 0.02 | 14.40 |
| Depression | Post-intervention | 0.01 | 0.77 |

**Table S37.** Direct-comparison heterogeneity for pain intensity at post-intervention

| Comparison | k | $\tau^2$ | $I^2$ (%) | Q statistic | P value |
|---|---|---|---|---|---|
| Biofeedback devices vs Control | 1 | NE | NE | NE | NE |
| Biofeedback devices vs Face-to-face care | 9 | 0.42 | 79.96 | 39.93 | **<0.001** |
| Biofeedback devices vs Telemedicine | 1 | NE | NE | NE | NE |
| Control vs Exergaming | 4 | 0.00 | 0.00 | 2.93 | 0.40 |
| Control vs Face-to-face care | 3 | 0.00 | 0.00 | 0.41 | 0.81 |
| Control vs Mobile-App | 8 | 0.33 | 88.07 | 58.68 | **<0.001** |
| Control vs Telemedicine | 9 | 0.04 | 50.16 | 16.05 | 0.04 |
| Control vs VR | 8 | 2.14 | 95.30 | 148.81 | **<0.001** |
| Control vs Web-App | 6 | 0.28 | 87.23 | 39.15 | **<0.001** |
| Exergaming vs Face-to-face care | 4 | 6.17 | 94.82 | 57.92 | **<0.001** |
| Face-to-face care vs Mobile-App | 11 | 0.10 | 66.00 | 29.41 | 0.001 |
| Face-to-face care vs Telemedicine | 5 | 0.00 | 3.69 | 4.15 | 0.39 |
| Face-to-face care vs VR | 8 | 1.29 | 91.65 | 83.86 | **<0.001** |
| Face-to-face care vs Web-App | 4 | 0.07 | 66.07 | 8.84 | 0.03 |
| Mobile-App vs Telemedicine | 1 | NE | NE | NE | NE |

**Table S38.** Direct-comparison heterogeneity for pain intensity at short-term

| Comparison | k | $\tau^2$ | $I^2$ (%) | Q statistic | P value |
|---|---|---|---|---|---|
| Biofeedback devices vs Face-to-face care | 1 | NE | NE | NE | NE |
| Control vs Mobile-App | 1 | NE | NE | NE | NE |
| Control vs Telemedicine | 1 | NE | NE | NE | NE |
| Control vs VR | 7 | 8.39 | 96.74 | 183.97 | **<0.001** |
| Face-to-face care vs Mobile-App | 2 | 0.00 | 0.00 | 0.19 | 0.66 |
| Face-to-face care vs Telemedicine | 2 | 0.05 | 38.64 | 1.63 | 0.20 |

| | | | | | |
|---|---|---|---|---|---|
| Face-to-face care vs Web-App | 2 | 0.00 | 0.00 | 0.00 | 1.00 |

**Table S39.** Direct-comparison heterogeneity for physical function at post-intervention

| Comparison | k | $\tau^2$ | $I^2$ (%) | Q statistic | P value |
|---|---|---|---|---|---|
| Biofeedback devices vs Control | 1 | NE | NE | NE | NE |
| Biofeedback devices vs Face-to-face care | 8 | 0.24 | 71.98 | 24.98 | **<0.001** |
| Control vs Exergaming | 2 | 0.00 | 0.00 | 0.56 | 0.45 |
| Control vs Face-to-face care | 2 | 0.00 | 0.00 | 0.64 | 0.42 |
| Control vs Mobile-App | 5 | 2.02 | 95.18 | 82.96 | **<0.001** |
| Control vs Telemedicine | 10 | 0.02 | 21.59 | 11.48 | 0.24 |
| Control vs VR | 3 | 0.01 | 1.91 | 2.04 | 0.36 |
| Control vs Web-App | 8 | 1.24 | 94.97 | 139.16 | **<0.001** |
| Exergaming vs Face-to-face care | 4 | 0.55 | 85.89 | 21.26 | **<0.001** |
| Face-to-face care vs Mobile-App | 10 | 0.04 | 53.09 | 19.19 | 0.02 |
| Face-to-face care vs Telemedicine | 3 | 0.45 | 82.68 | 11.55 | 0.003 |
| Face-to-face care vs VR | 6 | 1.29 | 92.11 | 63.41 | **<0.001** |
| Face-to-face care vs Web-App | 3 | 0.00 | 0.00 | 1.77 | 0.41 |
| Mobile-App vs Telemedicine | 1 | NE | NE | NE | NE |

**Table S40.** Direct-comparison heterogeneity for pain-related fear avoidance at post-intervention

| Comparison | k | $\tau^2$ | $I^2$ (%) | Q statistic | P value |
|---|---|---|---|---|---|
| Biofeedback devices vs Control | 1 | NE | NE | NE | NE |
| Biofeedback devices vs Face-to-face care | 5 | 0.43 | 83.37 | 24.06 | **<0.001** |
| Control vs Exergaming | 3 | 0.45 | 82.25 | 11.27 | 0.004 |
| Control vs Face-to-face care | 1 | NE | NE | NE | NE |

| | | | | | |
|---|---|---|---|---|---|
| Control vs Mobile-App | 4 | 0.05 | 64.13 | 8.36 | 0.04 |
| Control vs Telemedicine | 5 | 0.02 | 29.69 | 5.69 | 0.22 |
| Control vs VR | 4 | 1.34 | 90.44 | 31.39 | **<0.001** |
| Control vs Web-App | 3 | 0.00 | 0.00 | 0.12 | 0.94 |
| Exergaming vs Face-to-face care | 3 | 0.61 | 85.14 | 13.46 | 0.001 |
| Face-to-face care vs Mobile-App | 3 | 0.00 | 0.00 | 0.62 | 0.73 |
| Face-to-face care vs Telemedicine | 3 | 0.14 | 64.20 | 5.59 | 0.06 |
| Face-to-face care vs VR | 3 | 1.28 | 90.61 | 21.29 | **<0.001** |
| Face-to-face care vs Web-App | 2 | 0.00 | 0.00 | 0.05 | 0.83 |
| Mobile-App vs Telemedicine | 1 | NE | NE | NE | NE |

**Table S41.** Direct-comparison heterogeneity for health-related quality of life at short-term

| Comparison | k | $\tau^2$ | $I^2$ (%) | Q statistic | P value |
|---|---|---|---|---|---|
| Control vs Telemedicine | 2 | 0.00 | 0.00 | 0.27 | 0.60 |
| Control vs VR | 2 | 1.32 | 92.65 | 13.61 | **<0.001** |
| Face-to-face care vs Mobile-App | 1 | NE | NE | NE | NE |
| Face-to-face care vs Telemedicine | 1 | NE | NE | NE | NE |

**Table S42.** Direct-comparison heterogeneity for self-efficacy at post-intervention

| Comparison | k | $\tau^2$ | $I^2$ (%) | Q statistic | P value |
|---|---|---|---|---|---|
| Biofeedback devices vs Control | 1 | NE | NE | NE | NE |
| Biofeedback devices vs Face-to-face care | 1 | NE | NE | NE | NE |
| Control vs Exergaming | 2 | 0.39 | 80.43 | 5.11 | 0.02 |
| Control vs Face-to-face care | 1 | NE | NE | NE | NE |

| | | | | | |
|---|---|---|---|---|---|
| Control vs Mobile-App | 2 | 0.00 | 0.00 | 0.22 | 0.64 |
| Control vs Telemedicine | 3 | 0.00 | 20.61 | 2.52 | 0.28 |
| Control vs Web-App | 3 | 0.37 | 93.11 | 29.04 | **<0.001** |
| Face-to-face care vs Mobile-App | 3 | 1.12 | 85.92 | 14.20 | **<0.001** |
| Face-to-face care vs Telemedicine | 1 | NE | NE | NE | NE |
| Face-to-face care vs VR | 2 | 0.00 | 0.00 | 0.01 | 0.94 |
| Face-to-face care vs Web-App | 1 | NE | NE | NE | NE |

**Section 13.** Inconsistency

Global inconsistency was assessed using the design-by-treatment interaction model. Local inconsistency was assessed using node-splitting analyses comparing direct and indirect evidence. A two-sided *P* value <0.05 was considered evidence of statistically significant inconsistency.

**Table S43.** Global inconsistency

| Outcome | Time point | Q statistic | df | *P* value | Global inconsistency |
|---|---|---|---|---|---|
| Pain intensity | Post-intervention | 24.10 | 14 | **<0.05** | **Yes** |
| | Short-term | 0.32 | 1 | 0.57 | No |
| Physical function | Post-intervention | 9.06 | 10 | 0.52 | No |
| | Short-term | 0.01 | 1 | 0.91 | No |
| Pain-related fear avoidance | Post-intervention | 12.65 | 8 | 0.12 | No |
| Health-related quality of life | Post-intervention | 6.11 | 4 | 0.19 | No |
| Self-efficacy | Post-intervention | 2.05 | 3 | 0.56 | No |
| Anxiety | Post-intervention | 0.18 | 1 | 0.67 | No |
| Depression | Post-intervention | 0.03 | 1 | 0.87 | No |

**Table S44.** Local inconsistency assessment for pain intensity at post-intervention

| Comparison | k | Direct evidence proportion | NMA SMD | Direct SMD | Indirect SMD | Direct − indirect difference | P value |
|---|---|---|---|---|---|---|---|
| Biofeedback devices vs Control | 1 | 0.17 | −0.48 | −0.55 | −0.47 | −0.08 | 0.92 |
| Biofeedback devices vs Face-to-face care | 9 | 0.94 | −0.17 | −0.19 | 0.09 | −0.28 | 0.80 |
| Biofeedback devices vs Telemedicine | 1 | 0.18 | 0.02 | 0.84 | −0.15 | 0.99 | 0.26 |
| Exergaming vs Control | 4 | 0.61 | −1.09 | −0.31 | −2.28 | 1.98 | **<0.005** |
| Face-to-face care vs Control | 3 | 0.16 | −0.31 | −0.67 | −0.24 | −0.43 | 0.40 |
| Mobile-App vs Control | 8 | 0.58 | −0.67 | −0.71 | −0.62 | −0.08 | 0.85 |
| Telemedicine vs Control | 9 | 0.69 | −0.51 | −0.48 | −0.57 | 0.09 | 0.85 |
| VR vs Control | 8 | 0.60 | −1.43 | −1.66 | −1.08 | −0.57 | 0.22 |
| Web-App vs Control | 6 | 0.69 | −0.54 | −0.59 | −0.42 | −0.18 | 0.76 |

| | | | | | | | |
|---|---|---|---|---|---|---|---|
| Exergaming vs Face-to-face care | 4 | 0.57 | −0.78 | −1.38 | 0.03 | −1.41 | **<0.05** |
| Face-to-face care vs Mobile-App | 11 | 0.67 | 0.36 | 0.34 | 0.41 | −0.08 | 0.86 |
| Face-to-face care vs Telemedicine | 5 | 0.45 | 0.19 | 0.16 | 0.22 | −0.06 | 0.89 |
| Face-to-face care vs VR | 8 | 0.61 | 1.12 | 0.89 | 1.47 | −0.57 | 0.22 |
| Face-to-face care vs Web-App | 4 | 0.52 | 0.23 | 0.26 | 0.20 | 0.06 | 0.92 |
| Mobile-App vs Telemedicine | 1 | 0.11 | −0.17 | −0.22 | −0.16 | −0.06 | 0.94 |

**Section 14.** Comparison-adjusted funnel plots for assessment of small-study effects

Comparison-adjusted funnel plots were used to assess potential small-study effects in networks with sufficient data for this assessment. Each point represents an effect estimate from an individual study comparison. The horizontal axis represents the comparison-adjusted effect estimate, and the vertical axis represents its standard error. Funnel-plot asymmetry may indicate small-study effects, but it may also reflect heterogeneity or differences in network structure. Mobile-App, mobile applications; VR, virtual reality; Web-App, web applications.

**Figure S25.** Funnel plot for pain intensity at post-intervention

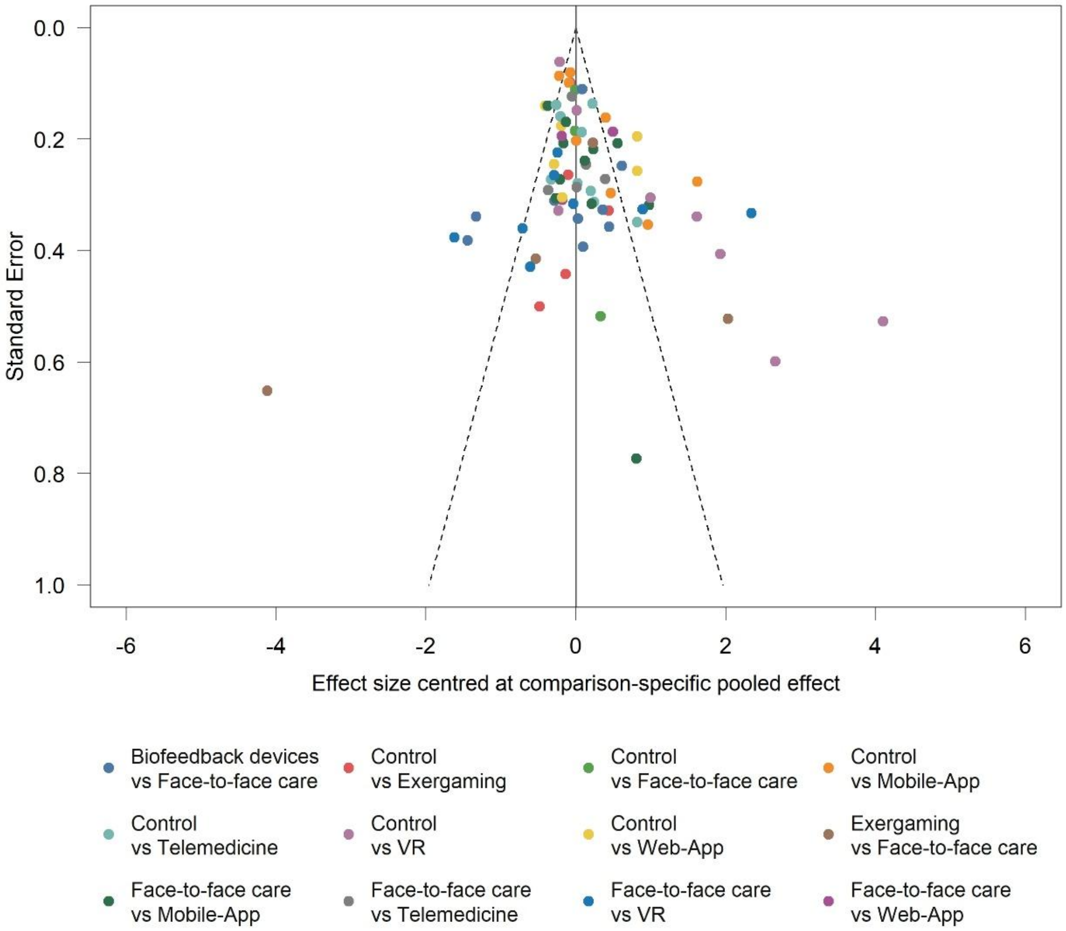

**Figure S26.** Funnel plot for physical function at post-intervention

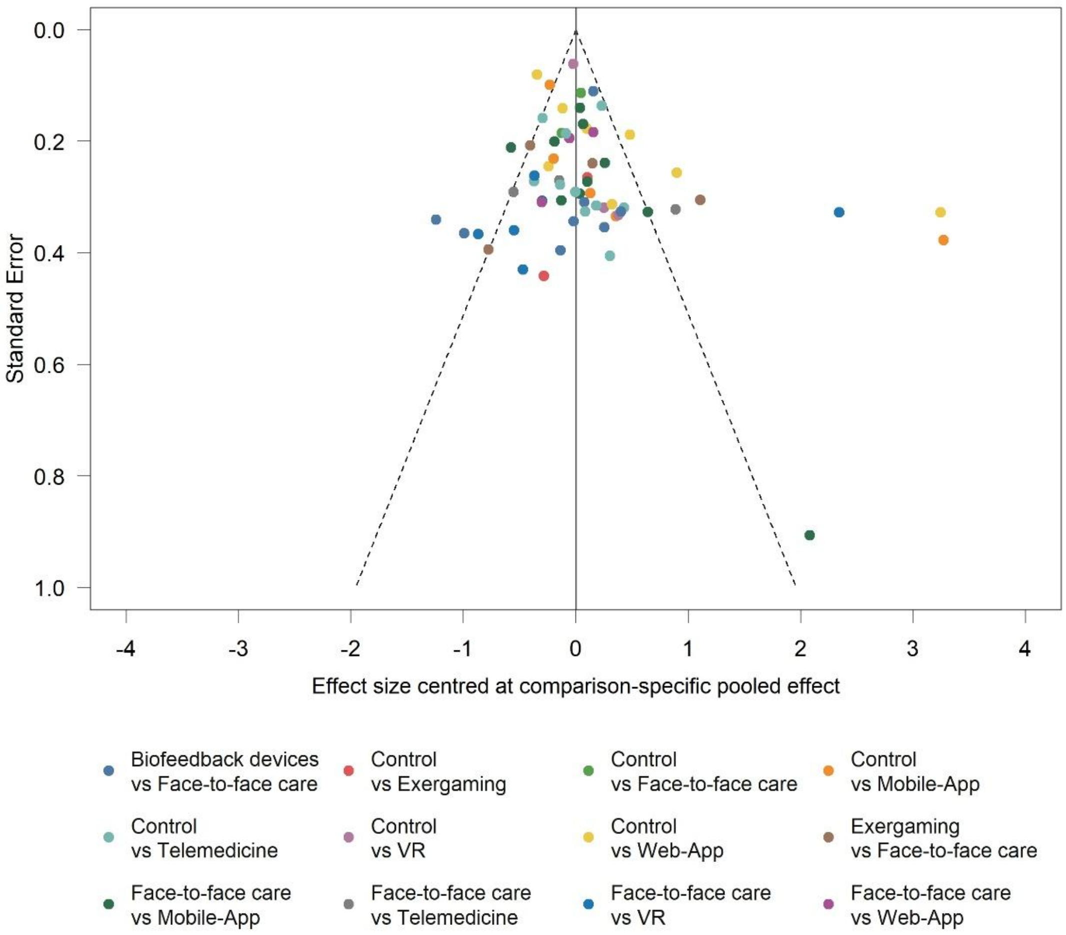

**Figure S27.** Funnel plot for pain-related fear avoidance at post-intervention

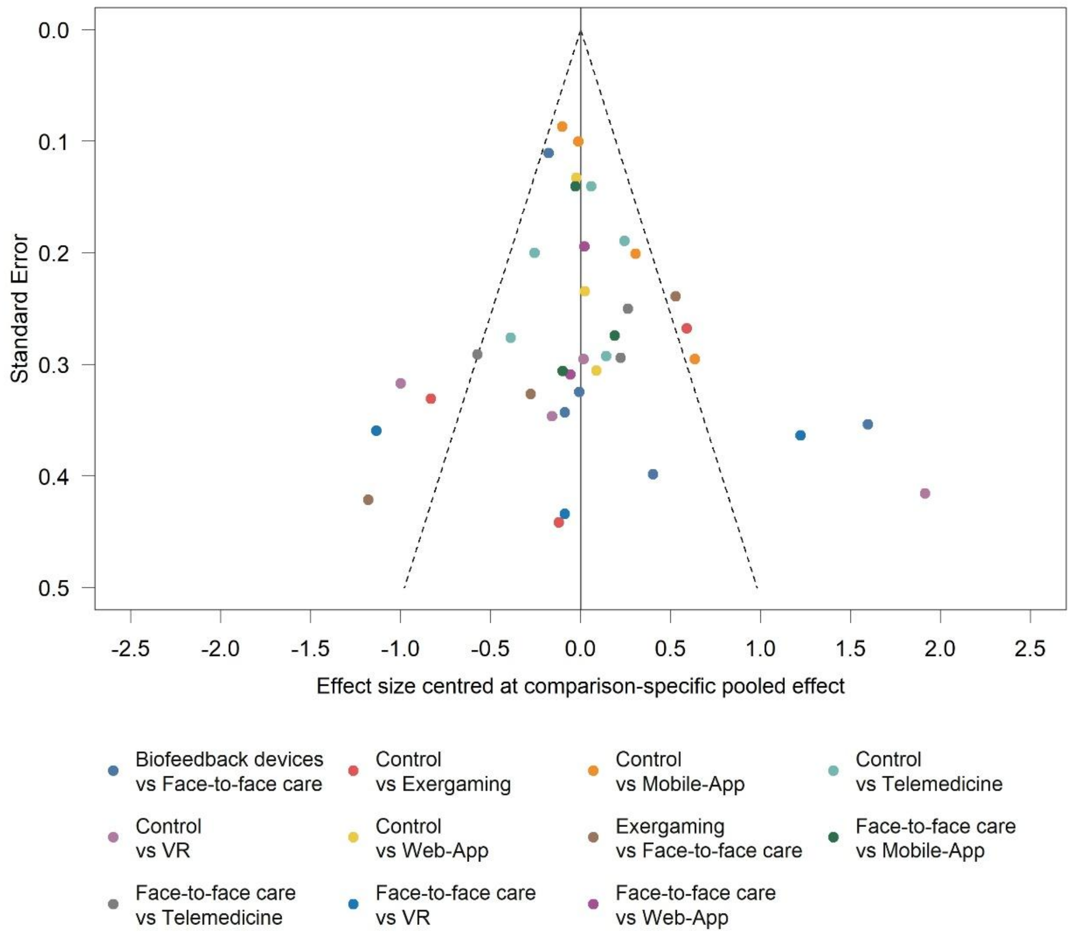

**Figure S28.** Funnel plot for health-related quality of life at post-intervention

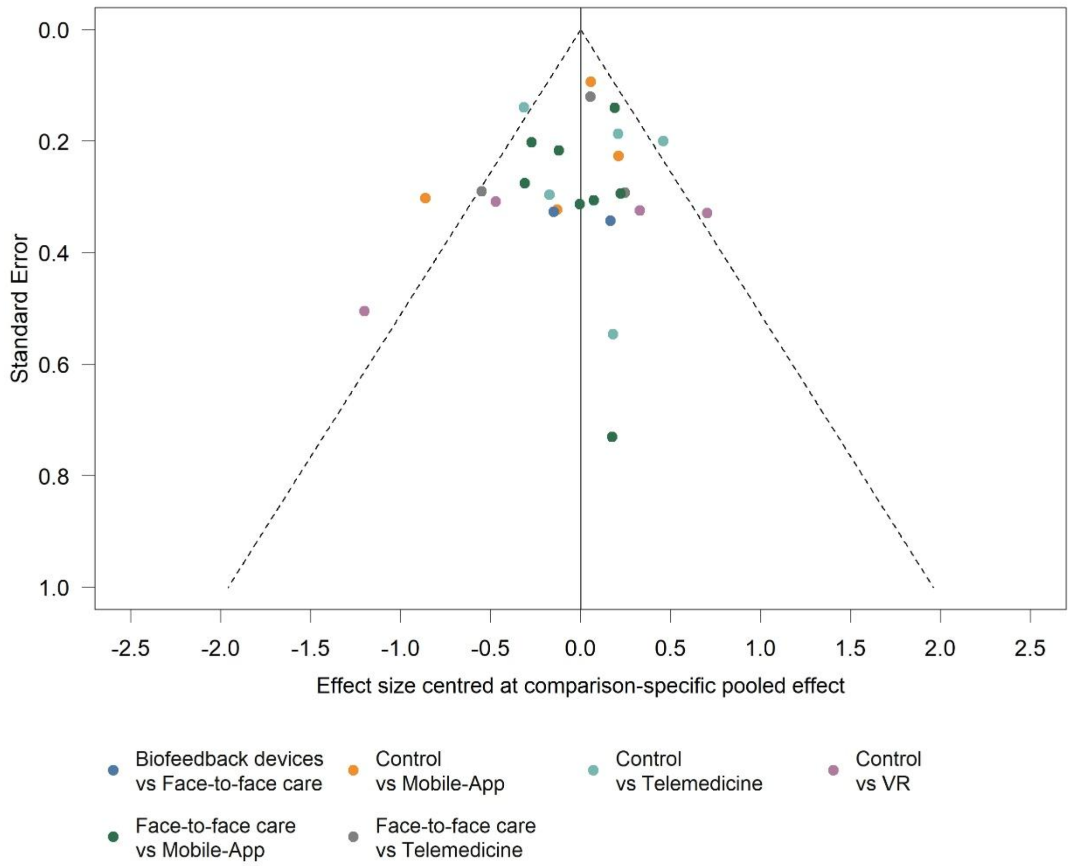

**Figure S29.** Funnel plot for self-efficacy at post-intervention

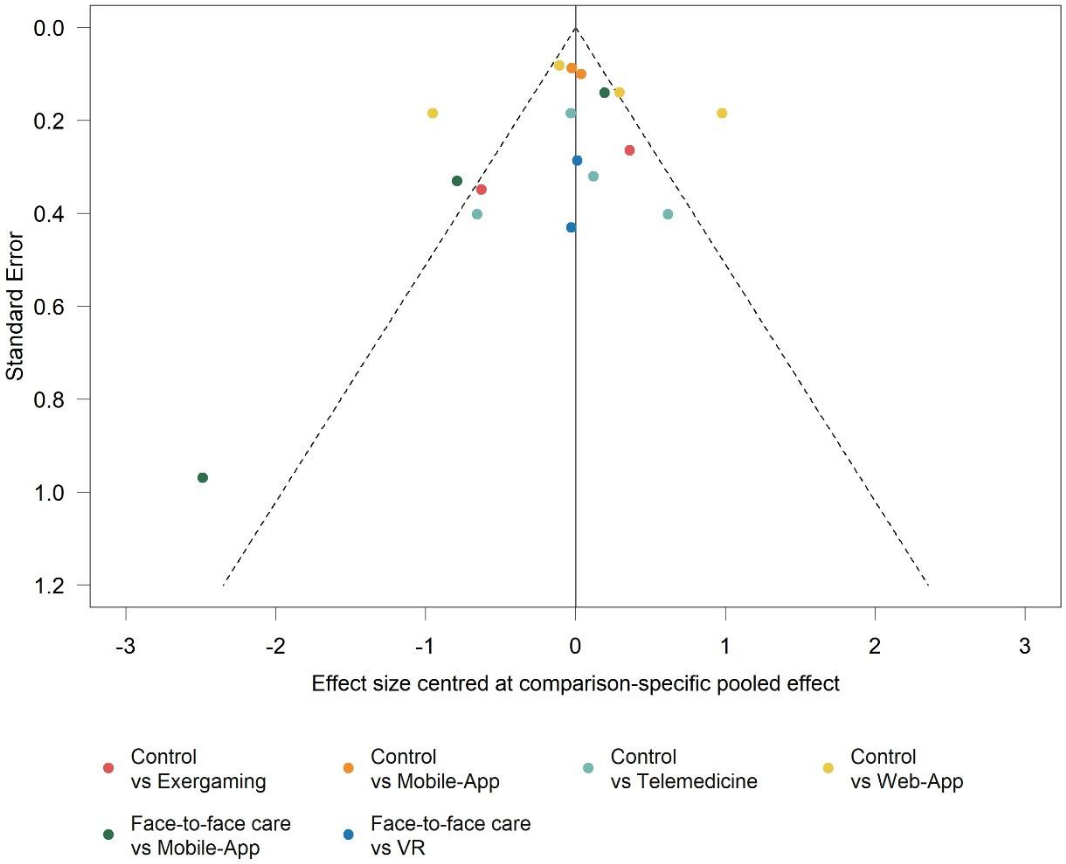

**Figure S30.** Funnel plot for anxiety at post-intervention

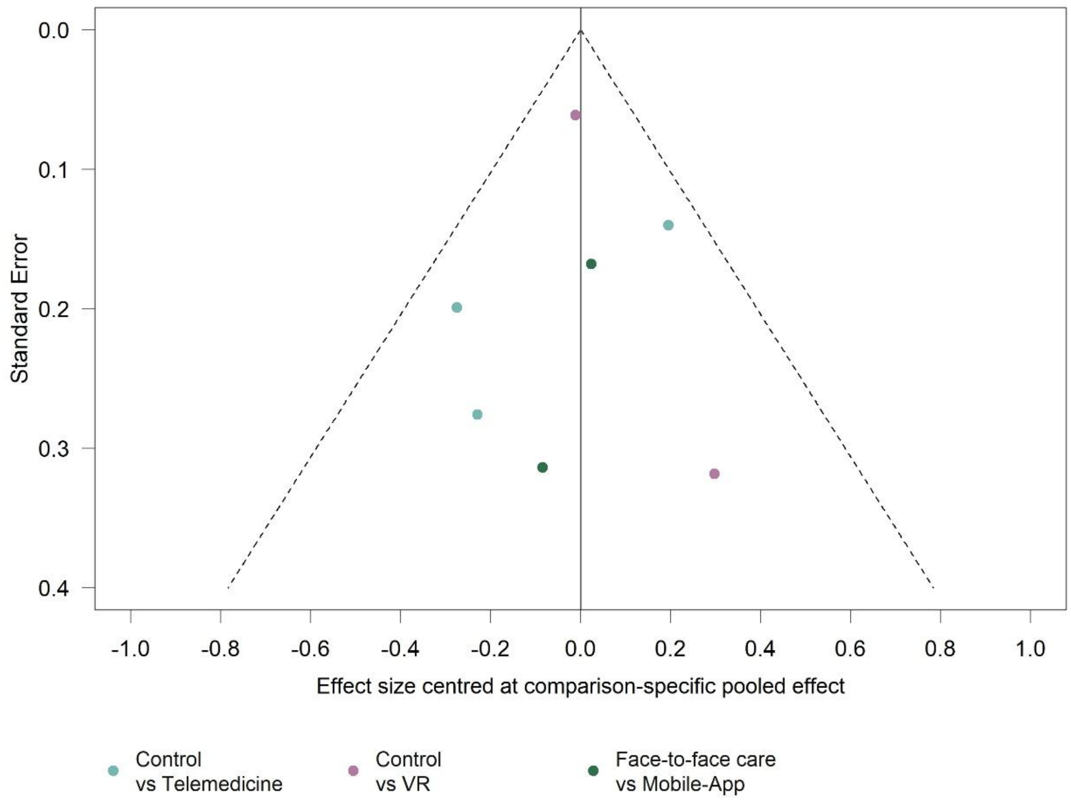

**Figure S31.** Funnel plot for depression at post-intervention

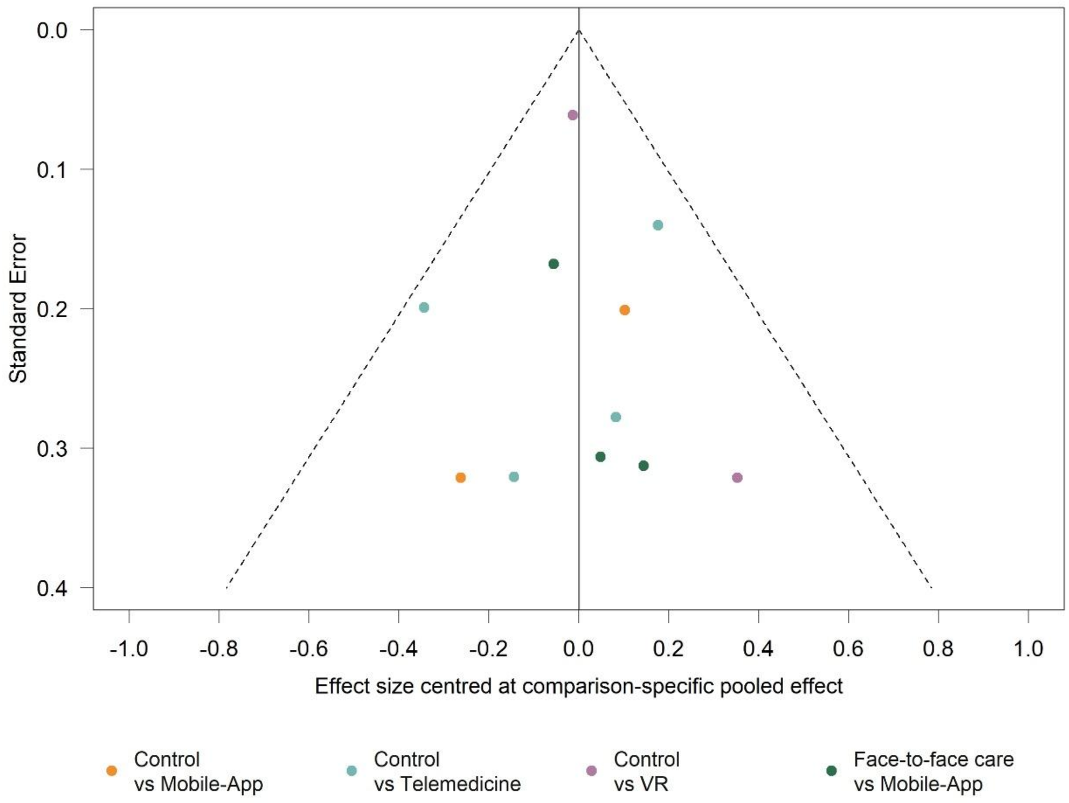

**Section 15.** Sensitivity analyses excluding studies at high risk of bias

This sensitivity analysis excluded studies rated as having a high risk of bias. Network meta-analysis estimates are presented as standardised mean differences (SMDs) with 95% confidence intervals (CIs) from random-effects models. For pain intensity, physical function, pain-related fear avoidance, anxiety, and depression, negative SMD values indicate improvement; for health-related quality of life and self-efficacy, positive SMD values indicate improvement. Estimates with 95% CIs excluding zero indicate evidence of a difference. In the network plots, node size is proportional to the number of participants assigned to each intervention, and line thickness is proportional to the number of randomised controlled trials contributing direct evidence; the absence of a line indicates that no direct comparison was available. Treatment-node definitions are provided in Supplementary Table 4.1, and study-level risk-of-bias assessments are provided in Supplementary 7. Mobile-App=mobile applications; VR=virtual reality; Web-App=web applications.

**Figure S32.** Network plots for pain intensity

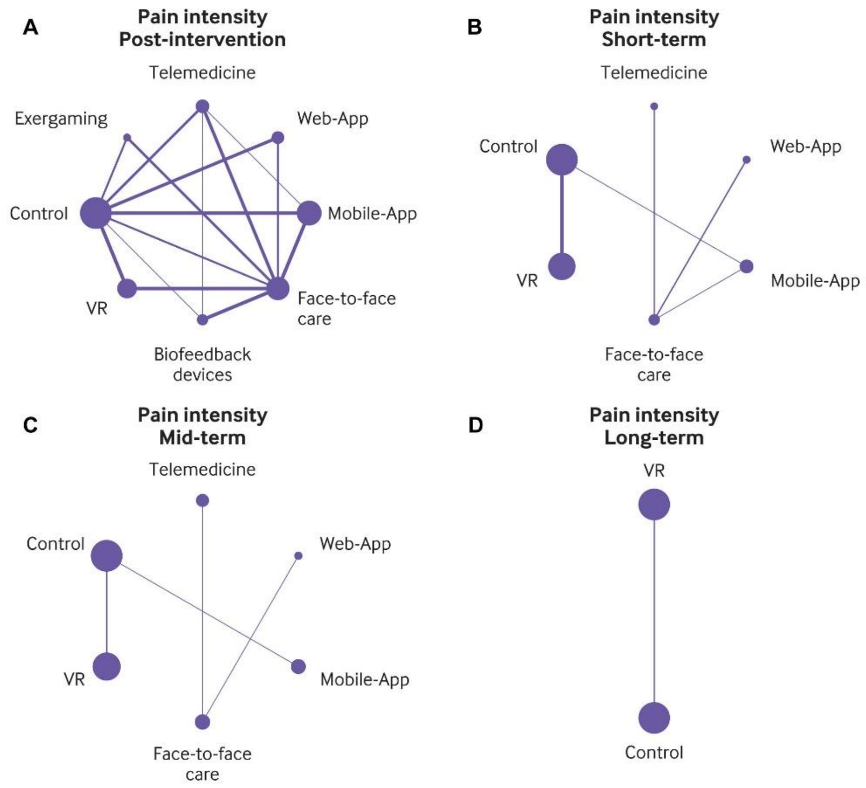

**Figure S33.** Network plots for physical function

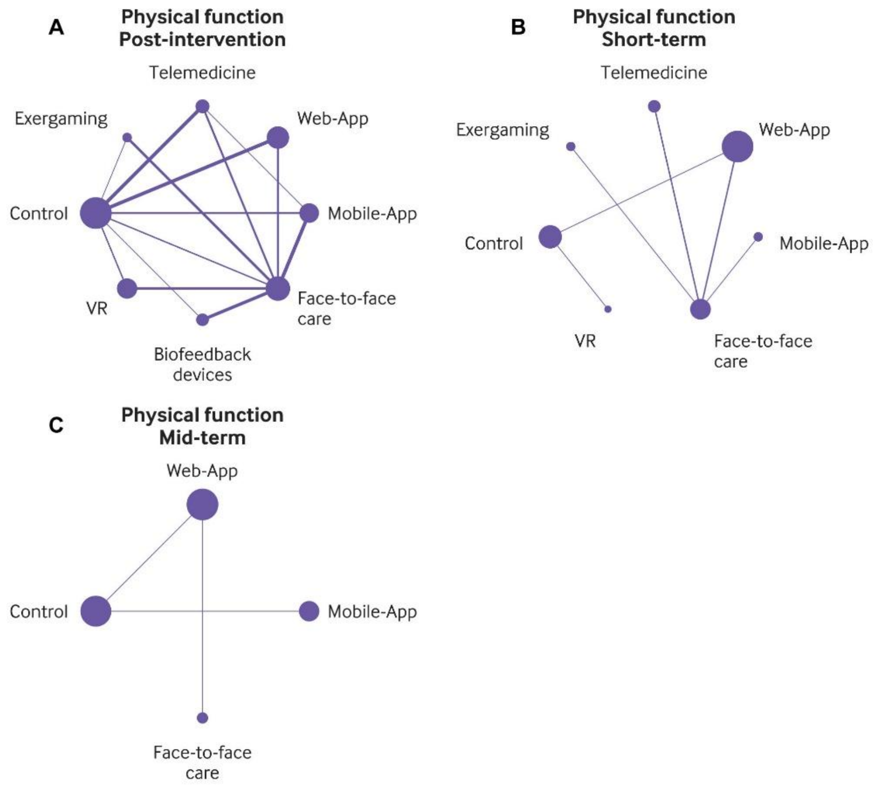

**Figure S34.** Network plots for pain-related fear avoidance

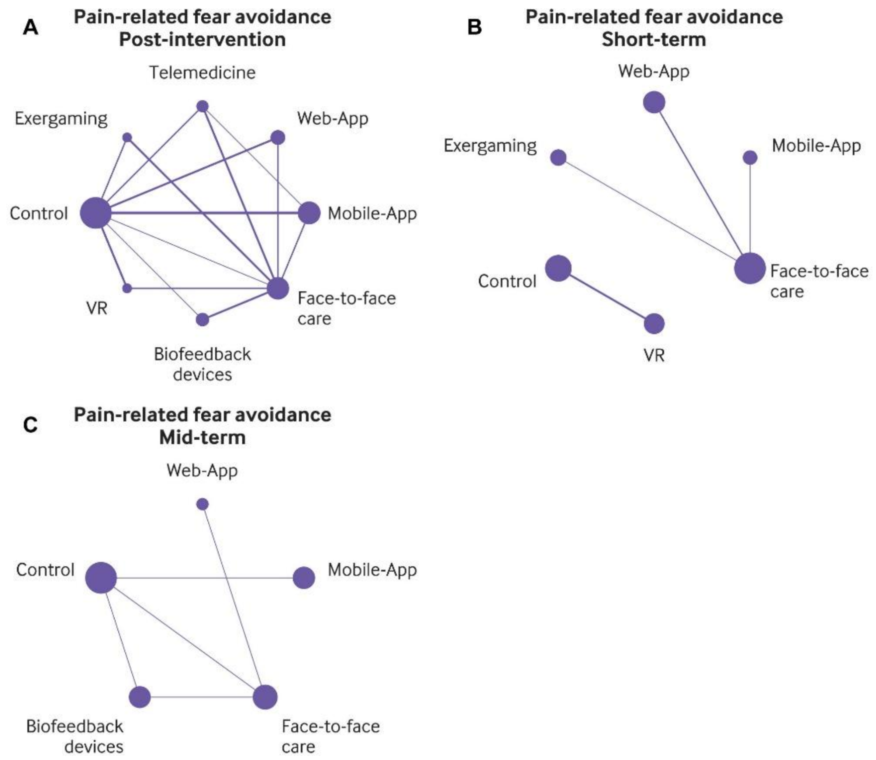

**Figure S35.** Network plots for health-related quality of life

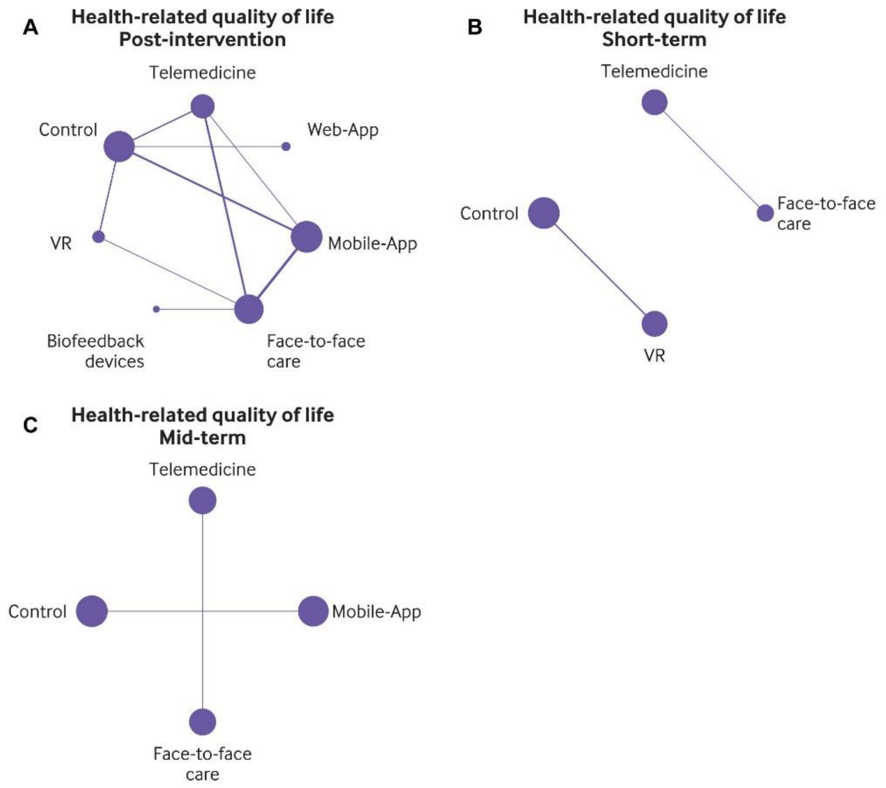

**Figure S36.** Network plots for self-efficacy

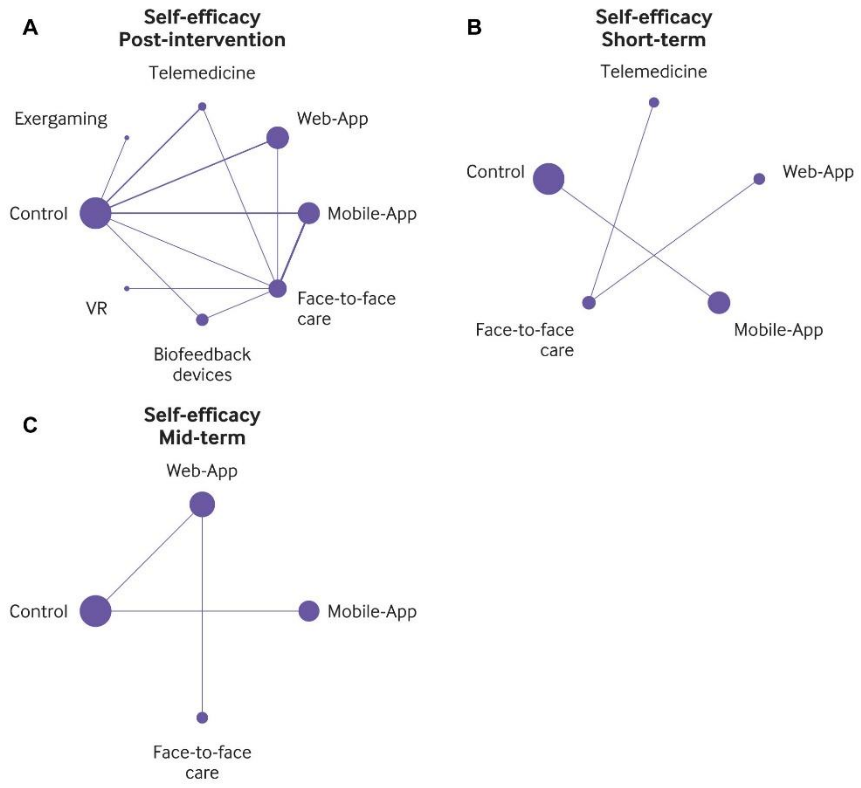

**Figure S37.** Network plots for anxiety

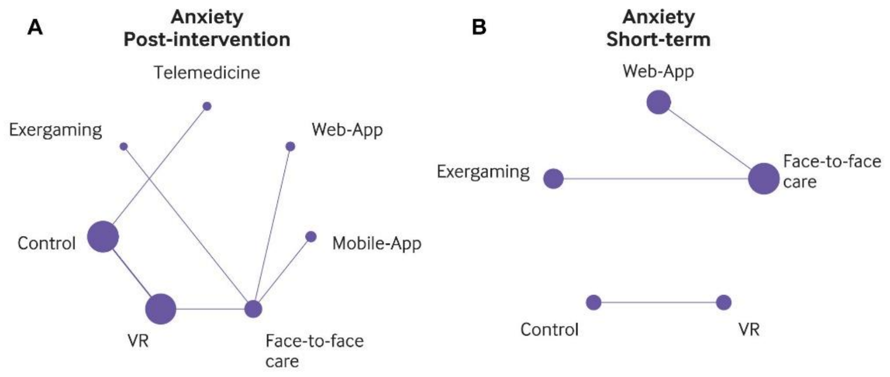

**Figure S38.** Network plots for depression

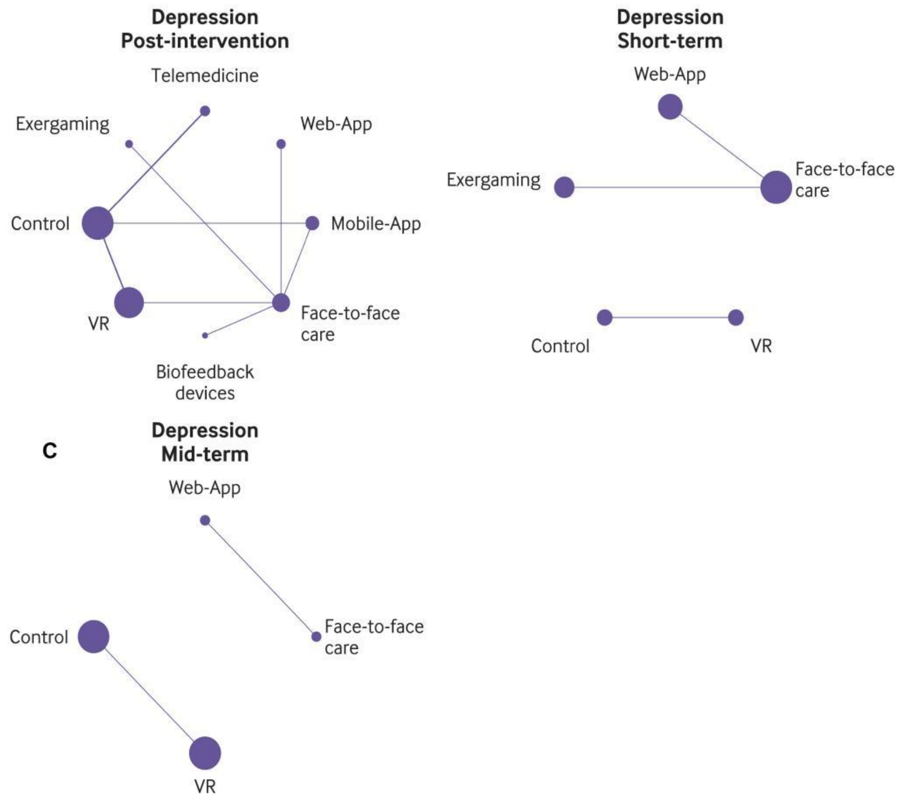

**Figure S39.** Forest plot for pain intensity at post-intervention

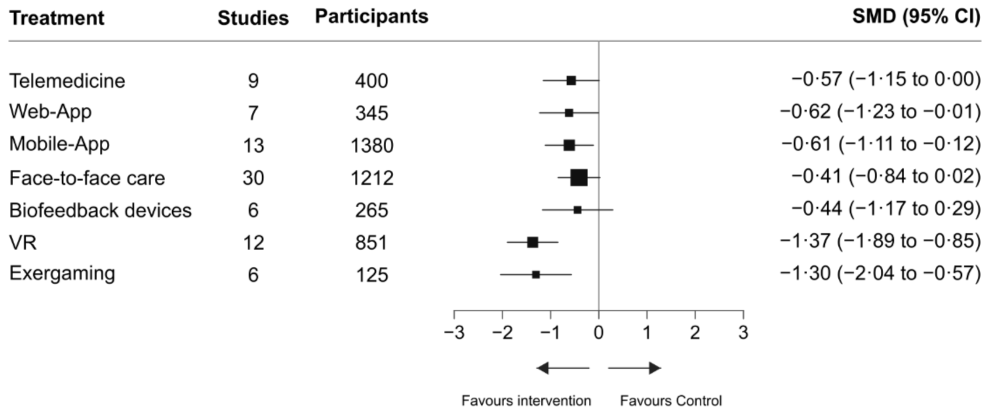

**Figure S40.** Forest plot for pain intensity at short-term

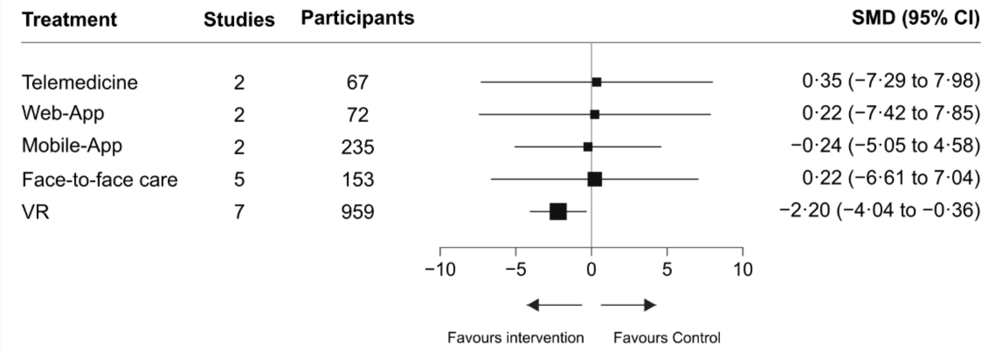

**Figure S41.** Forest plot for pain intensity at long-term

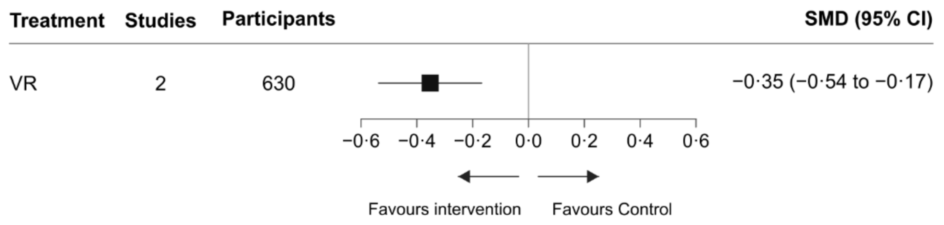

**Figure S42.** Forest plot for physical function at post-intervention

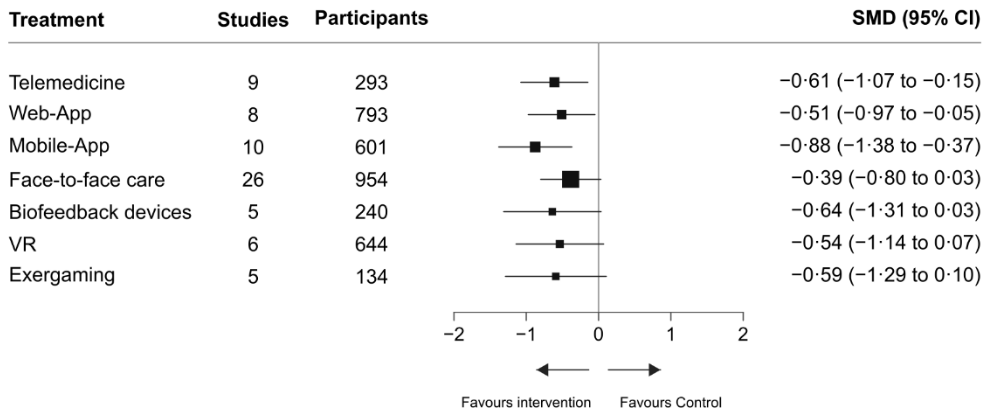

**Figure S43.** Forest plot for physical function at short-term

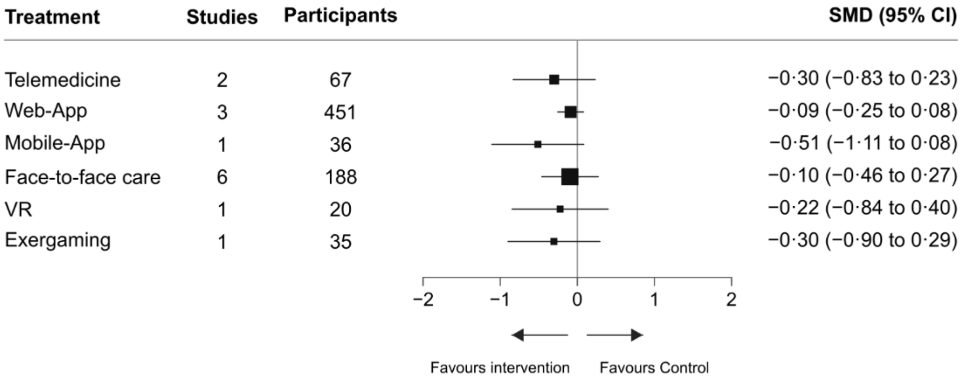

**Figure S44.** Forest plot for physical function at mid-term

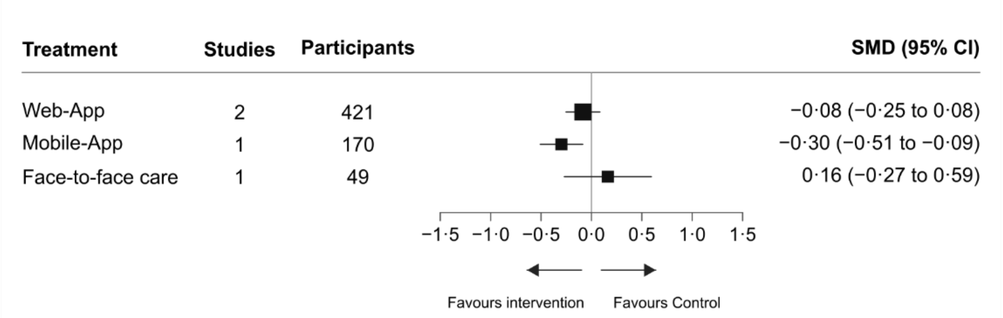

**Figure S45.** Forest plot for health-related quality of life at post-intervention

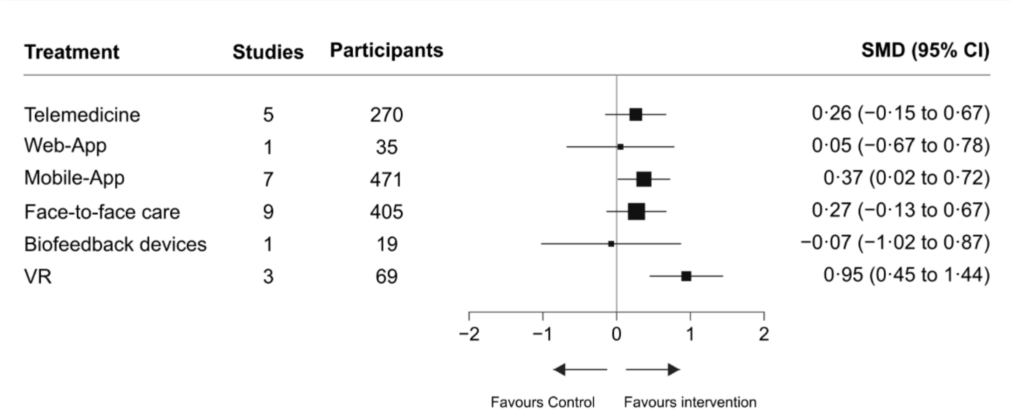

**Figure S46.** Forest plot for self-efficacy at post-intervention

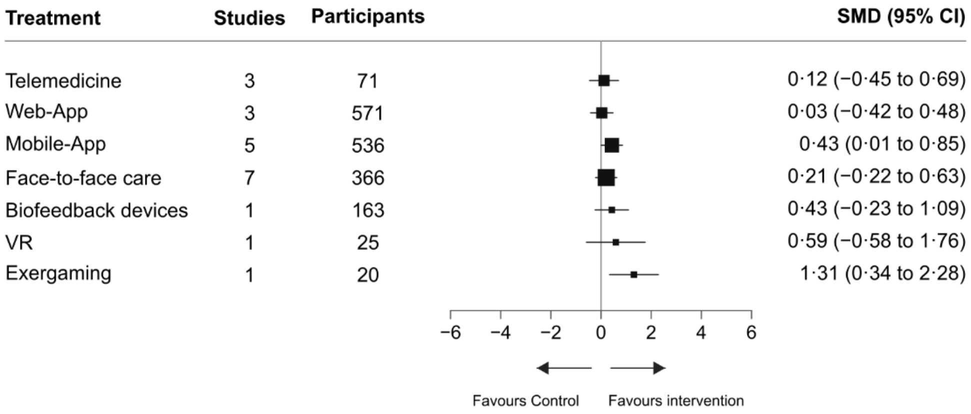

**Figure S47.** Forest plot for self-efficacy at mid-term

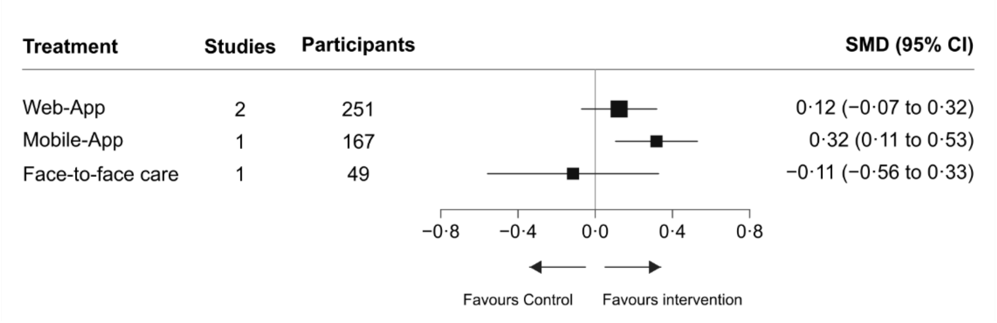

**Figure S48.** Forest plot for anxiety at post-intervention

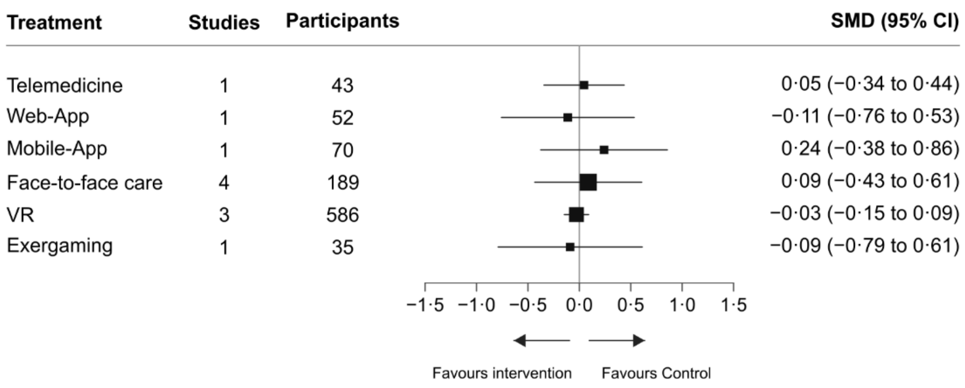

**Figure S49.** Forest plot for depression at post-intervention

| Treatment | Studies | Participants | SMD (95% CI) |
|---|---|---|---|
| Telemedicine | 2 | 62 | 0·05 (−0·28 to 0·38) |
| Web-App | 1 | 52 | −0·37 (−0·90 to 0·15) |
| Mobile-App | 2 | 121 | −0·12 (−0·45 to 0·22) |
| Face-to-face care | 5 | 208 | −0·37 (−0·74 to −0·01) |
| Biofeedback devices | 1 | 19 | −0·77 (−1·51 to −0·03) |
| VR | 3 | 586 | −0·16 (−0·27 to −0·04) |
| Exergaming | 1 | 35 | −0·54 (−1·13 to 0·06) |

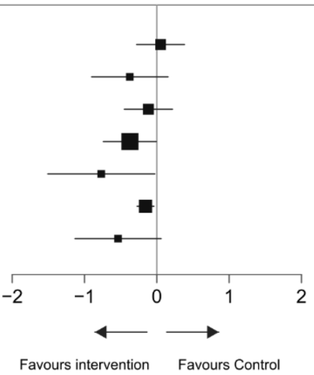

**Section 16.** Sensitivity analyses excluding studies with estimated or converted outcome data

This sensitivity analysis excluded studies contributing outcome data that were not directly reported but had been statistically derived or converted for the primary analysis. This included standard deviations derived from standard errors, confidence intervals, exact P values, or other reported measures of dispersion, and means and standard deviations estimated from medians with interquartile ranges, ranges, or other non-standard summary statistics. Network meta-analysis estimates are presented as standardised mean differences (SMDs) with 95% confidence intervals (CIs) from random-effects models. Negative SMDs indicate improvement for pain intensity, physical function, pain-related fear avoidance, anxiety, and depression, whereas positive SMDs indicate improvement for health-related quality of life and self-efficacy. In the network plots, node size is proportional to the number of participants and line thickness to the number of randomised controlled trials contributing direct evidence; no line indicates no direct comparison. Treatment-node definitions are provided in Supplementary Table 4.1. Mobile-App=mobile applications; VR=virtual reality; Web-App=web applications.

**Figure S50.** Network plots for pain intensity

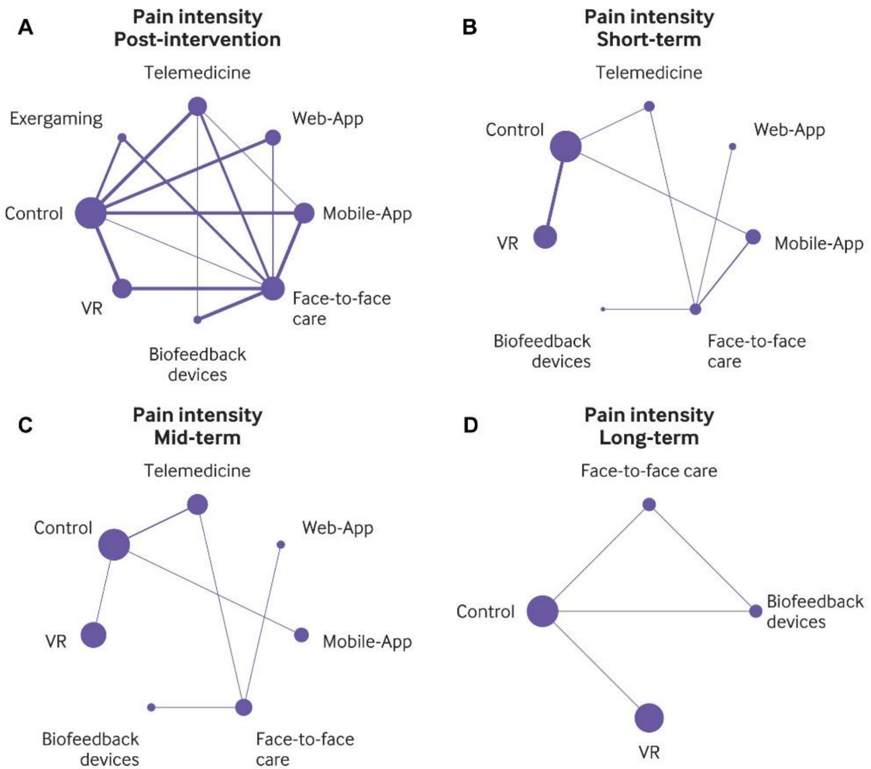

**Figure S51.** Network plots for physical function

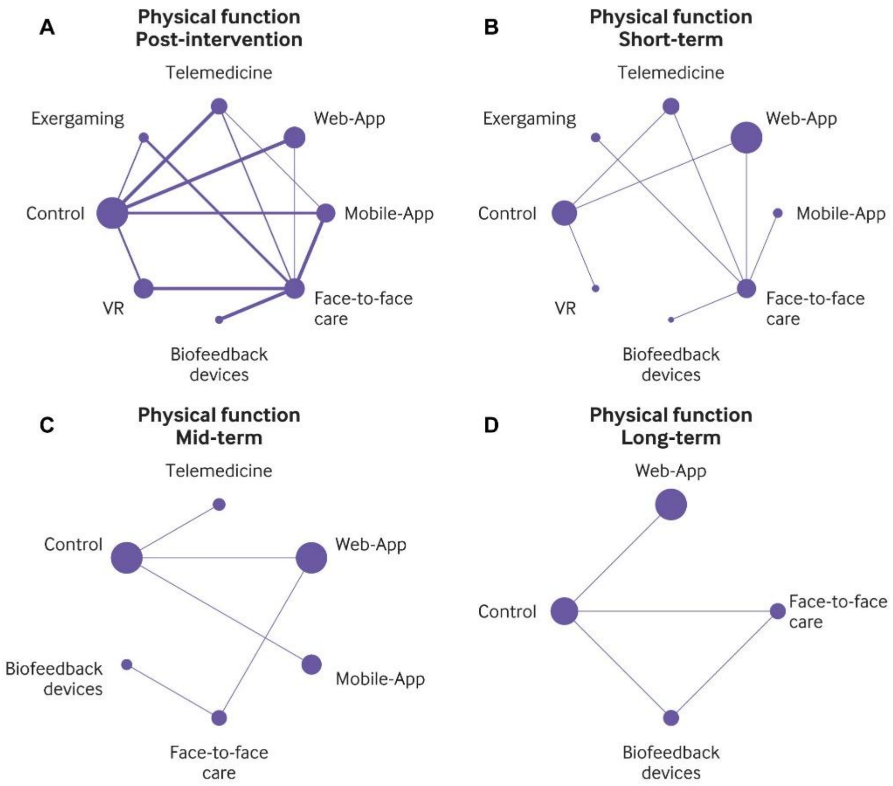

**Figure S52.** Network plots for pain-related fear avoidance

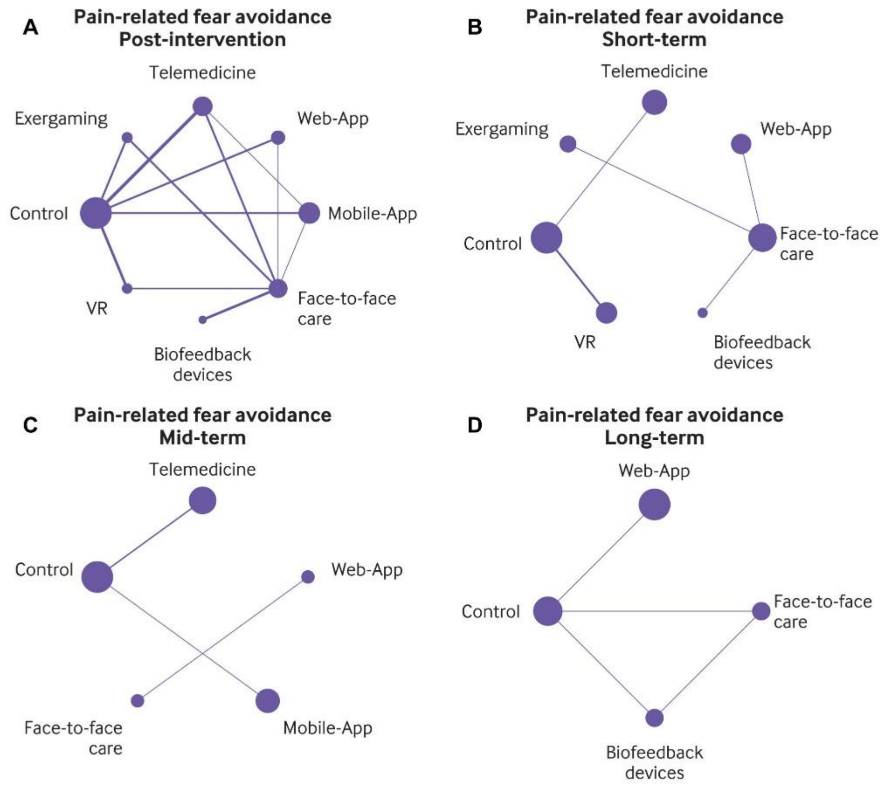

**Figure S53.** Network plots for health-related quality of life

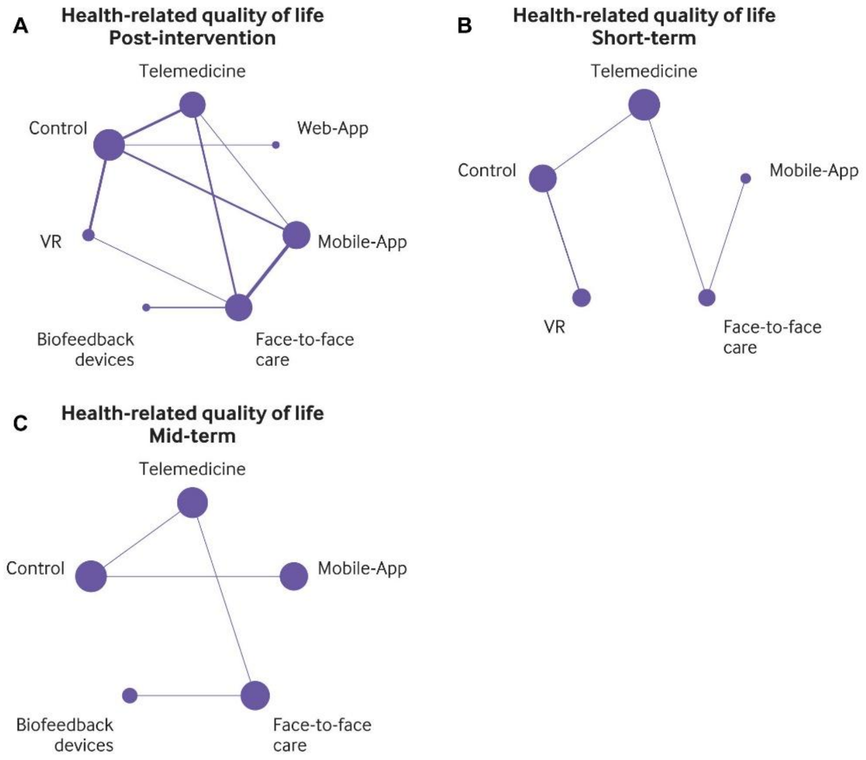

**Figure S54.** Network plots for self-efficacy

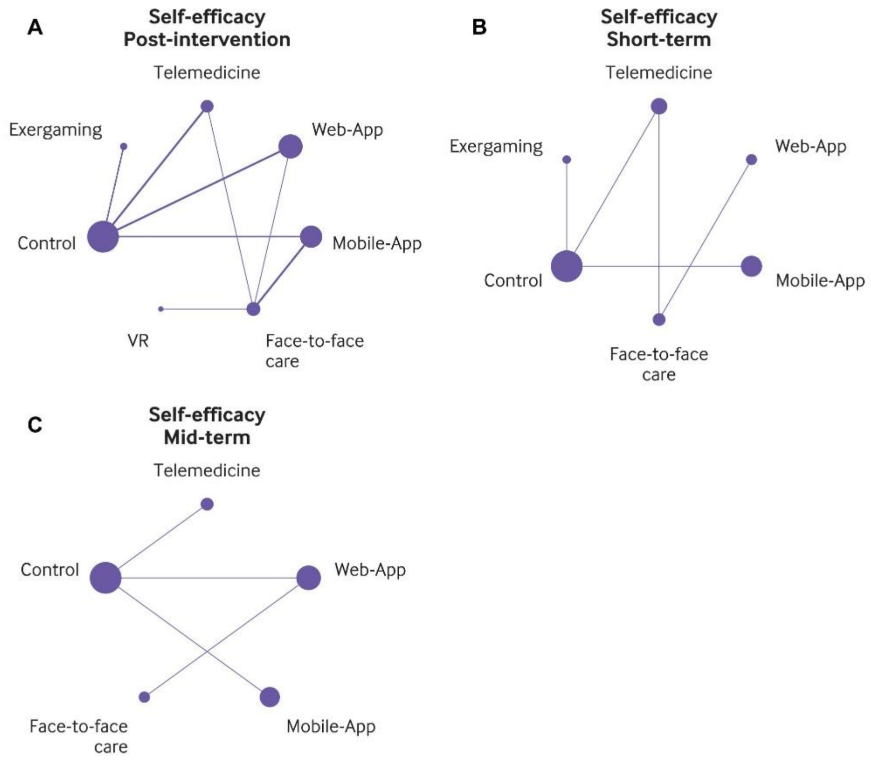

**Figure S55.** Network plots for anxiety

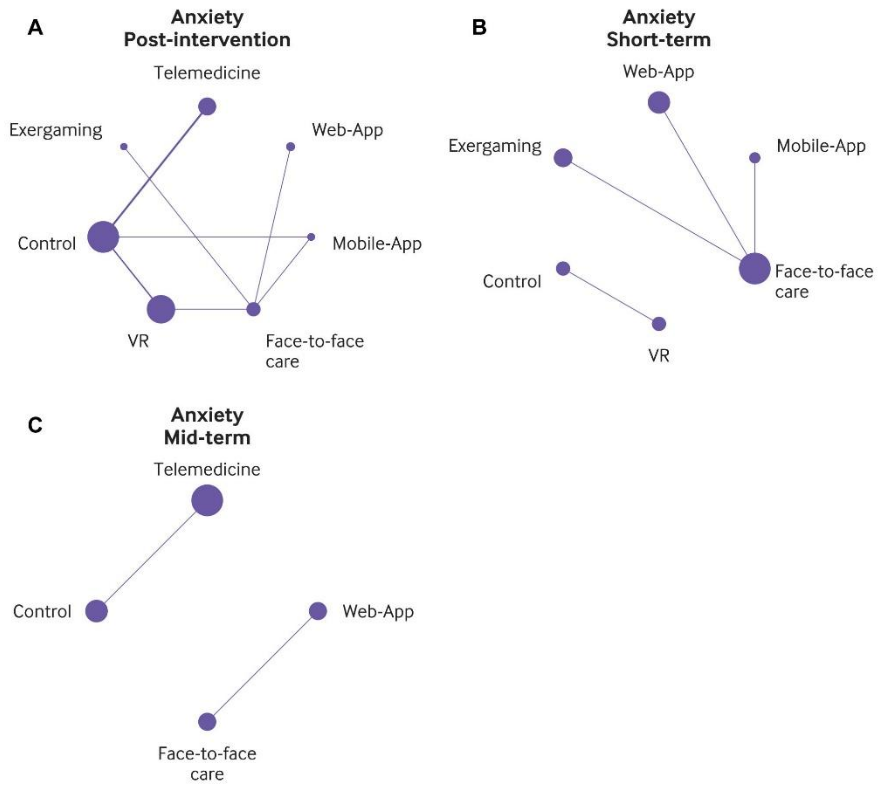

**Figure S56.** Network plots for depression

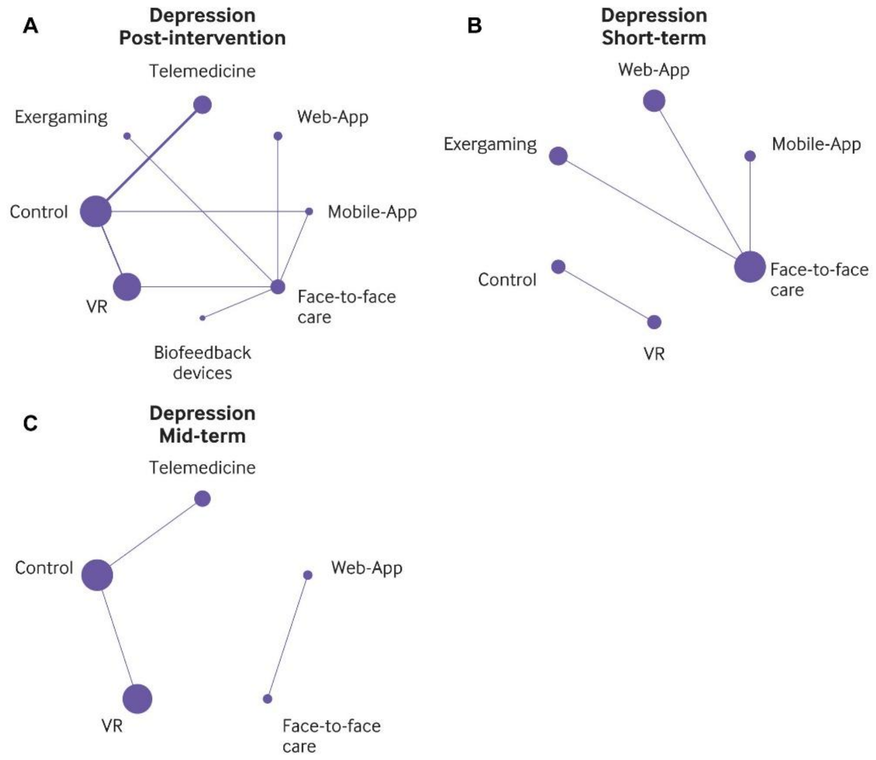

**Figure S57.** Forest plot for pain intensity at post-intervention

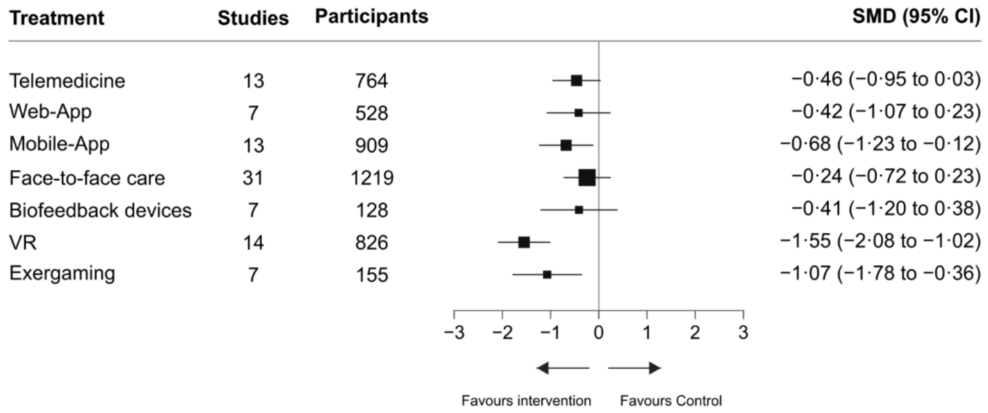

**Figure S58.** Forest plot for pain intensity at short-term

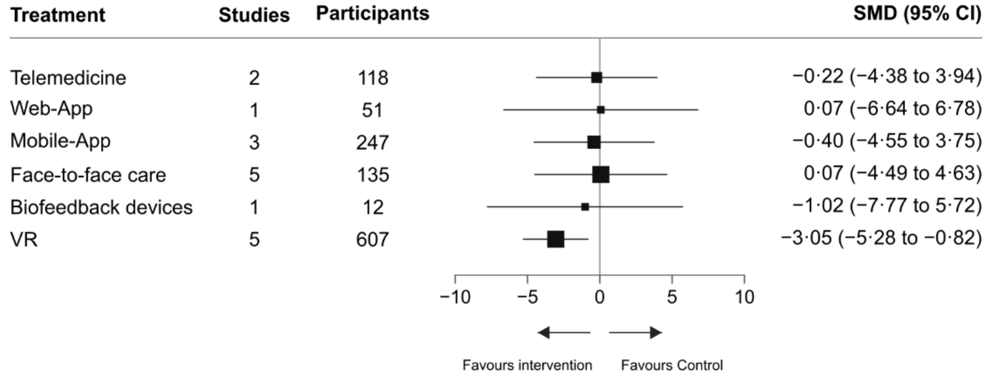

**Figure S59.** Forest plot for pain intensity at mid-term

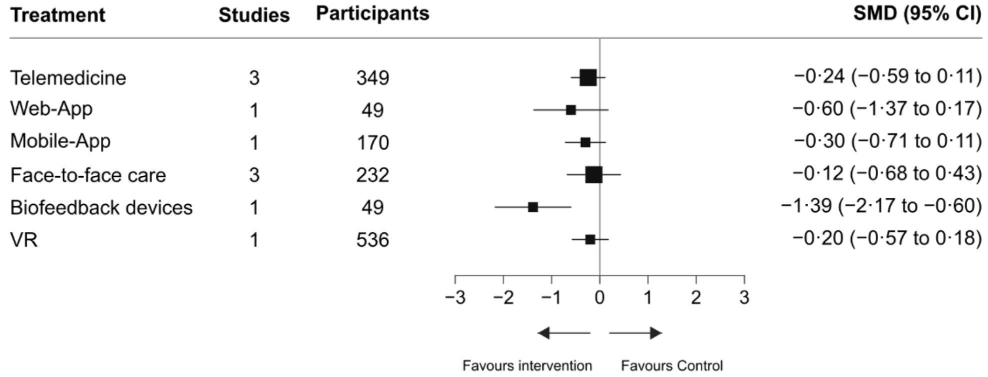

**Figure S60.** Forest plot for pain intensity at long-term

| Treatment | Studies | Participants | SMD (95% CI) |
|---|---|---|---|
| Face-to-face care | 1 | 106 | −0·13 (−0·40 to 0·14) |
| Biofeedback devices | 1 | 104 | 0·00 (−0·27 to 0·27) |
| VR | 1 | 536 | −0·29 (−0·41 to −0·17) |

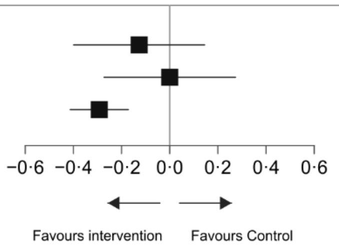

**Figure S61.** Forest plot for physical function at post-intervention

| Treatment | Studies | Participants | SMD (95% CI) |
|---|---|---|---|
| Telemedicine | 11 | 466 | −0·46 (−0·92 to −0·01) |
| Web-App | 8 | 825 | −0·80 (−1·32 to −0·28) |
| Mobile-App | 11 | 625 | −0·32 (−0·87 to 0·23) |
| Face-to-face care | 25 | 730 | 0·03 (−0·49 to 0·55) |
| Biofeedback devices | 6 | 102 | −0·41 (−1·23 to 0·41) |
| VR | 8 | 700 | −0·65 (−1·27 to −0·03) |
| Exergaming | 6 | 164 | −0·26 (−0·97 to 0·45) |

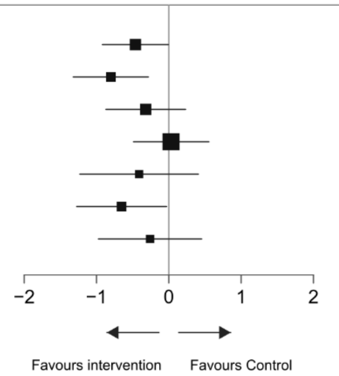

**Figure S62.** Forest plot for physical function at short-term

| Treatment | Studies | Participants | SMD (95% CI) |
|---|---|---|---|
| Telemedicine | 2 | 118 | −0·34 (−0·70 to 0·01) |
| Web-App | 2 | 430 | −0·09 (−0·25 to 0·07) |
| Mobile-App | 1 | 36 | −0·55 (−1·15 to 0·04) |
| Face-to-face care | 5 | 153 | −0·14 (−0·50 to 0·22) |
| Biofeedback devices | 1 | 12 | −0·61 (−1·47 to 0·25) |
| VR | 1 | 20 | −0·22 (−0·84 to 0·40) |
| Exergaming | 1 | 35 | −0·34 (−0·94 to 0·25) |

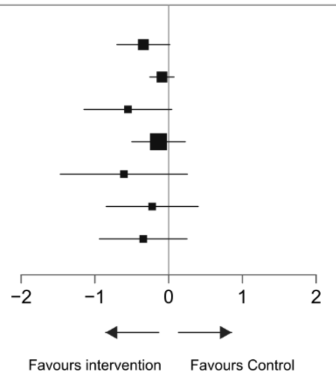

**Figure S63.** Forest plot for physical function at mid-term

| Treatment | Studies | Participants | SMD (95% CI) |
|---|---|---|---|
| Telemedicine | 1 | 64 | −0·41 (−0·84 to 0·01) |
| Web-App | 2 | 421 | −0·08 (−0·25 to 0·08) |
| Mobile-App | 1 | 170 | −0·30 (−0·51 to −0·09) |
| Face-to-face care | 2 | 100 | 0·16 (−0·27 to 0·59) |
| Biofeedback devices | 1 | 49 | −0·73 (−1·32 to −0·13) |

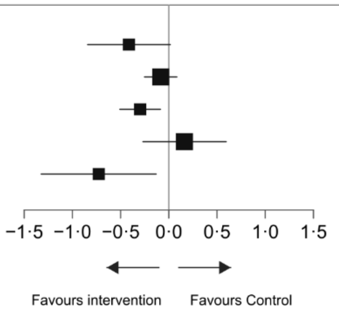

**Figure S64.** Forest plot for physical function at long-term

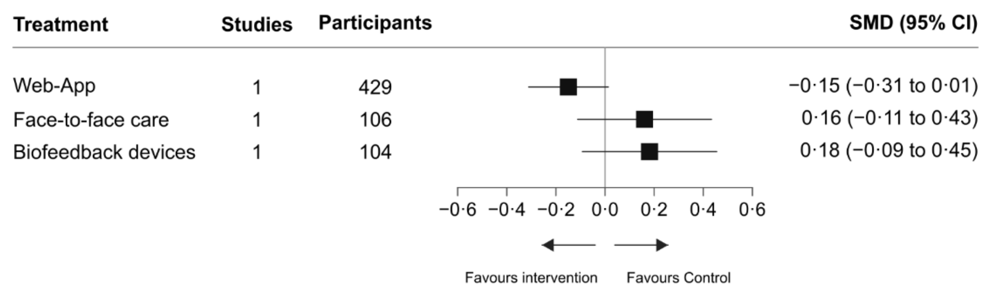

**Figure S65.** Forest plot for pain-related fear avoidance at post-intervention

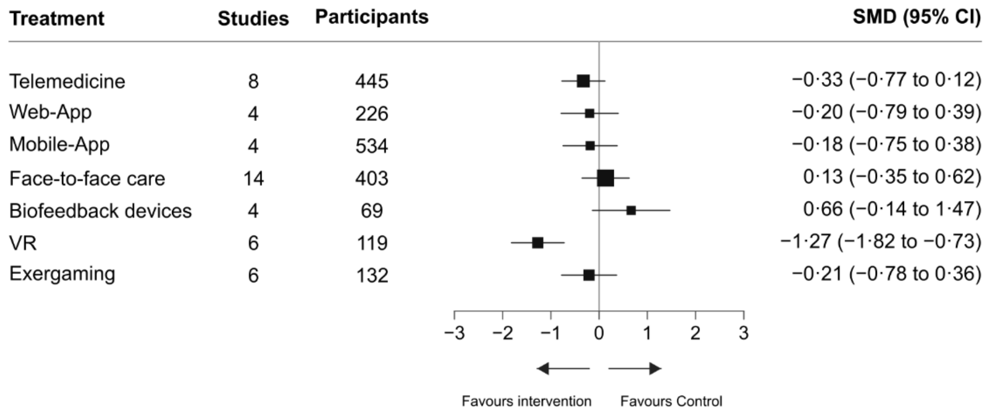

**Figure S66.** Forest plot for pain-related fear avoidance at long-term

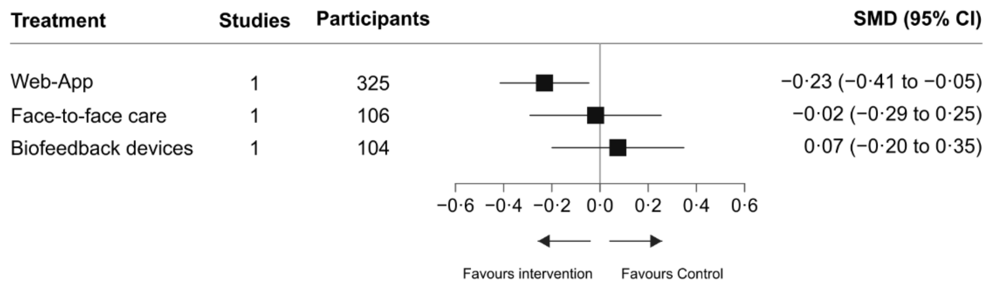

**Figure S67.** Forest plot for health-related quality of life at post-intervention

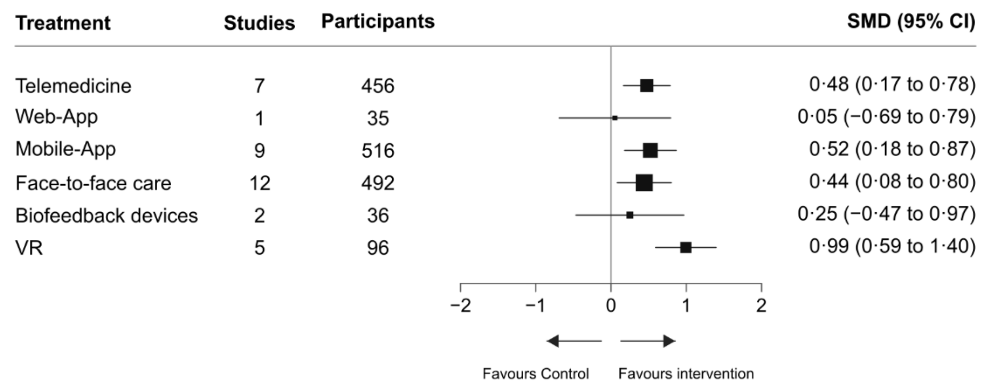

**Figure S68.** Forest plot for health-related quality of life at short-term

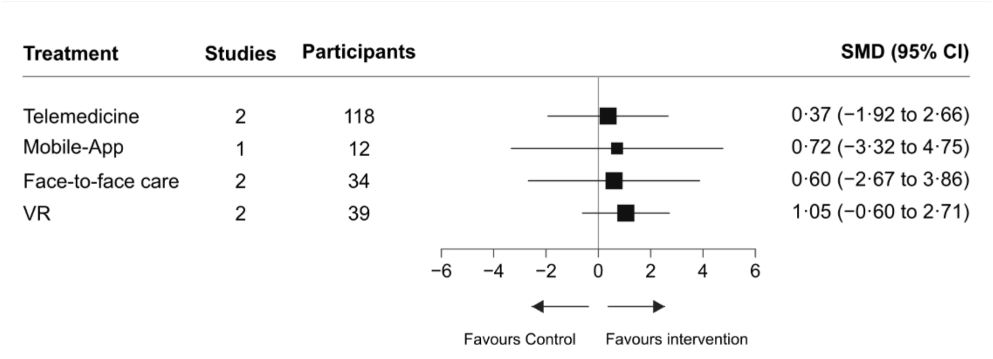

**Figure S69.** Forest plot for health-related quality of life at mid-term

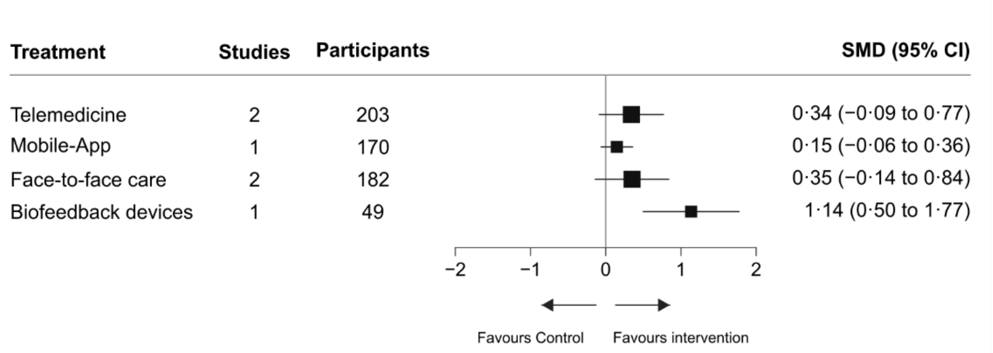

**Figure S70.** Forest plot for self-efficacy at post-intervention

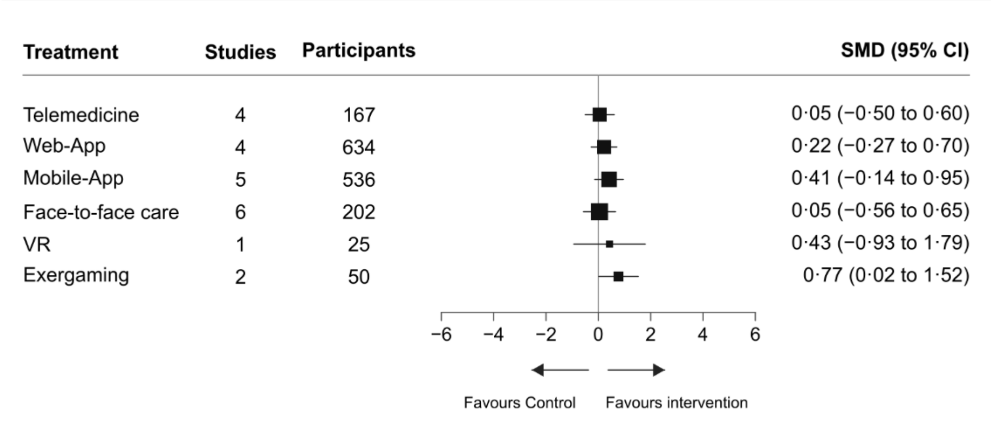

**Figure S71.** Forest plot for self-efficacy at short-term

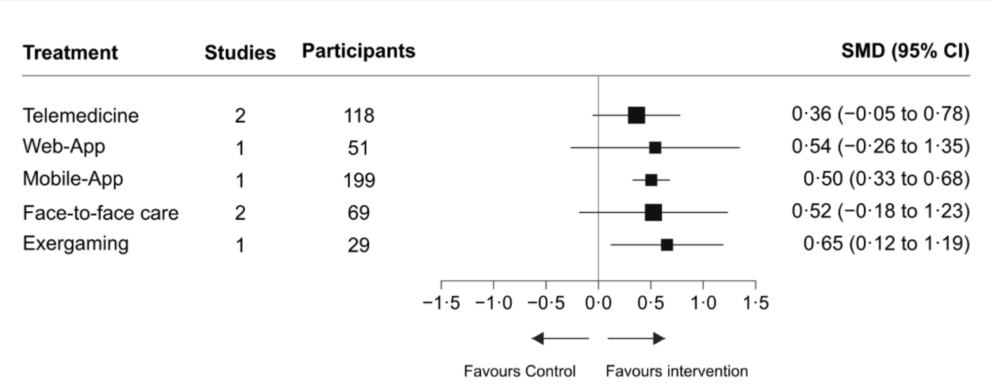

**Figure S72.** Forest plot for self-efficacy at mid-term

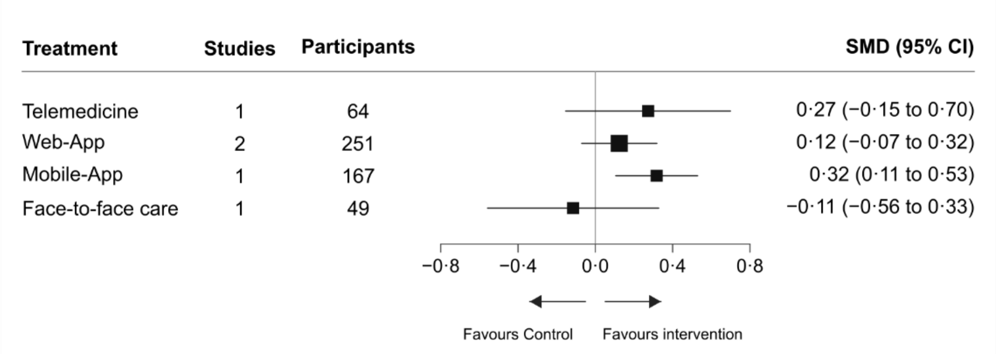

**Figure S73.** Forest plot for anxiety at post-intervention

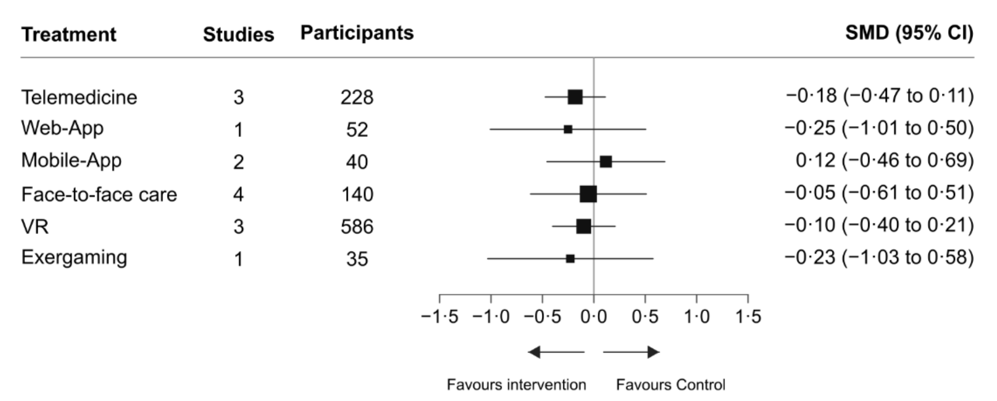

**Figure S74.** Forest plot for depression at post-intervention

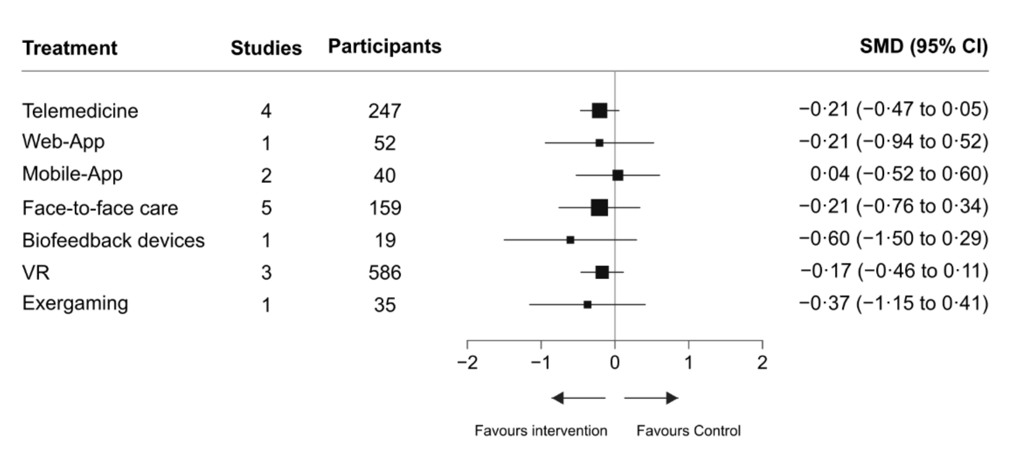

**Section 17.** Additional analyses restricted to participants with chronic low back pain

These analyses were restricted to trials enrolling participants with chronic low back pain, defined as symptoms lasting at least 12 weeks. Network meta-analysis estimates are presented as standardised mean differences (SMDs) with 95% confidence intervals (CIs) from random-effects models. Negative SMDs indicate improvement for pain intensity, physical function, pain-related fear avoidance, anxiety, and depression, whereas positive SMDs indicate improvement for health-related quality of life and self-efficacy. In the network plots, node size is proportional to the number of participants and line thickness to the number of randomised controlled trials contributing direct evidence; no line indicates no direct comparison. Treatment-node definitions are provided in Supplementary Table 4.1. Mobile-App=mobile applications; VR=virtual reality; Web-App=web applications.

**Figure S75.** Network plots for pain intensity

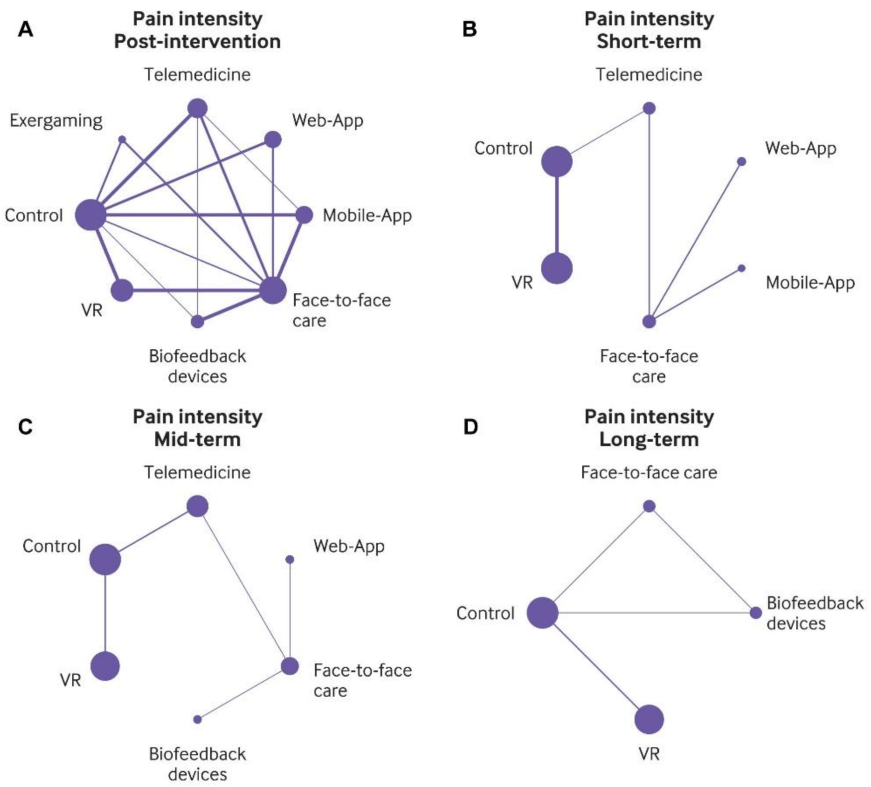

**Figure S76.** Network plots for physical function

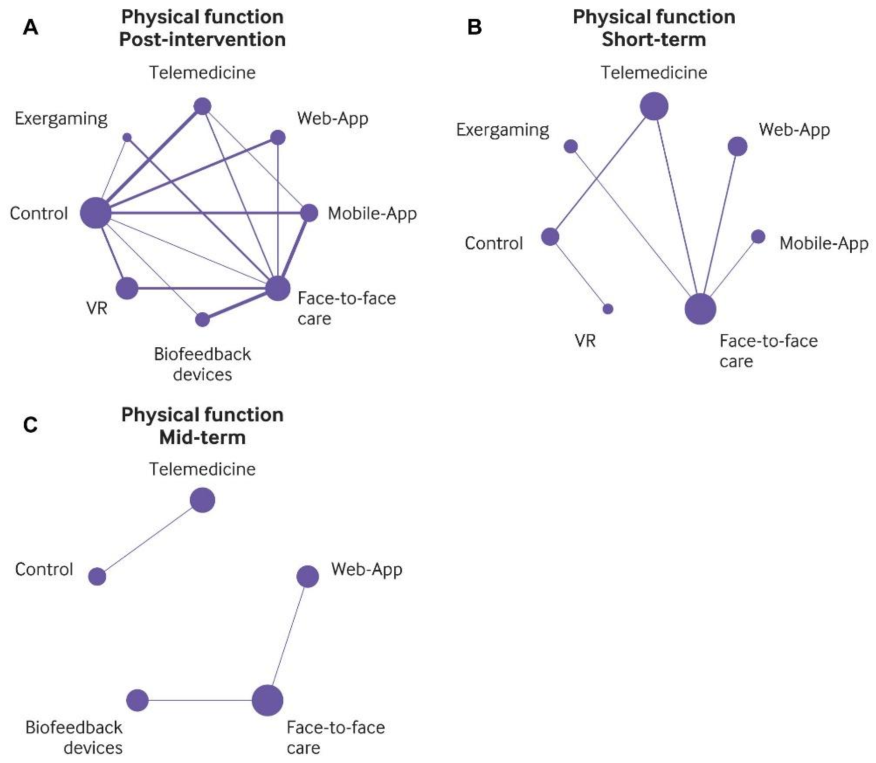

**Figure S77.** Network plots for pain-related fear avoidance

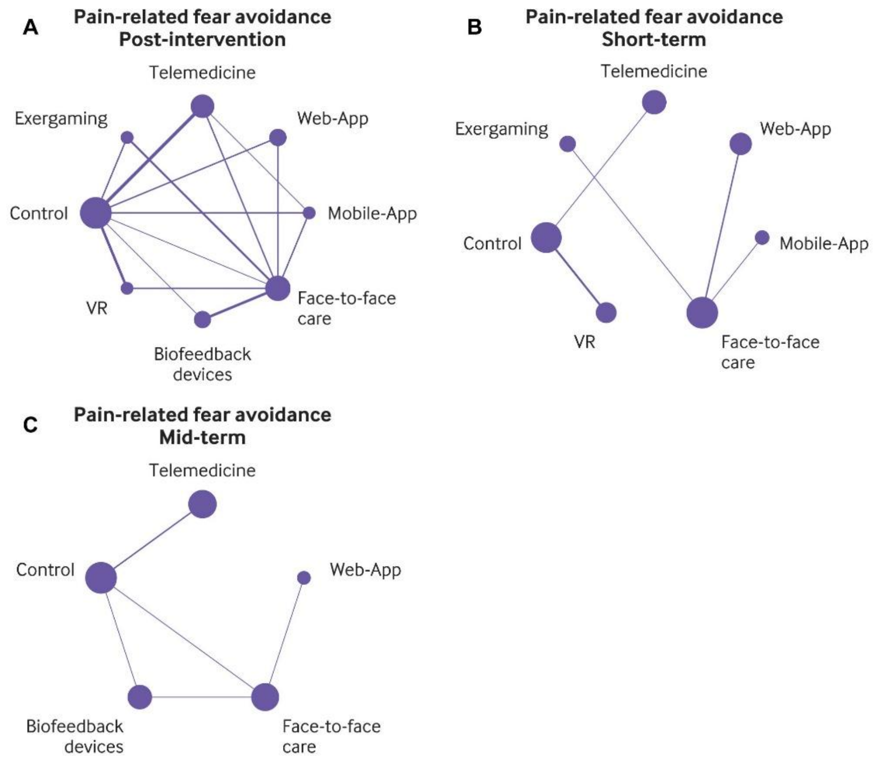

**Figure S78.** Network plots for health-related quality of life

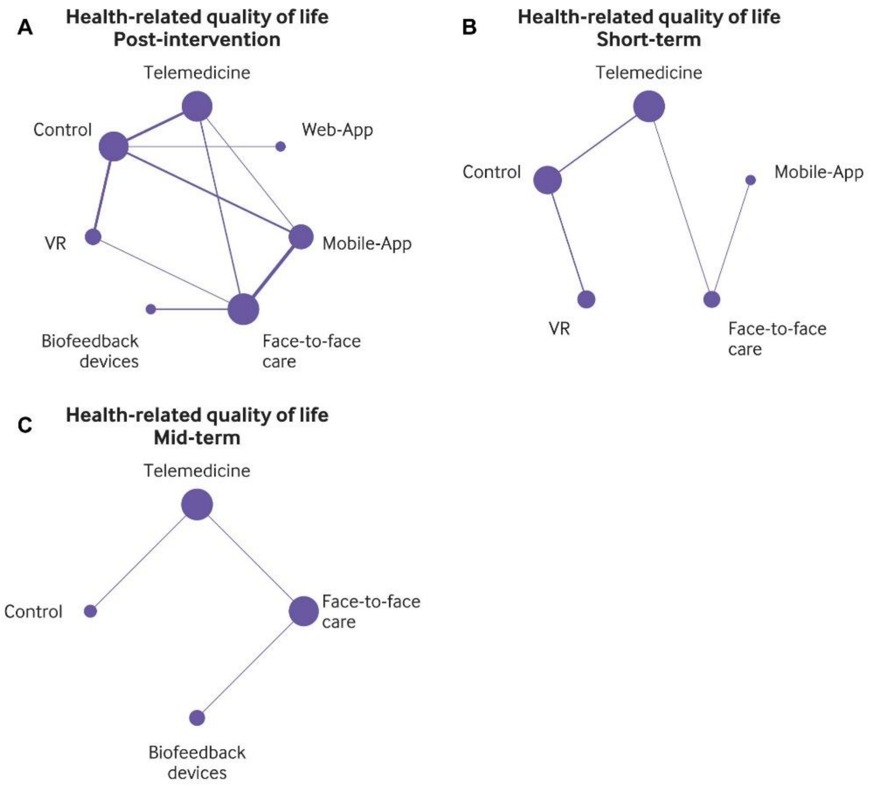

**Figure S79.** Network plots for self-efficacy

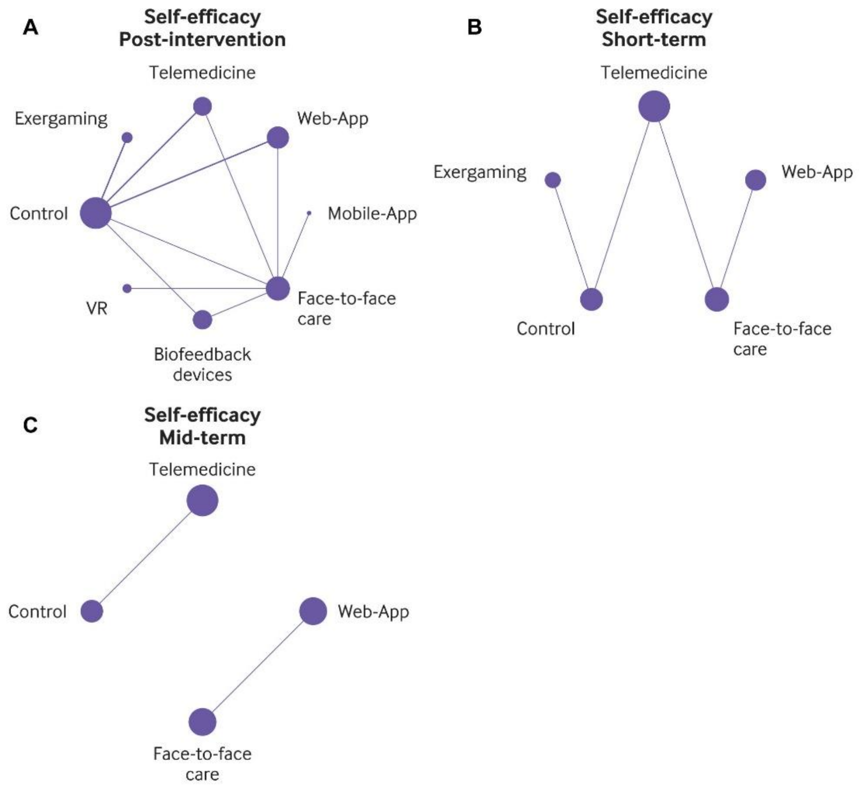

**Figure S80.** Network plots for anxiety

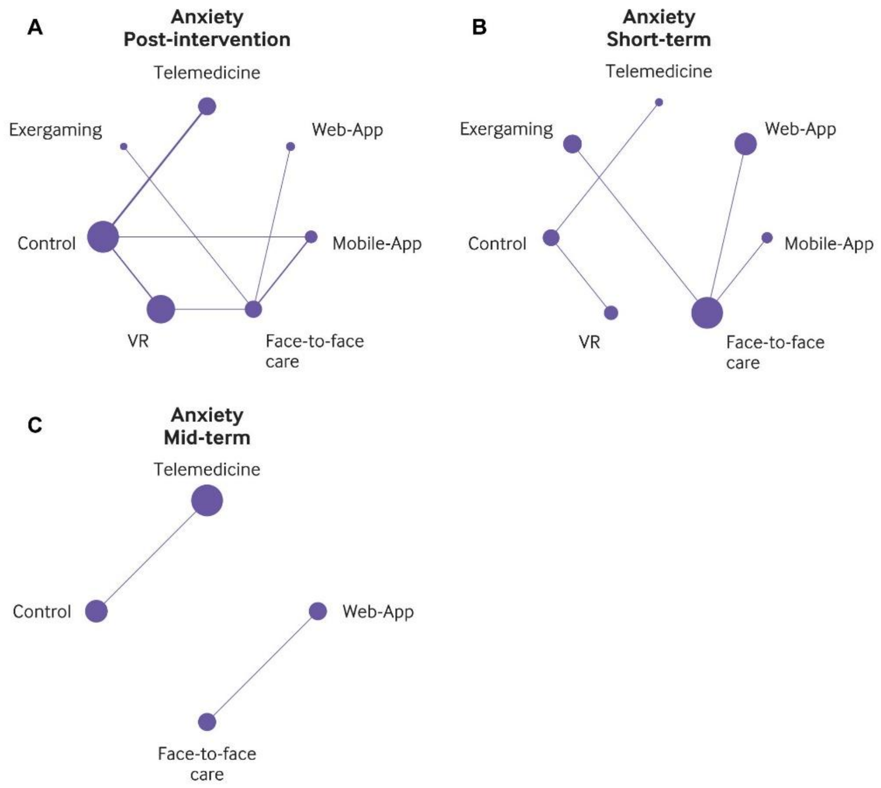

**Figure S81.** Network plots for depression

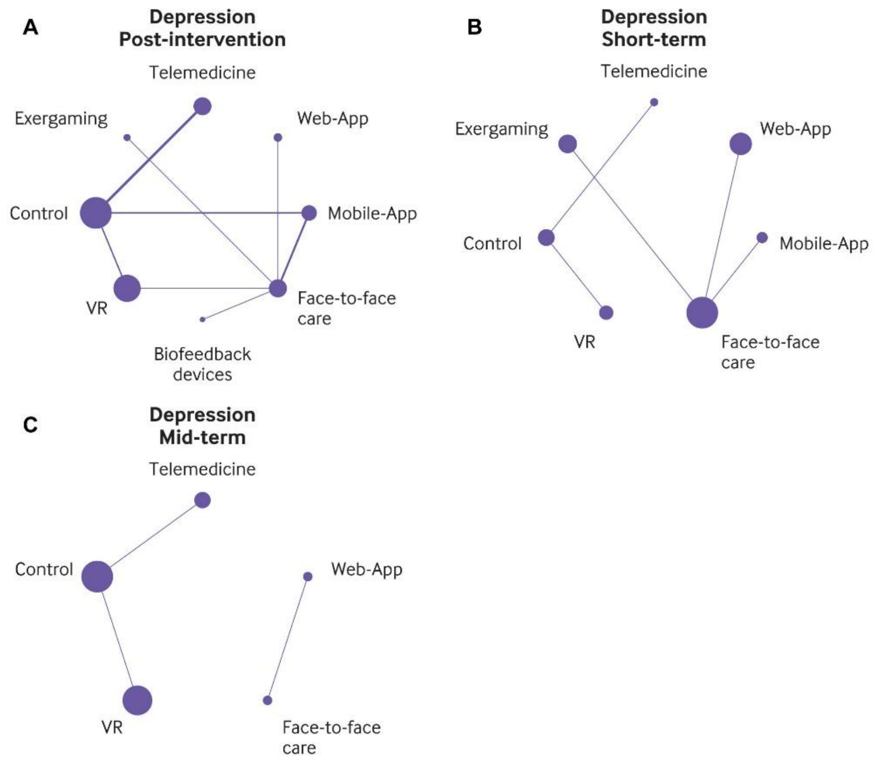

**Figure S82.** Forest plot for pain intensity at post-intervention

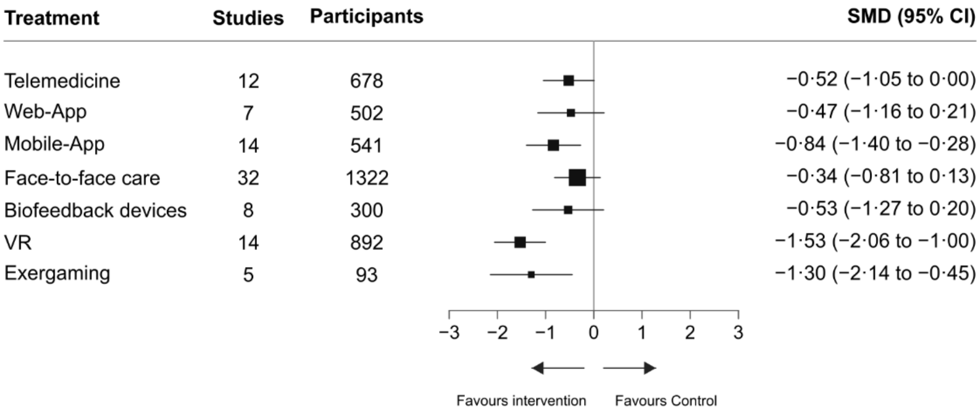

**Figure S83.** Forest plot for pain intensity at short-term

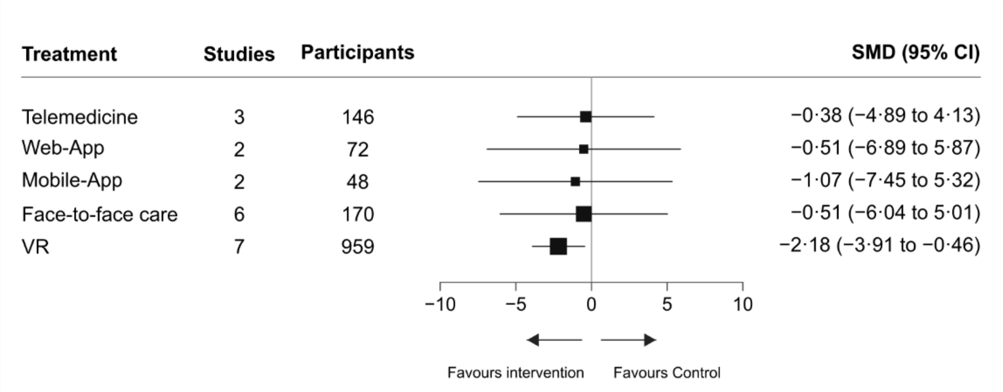

**Figure S84.** Forest plot for pain intensity at mid-term

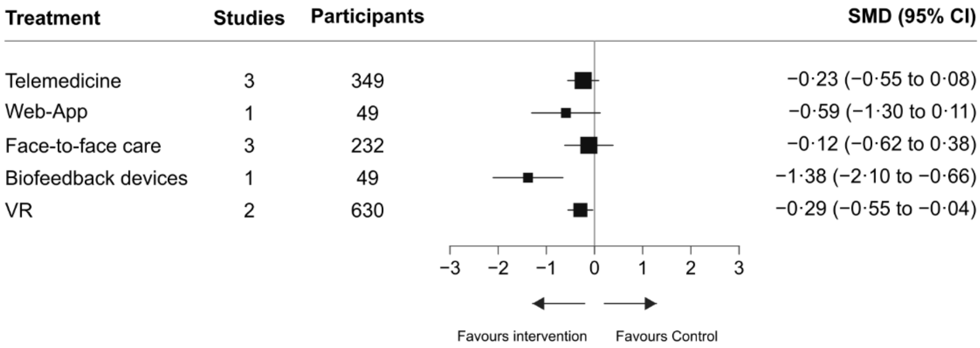

**Figure S85.** Forest plot for pain intensity at long-term

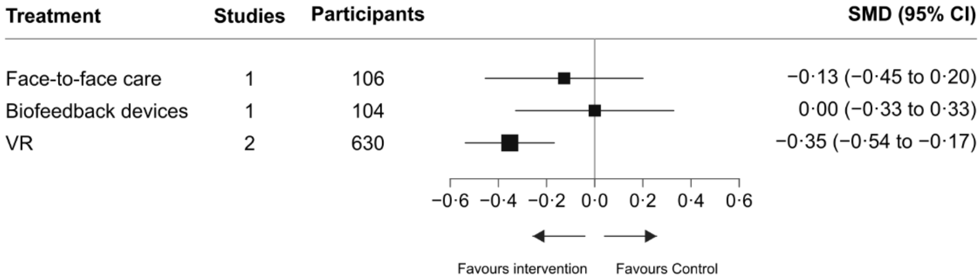

**Figure S86.** Forest plot for physical function at post-intervention

| Treatment | Studies | Participants | SMD (95% CI) |
|---|---|---|---|
| Telemedicine | 10 | 402 | −0·46 (−0·94 to 0·02) |
| Web-App | 6 | 301 | −0·61 (−1·23 to 0·01) |
| Mobile-App | 12 | 429 | −0·82 (−1·36 to −0·28) |
| Face-to-face care | 26 | 858 | −0·45 (−0·94 to 0·04) |
| Biofeedback devices | 7 | 274 | −0·80 (−1·51 to −0·09) |
| VR | 7 | 672 | −0·93 (−1·56 to −0·30) |
| Exergaming | 4 | 102 | −0·55 (−1·39 to 0·29) |

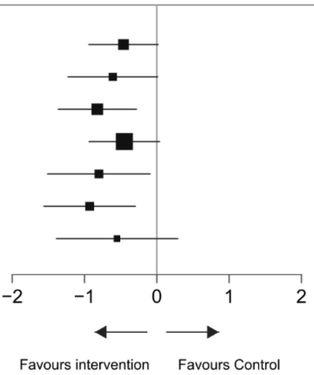

**Figure S87.** Forest plot for physical function at short-term

| Treatment | Studies | Participants | SMD (95% CI) |
|---|---|---|---|
| Telemedicine | 4 | 152 | −0·34 (−0·72 to 0·05) |
| Web-App | 2 | 72 | −0·12 (−0·76 to 0·51) |
| Mobile-App | 1 | 36 | −0·55 (−1·27 to 0·17) |
| Face-to-face care | 6 | 188 | −0·13 (−0·68 to 0·41) |
| VR | 1 | 20 | −0·22 (−0·84 to 0·40) |
| Exergaming | 1 | 35 | −0·34 (−1·06 to 0·38) |

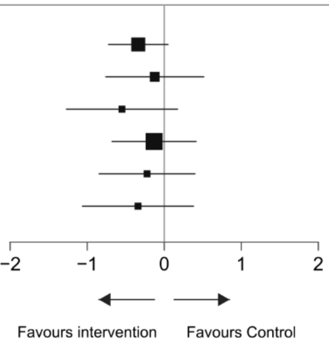

**Figure S88.** Forest plot for pain-related fear avoidance at post-intervention

| Treatment | Studies | Participants | SMD (95% CI) |
|---|---|---|---|
| Telemedicine | 7 | 421 | −0·47 (−0·99 to 0·06) |
| Web-App | 4 | 223 | −0·30 (−1·01 to 0·41) |
| Mobile-App | 4 | 119 | −0·58 (−1·28 to 0·13) |
| Face-to-face care | 15 | 488 | −0·39 (−0·90 to 0·13) |
| Biofeedback devices | 4 | 220 | −0·14 (−0·92 to 0·65) |
| VR | 6 | 116 | −1·05 (−1·66 to −0·44) |
| Exergaming | 5 | 120 | −0·58 (−1·28 to 0·11) |

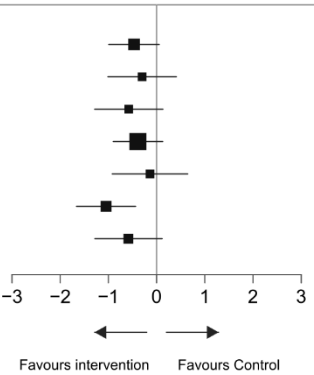

**Figure S89.** Forest plot for pain-related fear avoidance at mid-term

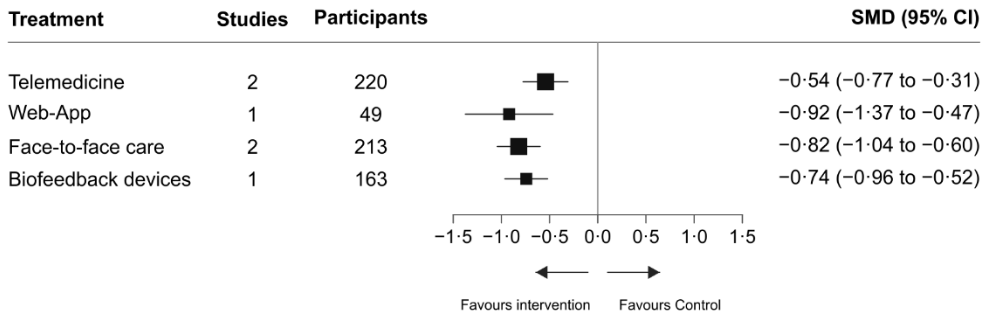

**Figure S90.** Forest plot for health-related quality of life at post-intervention

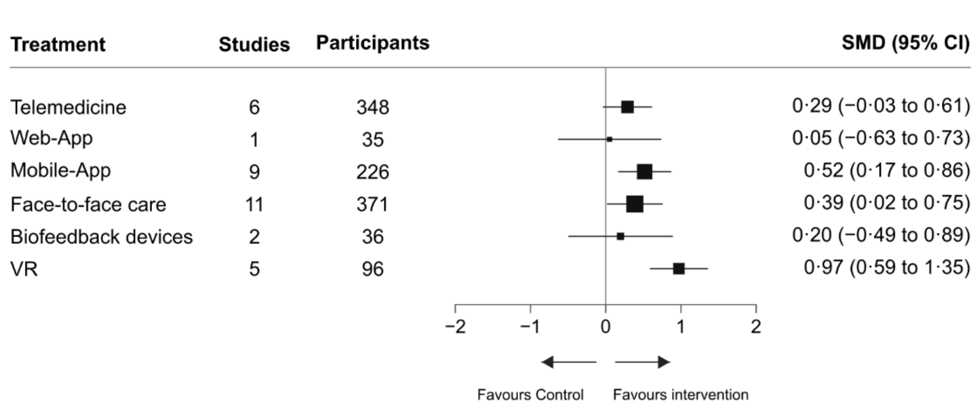

**Figure S91.** Forest plot for health-related quality of life at short-term

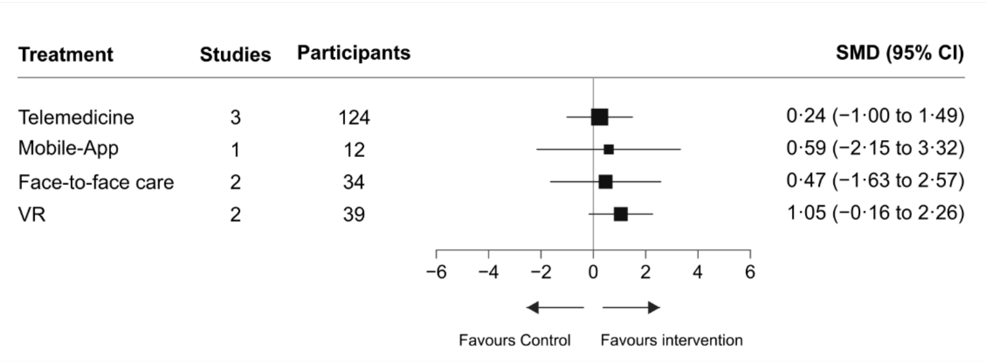

**Figure S92.** Forest plot for health-related quality of life at mid-term

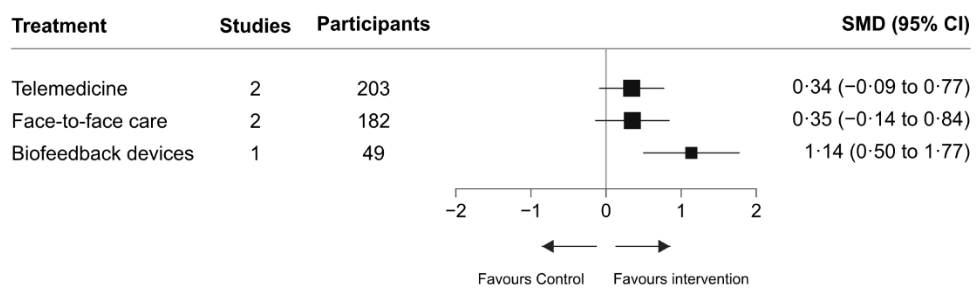

**Figure S93.** Forest plot for self-efficacy at post-intervention

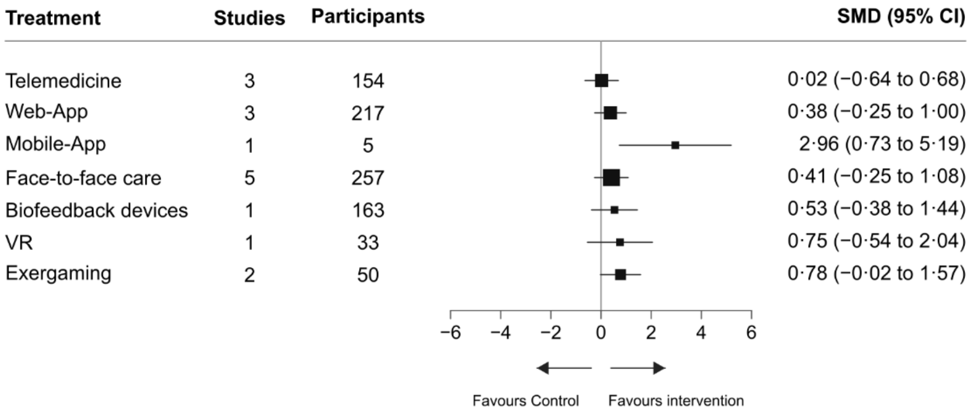

**Figure S94.** Forest plot for self-efficacy at short-term

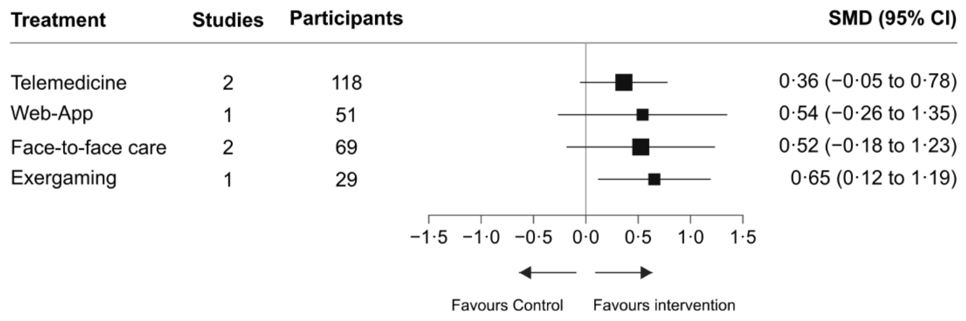

**Figure S95.** Forest plot for anxiety at post-intervention

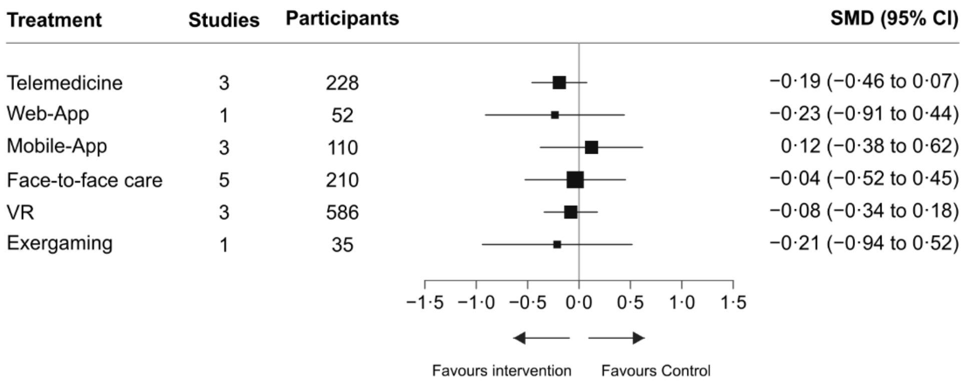

**Figure S96.** Forest plot for depression at post-intervention

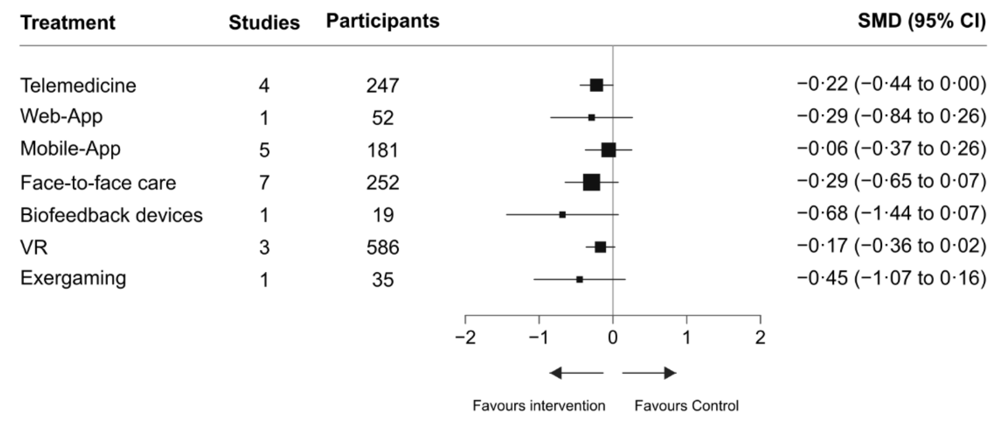

**Section 18.** Confidence in network meta-analysis (CINeMA) assessment of confidence in the network evidence

We assessed confidence in network estimates using the Confidence in Network Meta-Analysis (CINeMA) framework across six domains: within-study bias, reporting bias, indirectness, imprecision, heterogeneity, and incoherence. Each domain was rated as no concerns, some concerns, or major concerns. We derived overall confidence ratings according to CINeMA guidance. Mobile-App, mobile applications; VR, virtual reality; Web-App, web applications.

**Table S45.** Pain intensity at post-intervention

| Comparison | Number of studies | Within-study bias | Reporting bias | Indirectness | Imprecision | Heterogeneity | Incoherence | Confidence rating |
|---|---|---|---|---|---|---|---|---|
| Biofeedback devices: Control | 1 | Some concerns | Low risk | No concerns | No concerns | Major concerns | No concerns | Very low |
| Biofeedback devices: Face-to-face care | 9 | Some concerns | Low risk | No concerns | Major concerns | No concerns | No concerns | Very low |
| Biofeedback devices: Telemedicine | 1 | Some concerns | Low risk | No concerns | Major concerns | No concerns | No concerns | Very low |
| Control: Exergaming | 4 | Some concerns | Low risk | No concerns | No concerns | Some concerns | Major concerns | Very low |
| Control: Face-to-face care | 3 | Some concerns | Low risk | No concerns | No concerns | Major concerns | No concerns | Very low |
| Control: Mobile-App | 8 | Some concerns | Low risk | No concerns | No concerns | Major concerns | No concerns | Very low |
| Control: Telemedicine | 9 | Some concerns | Low risk | No concerns | No concerns | Major concerns | No concerns | Very low |
| Control: VR | 8 | Some concerns | Low risk | No concerns | No concerns | No concerns | No concerns | **Moderate** |
| Control: Web-App | 6 | Some concerns | Low risk | No concerns | No concerns | Major concerns | No concerns | Very low |
| Exergaming: Face-to-face care | 4 | Some concerns | Low risk | No concerns | No concerns | Major concerns | Major concerns | Very low |
| Face-to-face care: Mobile-App | 11 | Some concerns | Low risk | No concerns | No concerns | Major concerns | No concerns | Very low |
| Face-to-face care: Telemedicine | 5 | Some concerns | Low risk | No concerns | Some concerns | Some concerns | No concerns | Very low |
| Face-to-face care: VR | 8 | Some concerns | Low risk | No concerns | No concerns | Some concerns | No concerns | Very low |
| Face-to-face care: Web-App | 4 | Some concerns | Low risk | No concerns | Some concerns | Some concerns | No concerns | Very low |
| Mobile-App: Telemedicine | 1 | Some concerns | Low risk | No concerns | Major concerns | No concerns | No concerns | Very low |
| Biofeedback devices: Exergaming | 0 | Some concerns | Low risk | No concerns | Some concerns | Some concerns | Major concerns | Very low |
| Biofeedback devices: Mobile-App | 0 | Some concerns | Low risk | No concerns | Major concerns | No concerns | Major concerns | Very low |
| Biofeedback devices: VR | 0 | Some concerns | Low risk | No concerns | No concerns | Major concerns | Major concerns | Very low |
| Biofeedback devices: Web-App | 0 | Some concerns | Low risk | No concerns | Major concerns | No concerns | Major concerns | Very low |
| Exergaming: Mobile-App | 0 | Some concerns | Low risk | No concerns | Some concerns | Some concerns | Major concerns | Very low |

| | | | | | | | | |
|---|---|---|---|---|---|---|---|---|
| Exergaming: Telemedicine | 0 | Some concerns | Low risk | No concerns | Some concerns | Some concerns | Major concerns | Very low |
| Exergaming: VR | 0 | Some concerns | Low risk | No concerns | Major concerns | No concerns | Major concerns | Very low |
| Exergaming: Web-App | 0 | Some concerns | Low risk | No concerns | Some concerns | Some concerns | Major concerns | Very low |
| Mobile-App: VR | 0 | Some concerns | Low risk | No concerns | No concerns | Major concerns | Major concerns | Very low |
| Mobile-App: Web-App | 0 | Some concerns | Low risk | No concerns | Major concerns | No concerns | Major concerns | Very low |
| Telemedicine: VR | 0 | Some concerns | Low risk | No concerns | No concerns | Major concerns | Major concerns | Very low |
| Telemedicine: Web-App | 0 | Some concerns | Low risk | No concerns | Major concerns | No concerns | Major concerns | Very low |
| VR: Web-App | 0 | Some concerns | Low risk | No concerns | No concerns | Major concerns | Major concerns | Very low |

**Table S46.** Pain intensity at short-term

| Comparison | Number of studies | Within-study bias | Reporting bias | Indirectness | Imprecision | Heterogeneity | Incoherence | Confidence rating |
|---|---|---|---|---|---|---|---|---|
| Biofeedback devices: Face-to-face care | 1 | Major concerns | Low risk | No concerns | Major concerns | No concerns | Major concerns | Very low |
| Control: Mobile-App | 1 | Some concerns | Low risk | No concerns | Major concerns | No concerns | No concerns | Very low |
| Control: Telemedicine | 1 | Major concerns | Low risk | No concerns | Major concerns | No concerns | No concerns | Very low |
| Control: VR | 7 | Some concerns | Low risk | No concerns | No concerns | Some concerns | Major concerns | Very low |
| Face-to-face care: Mobile-App | 2 | Some concerns | Low risk | No concerns | Major concerns | No concerns | No concerns | Very low |
| Face-to-face care: Telemedicine | 2 | Some concerns | Low risk | No concerns | Major concerns | No concerns | No concerns | Very low |
| Face-to-face care: Web-App | 2 | Some concerns | Low risk | No concerns | Major concerns | No concerns | Major concerns | Very low |
| Biofeedback devices: Control | 0 | Major concerns | Low risk | No concerns | Major concerns | No concerns | Major concerns | Very low |
| Biofeedback devices: Mobile-App | 0 | Major concerns | Low risk | No concerns | Major concerns | No concerns | Major concerns | Very low |
| Biofeedback devices: Telemedicine | 0 | Major concerns | Low risk | No concerns | Major concerns | No concerns | Major concerns | Very low |
| Biofeedback devices: VR | 0 | Some concerns | Low risk | No concerns | Major concerns | No concerns | Major concerns | Very low |
| Biofeedback devices: Web-App | 0 | Major concerns | Low risk | No concerns | Major concerns | No concerns | Major concerns | Very low |
| Control: Face-to-face care | 0 | Some concerns | Low risk | No concerns | Major concerns | No concerns | Major concerns | Very low |
| Control: Web-App | 0 | Some concerns | Low risk | No concerns | Major concerns | No concerns | Major concerns | Very low |
| Face-to-face care: VR | 0 | Some concerns | Low risk | No concerns | No concerns | Major concerns | Major concerns | Very low |

| | | | | | | | | |
|---|---|---|---|---|---|---|---|---|
| Mobile-App: Telemedicine | 0 | Some concerns | Low risk | No concerns | Major concerns | No concerns | Major concerns | Very low |
| Mobile-App: VR | 0 | Some concerns | Low risk | No concerns | Major concerns | No concerns | Major concerns | Very low |
| Mobile-App: Web-App | 0 | Some concerns | Low risk | No concerns | Major concerns | No concerns | Major concerns | Very low |
| Telemedicine: VR | 0 | Some concerns | Low risk | No concerns | No concerns | Major concerns | Major concerns | Very low |
| Telemedicine: Web-App | 0 | Some concerns | Low risk | No concerns | Major concerns | No concerns | Major concerns | Very low |
| VR: Web-App | 0 | Some concerns | Low risk | No concerns | Major concerns | No concerns | Major concerns | Very low |

**Table S47.** Physical function at post-intervention

| Comparison | Number of studies | Within-study bias | Reporting bias | Indirectness | Imprecision | Heterogeneity | Incoherence | Confidence rating |
|---|---|---|---|---|---|---|---|---|
| Biofeedback devices: Control | 1 | Some concerns | Low risk | No concerns | No concerns | Major concerns | No concerns | Very low |
| Biofeedback devices: Face-to-face care | 8 | Some concerns | Low risk | No concerns | Some concerns | Some concerns | No concerns | Low |
| Control: Exergaming | 2 | Some concerns | Low risk | No concerns | Some concerns | Some concerns | No concerns | Low |
| Control: Face-to-face care | 2 | Some concerns | Low risk | No concerns | Some concerns | Some concerns | No concerns | Low |
| Control: Mobile-App | 5 | Some concerns | Low risk | No concerns | No concerns | Major concerns | Major concerns | Very low |
| Control: Telemedicine | 10 | Some concerns | Low risk | No concerns | No concerns | Major concerns | No concerns | Very low |
| Control: VR | 3 | Some concerns | Low risk | No concerns | No concerns | Major concerns | Major concerns | Very low |
| Control: Web-App | 8 | Some concerns | Low risk | No concerns | No concerns | Major concerns | No concerns | Very low |
| Exergaming: Face-to-face care | 4 | Some concerns | Low risk | No concerns | Major concerns | No concerns | No concerns | Very low |
| Face-to-face care: Mobile-App | 10 | Some concerns | Low risk | No concerns | No concerns | Major concerns | Some concerns | Very low |
| Face-to-face care: Telemedicine | 3 | Some concerns | Low risk | No concerns | Major concerns | No concerns | No concerns | Very low |
| Face-to-face care: VR | 6 | Some concerns | Low risk | No concerns | No concerns | Major concerns | Major concerns | Very low |
| Face-to-face care: Web-App | 3 | Some concerns | Low risk | No concerns | Some concerns | Some concerns | No concerns | Low |
| Mobile-App: Telemedicine | 1 | Some concerns | Low risk | No concerns | Major concerns | No concerns | No concerns | Very low |
| Biofeedback devices: Exergaming | 0 | Some concerns | Low risk | No concerns | Major concerns | No concerns | Major concerns | Very low |
| Biofeedback devices: Mobile-App | 0 | Some concerns | Low risk | No concerns | Major concerns | No concerns | Major concerns | Very low |
| Biofeedback devices: Telemedicine | 0 | Some concerns | Low risk | No concerns | Major concerns | No concerns | Major concerns | Very low |
| Biofeedback devices: VR | 0 | Some concerns | Low risk | No concerns | Major concerns | No concerns | Major concerns | Very low |

| | | | | | | | | |
|---|---|---|---|---|---|---|---|---|
| Biofeedback devices: Web-App | 0 | Some concerns | Low risk | No concerns | Major concerns | No concerns | Major concerns | Very low |
| Exergaming: Mobile-App | 0 | Some concerns | Low risk | No concerns | Major concerns | No concerns | Major concerns | Very low |
| Exergaming: Telemedicine | 0 | Some concerns | Low risk | No concerns | Major concerns | No concerns | Major concerns | Very low |
| Exergaming: VR | 0 | Some concerns | Low risk | No concerns | Major concerns | No concerns | Major concerns | Very low |
| Exergaming: Web-App | 0 | Some concerns | Low risk | No concerns | Major concerns | No concerns | Major concerns | Very low |
| Mobile-App: VR | 0 | Some concerns | Low risk | No concerns | Major concerns | No concerns | Major concerns | Very low |
| Mobile-App: Web-App | 0 | Some concerns | Low risk | No concerns | Major concerns | No concerns | Major concerns | Very low |
| Telemedicine: VR | 0 | Some concerns | Low risk | No concerns | Major concerns | No concerns | Major concerns | Very low |
| Telemedicine: Web-App | 0 | Some concerns | Low risk | No concerns | Major concerns | No concerns | Major concerns | Very low |
| VR: Web-App | 0 | Some concerns | Low risk | No concerns | Major concerns | No concerns | Major concerns | Very low |

**Table S48.** Physical function at short-term

| Comparison | Number of studies | Within-study bias | Reporting bias | Indirectness | Imprecision | Heterogeneity | Incoherence | Confidence rating |
|---|---|---|---|---|---|---|---|---|
| Biofeedback devices: Face-to-face care | 1 | Major concerns | Low risk | No concerns | Major concerns | No concerns | No concerns | Very low |
| Control: Telemedicine | 2 | Major concerns | Low risk | No concerns | No concerns | Some concerns | No concerns | Very low |
| Control: VR | 1 | Some concerns | Low risk | No concerns | Major concerns | No concerns | No concerns | Very low |
| Control: Web-App | 1 | Some concerns | Low risk | No concerns | Some concerns | No concerns | No concerns | Low |
| Exergaming: Face-to-face care | 1 | Some concerns | Low risk | No concerns | Major concerns | No concerns | No concerns | Very low |
| Face-to-face care: Mobile-App | 1 | No concerns | Low risk | No concerns | Some concerns | Some concerns | No concerns | Low |
| Face-to-face care: Telemedicine | 2 | Some concerns | Low risk | No concerns | Some concerns | No concerns | No concerns | Low |
| Face-to-face care: Web-App | 3 | Some concerns | Low risk | No concerns | Major concerns | No concerns | No concerns | Very low |
| Biofeedback devices: Control | 0 | Some concerns | Low risk | No concerns | Major concerns | No concerns | No concerns | Very low |
| Biofeedback devices: Exergaming | 0 | Major concerns | Low risk | No concerns | Major concerns | No concerns | No concerns | Very low |
| Biofeedback devices: Mobile-App | 0 | Some concerns | Low risk | No concerns | Major concerns | No concerns | No concerns | Very low |
| Biofeedback devices: Telemedicine | 0 | Some concerns | Low risk | No concerns | Major concerns | No concerns | No concerns | Very low |
| Biofeedback devices: VR | 0 | Some concerns | Low risk | No concerns | Major concerns | No concerns | No concerns | Very low |

| | | | | | | | | |
|---|---|---|---|---|---|---|---|---|
| Biofeedback devices: Web-App | 0 | Major concerns | Low risk | No concerns | Major concerns | No concerns | No concerns | Very low |
| Control: Exergaming | 0 | Some concerns | Low risk | No concerns | Major concerns | No concerns | No concerns | Very low |
| Control: Face-to-face care | 0 | Some concerns | Low risk | No concerns | Some concerns | Some concerns | No concerns | Low |
| Control: Mobile-App | 0 | Some concerns | Low risk | No concerns | Some concerns | No concerns | No concerns | Low |
| Exergaming: Mobile-App | 0 | Some concerns | Low risk | No concerns | Major concerns | No concerns | No concerns | Very low |
| Exergaming: Telemedicine | 0 | Some concerns | Low risk | No concerns | Major concerns | No concerns | No concerns | Very low |
| Exergaming: VR | 0 | Some concerns | Low risk | No concerns | Major concerns | No concerns | No concerns | Very low |
| Exergaming: Web-App | 0 | Some concerns | Low risk | No concerns | Major concerns | No concerns | No concerns | Very low |

**Table S49.** Pain-related fear avoidance at post-intervention

| Comparison | Number of studies | Within-study bias | Reporting bias | Indirectness | Imprecision | Heterogeneity | Incoherence | Confidence rating |
|---|---|---|---|---|---|---|---|---|
| Biofeedback devices: Control | 1 | Some concerns | Low risk | No concerns | Major concerns | No concerns | Major concerns | Very low |
| Biofeedback devices: Face-to-face care | 5 | Some concerns | Low risk | No concerns | Major concerns | No concerns | Some concerns | Very low |
| Control: Exergaming | 3 | Some concerns | Low risk | No concerns | Some concerns | Some concerns | Major concerns | Very low |
| Control: Face-to-face care | 1 | Some concerns | Low risk | No concerns | Some concerns | Some concerns | No concerns | Low |
| Control: Mobile-App | 4 | Some concerns | Low risk | No concerns | Some concerns | Some concerns | No concerns | Low |
| Control: Telemedicine | 5 | Some concerns | Low risk | No concerns | No concerns | Major concerns | No concerns | Very low |
| Control: VR | 4 | Some concerns | Low risk | No concerns | No concerns | No concerns | No concerns | **Moderate** |
| Control: Web-App | 3 | Some concerns | Low risk | No concerns | Major concerns | No concerns | No concerns | Very low |
| Exergaming: Face-to-face care | 3 | Some concerns | Low risk | No concerns | Major concerns | No concerns | Some concerns | Very low |
| Face-to-face care: Mobile-App | 3 | Some concerns | Low risk | No concerns | Major concerns | No concerns | No concerns | Very low |
| Face-to-face care: Telemedicine | 3 | Some concerns | Low risk | No concerns | Some concerns | Some concerns | No concerns | Low |
| Face-to-face care: VR | 3 | Some concerns | Low risk | No concerns | No concerns | Some concerns | No concerns | Low |
| Face-to-face care: Web-App | 2 | Some concerns | Low risk | No concerns | Major concerns | No concerns | No concerns | Very low |
| Mobile-App: Telemedicine | 1 | Some concerns | Low risk | No concerns | Major concerns | No concerns | No concerns | Very low |
| Biofeedback devices: Exergaming | 0 | Some concerns | Low risk | No concerns | Major concerns | No concerns | Major concerns | Very low |
| Biofeedback devices: Mobile-App | 0 | Some concerns | Low risk | No concerns | Major concerns | No concerns | Major concerns | Very low |

| | | | | | | | | |
|---|---|---|---|---|---|---|---|---|
| Biofeedback devices: Telemedicine | 0 | Some concerns | Low risk | No concerns | Some concerns | Some concerns | Major concerns | Very low |
| Biofeedback devices: VR | 0 | Some concerns | Low risk | No concerns | No concerns | No concerns | Major concerns | Very low |
| Biofeedback devices: Web-App | 0 | Some concerns | Low risk | No concerns | Major concerns | No concerns | Major concerns | Very low |
| Exergaming: Mobile-App | 0 | Some concerns | Low risk | No concerns | Major concerns | No concerns | Major concerns | Very low |
| Exergaming: Telemedicine | 0 | Some concerns | Low risk | No concerns | Major concerns | No concerns | Major concerns | Very low |
| Exergaming: VR | 0 | Some concerns | Low risk | No concerns | No concerns | Major concerns | Major concerns | Very low |
| Exergaming: Web-App | 0 | Some concerns | Low risk | No concerns | Major concerns | No concerns | Major concerns | Very low |
| Mobile-App: VR | 0 | Some concerns | Low risk | No concerns | No concerns | Major concerns | Major concerns | Very low |
| Mobile-App: Web-App | 0 | Some concerns | Low risk | No concerns | Major concerns | No concerns | Major concerns | Very low |
| Telemedicine: VR | 0 | Some concerns | Low risk | No concerns | No concerns | Major concerns | Major concerns | Very low |

**Table S50.** Health-related quality of life at post-intervention

| Comparison | Number of studies | Within-study bias | Reporting bias | Indirectness | Imprecision | Heterogeneity | Incoherence | Confidence rating |
|---|---|---|---|---|---|---|---|---|
| Biofeedback devices: Face-to-face care | 2 | Some concerns | Low risk | No concerns | Major concerns | No concerns | Major concerns | Very low |
| Control: Mobile-App | 4 | Some concerns | Low risk | No concerns | No concerns | Major concerns | No concerns | Very low |
| Control: Telemedicine | 5 | Some concerns | Low risk | No concerns | No concerns | Major concerns | No concerns | Very low |
| Control: VR | 4 | Some concerns | Low risk | No concerns | No concerns | No concerns | No concerns | **Moderate** |
| Control: Web-App | 1 | Some concerns | Low risk | No concerns | Major concerns | No concerns | Major concerns | Very low |
| Face-to-face care: Mobile-App | 8 | Some concerns | Low risk | No concerns | Some concerns | Some concerns | No concerns | Low |
| Face-to-face care: Telemedicine | 3 | Some concerns | Low risk | No concerns | Major concerns | No concerns | No concerns | Very low |
| Face-to-face care: VR | 1 | Some concerns | Low risk | No concerns | No concerns | Some concerns | No concerns | Low |
| Mobile-App: Telemedicine | 1 | Some concerns | Low risk | No concerns | Major concerns | No concerns | No concerns | Very low |
| Biofeedback devices: Control | 0 | Some concerns | Low risk | No concerns | Major concerns | No concerns | Major concerns | Very low |
| Biofeedback devices: Mobile-App | 0 | Some concerns | Low risk | No concerns | Major concerns | No concerns | Major concerns | Very low |
| Biofeedback devices: Telemedicine | 0 | Some concerns | Low risk | No concerns | Major concerns | No concerns | Major concerns | Very low |
| Biofeedback devices: VR | 0 | Some concerns | Low risk | No concerns | No concerns | Major concerns | Major concerns | Very low |
| Biofeedback devices: Web-App | 0 | Some concerns | Low risk | No concerns | Major concerns | No concerns | Major concerns | Very low |

| | | | | | | | | |
|---|---|---|---|---|---|---|---|---|
| Control: Face-to-face care | 0 | Some concerns | Low risk | No concerns | No concerns | Major concerns | Major concerns | Very low |
| Face-to-face care: Web-App | 0 | Some concerns | Low risk | No concerns | Major concerns | No concerns | Major concerns | Very low |
| Mobile-App: VR | 0 | Some concerns | Low risk | No concerns | No concerns | Major concerns | Major concerns | Very low |
| Mobile-App: Web-App | 0 | Some concerns | Low risk | No concerns | Major concerns | No concerns | Major concerns | Very low |
| Telemedicine: VR | 0 | Some concerns | Low risk | No concerns | No concerns | Major concerns | Major concerns | Very low |
| Telemedicine: Web-App | 0 | Some concerns | Low risk | No concerns | Major concerns | No concerns | Major concerns | Very low |
| VR: Web-App | 0 | Some concerns | Low risk | No concerns | No concerns | Some concerns | Major concerns | Very low |

**Table S51.** Self-efficacy at post-intervention

| Comparison | Number of studies | Within-study bias | Reporting bias | Indirectness | Imprecision | Heterogeneity | Incoherence | Confidence rating |
|---|---|---|---|---|---|---|---|---|
| Biofeedback devices: Control | 1 | Some concerns | Low risk | No concerns | Some concerns | Some concerns | No concerns | Low |
| Biofeedback devices: Face-to-face care | 1 | Some concerns | Low risk | No concerns | Major concerns | No concerns | No concerns | Very low |
| Control: Exergaming | 2 | Major concerns | Low risk | No concerns | No concerns | Major concerns | Major concerns | Very low |
| Control: Face-to-face care | 1 | Some concerns | Low risk | No concerns | Some concerns | Some concerns | No concerns | Low |
| Control: Mobile-App | 2 | Some concerns | Low risk | No concerns | No concerns | Major concerns | No concerns | Very low |
| Control: Telemedicine | 3 | Some concerns | Low risk | No concerns | Major concerns | No concerns | No concerns | Very low |
| Control: Web-App | 3 | Some concerns | Low risk | No concerns | Some concerns | Some concerns | No concerns | Low |
| Face-to-face care: Mobile-App | 3 | Some concerns | Low risk | No concerns | Major concerns | No concerns | No concerns | Very low |
| Face-to-face care: Telemedicine | 1 | Some concerns | Low risk | No concerns | Major concerns | No concerns | No concerns | Very low |
| Face-to-face care: VR | 2 | Major concerns | Low risk | No concerns | Major concerns | No concerns | Major concerns | Very low |
| Face-to-face care: Web-App | 1 | Some concerns | Low risk | No concerns | Major concerns | No concerns | No concerns | Very low |
| Biofeedback devices: Exergaming | 0 | Some concerns | Low risk | No concerns | Major concerns | No concerns | Major concerns | Very low |
| Biofeedback devices: Mobile-App | 0 | Some concerns | Low risk | No concerns | Major concerns | No concerns | Major concerns | Very low |
| Biofeedback devices: Telemedicine | 0 | Some concerns | Low risk | No concerns | Major concerns | No concerns | Major concerns | Very low |
| Biofeedback devices: VR | 0 | Some concerns | Low risk | No concerns | Major concerns | No concerns | Major concerns | Very low |
| Biofeedback devices: Web-App | 0 | Some concerns | Low risk | No concerns | Major concerns | No concerns | Major concerns | Very low |

| | | | | | | | | |
|---|---|---|---|---|---|---|---|---|
| Control: VR | 0 | Some concerns | Low risk | No concerns | Some concerns | Some concerns | Major concerns | Very low |
| Exergaming: Face-to-face care | 0 | Some concerns | Low risk | No concerns | Major concerns | No concerns | Major concerns | Very low |
| Exergaming: Mobile-App | 0 | Some concerns | Low risk | No concerns | Major concerns | No concerns | Major concerns | Very low |
| Exergaming: Telemedicine | 0 | Some concerns | Low risk | No concerns | Some concerns | Some concerns | Major concerns | Very low |
| Exergaming: VR | 0 | Some concerns | Low risk | No concerns | Major concerns | No concerns | Major concerns | Very low |
| Exergaming: Web-App | 0 | Some concerns | Low risk | No concerns | Major concerns | No concerns | Major concerns | Very low |
| Mobile-App: Telemedicine | 0 | Some concerns | Low risk | No concerns | Major concerns | No concerns | Major concerns | Very low |
| Mobile-App: VR | 0 | Some concerns | Low risk | No concerns | Major concerns | No concerns | Major concerns | Very low |
| Mobile-App: Web-App | 0 | Some concerns | Low risk | No concerns | Major concerns | No concerns | Major concerns | Very low |
| Telemedicine: VR | 0 | Some concerns | Low risk | No concerns | Major concerns | No concerns | Major concerns | Very low |
| Telemedicine: Web-App | 0 | Some concerns | Low risk | No concerns | Major concerns | No concerns | Major concerns | Very low |
| VR: Web-App | 0 | Some concerns | Low risk | No concerns | Major concerns | No concerns | Major concerns | Very low |

**Table S52.** Anxiety at post-intervention

| Comparison | Number of studies | Within-study bias | Reporting bias | Indirectness | Imprecision | Heterogeneity | Incoherence | Confidence rating |
|---|---|---|---|---|---|---|---|---|
| Control: Mobile-App | 1 | Some concerns | Low risk | No concerns | Major concerns | No concerns | No concerns | Very low |
| Control: Telemedicine | 3 | Major concerns | Low risk | No concerns | Some concerns | No concerns | No concerns | Very low |
| Control: VR | 2 | No concerns | Low risk | No concerns | Some concerns | Some concerns | No concerns | **Moderate** |
| Exergaming: Face-to-face care | 1 | Some concerns | Low risk | No concerns | Major concerns | No concerns | No concerns | Very low |
| Face-to-face care: Mobile-App | 2 | Some concerns | Low risk | No concerns | Some concerns | Some concerns | No concerns | Low |
| Face-to-face care: VR | 1 | Some concerns | Low risk | No concerns | Major concerns | No concerns | No concerns | Very low |
| Face-to-face care: Web-App | 1 | Major concerns | Low risk | No concerns | Major concerns | No concerns | No concerns | Very low |
| Control: Exergaming | 0 | Some concerns | Low risk | No concerns | Major concerns | No concerns | No concerns | Very low |
| Control: Face-to-face care | 0 | Some concerns | Low risk | No concerns | Major concerns | No concerns | No concerns | Very low |
| Control: Web-App | 0 | Some concerns | Low risk | No concerns | Major concerns | No concerns | No concerns | Very low |
| Exergaming: Mobile-App | 0 | Some concerns | Low risk | No concerns | Major concerns | No concerns | No concerns | Very low |
| Exergaming: Telemedicine | 0 | Some concerns | Low risk | No concerns | Major concerns | No concerns | No concerns | Very low |

| | | | | | | | | |
|---|---|---|---|---|---|---|---|---|
| Exergaming: VR | 0 | Some concerns | Low risk | No concerns | Major concerns | No concerns | No concerns | Very low |
| Exergaming: Web-App | 0 | Major concerns | Low risk | No concerns | Major concerns | No concerns | No concerns | Very low |
| Face-to-face care: Telemedicine | 0 | Some concerns | Low risk | No concerns | Major concerns | No concerns | No concerns | Very low |
| Mobile-App: Telemedicine | 0 | Some concerns | Low risk | No concerns | Some concerns | Some concerns | No concerns | Low |
| Mobile-App: VR | 0 | Some concerns | Low risk | No concerns | Major concerns | No concerns | No concerns | Very low |
| Mobile-App: Web-App | 0 | Major concerns | Low risk | No concerns | Some concerns | Some concerns | No concerns | Very low |
| Telemedicine: VR | 0 | Some concerns | Low risk | No concerns | Some concerns | Some concerns | No concerns | Low |
| Telemedicine: Web-App | 0 | Some concerns | Low risk | No concerns | Major concerns | No concerns | No concerns | Very low |
| VR: Web-App | 0 | Some concerns | Low risk | No concerns | Major concerns | No concerns | No concerns | Very low |

**Table S53.** Depression at post-intervention

| Comparison | Number of studies | Within-study bias | Reporting bias | Indirectness | Imprecision | Heterogeneity | Incoherence | Confidence rating |
|---|---|---|---|---|---|---|---|---|
| Biofeedback devices: Face-to-face care | 1 | Some concerns | Low risk | No concerns | Major concerns | No concerns | No concerns | Very low |
| Control: Mobile-App | 2 | Some concerns | Low risk | No concerns | Major concerns | No concerns | No concerns | Very low |
| Control: Telemedicine | 3 | Major concerns | Low risk | No concerns | No concerns | No concerns | No concerns | Low |
| Control: VR | 2 | No concerns | Low risk | No concerns | No concerns | Some concerns | No concerns | **Moderate** |
| Exergaming: Face-to-face care | 1 | Some concerns | Low risk | No concerns | Major concerns | No concerns | No concerns | Very low |
| Face-to-face care: Mobile-App | 2 | Some concerns | Low risk | No concerns | Some concerns | No concerns | No concerns | Low |
| Face-to-face care: Telemedicine | 1 | Some concerns | Low risk | No concerns | Major concerns | No concerns | No concerns | Very low |
| Face-to-face care: VR | 1 | Some concerns | Low risk | No concerns | Some concerns | Some concerns | No concerns | Low |
| Face-to-face care: Web-App | 1 | No concerns | Low risk | No concerns | Major concerns | No concerns | No concerns | Low |
| Biofeedback devices: Control | 0 | Some concerns | Low risk | No concerns | Some concerns | No concerns | No concerns | Low |
| Biofeedback devices: Exergaming | 0 | Some concerns | Low risk | No concerns | Major concerns | No concerns | No concerns | Very low |
| Biofeedback devices: Mobile-App | 0 | Some concerns | Low risk | No concerns | Some concerns | No concerns | No concerns | Low |
| Biofeedback devices: Telemedicine | 0 | Some concerns | Low risk | No concerns | Major concerns | No concerns | No concerns | Very low |
| Biofeedback devices: VR | 0 | Some concerns | Low risk | No concerns | Some concerns | Some concerns | No concerns | Low |
| Biofeedback devices: Web-App | 0 | Some concerns | Low risk | No concerns | Major concerns | No concerns | No concerns | Very low |

| | | | | | | | | |
|---|---|---|---|---|---|---|---|---|
| Control: Exergaming | 0 | Some concerns | Low risk | No concerns | Some concerns | Some concerns | No concerns | Low |
| Control: Face-to-face care | 0 | Some concerns | Low risk | No concerns | Some concerns | No concerns | No concerns | Low |
| Control: Web-App | 0 | Some concerns | Low risk | No concerns | Major concerns | No concerns | No concerns | Very low |
| Exergaming: Mobile-App | 0 | Some concerns | Low risk | No concerns | Some concerns | Some concerns | No concerns | Low |
| Exergaming: Telemedicine | 0 | Some concerns | Low risk | No concerns | Major concerns | No concerns | No concerns | Very low |
| Exergaming: VR | 0 | Some concerns | Low risk | No concerns | Major concerns | No concerns | No concerns | Very low |
| Exergaming: Web-App | 0 | Some concerns | Low risk | No concerns | Major concerns | No concerns | No concerns | Very low |
| Mobile-App: Telemedicine | 0 | Some concerns | Low risk | No concerns | Some concerns | No concerns | No concerns | Low |
| Mobile-App: VR | 0 | Some concerns | Low risk | No concerns | Some concerns | Some concerns | No concerns | Low |
| Mobile-App: Web-App | 0 | Some concerns | Low risk | No concerns | Major concerns | No concerns | No concerns | Very low |
| Telemedicine: VR | 0 | Some concerns | Low risk | No concerns | Some concerns | No concerns | No concerns | Low |
| Telemedicine: Web-App | 0 | Some concerns | Low risk | No concerns | Major concerns | No concerns | No concerns | Very low |
| VR: Web-App | 0 | Some concerns | Low risk | No concerns | Major concerns | No concerns | No concerns | Very low |

**Section 19.** Back-transformation to clinical scales

To aid clinical interpretation, we performed a post hoc back-transformation for the two primary outcomes at post-intervention. Following established guidance for interpreting standardised mean differences, we multiplied each network SMD and its confidence limits by a representative SD for the corresponding instrument.[104-106] Pain-intensity estimates were re-expressed on the 0–10 numerical rating scale (NRS)[107] using a reference SD of 1.70, defined as the median baseline SD across eligible control groups reporting 0–10 numerical pain rating scales. Physical-function estimates were re-expressed on the 0–100 Oswestry Disability Index (ODI)[108] using a reference SD of 13.55, defined as the median baseline SD across eligible control groups directly reporting the ODI on a 0–100 scale. These back-transformed estimates were used solely to aid clinical interpretation and were not used for network estimation, treatment ranking, or CINeMA assessment.

Back-transformed differences were calculated by multiplying the network SMD and each bound of its 95% confidence interval by the corresponding reference SD:

$$\text{Back-transformed difference} = \text{SMD} \times \text{reference SD}$$

$$\text{Lower 95\% CI} = \text{lower 95\% CI of SMD} \times \text{reference SD}$$

$$\text{Upper 95\% CI} = \text{upper 95\% CI of SMD} \times \text{reference SD}$$

**Table S54.** Reference SD for pain intensity

| Study | Control n | Baseline mean | Baseline SD |
|---|---|---|---|
| Krein 2013 | 118 | 6.10 | 1.60 |
| Irvine 2015 | 398 | 2.74 | 1.18 |
| Geraghty 2018 | 26 | 3.80 | 2.30 |
| Williams 2018 | 80 | 6.80 | 1.60 |
| Amorim 2019 | 34 | 5.10 | 1.40 |
| Zadro 2019 | 30 | 4.80 | 1.70 |
| Sandal 2021 | 229 | 4.90 | 1.90 |
| Eccleston 2022 | 28 | 5.52 | 1.91 |
| Itoh 2022 | 51 | 5.10 | 2.10 |
| Meinke 2022 | 14 | 2.57 | 1.60 |
| Kent 2023 | 165 | 6.20 | 1.28 |

| | | | |
|---|---|---|---|
| Sanabria-Mazo 2023 | 78 | 6.95 | 1.69 |
| Priebe 2024 | 307 | 5.27 | 1.76 |
| Santos 2024 | 19 | 6.50 | 1.88 |
| Feng 2025 | 39 | 3.47 | 1.28 |
| Spiegel 2026 | 127 | 6.00 | 1.90 |

Note: The reference SD was defined as the median baseline SD across eligible control groups that reported pain intensity on a 0–10 numerical rating scale. Sixteen eligible trials contributed to this calculation, yielding a median baseline SD of 1.70 points (IQR 1.55–1.90), which was used to back-transform the corresponding standardised mean differences. The term NRS denotes 0–10 numerical pain rating scales reported as NRS in the source reports.

**Table S55.** Reference SD for physical function

| Study | Control n | Baseline mean | Baseline SD |
|---|---|---|---|
| Iles 2011 | 15 | 41.00 | 13.00 |
| Shebib 2019 | 64 | 21.00 | 9.66 |
| Almhdawi 2020 | 20 | 31.05 | 10.75 |
| Kim 2021 | 22 | 33.00 | 14.10 |
| Lazaridou 2023 | 29 | 32.30 | 15.40 |
| Maddox 2023 | 531 | 40.95 | 16.93 |

Note: The reference SD was defined as the median baseline SD across eligible control groups reporting physical function on the 0–100 Oswestry Disability Index (ODI). Six eligible trials contributed to this calculation, yielding a median baseline SD of 13.55 points (IQR 11.31–15.08), which was used to back-transform the corresponding standardised mean differences. Trials reporting the ODI on other scoring ranges or modified versions of the ODI were not included in the reference-SD calculation.

**Table S56.** Back-transformed post-intervention estimates

| Outcome | Comparison | SMD (95% CI) | Reference scale | Reference SD | Back-transformed difference (95% CI) |
|---|---|---|---|---|---|
| Pain intensity | Telemedicine vs Control | −0.51 (−0.94 to −0.07) | NRS (0–10) | 1.70 | **−0.87 (−1.60 to −0.12)** |
| | Web-App vs Control | −0.54 (−1.06 to −0.02) | NRS (0–10) | 1.70 | **−0.92 (−1.80 to −0.03)** |
| | Mobile-App vs Control | −0.67 (−1.08 to −0.26) | NRS (0–10) | 1.70 | **−1.14 (−1.84 to −0.44)** |
| | VR vs Control | −1.43 (−1.87 to −0.98) | NRS (0–10) | 1.70 | **−2.43 (−3.18 to −1.67)** |
| | Exergaming vs Control | −1.09 (−1.73 to −0.44) | NRS (0–10) | 1.70 | **−1.85 (−2.94 to −0.75)** |
| | Biofeedback devices vs Control | −0.48 (−1.09 to 0.13) | NRS (0–10) | 1.70 | **−0.82 (−1.85 to 0.22)** |
| | Face-to-face care vs Control | −0.31 (−0.68 to 0.06) | NRS (0–10) | 1.70 | **−0.53 (−1.16 to 0.10)** |
| Physical function | Telemedicine vs Control | −0.57 (−1.01 to −0.13) | ODI (0–100) | 13.55 | **−7.72 (−13.69 to −1.76)** |
| | Web-App vs Control | −0.78 (−1.27 to −0.30) | ODI (0–100) | 13.55 | **−10.57 (−17.21 to −4.07)** |
| | Mobile-App vs Control | −0.73 (−1.22 to −0.23) | ODI (0–100) | 13.55 | **−9.89 (−16.53 to −3.12)** |
| | VR vs Control | −0.86 (−1.45 to −0.26) | ODI (0–100) | 13.55 | **−11.65 (−19.65 to −3.52)** |
| | Exergaming vs Control | −0.52 (−1.23 to 0.19) | ODI (0–100) | 13.55 | **−7.05 (−16.67 to 2.57)** |
| | Biofeedback devices vs Control | −0.70 (−1.38 to −0.03) | ODI (0–100) | 13.55 | **−9.49 (−18.70 to −0.41)** |
| | Face-to-face care vs Control | −0.35 (−0.78 to 0.09) | ODI (0–100) | 13.55 | **−4.74 (−10.57 to 1.22)** |

Note: Back-transformed differences were calculated by multiplying each SMD and its 95% confidence limits by the corresponding reference SD. A reference SD of 1.70 points was used for pain intensity on the 0–10 NRS, and a reference SD of 13.55 points was used for physical function on the 0–100 ODI. Negative values indicate lower pain intensity or lower disability and therefore favour the intervention. Back-transformed estimates are approximate transformations intended to facilitate clinical interpretation and should not be interpreted as directly pooled mean differences on the corresponding instruments or as estimates of the proportion of participants achieving a minimal clinically important difference. CI, confidence interval; NRS, numerical rating scale; ODI, Oswestry Disability Index; SD, standard deviation; SMD, standardised mean difference.
